# Risk of Severe Outcomes From COVID-19 in Comorbid Populations in the Omicron Era: A Meta-analysis

**DOI:** 10.1101/2024.12.02.24317727

**Authors:** Akvile Chapman, Dan H. Barouch, Gregory Y. H. Lip, Triantafyllos Pliakas, Eva Polverino, Harald Sourij, Sultan Abduljawad

## Abstract

**Importance:** This is the first meta-analysis to investigate risk of death and hospitalization in individuals with comorbidities, specifically during the Omicron era.

**Objective:** To assess the risk of mortality and hospitalization from COVID-19 in individuals with comorbidities in comparison with individuals without comorbidities during the Omicron era.

**Data Sources:** A systematic search of Embase, MEDLINE, PubMed, Europe PMC, Latin American and Caribbean Health Sciences Literature, Cochrane COVID-19 Study Register, and WHO COVID-19 Database was performed to identify studies published between 1 January 2022 and 13 March 2024.

**Study Selection:** Inclusion criteria were observational studies including people (all ages) with at least 1 of the following comorbidities: cardiovascular/ cerebrovascular disease, chronic lung conditions, diabetes, and obesity. In total, 72 studies were included in the review, of which 68 were meta-analyzed.

**Data Extraction and Synthesis:** Data were extracted by one reviewer and verified by a second. Studies were synthesized quantitively (meta-analysis) using random-effect models. PRISMA guidelines were followed.

**Main Outcomes and Measures:** Evaluated outcomes were the risks of death, hospitalization, intensive care unit (ICU) admission, and any combination of these outcomes. Odds ratios, hazard ratios, and rate ratios were extracted; pooled relative risk (RR) and 95% confidence intervals (CI) were calculated.

**Results:** Minimum numbers of participants per comorbidity across included studies ranged from 328 870 for thrombosis to 13 720 480 for hypertension. Risks of death, hospitalization, and the combined outcome were increased in individuals with cerebrovascular disease, COPD, diabetes, respiratory diseases, heart disease, and heart failure versus those without (pooled RRs ranged from 1.27 [heart disease, hospitalization; 95% CI, 1.17-1.38, *P* < .001] to 1.78 [heart failure, death: 95% CI, 1.46-2.16, *P* < .001]). Individuals with diabetes and obesity had increased risk of ICU admission (RR: 1.20; 95% CI: 1.04-1.38, *P* = .0141 and RR: 1.32; 95% CI: 1.11-1.57, *P* = .00158, respectively).

**Conclusions:** During the Omicron era, risk of death and hospitalization from COVID-19 is increased amongst individuals with comorbidities including cerebrovascular/cardiovascular conditions, chronic lung diseases, and diabetes, with the highest risk in those with heart failure. Individuals with diabetes and obesity are at increased risk of ICU admission.

**Key Points:** *Question:* What are the risks of severe outcomes from COVID-19 in individuals with comorbidities during the Omicron era?

*Findings:* This systematic review and meta-analysis found increased risk of mortality and hospitalization among individuals with a range of comorbidities, including cerebrovascular/cardiovascular conditions, chronic lung diseases, and diabetes, with the highest risk in those with heart failure, versus those without. Risk of ICU admission was higher in individuals with obesity and diabetes.

*Meaning:* This study identified comorbid populations most at risk of severe outcomes from COVID-19. Targeting these populations with public health measures, such as vaccination, may be beneficial.

## Introduction

At an early stage of the COVID-19 pandemic, individuals with comorbidities, including diabetes, obesity, respiratory, and cardiovascular diseases were more likely to experience adverse health outcomes caused by SARS-CoV-2 infection. ^1,2^ Such populations may be at increased risk of death and hospitalization in comparison with the general population.^1–4^

Patients infected with SARS-CoV-2 Omicron variants have exhibited milder infections and reduced hospitalization compared with the earlier Delta (B.1.617.2) variant.^5–7^ Despite this, Omicron (B.1.1.529) was classified as a variant of concern by the World Health Organization (WHO) in November 2021 due to a detrimental change in COVID-19 epidemiology.^8^ In particular, individuals with pre-existing comorbidities may still be at a higher risk of complications and death from SARS-CoV-2 infection in the current Omicron era than individuals in the general population.^9^ As SARS-CoV-2 Omicron variants evolve, understanding the impact and risk of infection on individuals with comorbidities is crucial for informed treatment and risk mitigation through vaccination or other preventative measures.

To our knowledge, this is the first systematic literature review (SLR) and meta-analysis aimed at assessing the risk of mortality and hospitalization from COVID-19 in individuals with comorbidities, including cardiovascular/cerebrovascular diseases, chronic lung conditions, diabetes, and obesity, during the Omicron era.

## Methods

The SLR protocol is registered with PROSPERO (CRD42024501163). This SLR and meta-analysis adheres to the Preferred Reporting Items for Systematic Reviews and Meta-Analysis guidelines (PRISMA).

### Search Strategy

The following databases were searched: Embase, MEDLINE, PubMed, Europe PMC (including MedRix and bioRxiv preprints), Latin American and Caribbean Health Sciences Literature, the Cochrane COVID-19 Study Register, and the WHO COVID-19 Database. Search strategies were structured using terms related to COVID-19 infection, risk, and burden of illness (**eMethods**).

### Eligibility Criteria

Eligible studies included people (all ages) with at least 1 of the following comorbidities: cardiovascular/cerebrovascular disease, chronic lung conditions, diabetes, and obesity (**eTable 1**).

Individuals without the respective comorbidities or the general population were used as the comparator group. Evaluated outcomes were the risks of death, hospitalization (for any reason), intensive care unit (ICU) admission (for any reason), and the combined outcome (any combination of the other outcomes). COVID-19 outcomes were determined by including studies where either all patients had COVID-19 at the start of the study or all deaths and hospitalisations were related to COVID-19 (defined by the studies).

Included studies were observational (cohort, case-control, cross-sectional), published between 1 January 2022 and 13 March 2024, with full texts published in English.

### Study Selection, Data Extraction, and Quality Assessment

Identified studies were first assessed at title and abstract level using the Rayyan tool^10^ by 2 independent reviewers to determine whether inclusion criteria were met, followed by full text screening of all studies found to be eligible. For details of data extraction and quality assessment, see **eMethods**.

### Data Synthesis and Analysis

#### Qualitative Data Synthesis

All studies included in the review were assessed qualitatively to identify which studies could be combined in a meta-analysis. Comorbidities were grouped as follows, based on recommendations from clinical experts: asthma, cerebrovascular disease, COPD, diabetes, obesity, peripheral vascular disease (PVD), respiratory diseases (excluding asthma and COPD), atrial fibrillation, heart disease, heart failure, hypertension, and thrombosis.

#### Statistical Analysis

For the primary analyses, pairwise meta-analyses were performed for the risks of death, hospitalization, ICU admission, and the combined outcome for the comorbid populations, using the most adjusted reported outcome estimates. The robustness of the results was assessed using ‘Leave-1-out’,^11^ ‘Least adjusted’, ‘Only adjusted’, and ‘Excluding studies for population overlap’ sensitivity analyses.

Subgroup analyses were conducted for the ‘Hospitalized’ or ‘General’ populations, which included only individuals who were or were not already hospitalized when they started the study, respectively. Additional subgroup analyses were performed for the ‘>50 years’ population, which included only participants older than 50 years, and the ‘COVID-19-related outcomes only’ population, which included only outcomes explicitly caused by COVID-19 (**eMethods**).

All statistical analyses were performed in R version 4.1.1 (R Foundation for Statistical Computing) using the meta package. A statistically significant (*P* < .05) result is referred to as significant thereafter.

## Results

In total, 21 937 records were identified through searches and 1 study was identified via reference checking. Following elimination of duplicates, 11 593 remaining studies underwent title and abstract screening of which 3123 studies were assessed in full text screening. A total of 72 studies were selected for inclusion (**Figure 1**).

**Figure 1.**
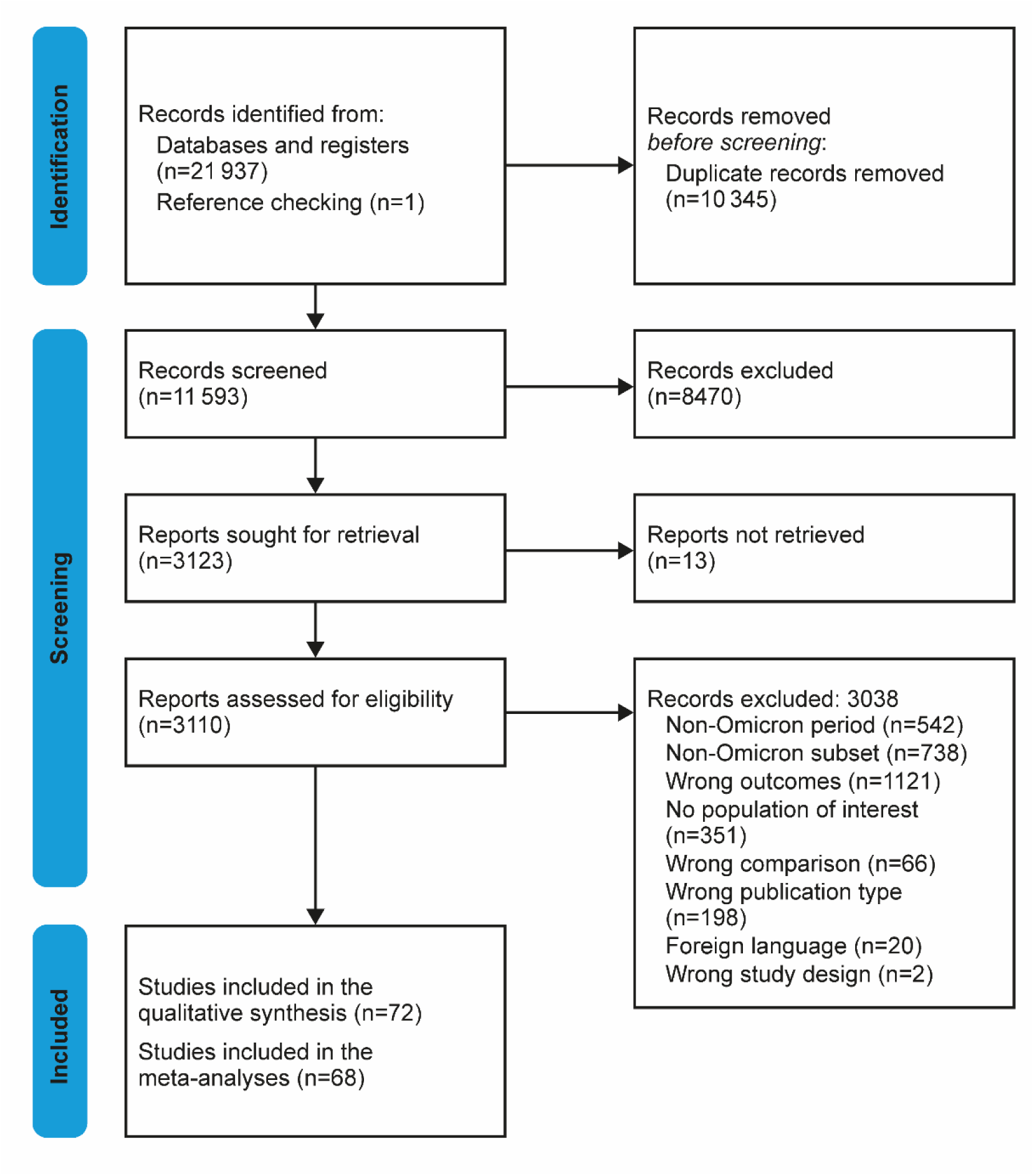
PRISMA Flow Diagram.

### Study Characteristics

Study characteristics are summarized in **Table 1**. The studies were performed in 26 different countries, primarily in Europe (n = 15), China (n = 14), and the USA (n = 13). Most studies were retrospective cohort studies (n = 58), followed by prospective cohort (n = 7), cross-sectional (n = 6), and case-control (n = 1) studies. The ‘Death’, ‘Hospitalization’, ‘ICU admission’, and ‘Combined’ outcomes were reported in 45, 17, 20, and 17 studies, respectively. Most studies did not report Omicron subvariants, but in studies that did, BA.1 was the most common. Other reported subvariants included BA.2, BA.4, BA.5, BF.7, BQ.1, and XXB. **eTable 2** provides visualization of the Omicron period in the included studies.

**Table 1.**
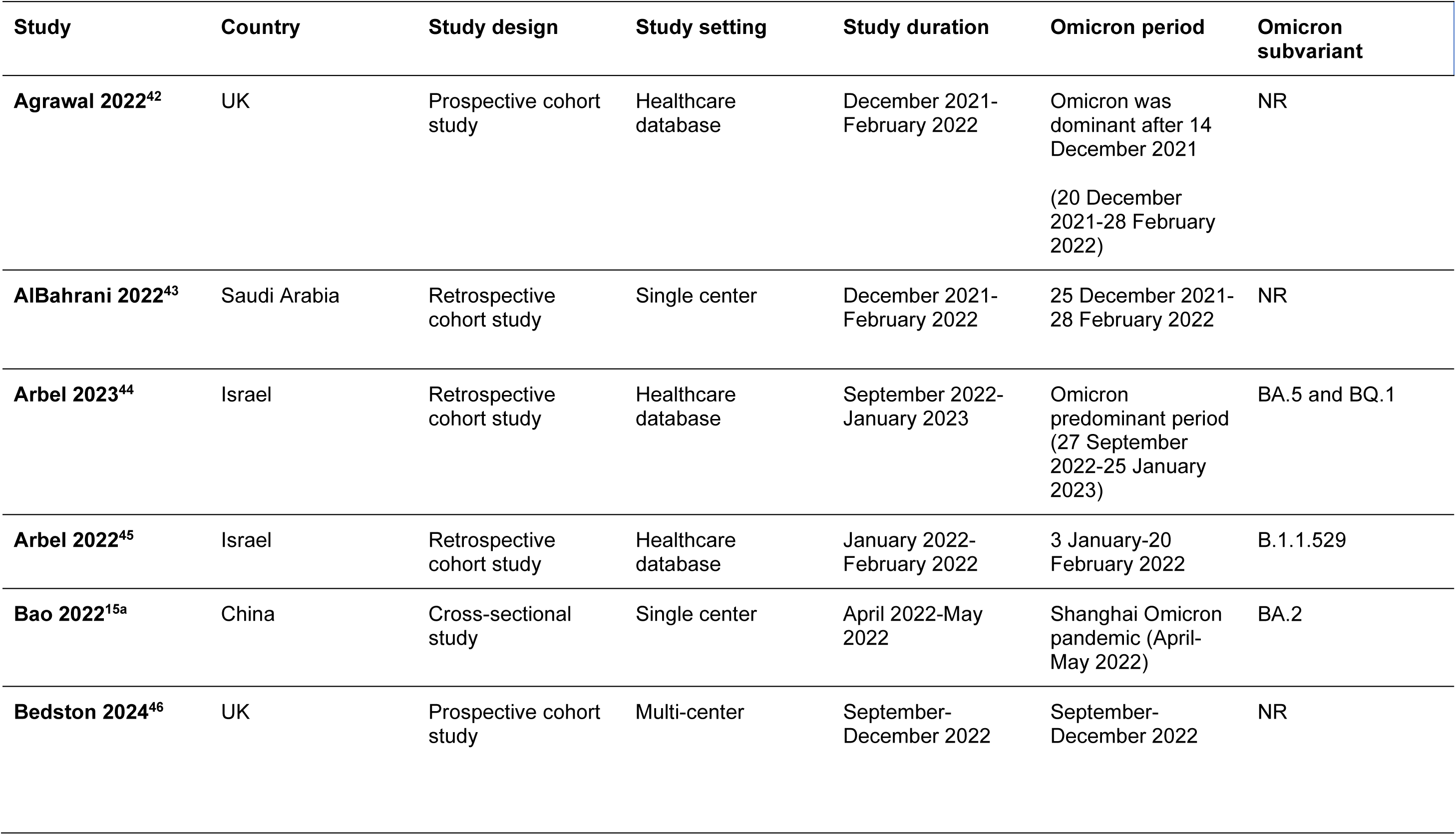
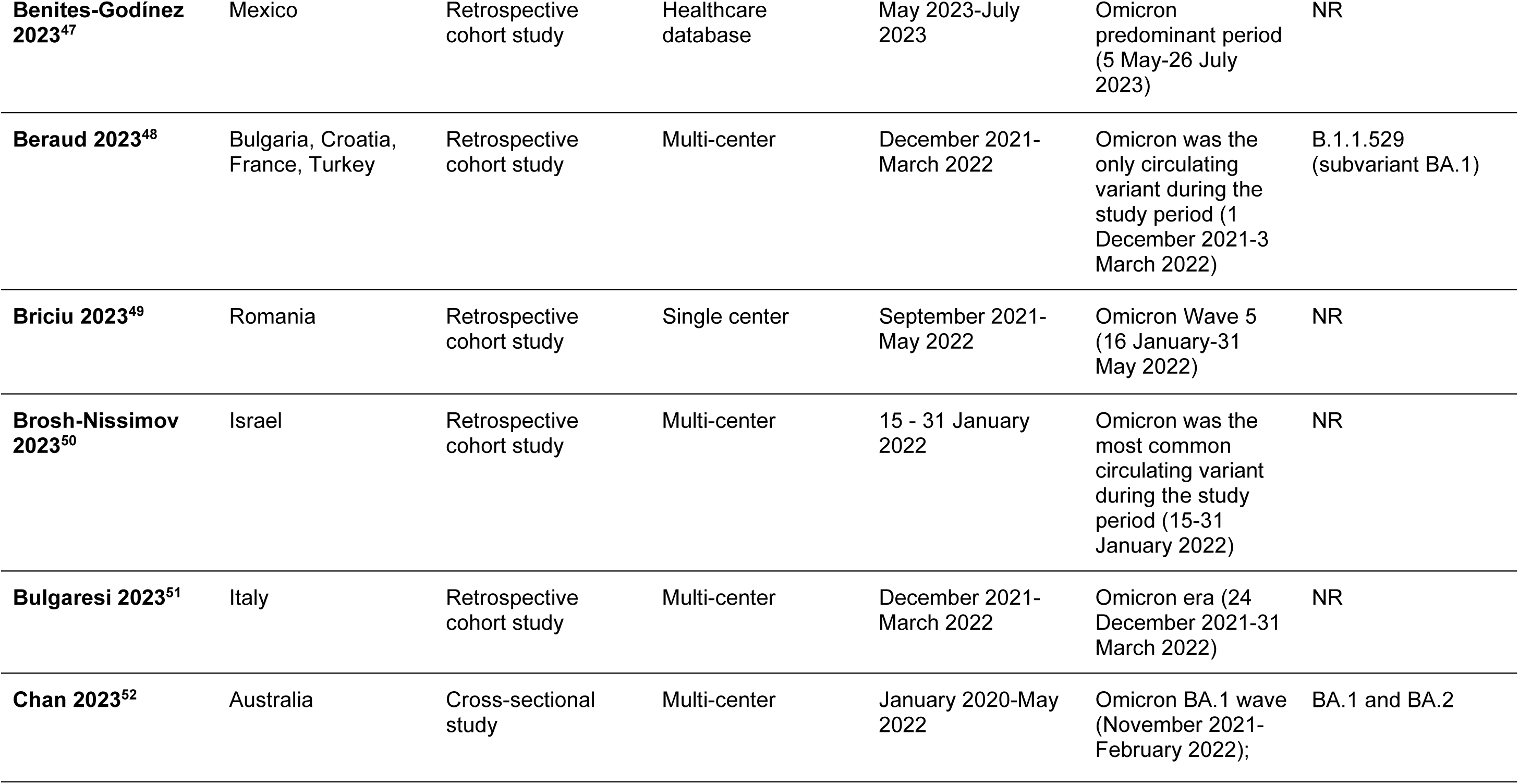
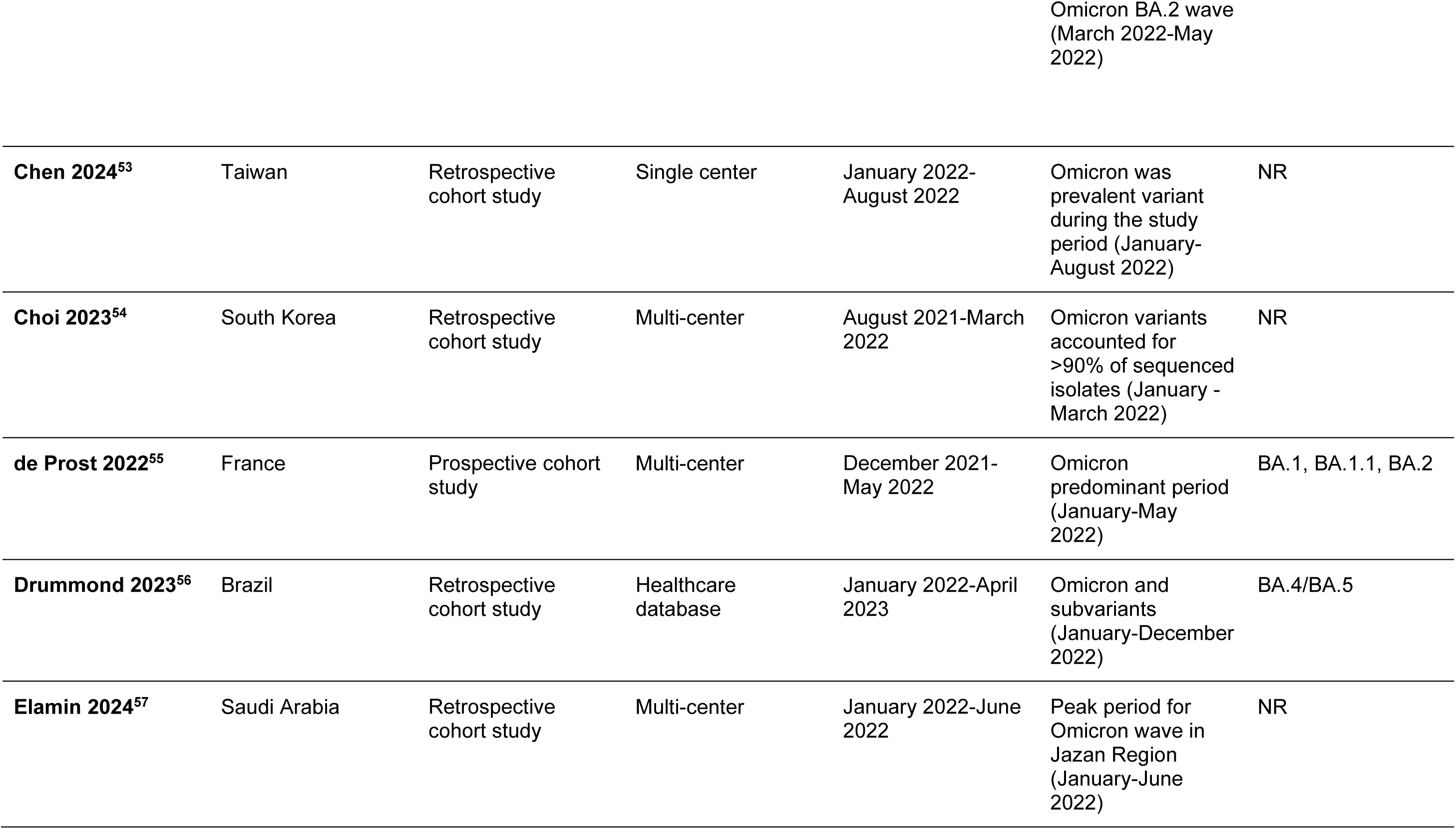
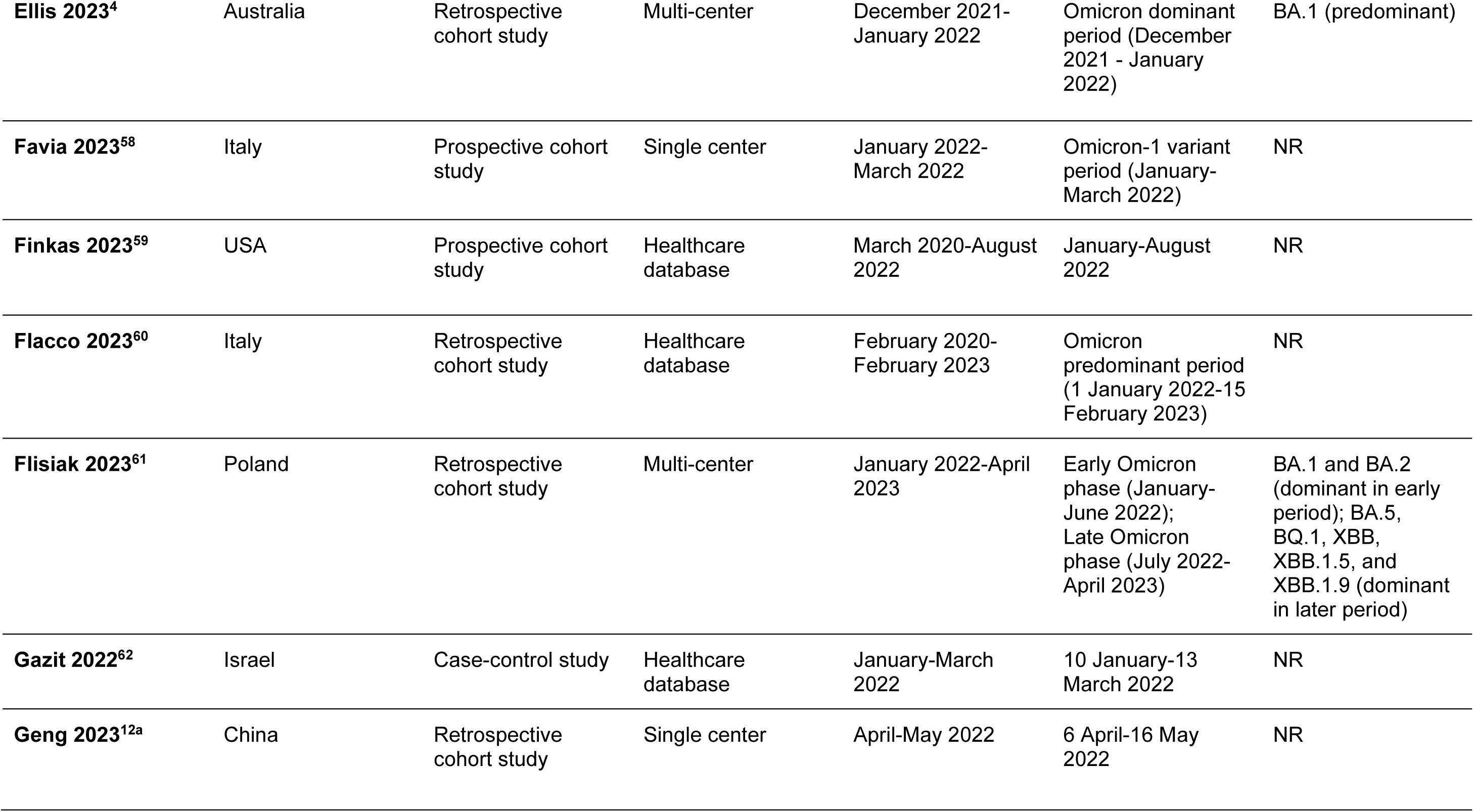
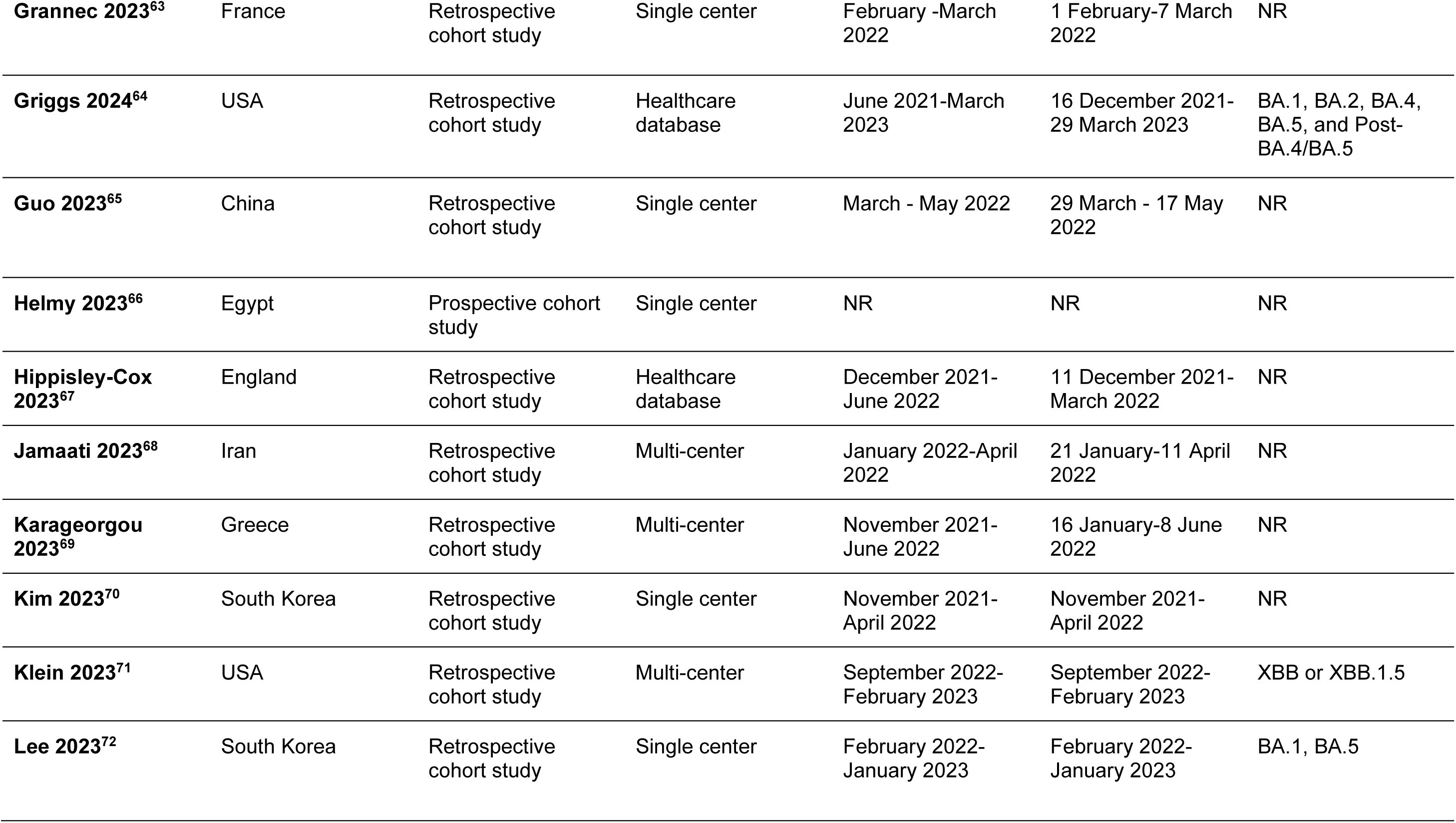
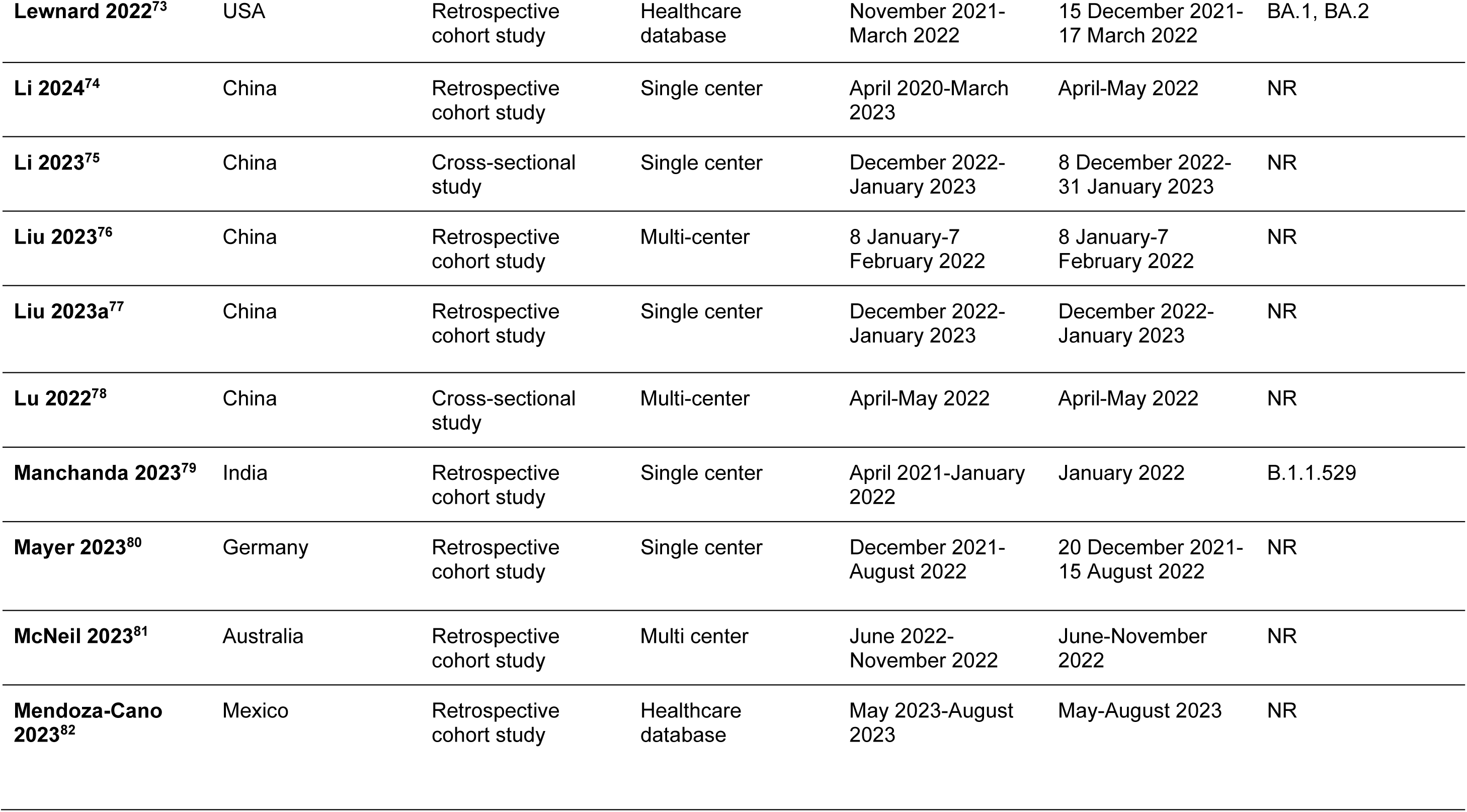
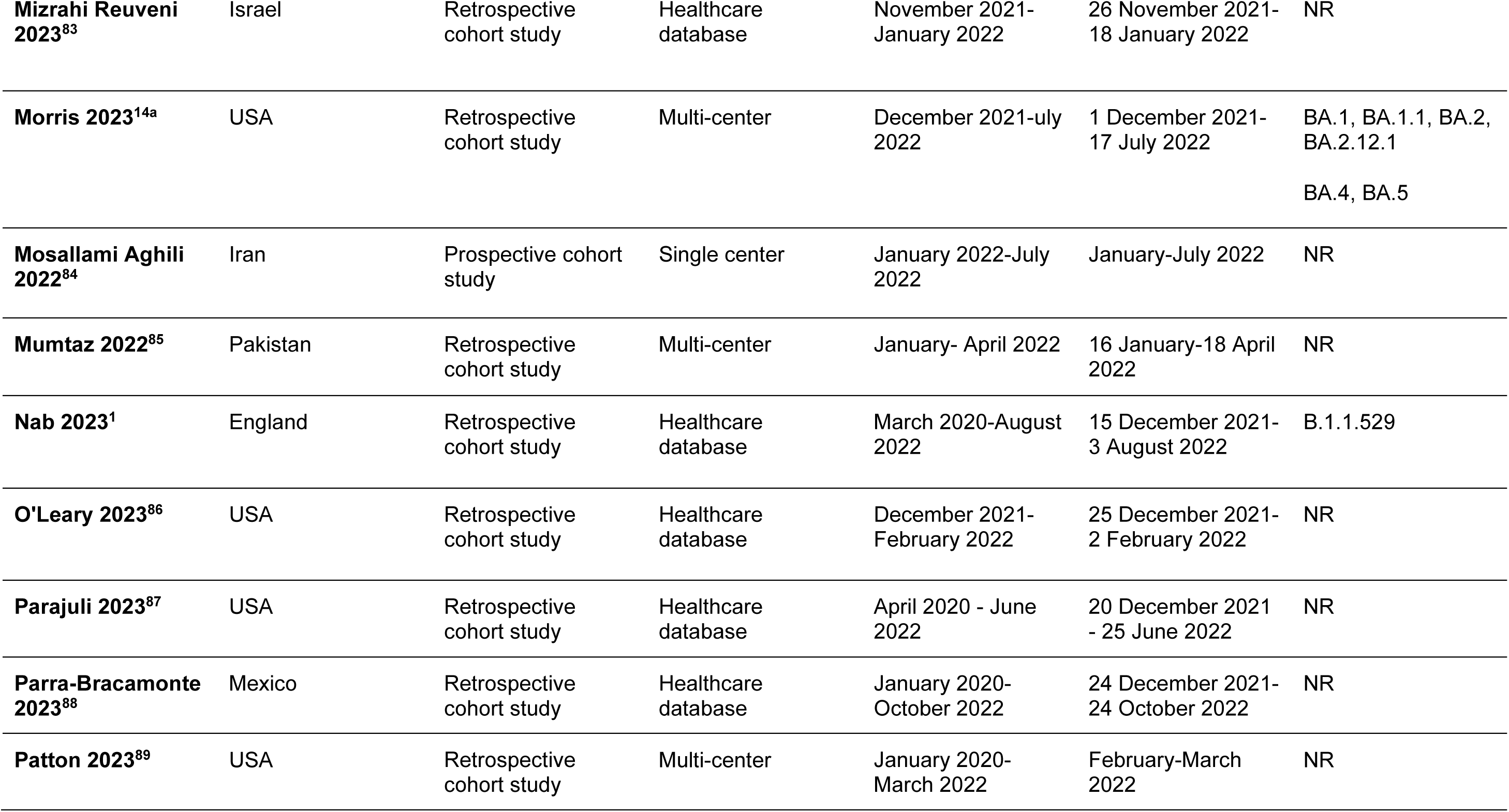
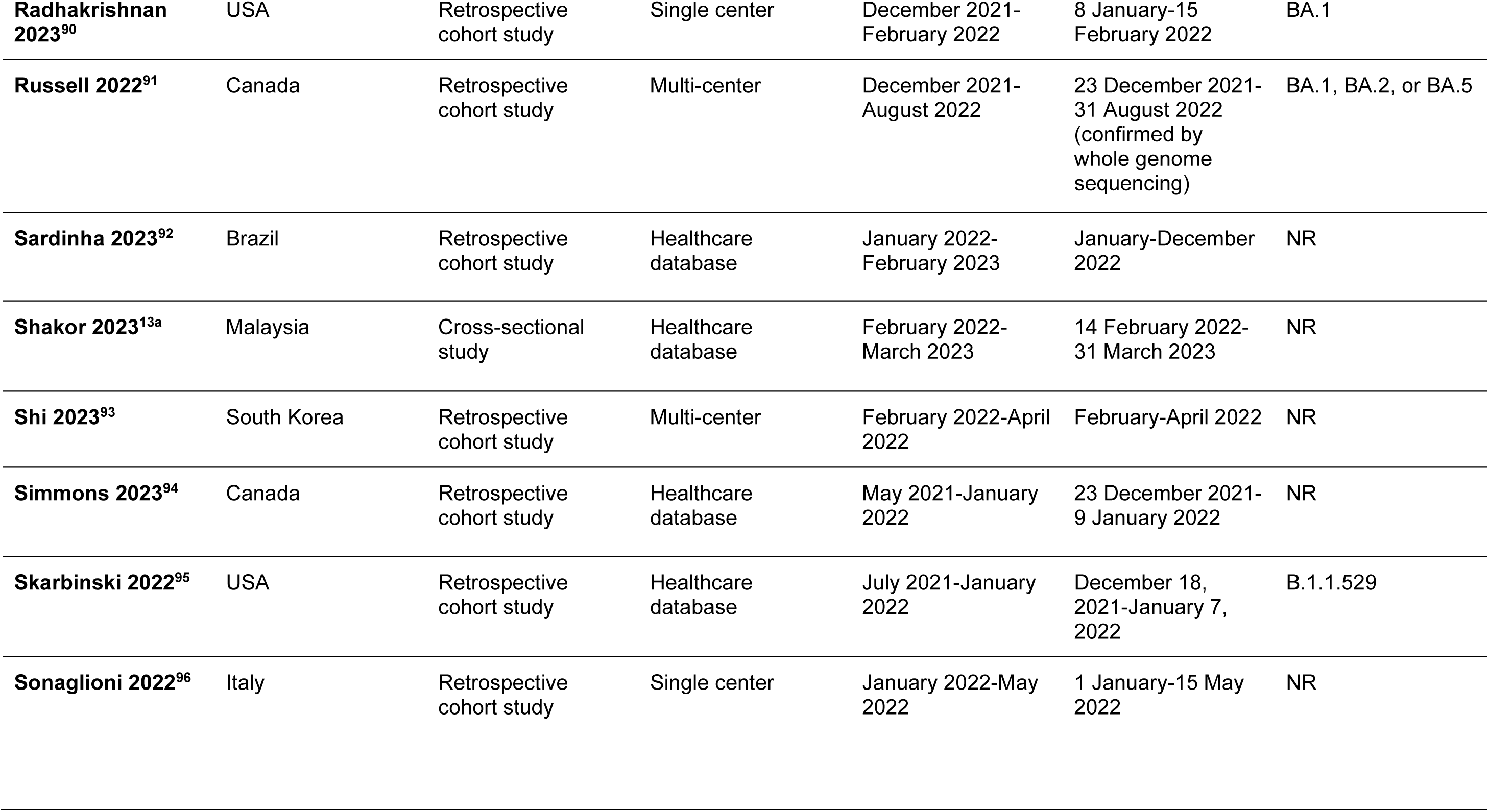
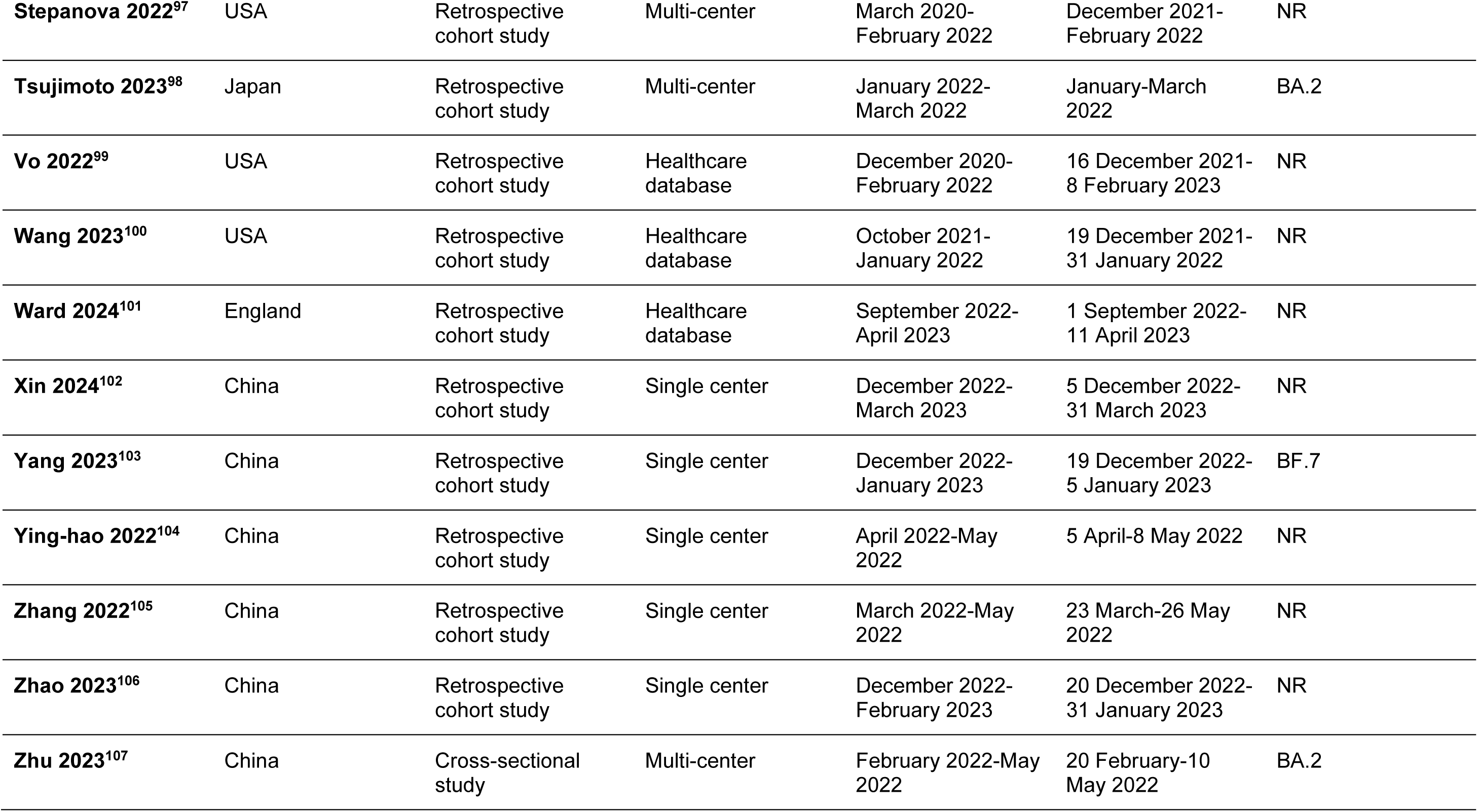
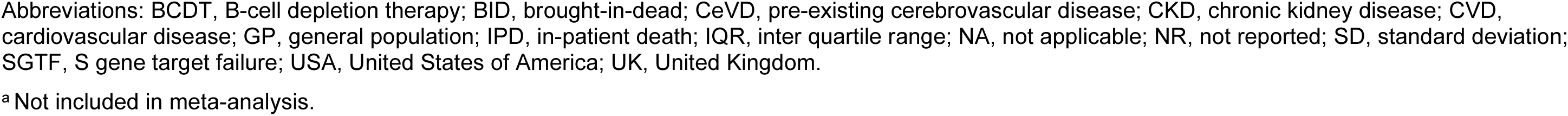
Characteristics of the Included Studies.

| Study | Country | Study design | Study setting | Study duration | Omicron period | Omicron subvariant |
| --- | --- | --- | --- | --- | --- | --- |
| **Agrawal 2022<sup>42</sup>** | UK | Prospective cohort study | Healthcare database | December 2021-February 2022 | Omicron was dominant after 14 December 2021<br>(20 December 2021-28 February 2022) | NR |
| **AlBahrani 2022<sup>43</sup>** | Saudi Arabia | Retrospective cohort study | Single center | December 2021-February 2022 | 25 December 2021-28 February 2022 | NR |
| **Arbel 2023<sup>44</sup>** | Israel | Retrospective cohort study | Healthcare database | September 2022-January 2023 | Omicron predominant period (27 September 2022-25 January 2023) | BA.5 and BQ.1 |
| **Arbel 2022<sup>45</sup>** | Israel | Retrospective cohort study | Healthcare database | January 2022-February 2022 | 3 January-20 February 2022 | B.1.1.529 |
| **Bao 2022<sup>15a</sup>** | China | Cross-sectional study | Single center | April 2022-May 2022 | Shanghai Omicron pandemic (April-May 2022) | BA.2 |
| **Bedston 2024<sup>46</sup>** | UK | Prospective cohort study | Multi-center | September-December 2022 | September-December 2022 | NR |
| **Benites-Godínez 2023<sup>47</sup>** | Mexico | Retrospective cohort study | Healthcare database | May 2023-July 2023 | Omicron predominant period (5 May-26 July 2023) | NR |
| **Beraud 2023<sup>48</sup>** | Bulgaria, Croatia, France, Turkey | Retrospective cohort study | Multi-center | December 2021-March 2022 | Omicron was the only circulating variant during the study period (1 December 2021-3 March 2022) | B.1.1.529 (subvariant BA.1) |
| **Briciu 2023<sup>49</sup>** | Romania | Retrospective cohort study | Single center | September 2021-May 2022 | Omicron Wave 5 (16 January-31 May 2022) | NR |
| **Brosh-Nissimov 2023<sup>50</sup>** | Israel | Retrospective cohort study | Multi-center | 15 - 31 January 2022 | Omicron was the most common circulating variant during the study period (15-31 January 2022) | NR |
| **Bulgaresi 2023<sup>51</sup>** | Italy | Retrospective cohort study | Multi-center | December 2021-March 2022 | Omicron era (24 December 2021-31 March 2022) | NR |
| **Chan 2023<sup>52</sup>** | Australia | Cross-sectional study | Multi-center | January 2020-May 2022 | Omicron BA.1 wave (November 2021-February 2022); | BA.1 and BA.2 |
|  |  |  |  |  | Omicron BA.2 wave (March 2022-May 2022) |  |
| **Chen 2024<sup>53</sup>** | Taiwan | Retrospective cohort study | Single center | January 2022-August 2022 | Omicron was prevalent variant during the study period (January-August 2022) | NR |
| **Choi 2023<sup>54</sup>** | South Korea | Retrospective cohort study | Multi-center | August 2021-March 2022 | Omicron variants accounted for >90% of sequenced isolates (January - March 2022) | NR |
| **de Prost 2022<sup>55</sup>** | France | Prospective cohort study | Multi-center | December 2021-May 2022 | Omicron predominant period (January-May 2022) | BA.1, BA.1.1, BA.2 |
| **Drummond 2023<sup>56</sup>** | Brazil | Retrospective cohort study | Healthcare database | January 2022-April 2023 | Omicron and subvariants (January-December 2022) | BA.4/BA.5 |
| **Elamin 2024<sup>57</sup>** | Saudi Arabia | Retrospective cohort study | Multi-center | January 2022-June 2022 | Peak period for Omicron wave in Jazan Region (January-June 2022) | NR |
| **Ellis 2023<sup>4</sup>** | Australia | Retrospective cohort study | Multi-center | December 2021-January 2022 | Omicron dominant period (December 2021 - January 2022) | BA.1 (predominant) |
| **Favia 2023<sup>58</sup>** | Italy | Prospective cohort study | Single center | January 2022-March 2022 | Omicron-1 variant period (January-March 2022) | NR |
| **Finkas 2023<sup>59</sup>** | USA | Prospective cohort study | Healthcare database | March 2020-August 2022 | January-August 2022 | NR |
| **Flacco 2023<sup>60</sup>** | Italy | Retrospective cohort study | Healthcare database | February 2020-February 2023 | Omicron predominant period (1 January 2022-15 February 2023) | NR |
| **Flisiak 2023<sup>61</sup>** | Poland | Retrospective cohort study | Multi-center | January 2022-April 2023 | Early Omicron phase (January-June 2022); Late Omicron phase (July 2022-April 2023) | BA.1 and BA.2 (dominant in early period); BA.5, BQ.1, XBB, XBB.1.5, and XBB.1.9 (dominant in later period) |
| **Gazit 2022<sup>62</sup>** | Israel | Case-control study | Healthcare database | January-March 2022 | 10 January-13 March 2022 | NR |
| **Geng 2023<sup>12a</sup>** | China | Retrospective cohort study | Single center | April-May 2022 | 6 April-16 May 2022 | NR |
| **Grannec 2023<sup>63</sup>** | France | Retrospective cohort study | Single center | February -March 2022 | 1 February-7 March 2022 | NR |
| **Griggs 2024<sup>64</sup>** | USA | Retrospective cohort study | Healthcare database | June 2021-March 2023 | 16 December 2021-29 March 2023 | BA.1, BA.2, BA.4, BA.5, and Post-BA.4/BA.5 |
| **Guo 2023<sup>65</sup>** | China | Retrospective cohort study | Single center | March - May 2022 | 29 March - 17 May 2022 | NR |
| **Helmy 2023<sup>66</sup>** | Egypt | Prospective cohort study | Single center | NR | NR | NR |
| **Hippisley-Cox 2023<sup>67</sup>** | England | Retrospective cohort study | Healthcare database | December 2021-June 2022 | 11 December 2021-March 2022 | NR |
| **Jamaati 2023<sup>68</sup>** | Iran | Retrospective cohort study | Multi-center | January 2022-April 2022 | 21 January-11 April 2022 | NR |
| **Karageorgou 2023<sup>69</sup>** | Greece | Retrospective cohort study | Multi-center | November 2021-June 2022 | 16 January-8 June 2022 | NR |
| **Kim 2023<sup>70</sup>** | South Korea | Retrospective cohort study | Single center | November 2021-April 2022 | November 2021-April 2022 | NR |
| **Klein 2023<sup>71</sup>** | USA | Retrospective cohort study | Multi-center | September 2022-February 2023 | September 2022-February 2023 | XBB or XBB.1.5 |
| **Lee 2023<sup>72</sup>** | South Korea | Retrospective cohort study | Single center | February 2022-January 2023 | February 2022-January 2023 | BA.1, BA.5 |
| **Lewnard 2022<sup>73</sup>** | USA | Retrospective cohort study | Healthcare database | November 2021-March 2022 | 15 December 2021-17 March 2022 | BA.1, BA.2 |
| **Li 2024<sup>74</sup>** | China | Retrospective cohort study | Single center | April 2020-March 2023 | April-May 2022 | NR |
| **Li 2023<sup>75</sup>** | China | Cross-sectional study | Single center | December 2022-January 2023 | 8 December 2022-31 January 2023 | NR |
| **Liu 2023<sup>76</sup>** | China | Retrospective cohort study | Multi-center | 8 January-7 February 2022 | 8 January-7 February 2022 | NR |
| **Liu 2023a<sup>77</sup>** | China | Retrospective cohort study | Single center | December 2022-January 2023 | December 2022-January 2023 | NR |
| **Lu 2022<sup>78</sup>** | China | Cross-sectional study | Multi-center | April-May 2022 | April-May 2022 | NR |
| **Manchanda 2023<sup>79</sup>** | India | Retrospective cohort study | Single center | April 2021-January 2022 | January 2022 | B.1.1.529 |
| **Mayer 2023<sup>80</sup>** | Germany | Retrospective cohort study | Single center | December 2021-August 2022 | 20 December 2021-15 August 2022 | NR |
| **McNeil 2023<sup>81</sup>** | Australia | Retrospective cohort study | Multi center | June 2022-November 2022 | June-November 2022 | NR |
| **Mendoza-Cano 2023<sup>82</sup>** | Mexico | Retrospective cohort study | Healthcare database | May 2023-August 2023 | May-August 2023 | NR |
| **Mizrahi Reuveni 2023<sup>83</sup>** | Israel | Retrospective cohort study | Healthcare database | November 2021-January 2022 | 26 November 2021-18 January 2022 | NR |
| **Morris 2023<sup>14a</sup>** | USA | Retrospective cohort study | Multi-center | December 2021-July 2022 | 1 December 2021-17 July 2022 | BA.1, BA.1.1, BA.2, BA.2.12.1<br>BA.4, BA.5 |
| **Mosallami Aghili 2022<sup>84</sup>** | Iran | Prospective cohort study | Single center | January 2022-July 2022 | January-July 2022 | NR |
| **Mumtaz 2022<sup>85</sup>** | Pakistan | Retrospective cohort study | Multi-center | January- April 2022 | 16 January-18 April 2022 | NR |
| **Nab 2023<sup>1</sup>** | England | Retrospective cohort study | Healthcare database | March 2020-August 2022 | 15 December 2021-3 August 2022 | B.1.1.529 |
| **O'Leary 2023<sup>86</sup>** | USA | Retrospective cohort study | Healthcare database | December 2021-February 2022 | 25 December 2021-2 February 2022 | NR |
| **Parajuli 2023<sup>87</sup>** | USA | Retrospective cohort study | Healthcare database | April 2020 - June 2022 | 20 December 2021 - 25 June 2022 | NR |
| **Parra-Bracamonte 2023<sup>88</sup>** | Mexico | Retrospective cohort study | Healthcare database | January 2020-October 2022 | 24 December 2021-24 October 2022 | NR |
| **Patton 2023<sup>89</sup>** | USA | Retrospective cohort study | Multi-center | January 2020-March 2022 | February-March 2022 | NR |
| **Radhakrishnan 2023<sup>90</sup>** | USA | Retrospective cohort study | Single center | December 2021-February 2022 | 8 January-15 February 2022 | BA.1 |
| **Russell 2022<sup>91</sup>** | Canada | Retrospective cohort study | Multi-center | December 2021-August 2022 | 23 December 2021-31 August 2022 (confirmed by whole genome sequencing) | BA.1, BA.2, or BA.5 |
| **Sardinha 2023<sup>92</sup>** | Brazil | Retrospective cohort study | Healthcare database | January 2022-February 2023 | January-December 2022 | NR |
| **Shakor 2023<sup>13a</sup>** | Malaysia | Cross-sectional study | Healthcare database | February 2022-March 2023 | 14 February 2022-31 March 2023 | NR |
| **Shi 2023<sup>93</sup>** | South Korea | Retrospective cohort study | Multi-center | February 2022-April 2022 | February-April 2022 | NR |
| **Simmons 2023<sup>94</sup>** | Canada | Retrospective cohort study | Healthcare database | May 2021-January 2022 | 23 December 2021-9 January 2022 | NR |
| **Skarbinski 2022<sup>95</sup>** | USA | Retrospective cohort study | Healthcare database | July 2021-January 2022 | December 18, 2021-January 7, 2022 | B.1.1.529 |
| **Sonaglioni 2022<sup>96</sup>** | Italy | Retrospective cohort study | Single center | January 2022-May 2022 | 1 January-15 May 2022 | NR |
| **Stepanova 2022<sup>97</sup>** | USA | Retrospective cohort study | Multi-center | March 2020-February 2022 | December 2021-February 2022 | NR |
| **Tsujimoto 2023<sup>98</sup>** | Japan | Retrospective cohort study | Multi-center | January 2022-March 2022 | January-March 2022 | BA.2 |
| **Vo 2022<sup>99</sup>** | USA | Retrospective cohort study | Healthcare database | December 2020-February 2022 | 16 December 2021-8 February 2023 | NR |
| **Wang 2023<sup>100</sup>** | USA | Retrospective cohort study | Healthcare database | October 2021-January 2022 | 19 December 2021-31 January 2022 | NR |
| **Ward 2024<sup>101</sup>** | England | Retrospective cohort study | Healthcare database | September 2022-April 2023 | 1 September 2022-11 April 2023 | NR |
| **Xin 2024<sup>102</sup>** | China | Retrospective cohort study | Single center | December 2022-March 2023 | 5 December 2022-31 March 2023 | NR |
| **Yang 2023<sup>103</sup>** | China | Retrospective cohort study | Single center | December 2022-January 2023 | 19 December 2022-5 January 2023 | BF.7 |
| **Ying-hao 2022<sup>104</sup>** | China | Retrospective cohort study | Single center | April 2022-May 2022 | 5 April-8 May 2022 | NR |
| **Zhang 2022<sup>105</sup>** | China | Retrospective cohort study | Single center | March 2022-May 2022 | 23 March-26 May 2022 | NR |
| **Zhao 2023<sup>106</sup>** | China | Retrospective cohort study | Single center | December 2022-February 2023 | 20 December 2022-31 January 2023 | NR |
| **Zhu 2023<sup>107</sup>** | China | Cross-sectional study | Multi-center | February 2022-May 2022 | 20 February-10 May 2022 | BA.2 |

Abbreviations: BCDT, B-cell depletion therapy; BID, brought-in-dead; CeVD, pre-existing cerebrovascular disease; CKD, chronic kidney disease; CVD, cardiovascular disease; GP, general population; IPD, in-patient death; IQR, inter quartile range; NA, not applicable; NR, not reported; SD, standard deviation; SGTF, S gene target failure; USA, United States of America; UK, United Kingdom.
<sup>a</sup> Not included in meta-analysis.

### Patient Characteristics

Patient characteristics are summarized in **Table 2**. Minimum numbers of participants included in analyses for each comorbidity across the included studies are reported in **Table 3**. Most studies (n = 42) included a ‘Hospitalized Population’, and the rest (n = 30) included ‘General Population’. These populations included individuals who were and were not hospitalized at the start of the study, respectively (**Table 2**). Diabetes was the most commonly reported comorbidity (n = 50 studies; **Table 3**) and thrombosis the least commonly reported (n = 4 studies). The majority of studies (>50%) that reported vaccination status included fully vaccinated individuals.

**Table 2.**
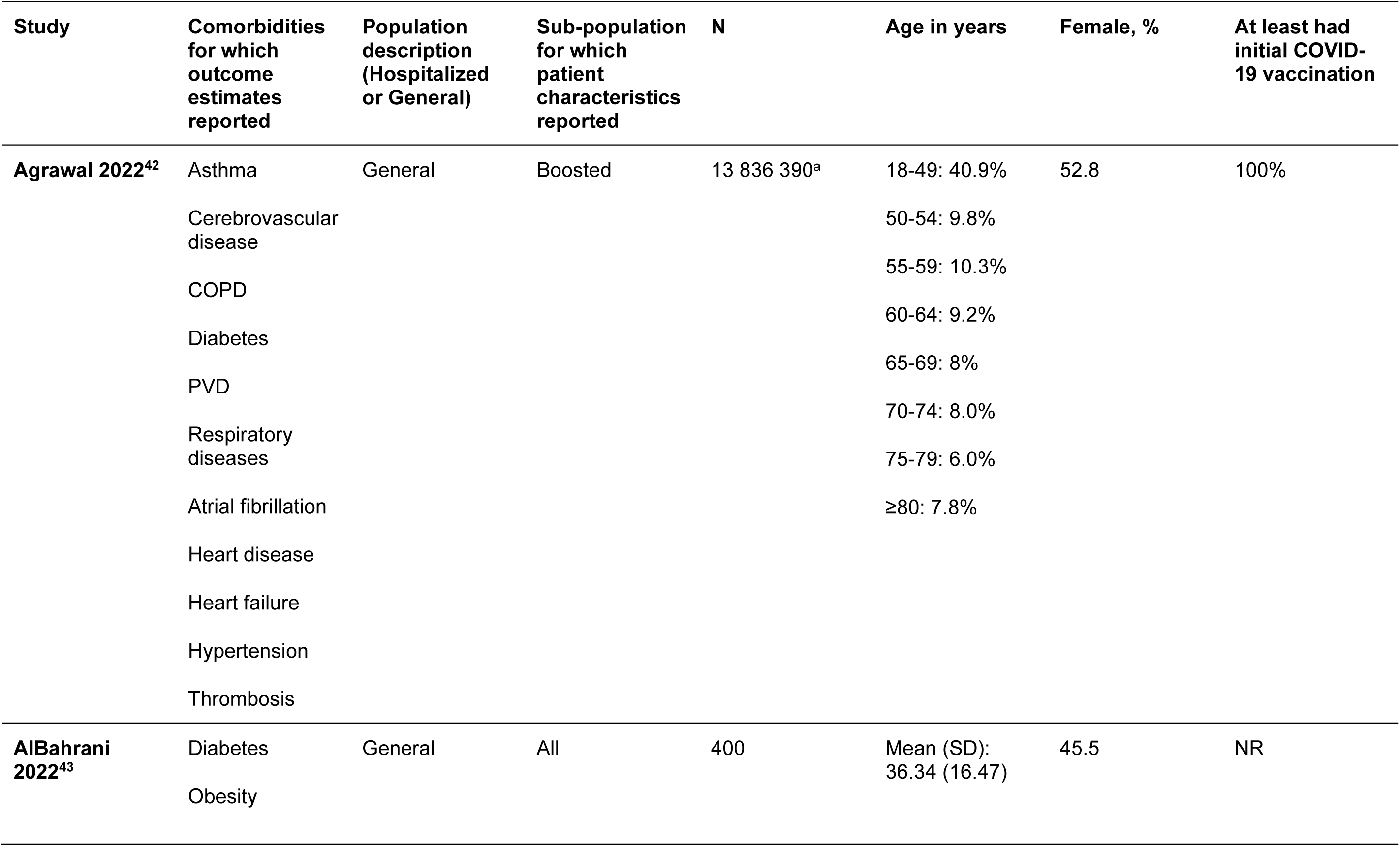
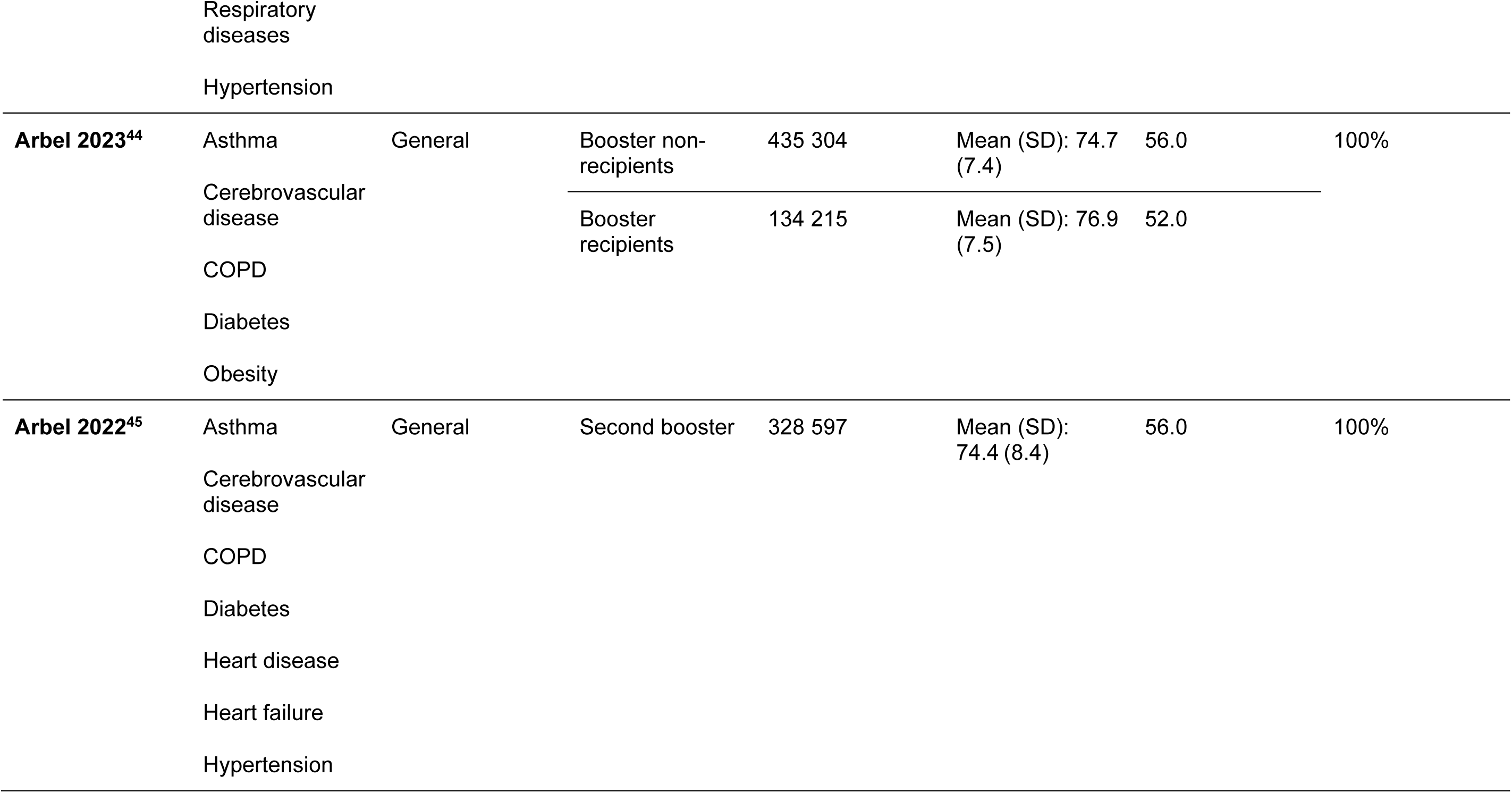
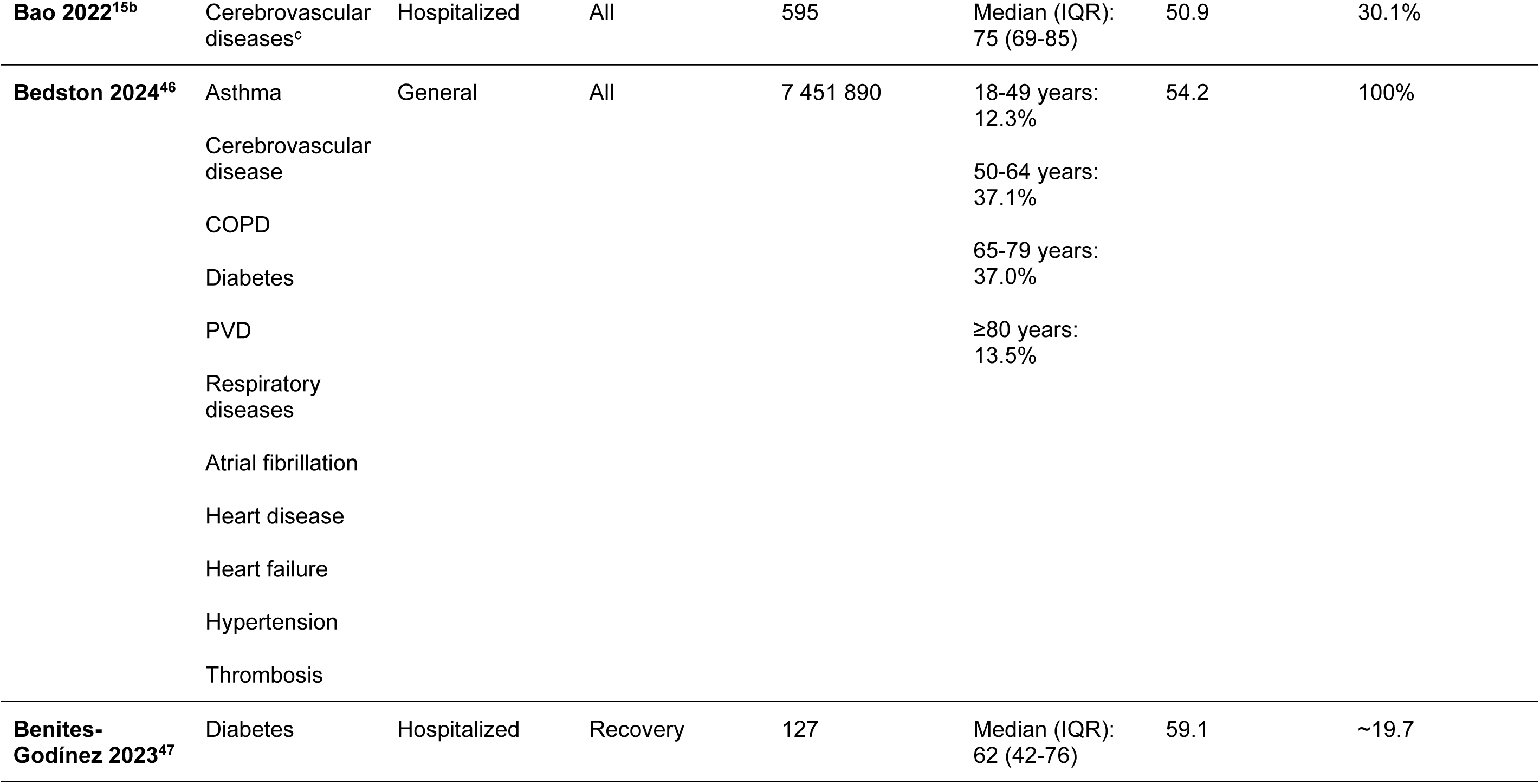
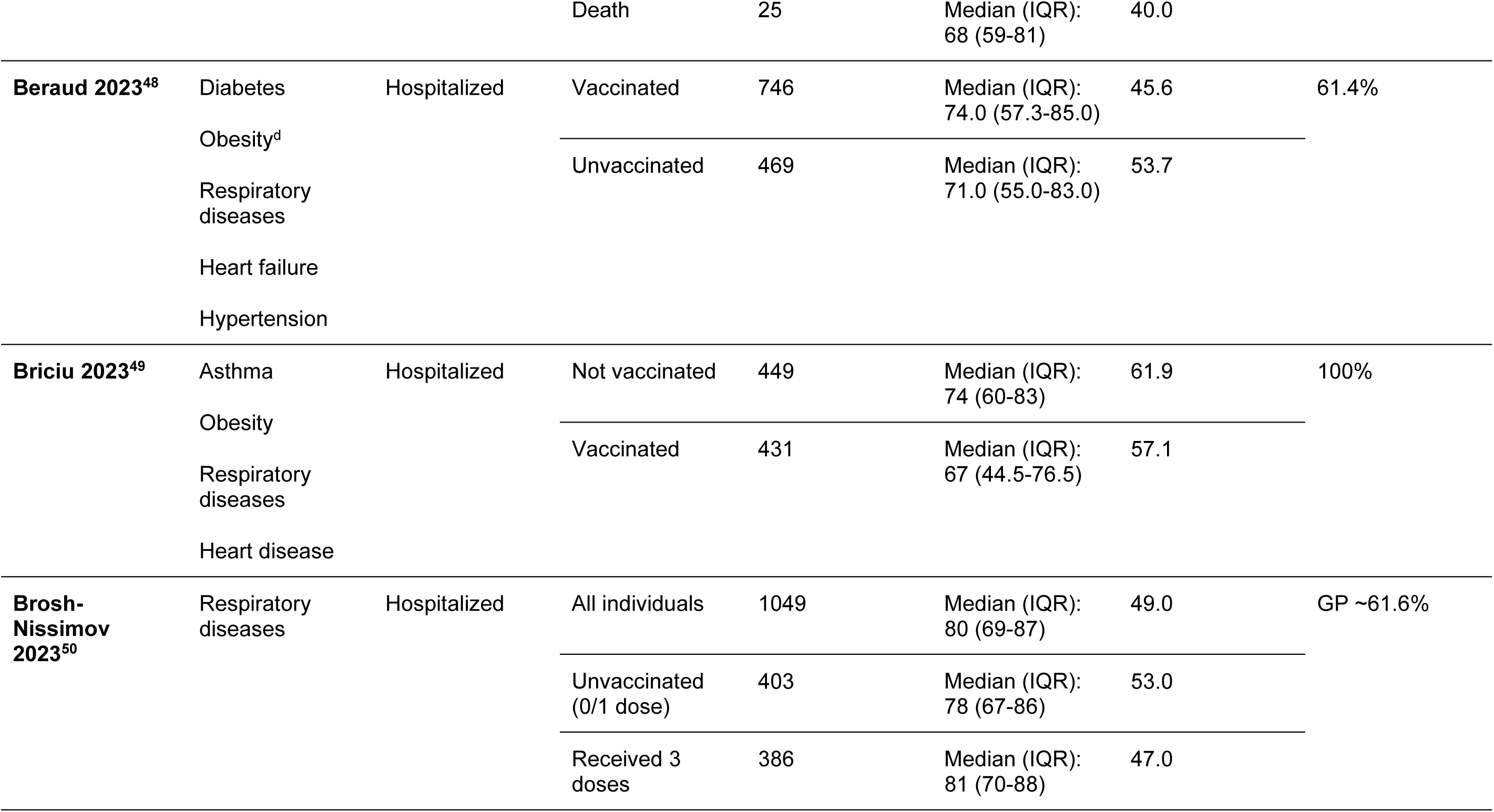
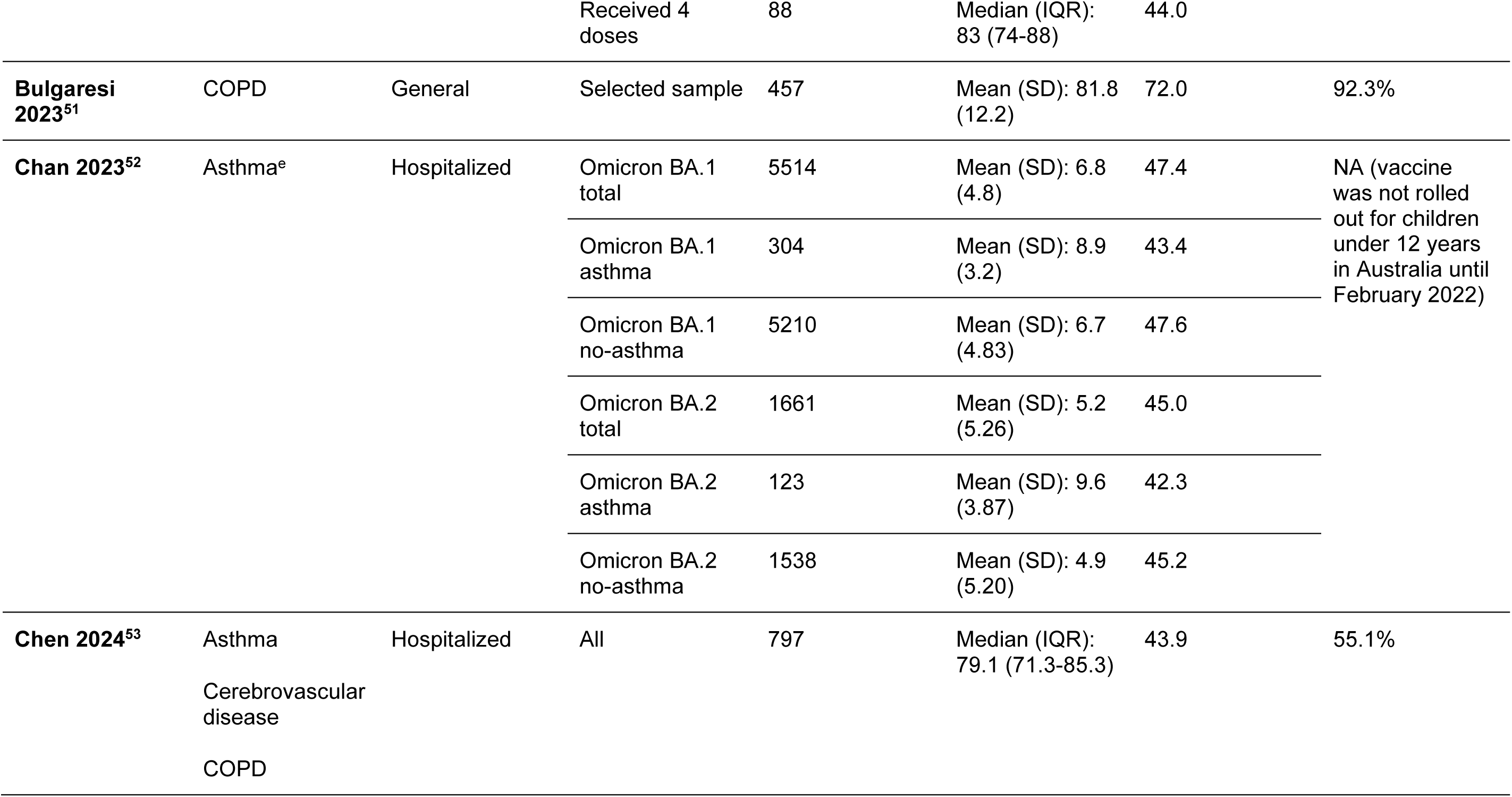
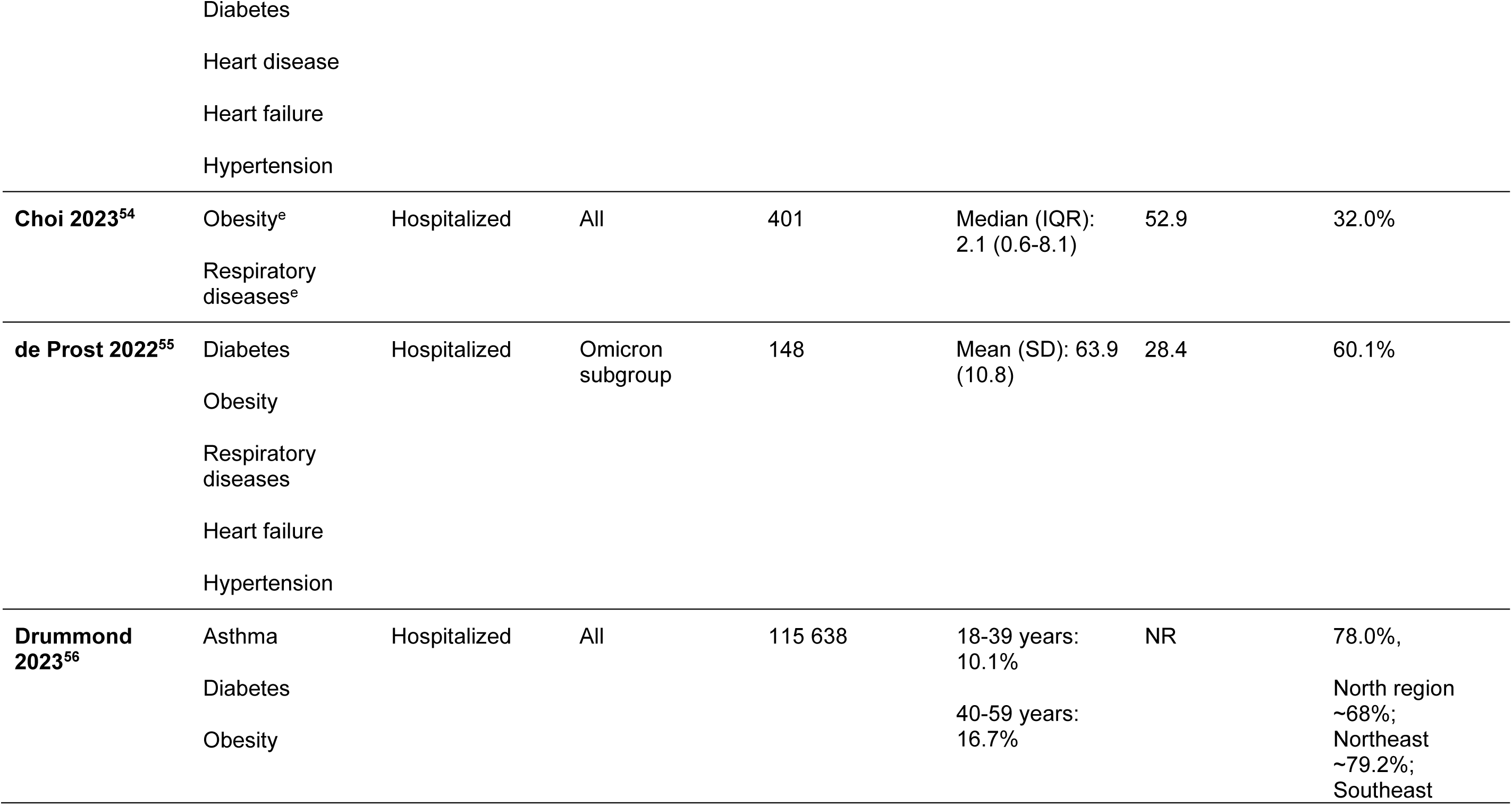
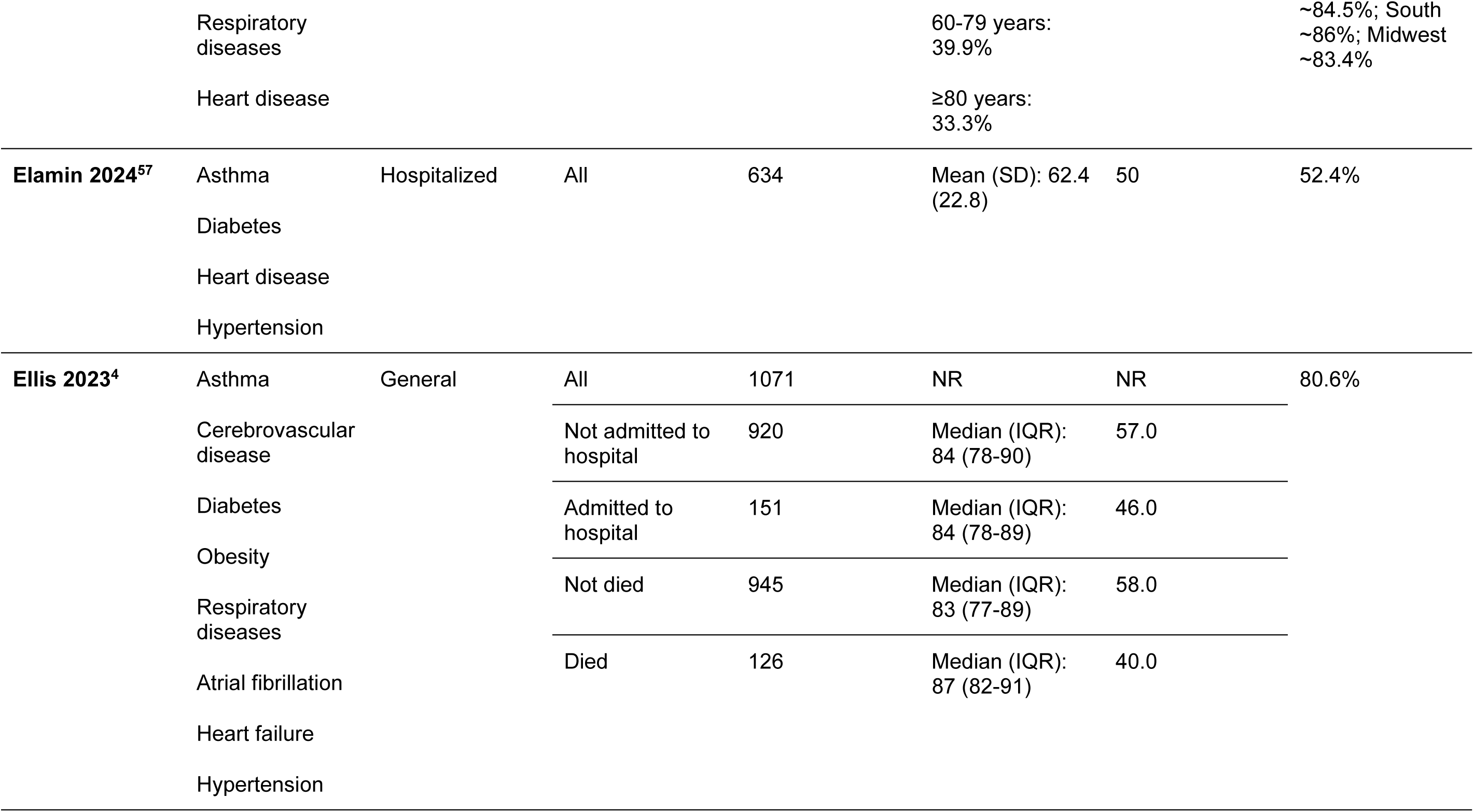
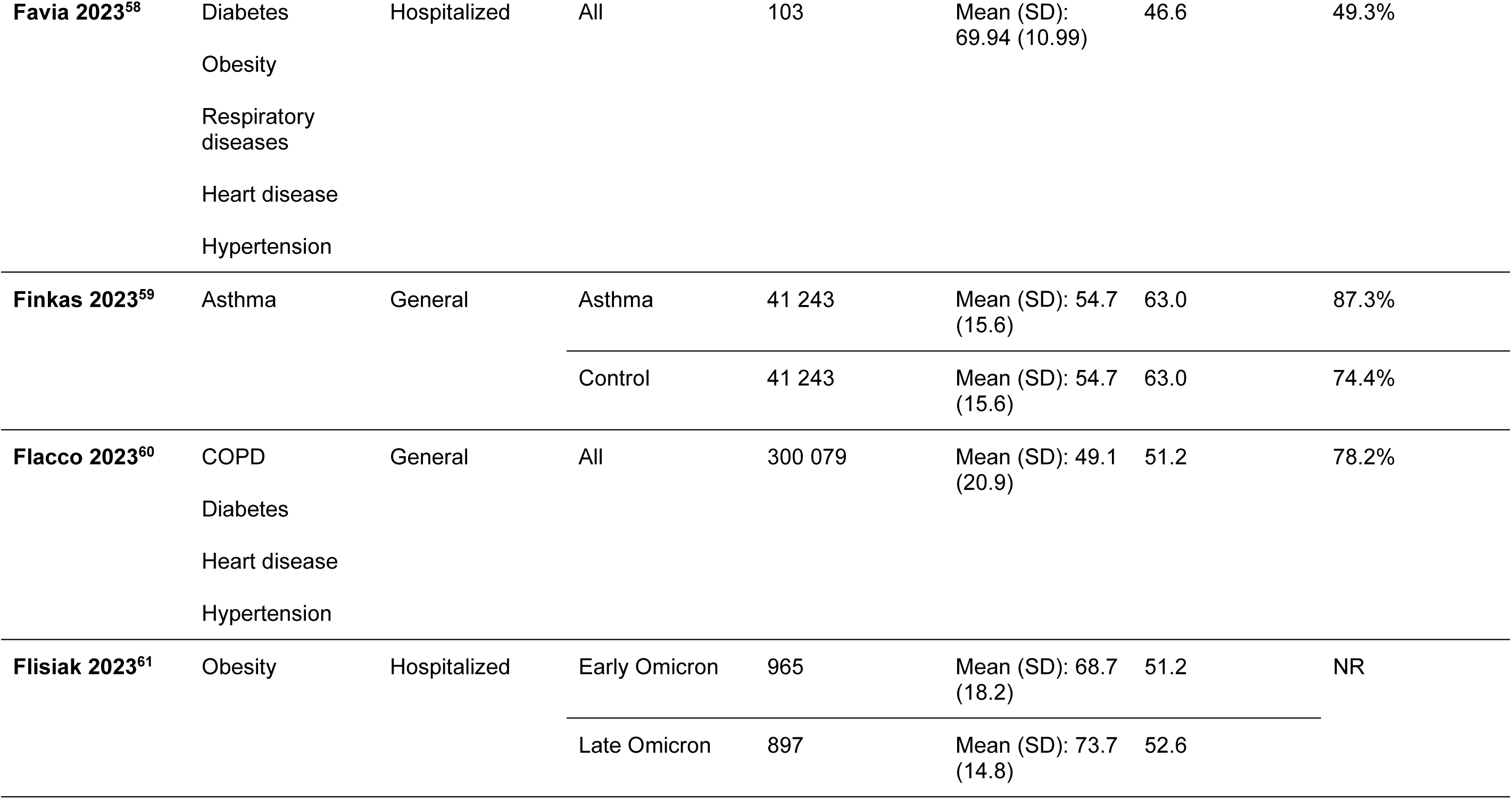
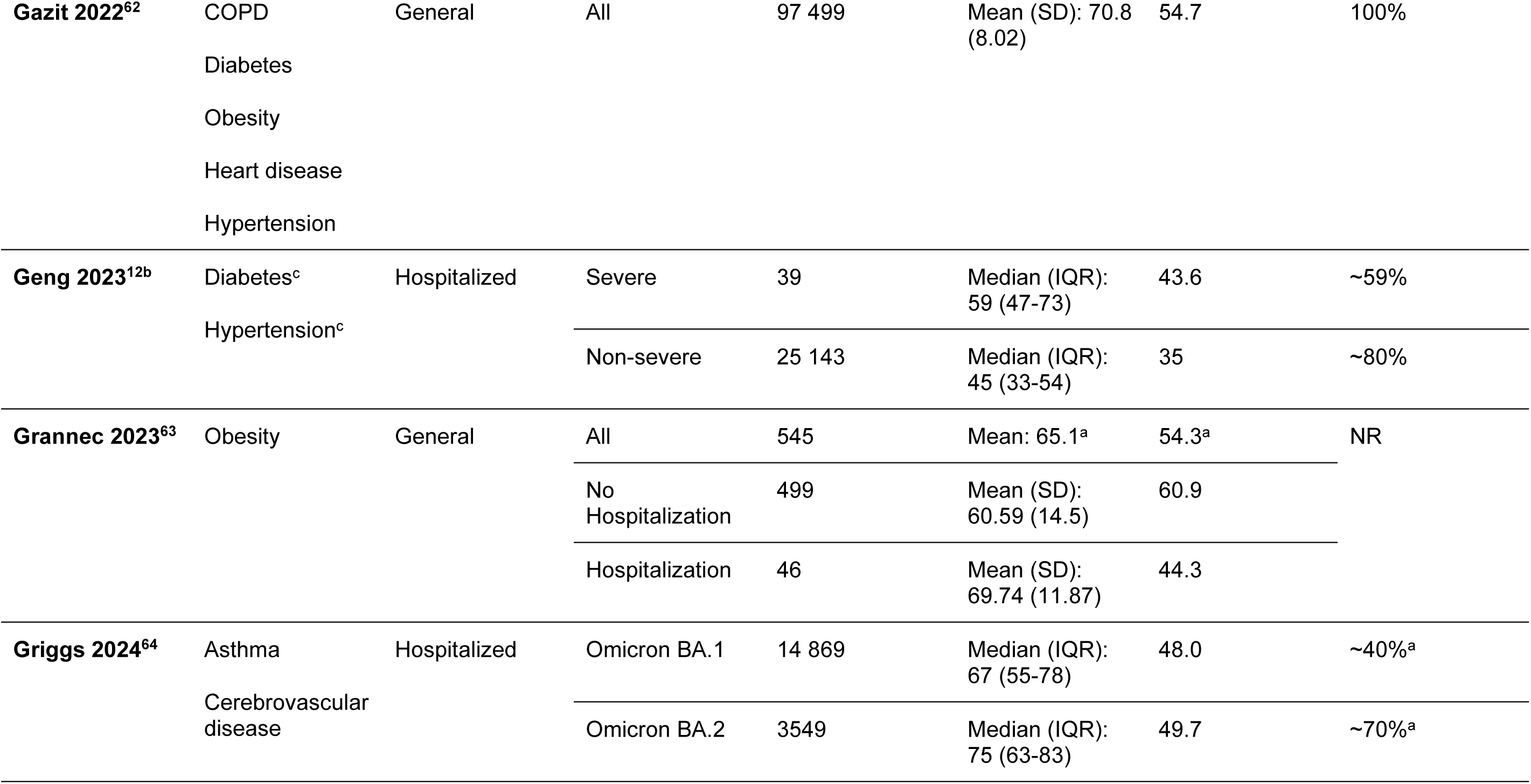
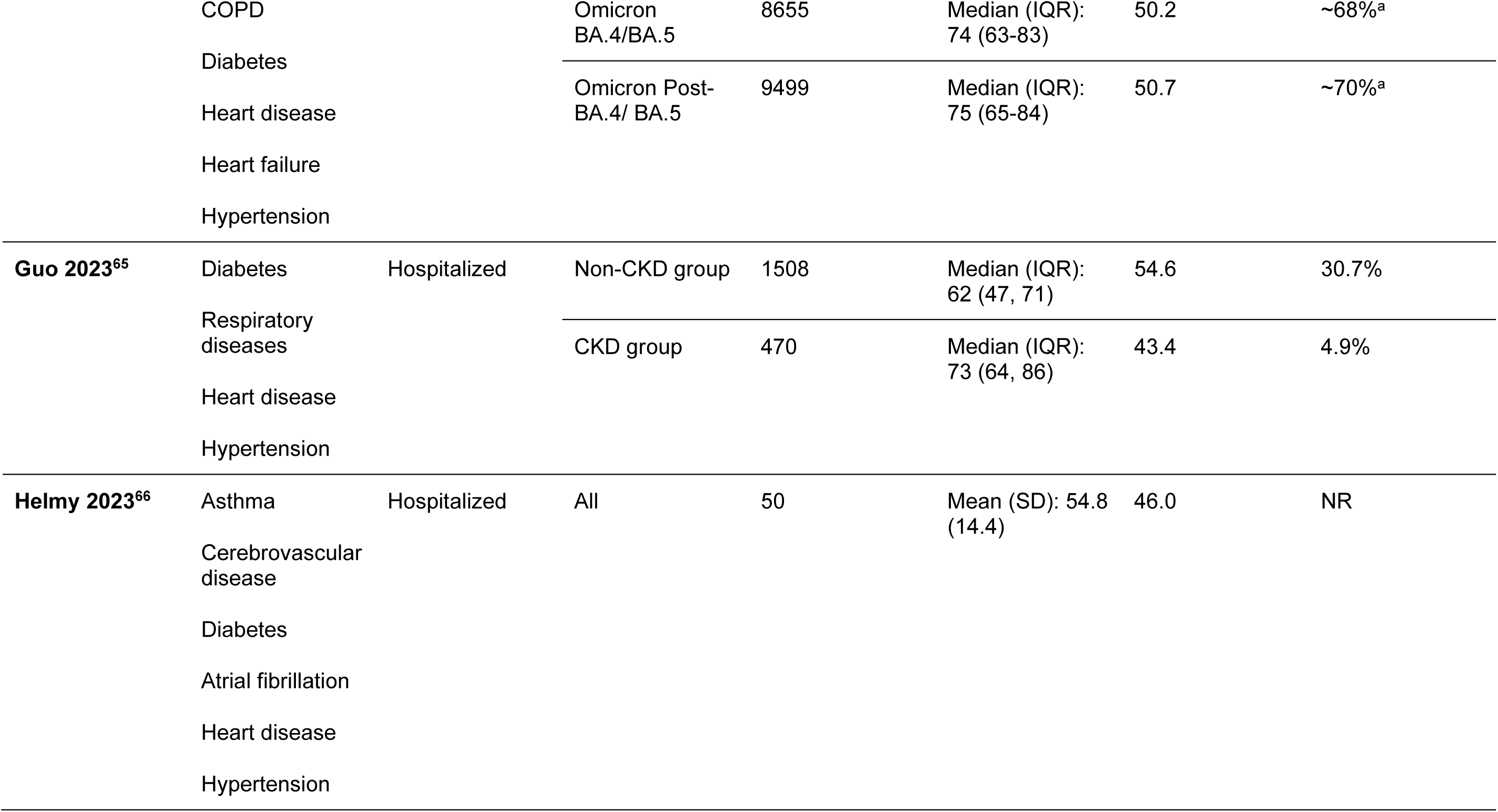
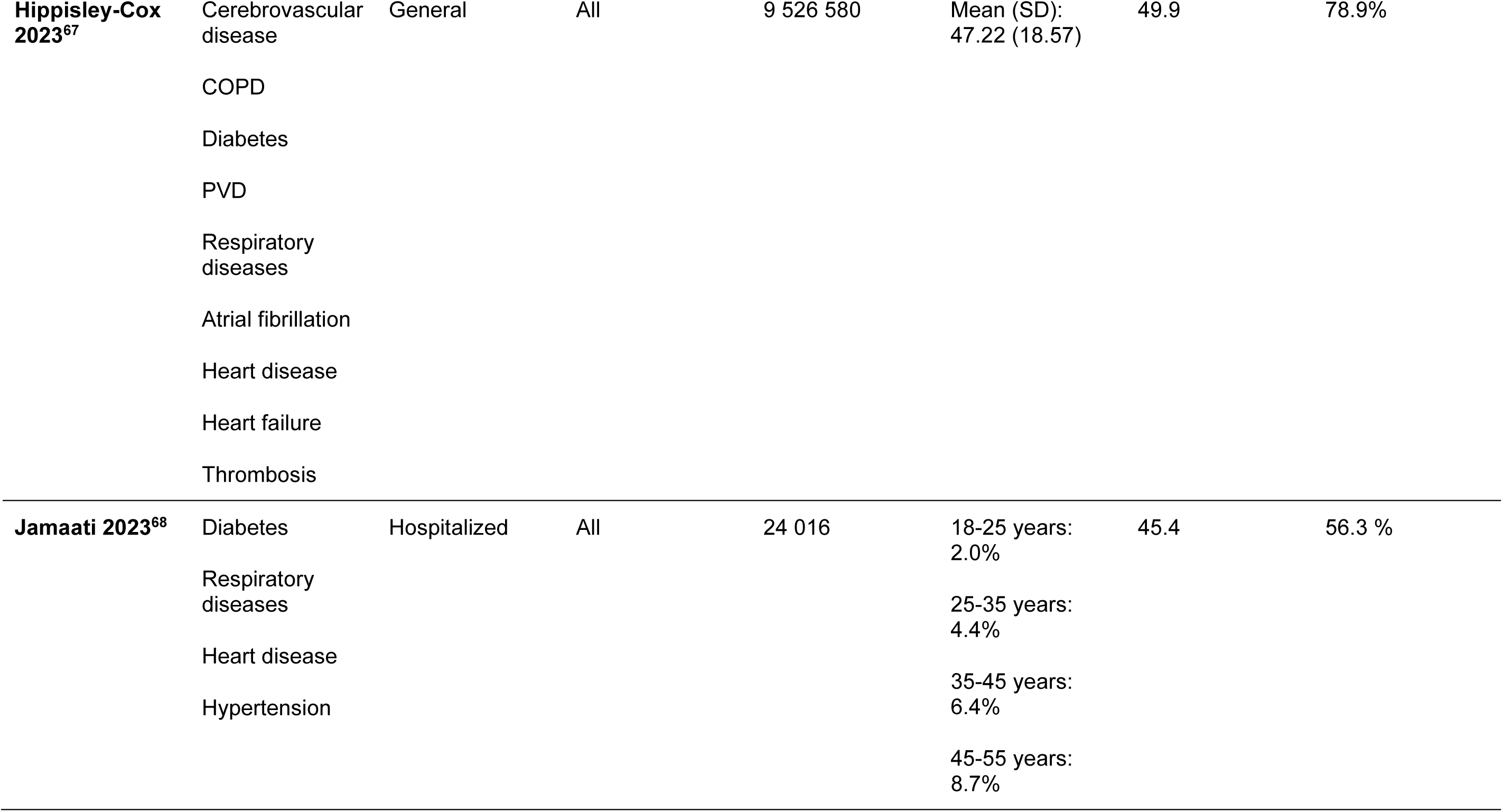
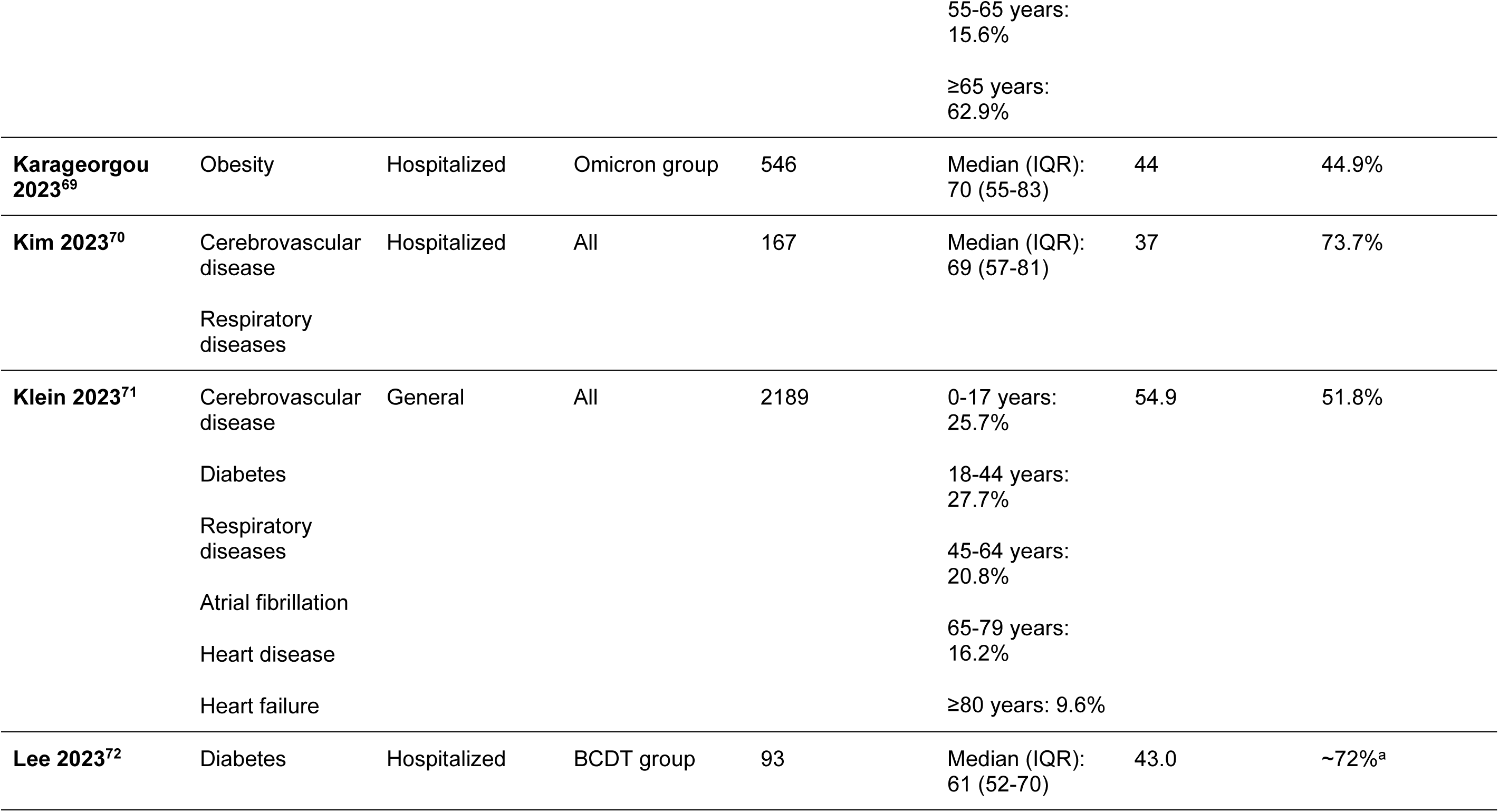
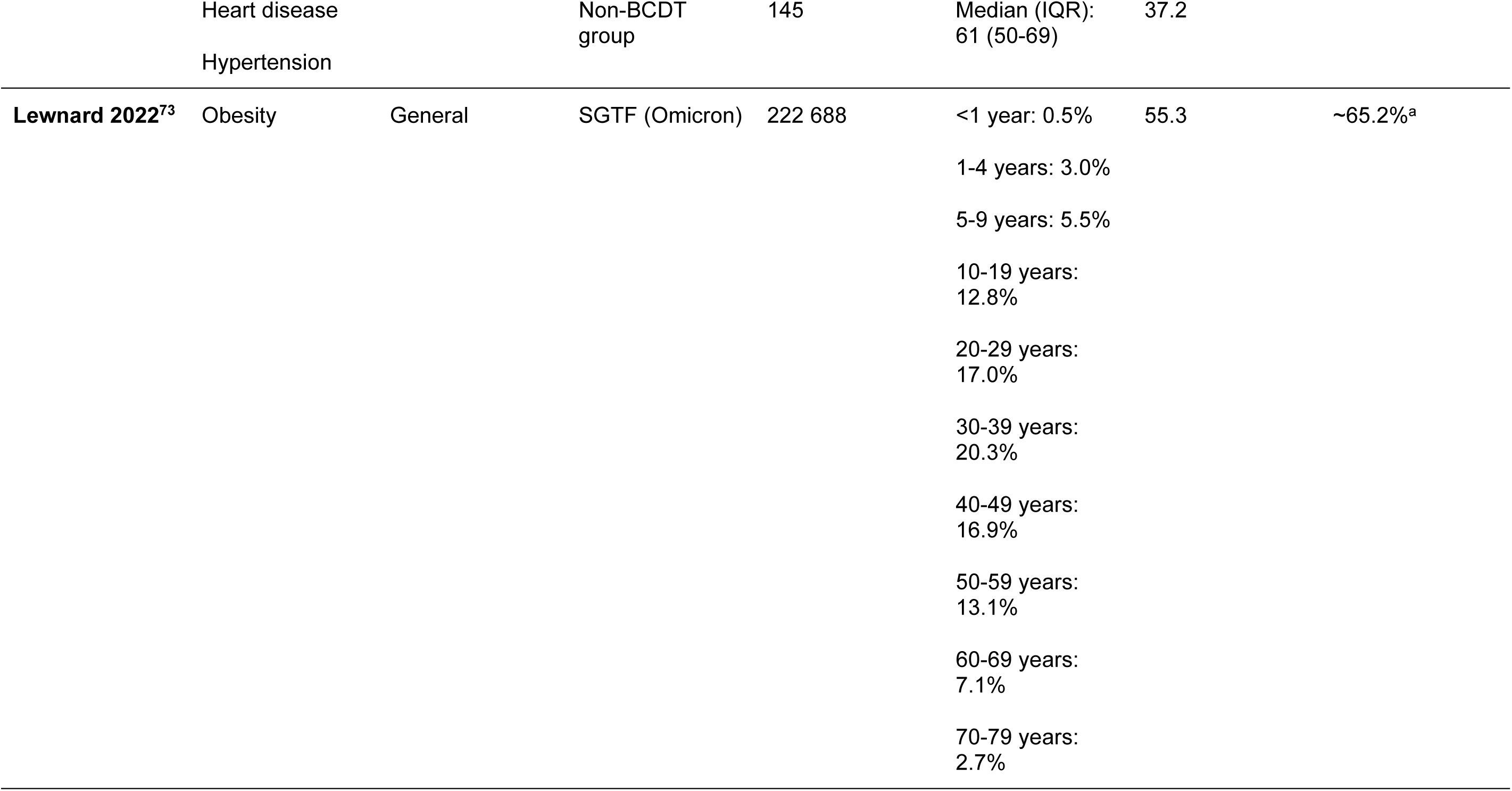
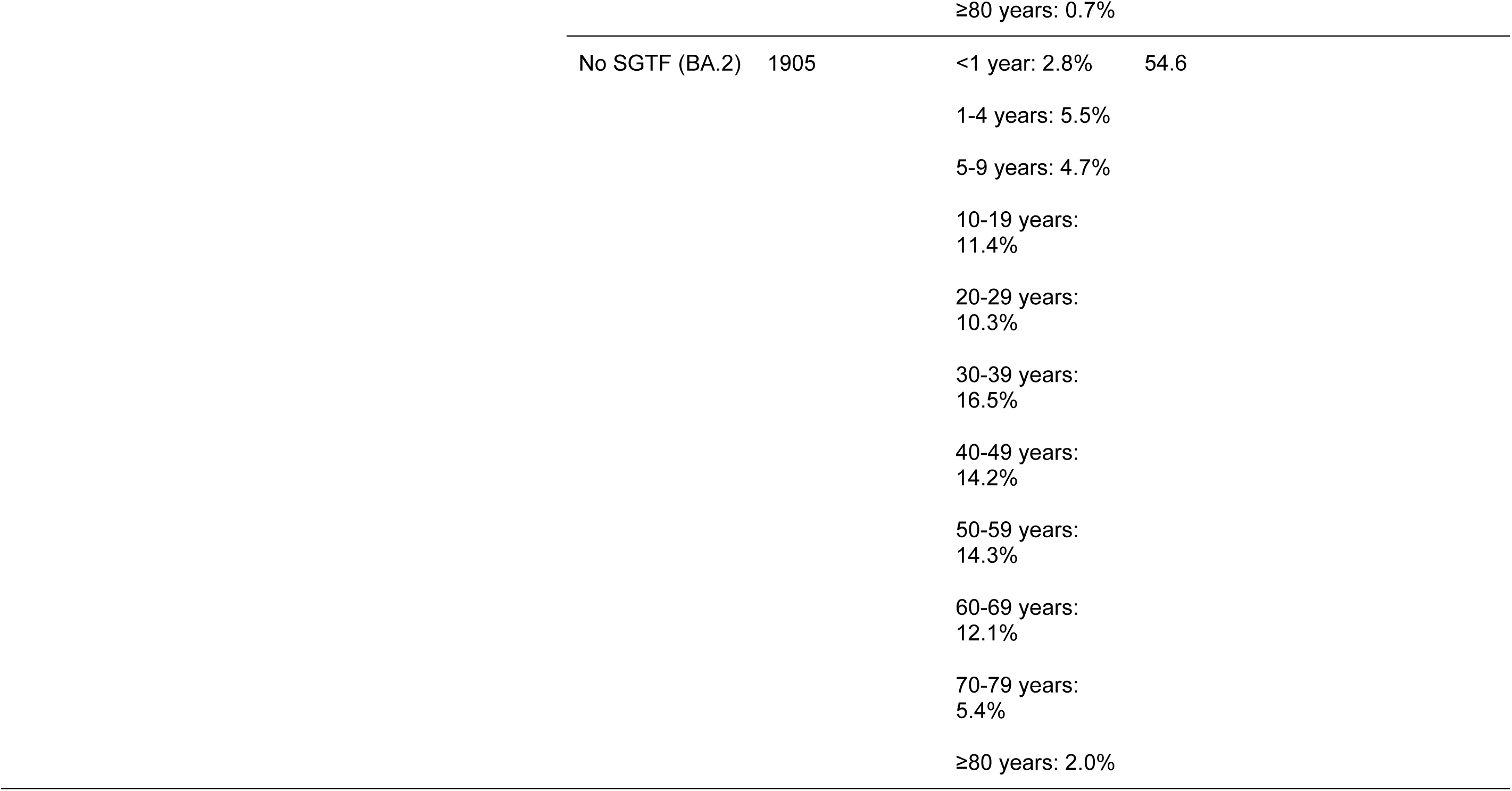
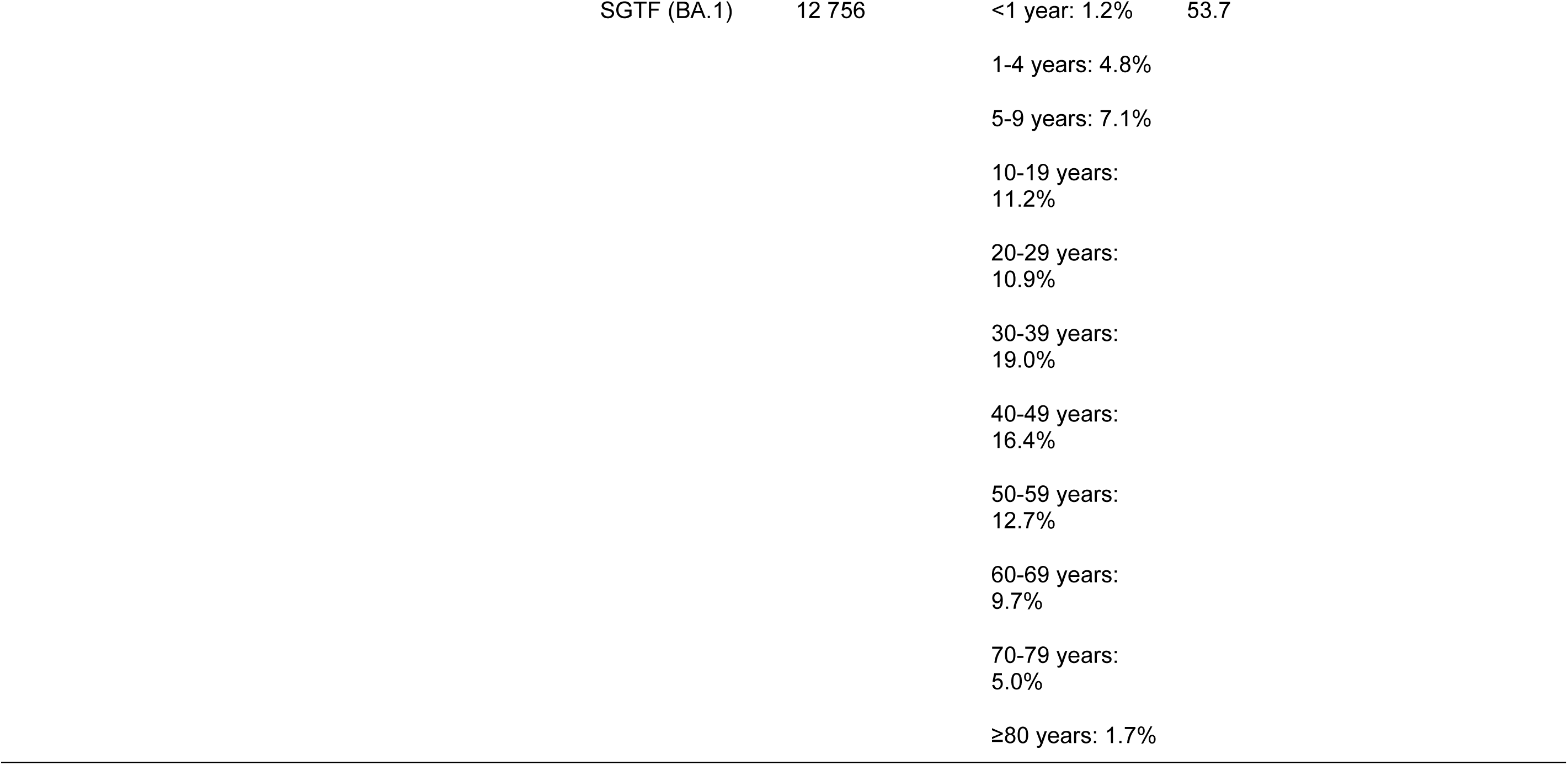
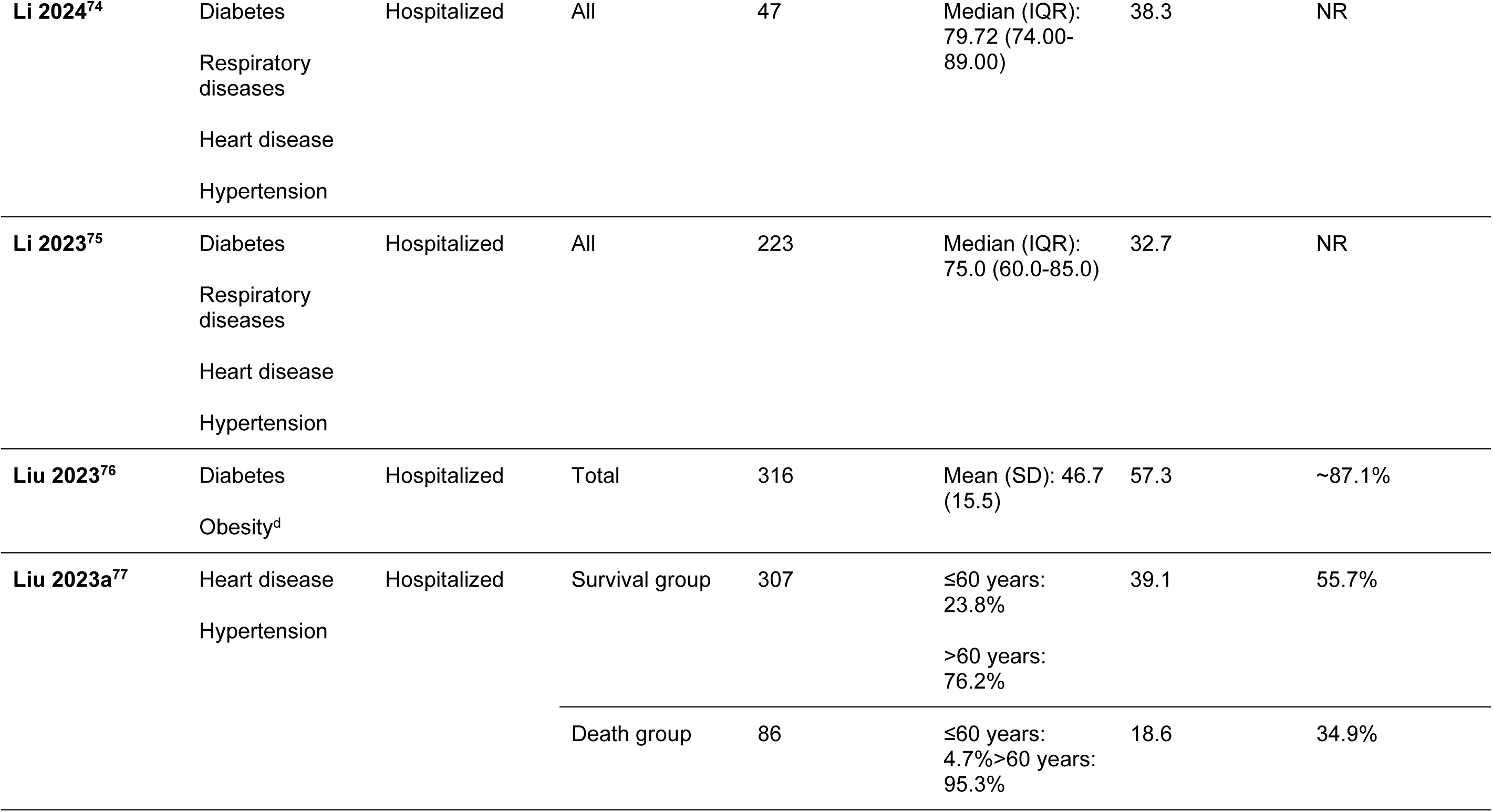
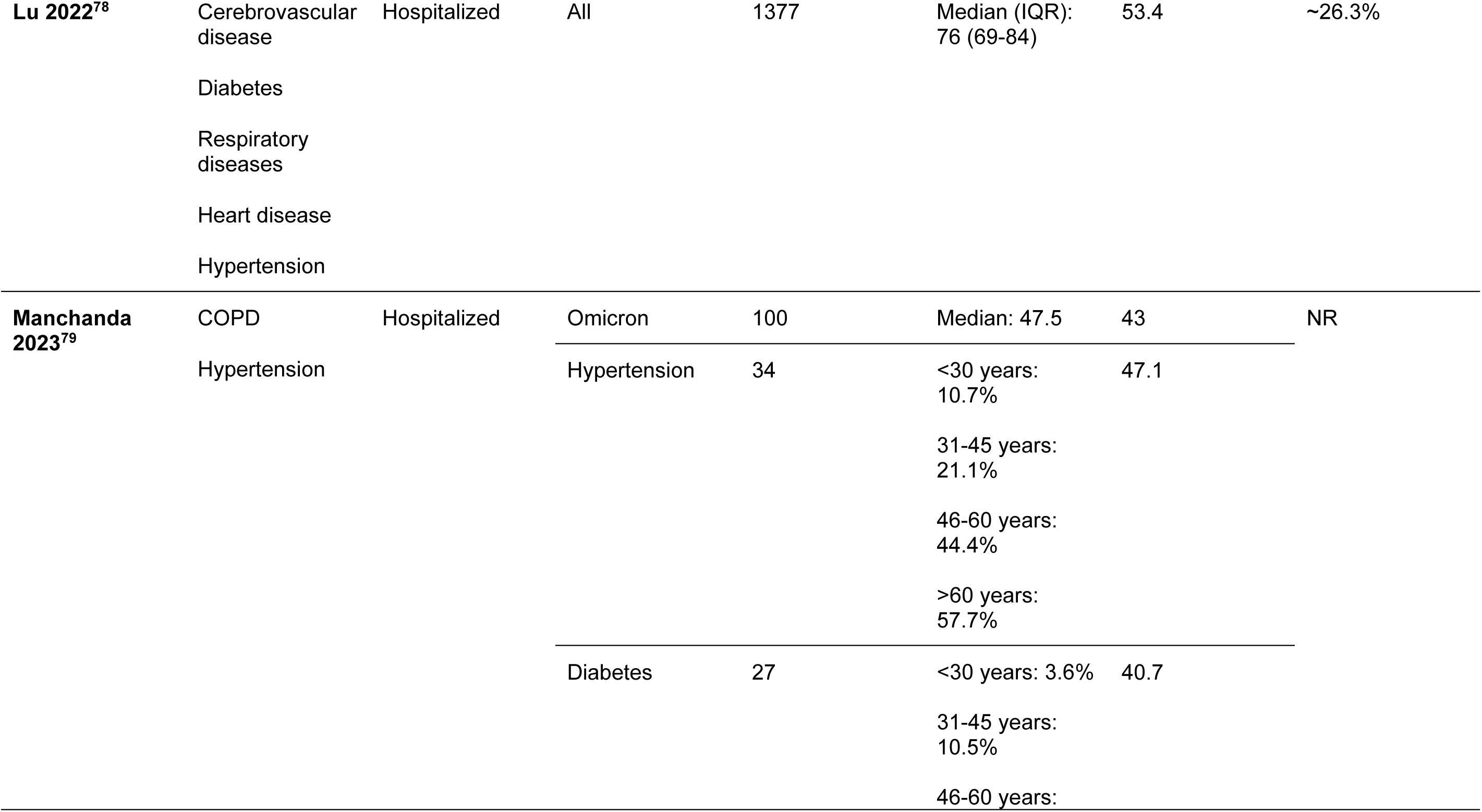
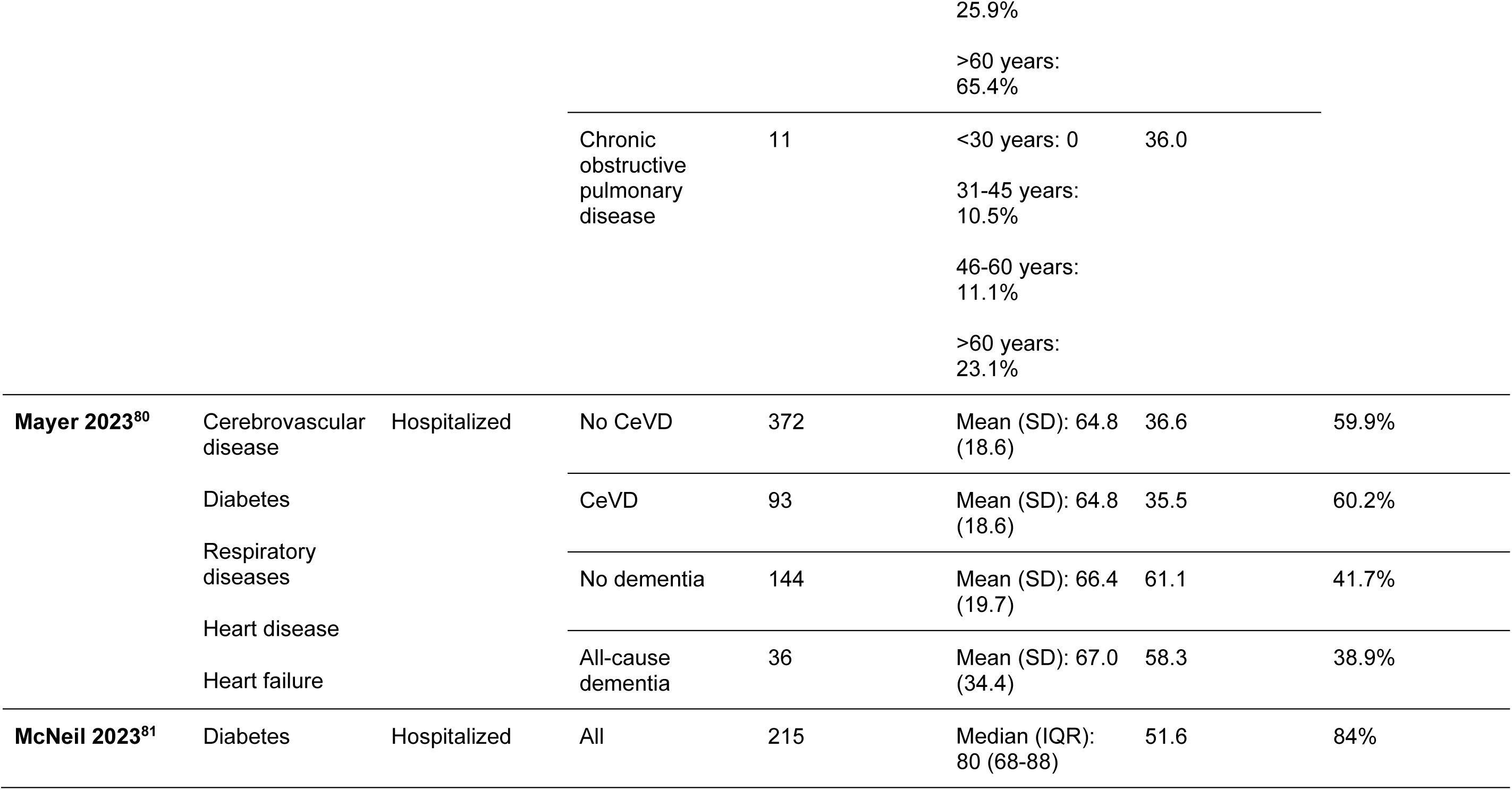
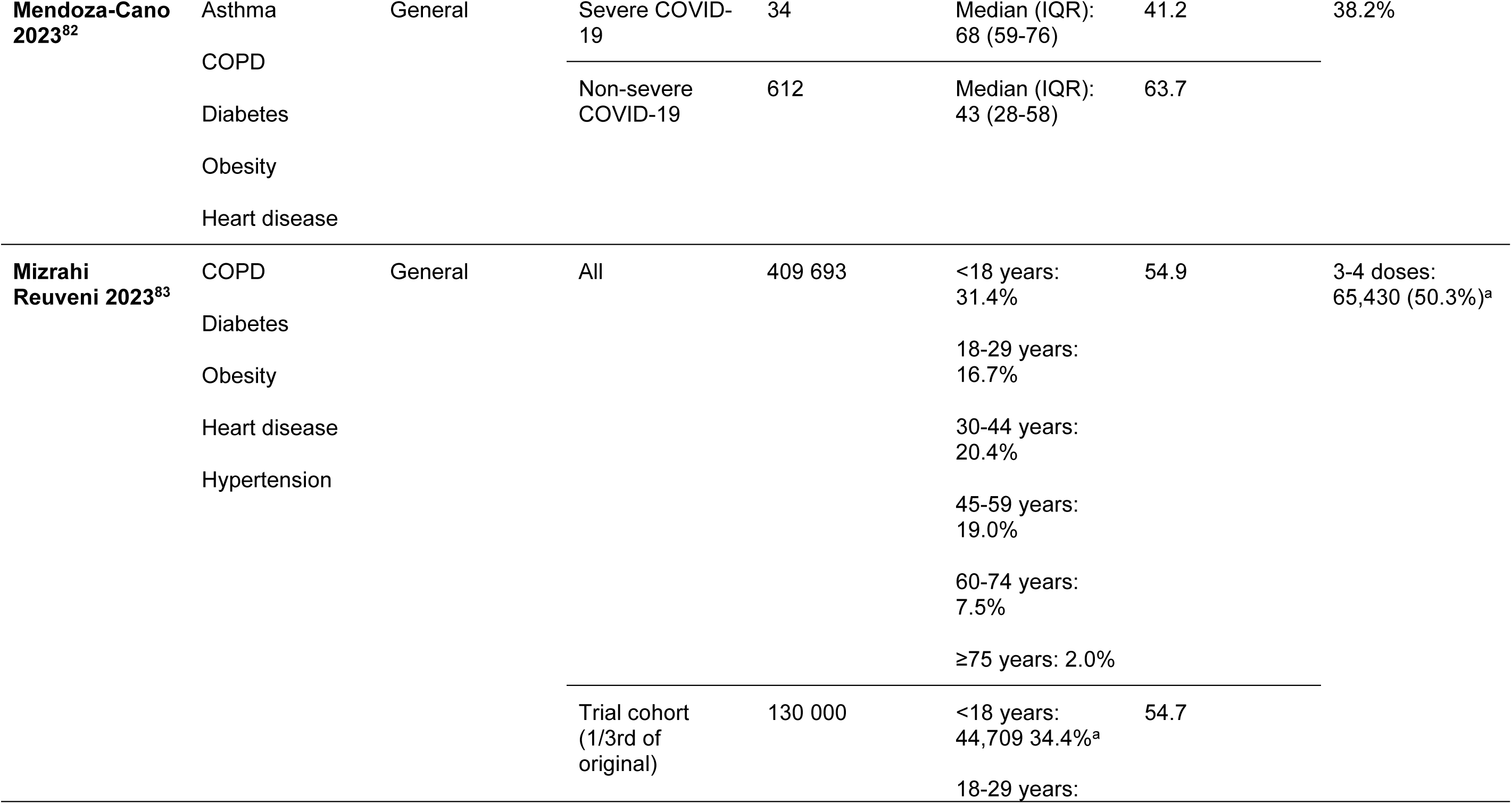
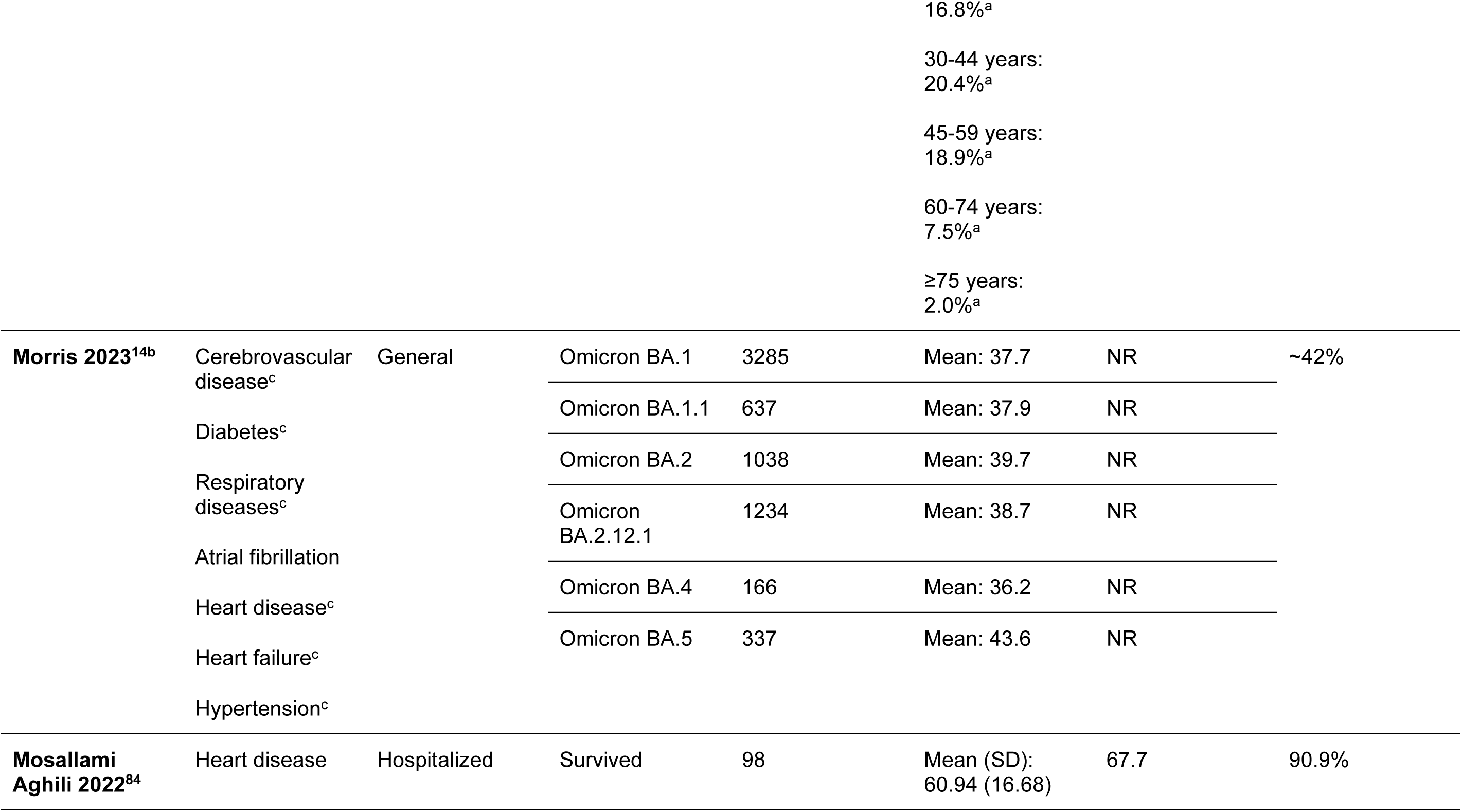
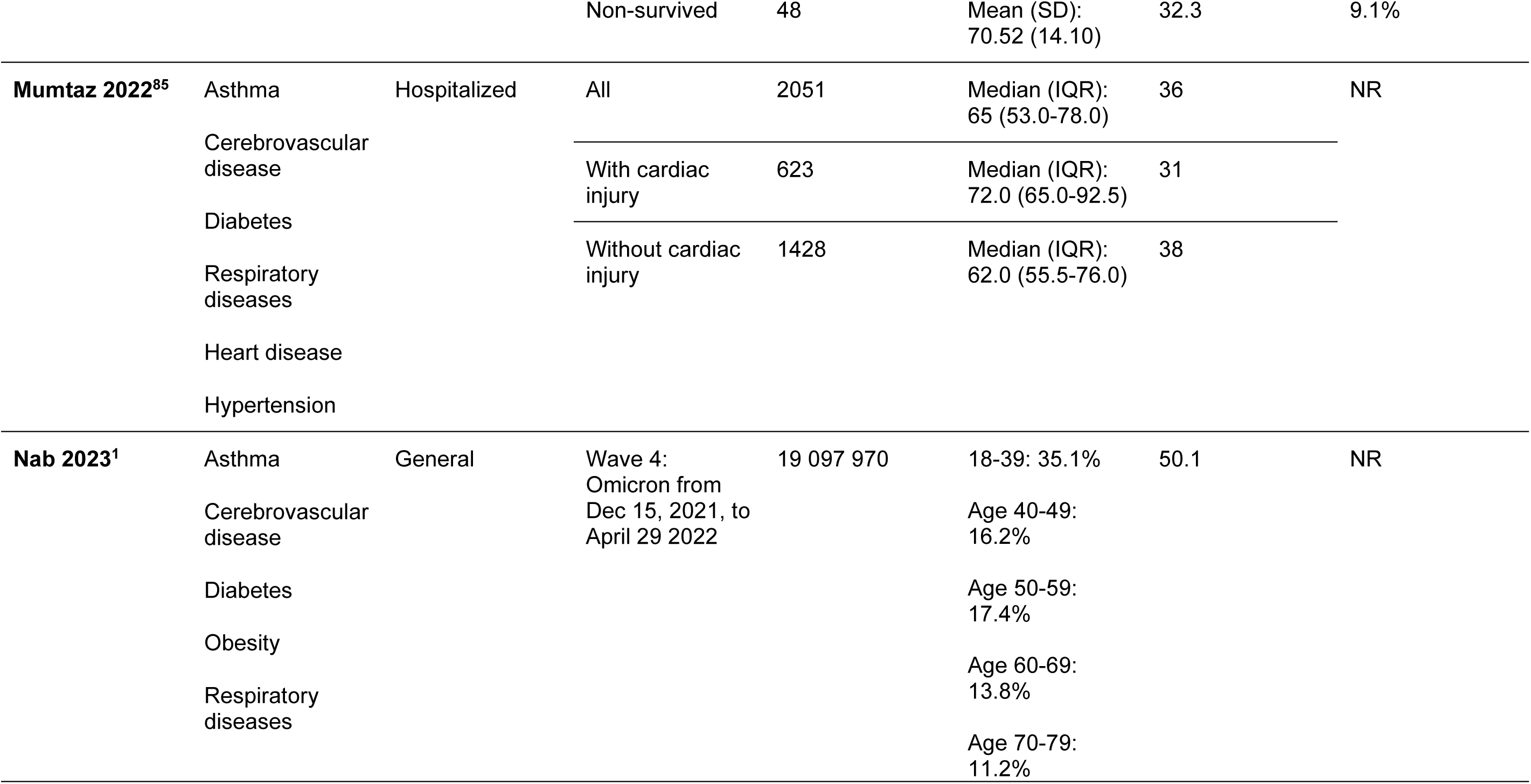
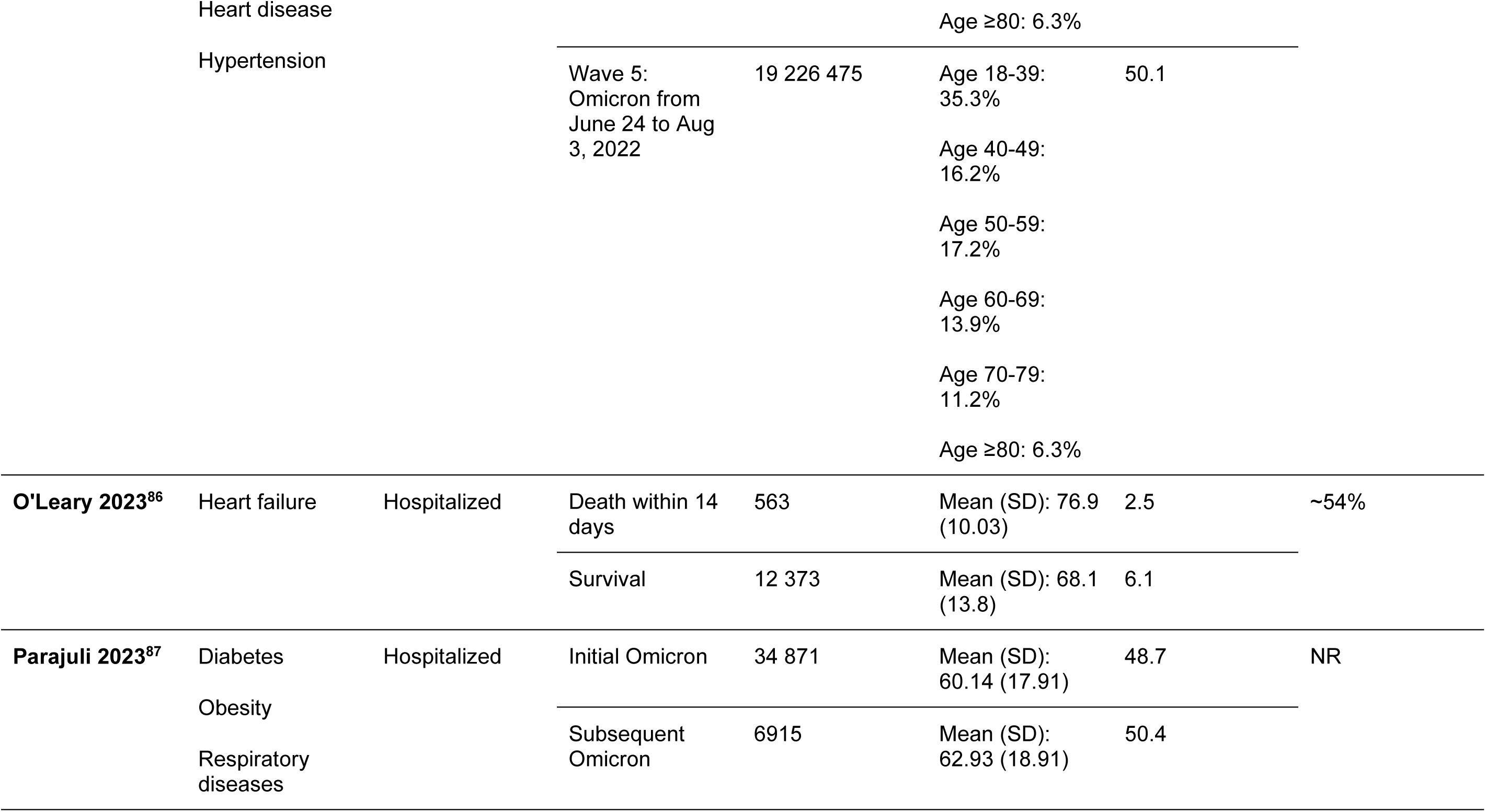
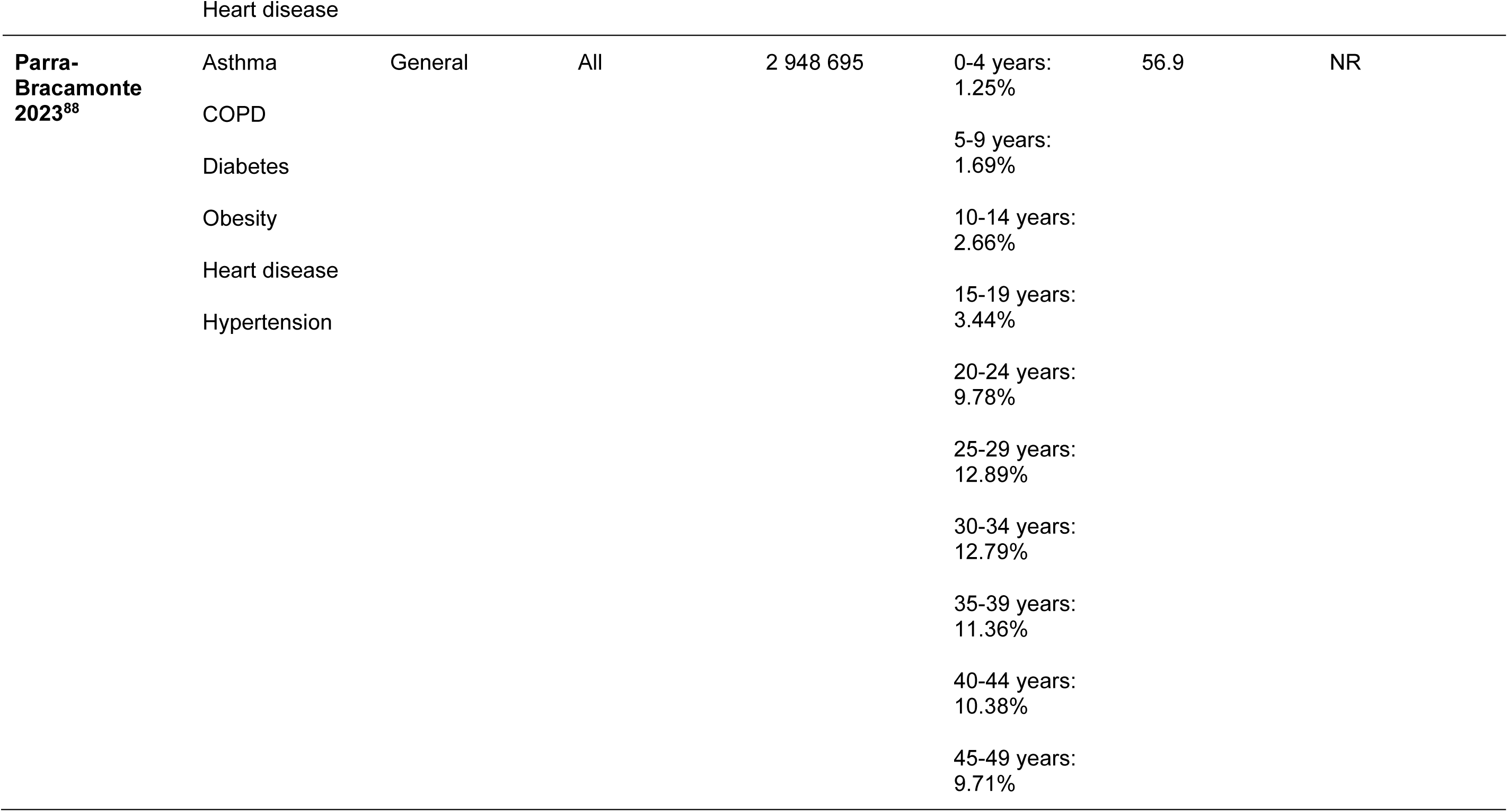
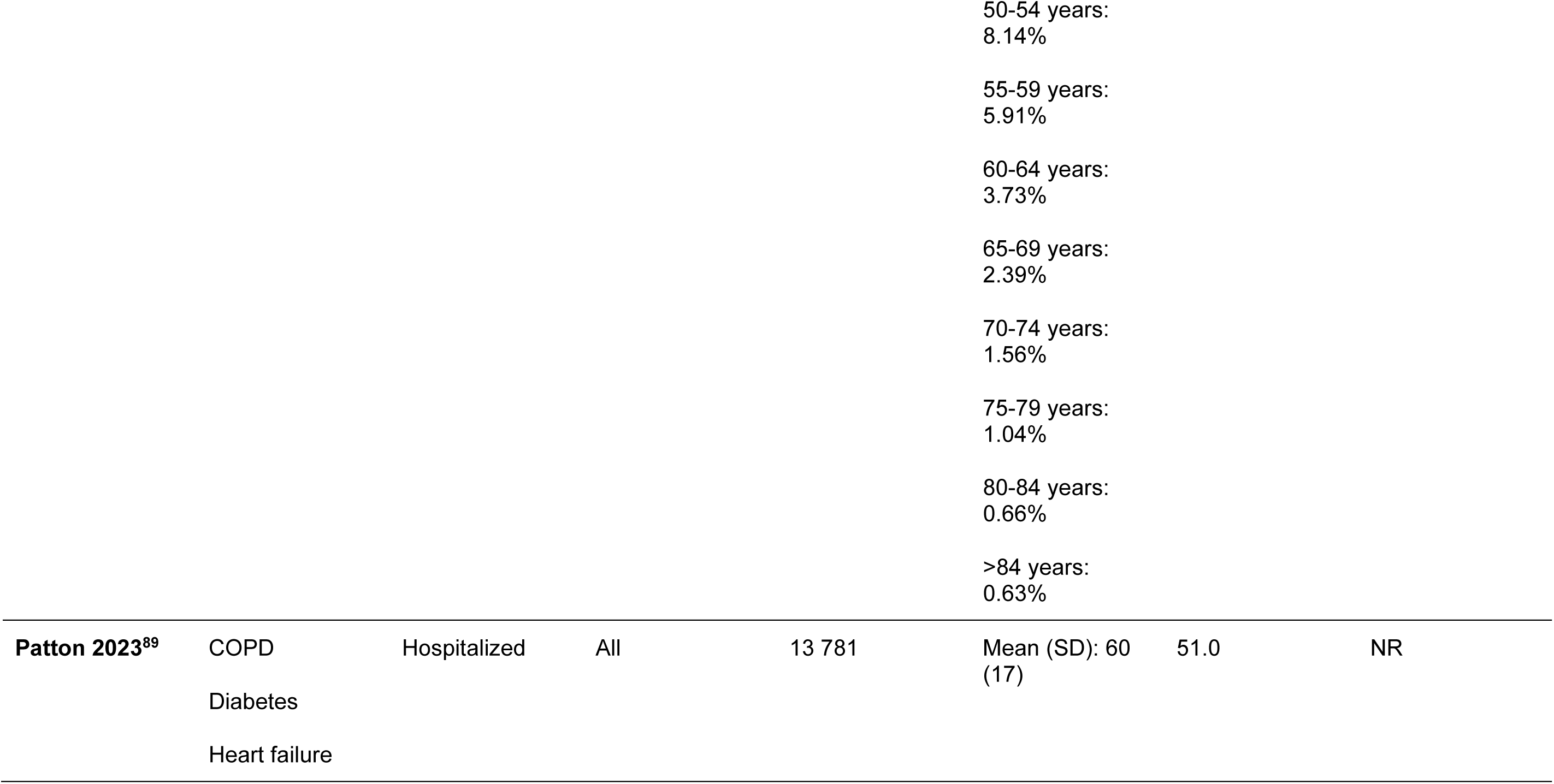
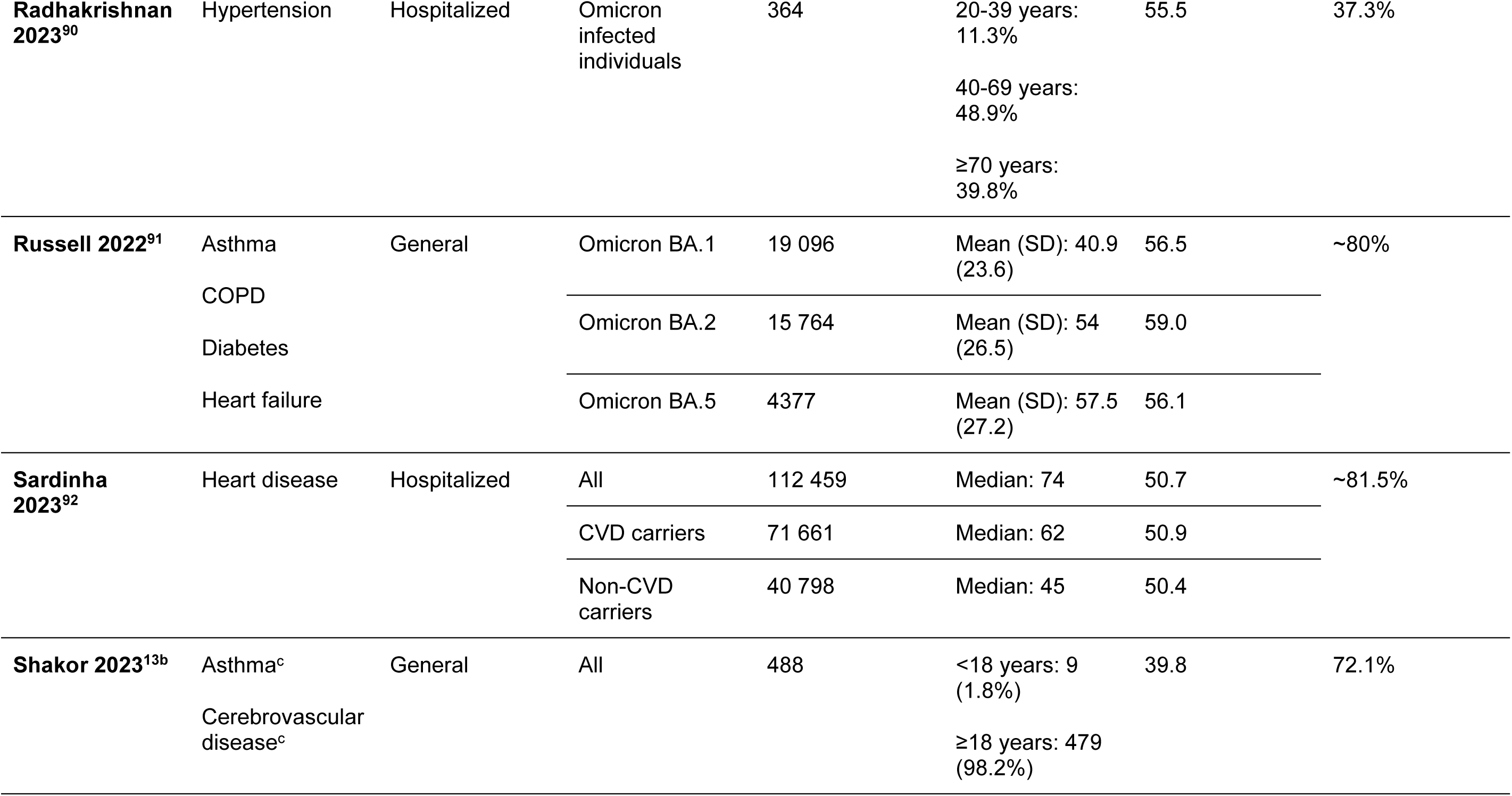
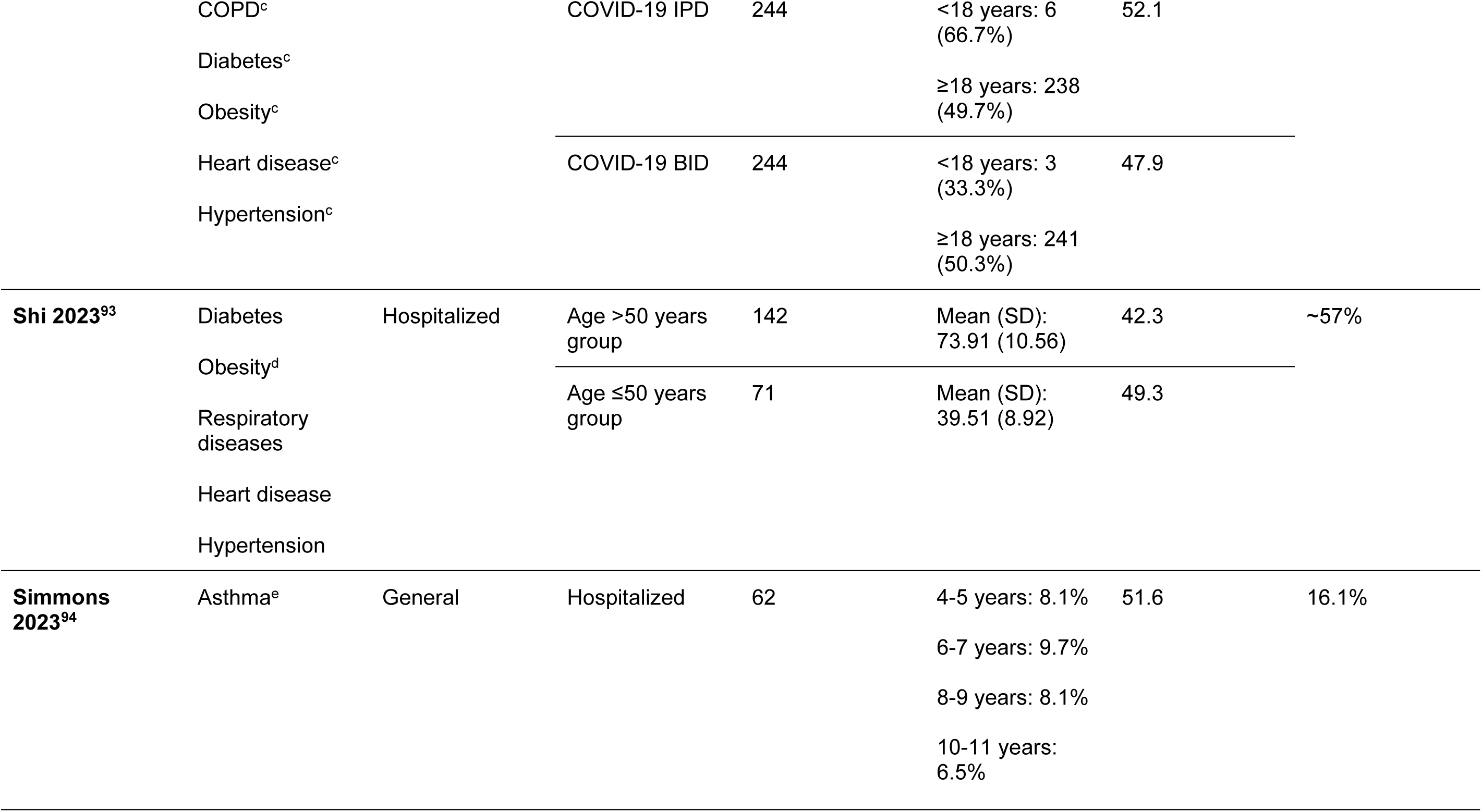
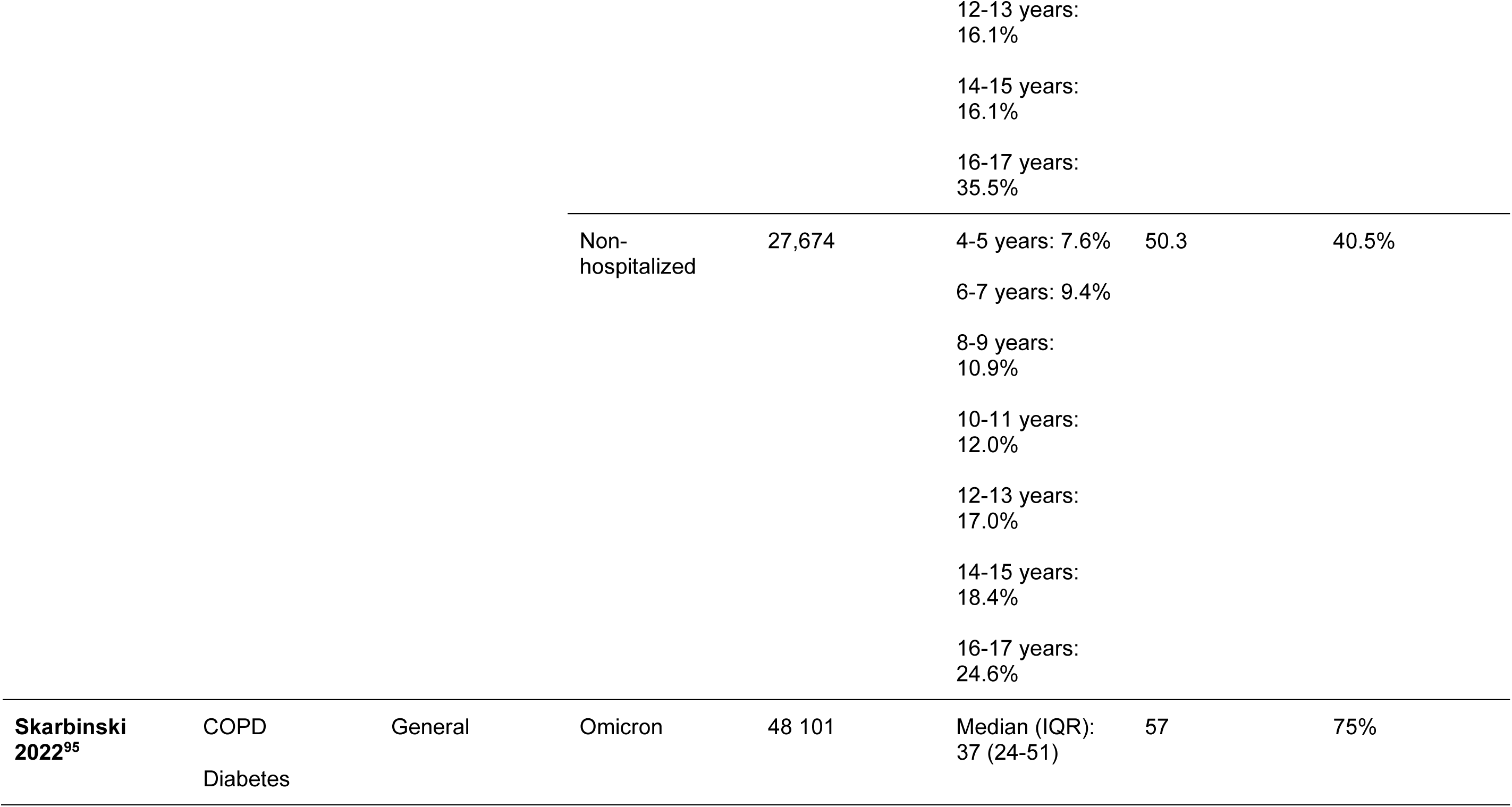
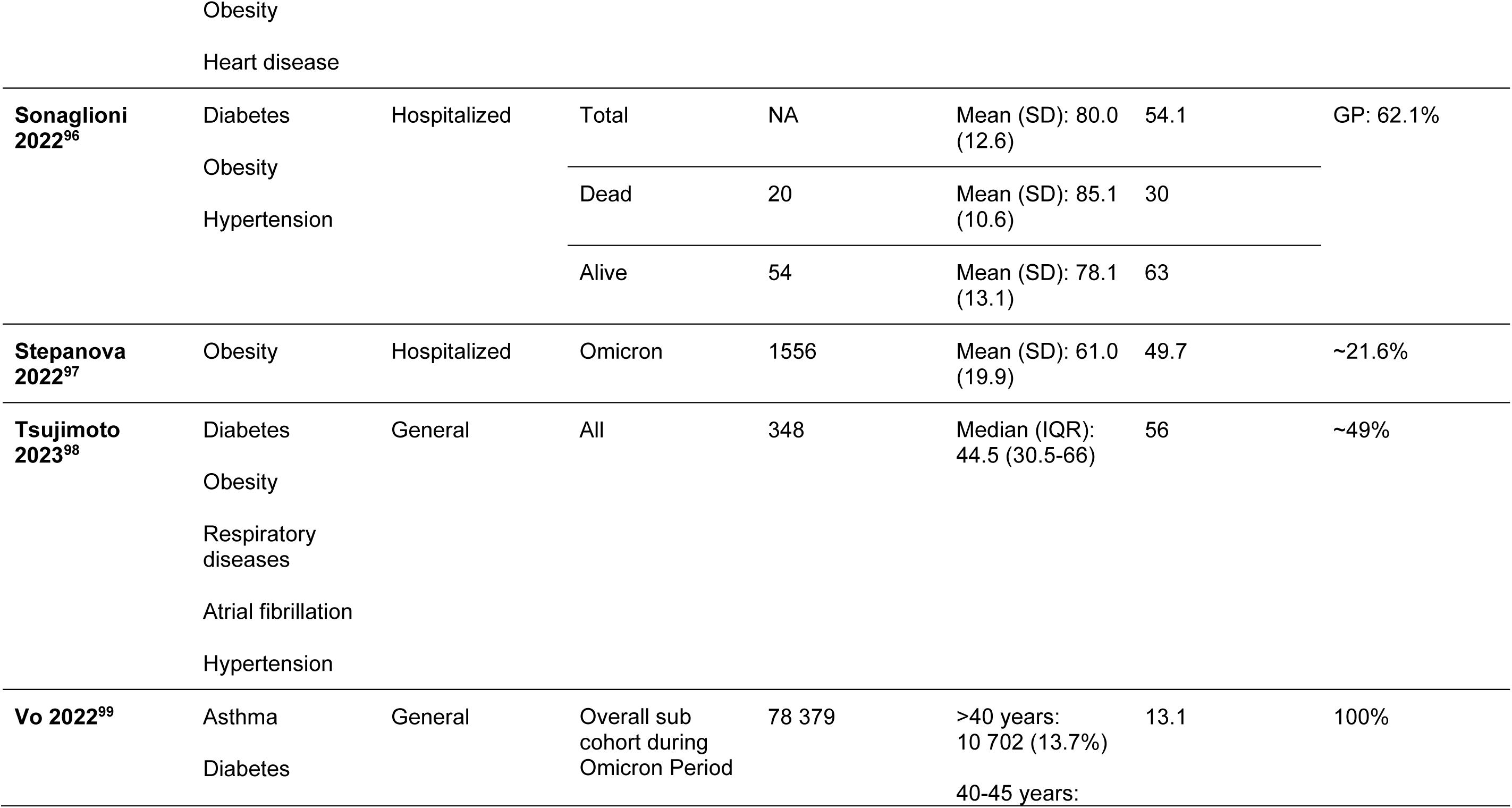
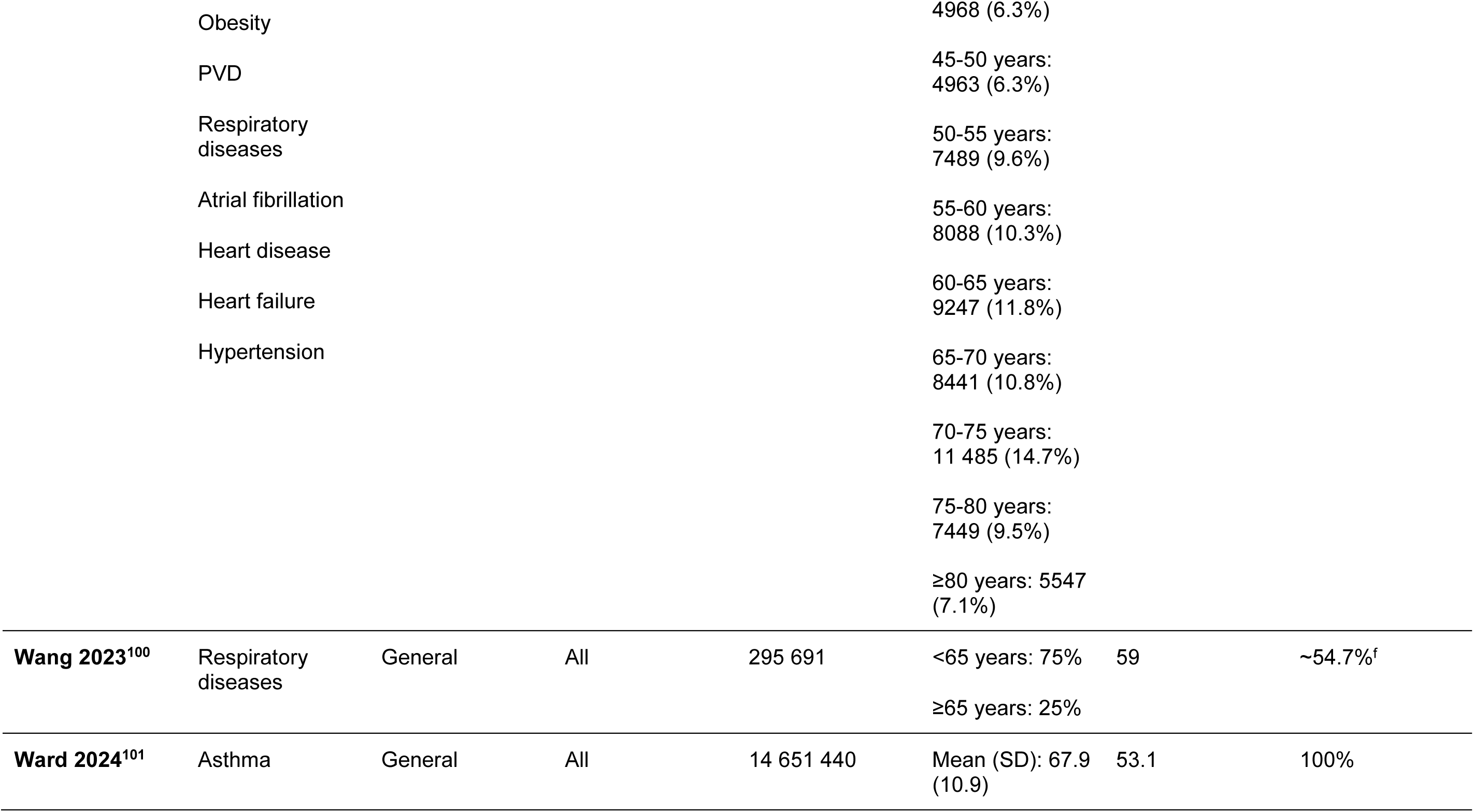
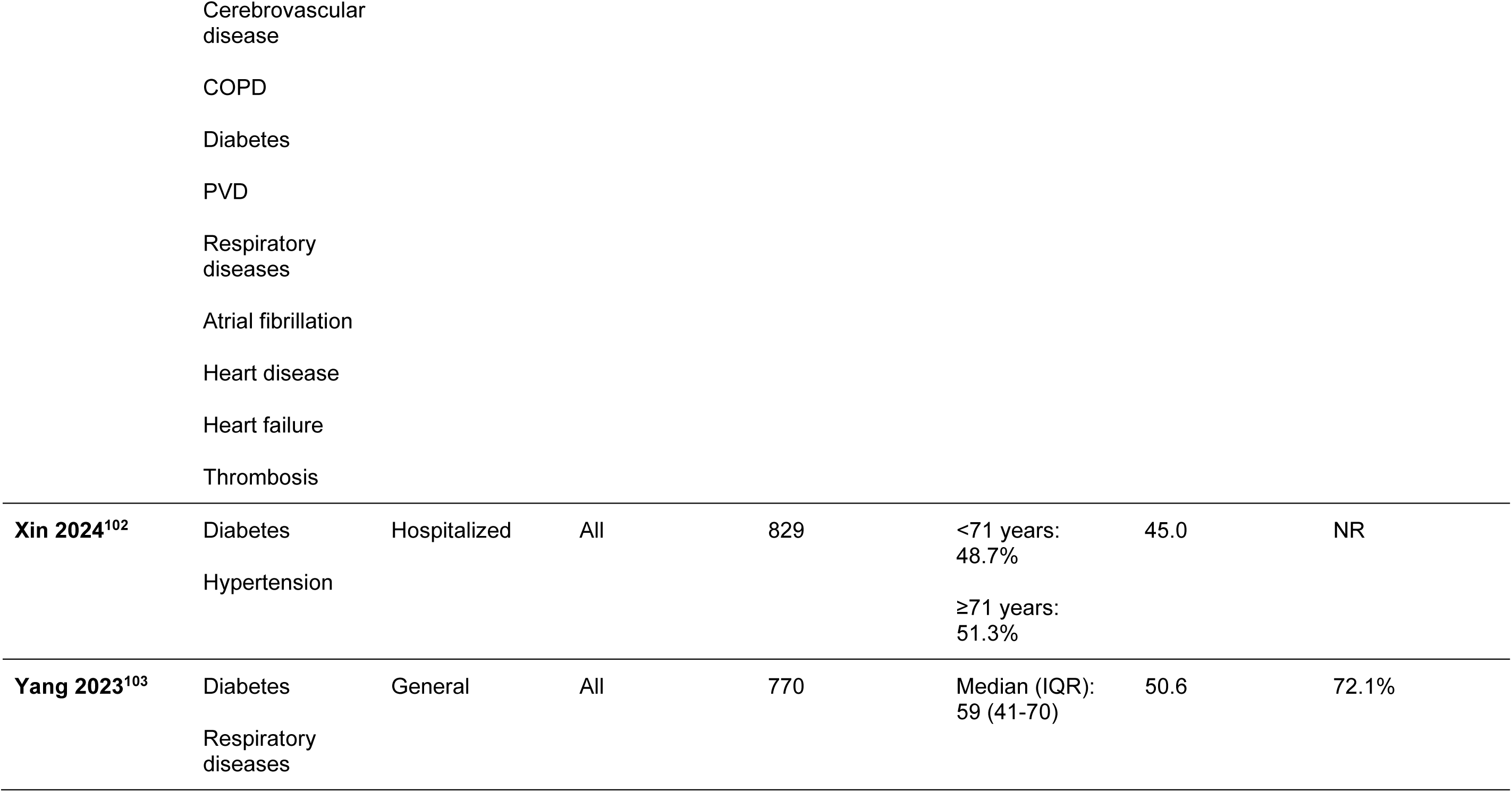
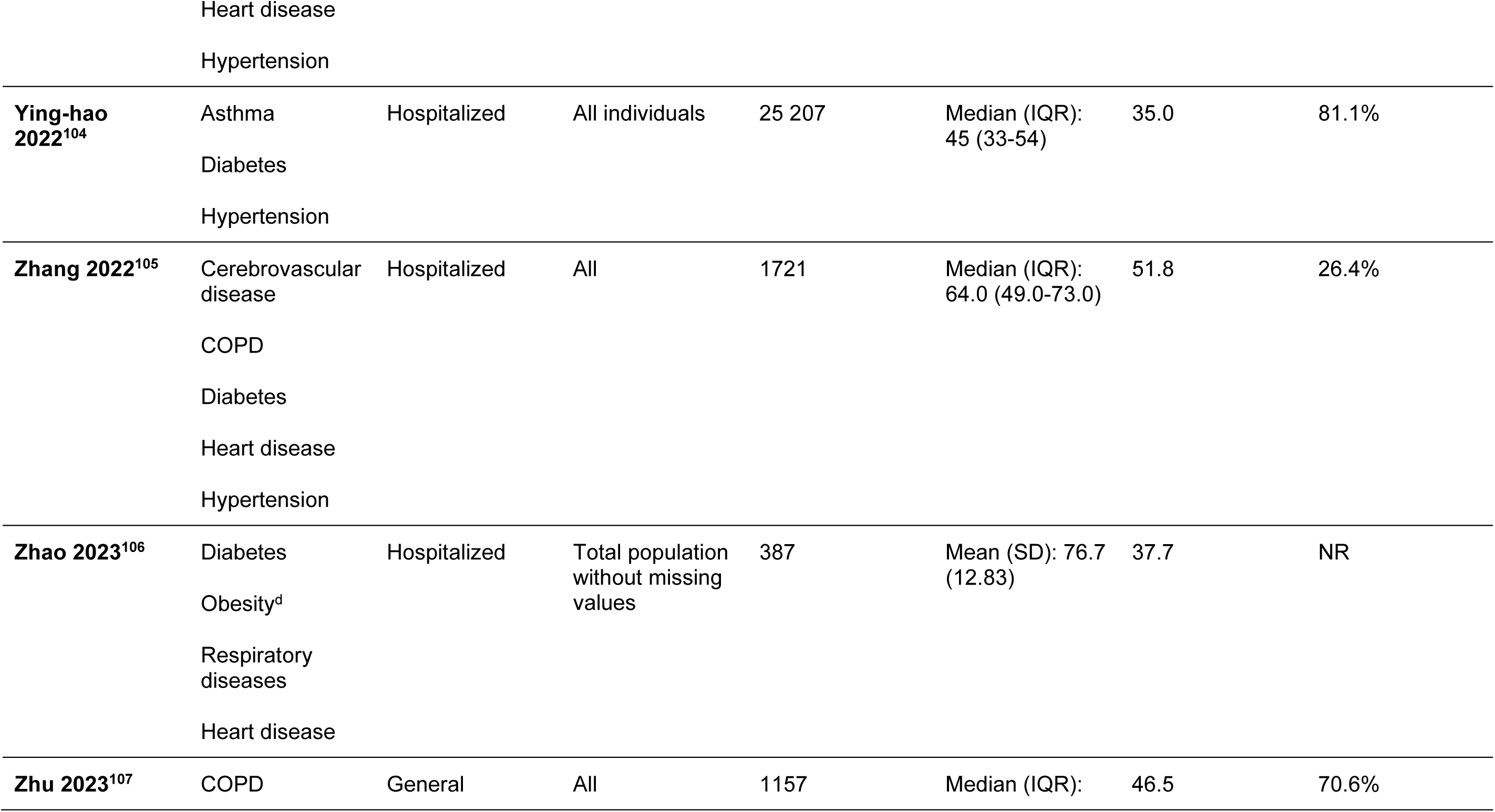
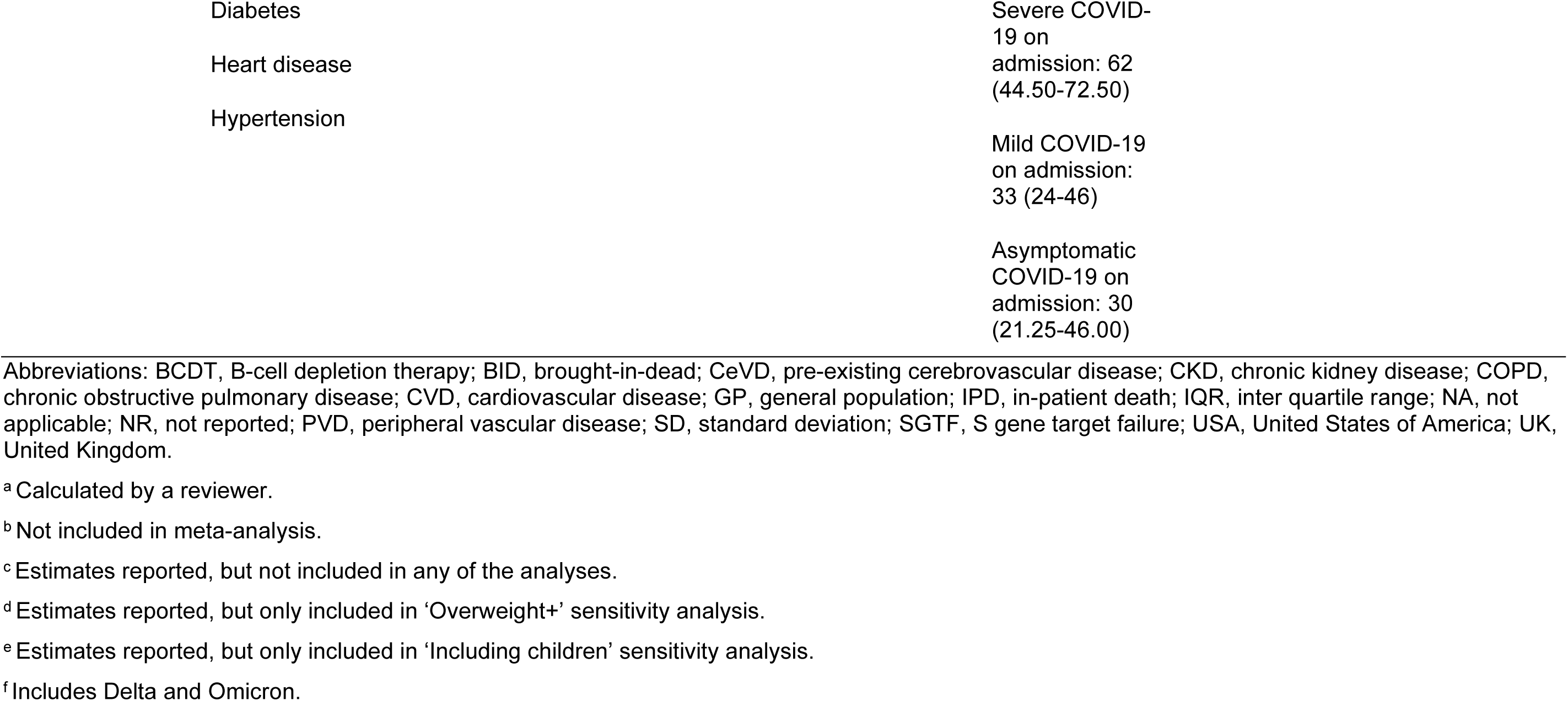
Patient characteristics of the Included Studies.

**Table 3.**
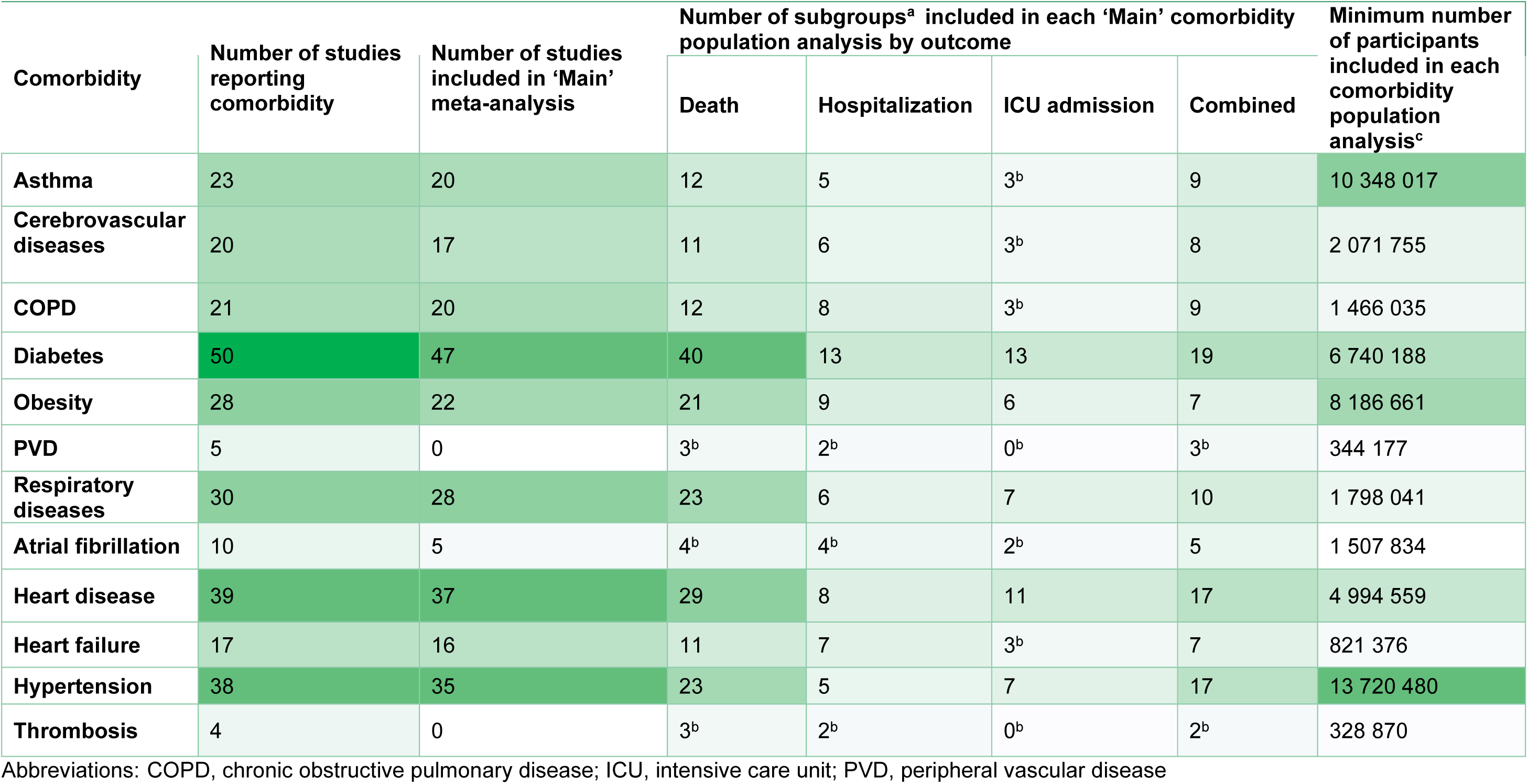
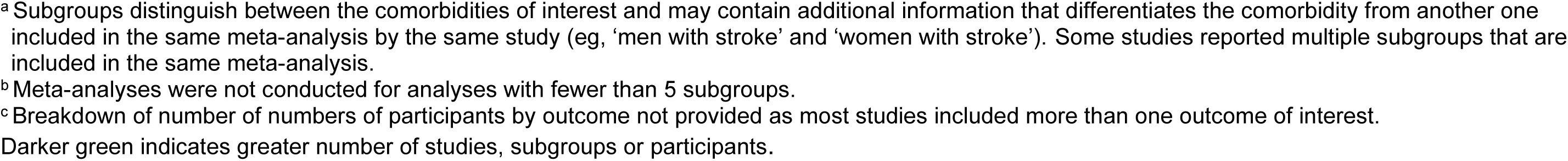
Number of Studies, Subgroups and Minimum Number of Participants Included in Each Comorbidity Population Analysis.

| Comorbidity | Number of studies reporting comorbidity | Number of studies included in 'Main' meta-analysis | Number of subgroups<sup>a</sup> included in each 'Main' comorbidity population analysis by outcome |  |  |  | Minimum number of participants included in each comorbidity population analysis<sup>c</sup> |
| --- | --- | --- | --- | --- | --- | --- | --- |
|  |  |  | Death | Hospitalization | ICU admission | Combined |  |
| **Asthma** | 23 | 20 | 12 | 5 | 3<sup>b</sup> | 9 | 10 348 017 |
| **Cerebrovascular diseases** | 20 | 17 | 11 | 6 | 3<sup>b</sup> | 8 | 2 071 755 |
| **COPD** | 21 | 20 | 12 | 8 | 3<sup>b</sup> | 9 | 1 466 035 |
| **Diabetes** | 50 | 47 | 40 | 13 | 13 | 19 | 6 740 188 |
| **Obesity** | 28 | 22 | 21 | 9 | 6 | 7 | 8 186 661 |
| **PVD** | 5 | 0 | 3<sup>b</sup> | 2<sup>b</sup> | 0<sup>b</sup> | 3<sup>b</sup> | 344 177 |
| **Respiratory diseases** | 30 | 28 | 23 | 6 | 7 | 10 | 1 798 041 |
| **Atrial fibrillation** | 10 | 5 | 4<sup>b</sup> | 4<sup>b</sup> | 2<sup>b</sup> | 5 | 1 507 834 |
| **Heart disease** | 39 | 37 | 29 | 8 | 11 | 17 | 4 994 559 |
| **Heart failure** | 17 | 16 | 11 | 7 | 3<sup>b</sup> | 7 | 821 376 |
| **Hypertension** | 38 | 35 | 23 | 5 | 7 | 17 | 13 720 480 |
| **Thrombosis** | 4 | 0 | 3<sup>b</sup> | 2<sup>b</sup> | 0<sup>b</sup> | 2<sup>b</sup> | 328 870 |

Abbreviations: COPD, chronic obstructive pulmonary disease; ICU, intensive care unit; PVD, peripheral vascular disease
<sup>a</sup> Subgroups distinguish between the comorbidities of interest and may contain additional information that differentiates the comorbidity from another one included in the same meta-analysis by the same study (eg, 'men with stroke' and 'women with stroke'). Some studies reported multiple subgroups that are included in the same meta-analysis.
<sup>b</sup> Meta-analyses were not conducted for analyses with fewer than 5 subgroups.
<sup>c</sup> Breakdown of number of numbers of participants by outcome not provided as most studies included more than one outcome of interest.
Darker green indicates greater number of studies, subgroups or participants.

### Risk of Bias Assessment

Risk of bias (**eTable 3**) was assessed in all included studies (n = 72); 56 and 14 were found to be low and medium risk, respectively. Two studies^12,13^ were found to be high risk and were excluded from the analysis. Two additional studies were excluded from the analysis for not reporting confidence intervals (n = 1)^14^ and for conflicting interpretation of results within the study (n = 1).^15^

### Meta-Analysis

The risk ratios for death, hospitalization, ICU admission, and combined outcomes among individuals with each comorbidity compared with individuals without comorbidities are shown in **Figure 2**.

**Figure 2.**
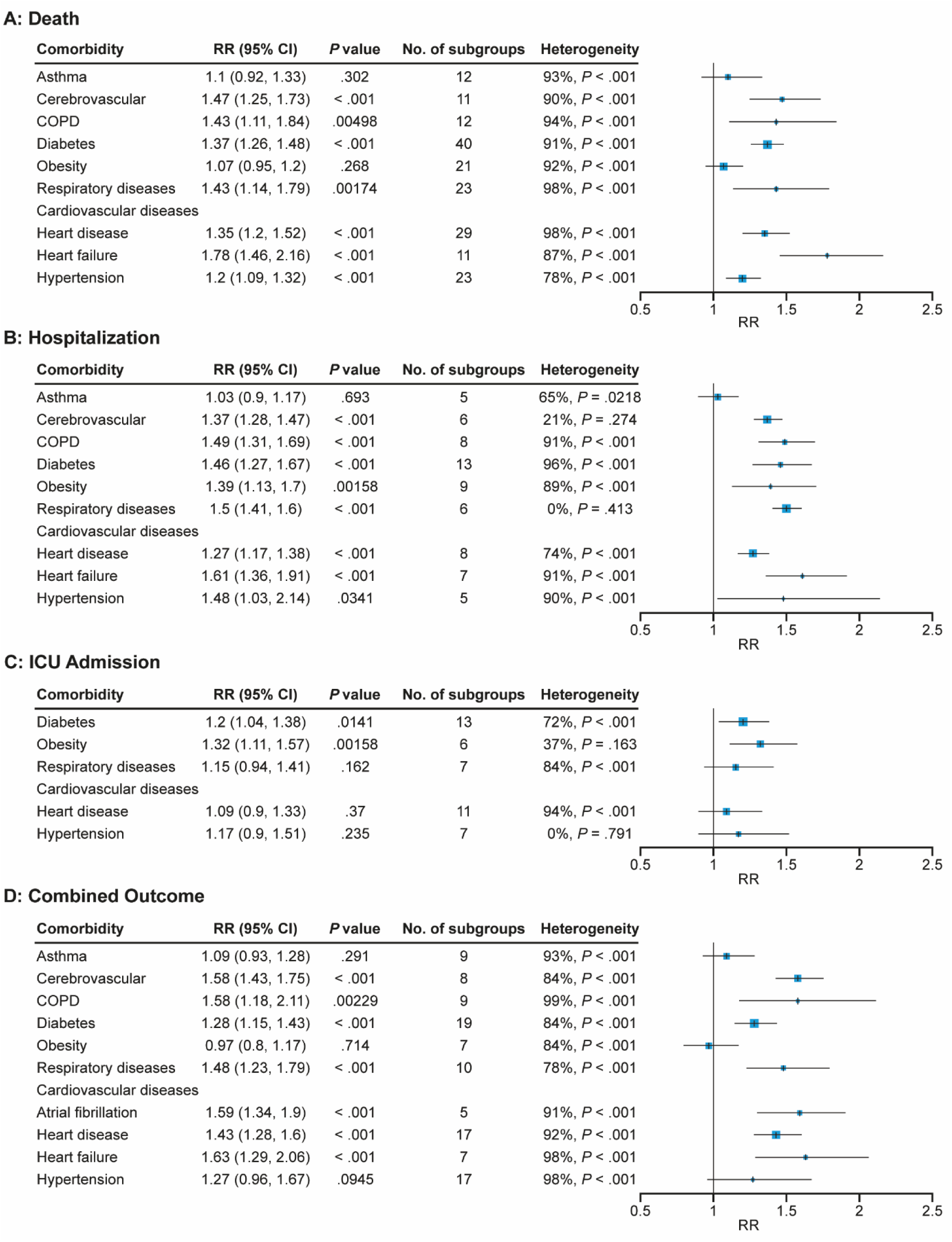
Forest Plots of the Risk of A) Death B) Hospitalization C) ICU Admission, and D) Combined Outcomes by Comorbid Population. Abbreviations: CI, confidence interval; COPD, chronic obstructive pulmonary disease; ICU, intensive care unit; RR, risk ratio.

Due to insufficient numbers of studies, meta-analysis was not conducted for the following outcomes and comorbid conditions: ICU admission for asthma, cerebrovascular diseases, COPD, and heart failure; all outcomes except the combined outcome for atrial fibrillation; and all outcomes for PVD and thrombosis (**Table 3**). These outcomes and comorbidities are excluded from the following meta-analysis but synthesized narratively in the supplement (**eResults**).

#### Risk of Death

Individuals with any of the comorbidities meta-analyzed, except asthma and obesity, had a significantly increased risk of death compared with individuals without the respective comorbidity (**Figure 2A**).

Statistical heterogeneity was considerable in all analyses except among the comorbid populations with heart failure and hypertension, which had substantial heterogeneity (**Figure 2A**). There was no statistical evidence of publication bias for any of the applicable comorbid populations analyzed, except diabetes (**eTable 4**).

All sensitivity analyses results had the same direction of effect and degree of significance as the main analysis for the risk of death outcome (**eTable 4**), indicating robustness of the main analysis results.

Subgroup analyses showed the direction of effect, versus the main analysis, changed for individuals with asthma in the ‘Hospitalized’ and ‘>50 years’ subgroups (ie. these groups with asthma had significantly lower risk of death versus those without asthma) (**eTable 5**).

#### Risk of Hospitalization

For all meta-analyzed comorbid conditions, except asthma, individuals with the comorbidity had a significantly increased risk of hospitalization in comparison with individuals without the comorbidity (**Figure 2B**). Statistical heterogeneity was substantial or considerable for most comorbidities (7/9 applicable comorbid populations; **Figure 2B**). Publication bias was not assessed in any of the analyses due to the small number of studies, except in the comorbid group with diabetes, where no bias was found (**eTable 6**).

All sensitivity analyses results had the same direction of effect, and nearly all had the same degree of significance, as the main analyses for the risk of hospitalization outcome (**eTable 6**), indicating robustness of the main analysis results.

In all subgroup analyses, the direction of effect was the same compared with the main analysis, except individuals with asthma had (non-significantly) lower risk of hospitalization. In nearly all analyses the significance level remained unchanged (**eTable 7**).

#### Risk of ICU Admission

While all 5 meta-analyzed comorbidities had increased risk of ICU admission compared with individuals without the comorbidity, this risk was significant in diabetes and obesity only (**Figure 2C**).

Statistical heterogeneity ranged from not important to considerable across the analyses. Publication bias was not assessed in any of the analyses due to the small number of studies, except in the groups with diabetes and heart disease, in which no bias was found (**eTable 8**).

Four sensitivity analyses resulted in loss or gain of significance compared with the main analysis for risk of admission to ICU, although the direction of effect (ie, the higher risk of ICU admission) remained the same (**eTable 8**).

In the ‘COVID-19 related only’ subgroup, the increased risk of ICU admission for comorbid populations with respiratory diseases and heart disease was significant (**eTable 9**), while this relationship was non-significant in the main analysis.

#### Risk of the Combined Outcome (Death, Hospitalization or ICU Admission)

For all comorbid conditions, individuals with the comorbidity had significantly increased risk of the combined outcome in comparison with individuals without the comorbidity, except in individuals with asthma and hypertension, where the increased risk was non-significant, and in individuals with obesity, who had lower (but non-significant) risk of the combined outcome in comparison with individuals without obesity (**Figure 2D**). For details of sensitivity and subgroup analyses for the combined outcome see **eResults** and **eTables 10** and **11**.

Individual meta-analyses for each comorbidity, stratified by outcome, can be found in the supplement (**eResults**).

## Discussion

This SLR and meta-analysis examined the impact of SARS-CoV-2 on four outcomes in populations with comorbidities versus those without during the Omicron era. For those with cerebrovascular disease, COPD, diabetes, respiratory diseases (excluding COPD and asthma), heart disease, and heart failure, the pooled relative risks (RR) of death, hospitalization, and a combined outcome were significantly higher, ranging from 1.27 (95% CI, 1.17-1.38) to 1.78 (95% CI, 1.46-2.16). Individuals with hypertension had a significantly increased risk of death and hospitalization, with pooled RRs of 1.20 (95% CI, 1.09-1.32) and 1.48 (95% CI, 1.03-2.14), respectively. Those with obesity and atrial fibrillation demonstrated a significantly increased risk of hospitalization and combined outcomes, respectively, with pooled RRs of 1.39 (95% CI, 1.13-1.70) and 1.59 (95% CI, 1.34-1.90). Additionally, individuals with diabetes and obesity had a significantly increased risk of ICU admission, with pooled RRs of 1.20 (95% CI, 1.04-1.38) and 1.32 (95% CI, 1.11-1.57), respectively.

Of the meta-analyzed comorbid populations, individuals with heart failure had the greatest increase in pooled RRs of death at 1.78 (95% CI, 1.46-2.16), hospitalization at 1.61 (95% CI, 1.36-1.91), and the combined outcome at 1.63 (95% CI, 1.29-2.06). Individuals with asthma had a non-significant increase in risk of any severe outcome, but hospitalized individuals with asthma had a significantly lower pooled RR of death at 0.84 (95% CI, 0.75-0.95) compared with hospitalized individuals without asthma.

### Cardiovascular and Cerebrovascular Disease

Individuals with cerebrovascular disease, heart disease, and heart failure had significantly higher risk of all severe outcomes from COVID-19, except ICU admission. This is consistent with several studies conducted prior to and during the Omicron era which reported longer time to recovery^16^ and greater risk of severe outcomes from COVID-19 in patients with cerebrovascular^17–20^ and cardiovascular diseases.^18–21^ In the current study, individuals with hypertension had significantly higher risk of death and hospitalization, but not ICU admission or combined outcomes, consistent with a pre-Omicron meta-analysis,^22^ although this previous study also reported significantly higher risk of ICU admission. Individuals with atrial fibrillation had significantly higher risk of the combined outcome.

The underlying mechanisms for these outcomes are not well understood, but several have been proposed. For cerebrovascular disease, proinflammatory and hypercoagulable states associated with COVID-19 have been suggested.^23,24^ For heart disease, possible mechanisms include COVID-19–associated hypercoagulation, direct viral damage to cardiomyocytes, or pneumonia-induced gas exchange obstruction leading to cardiomyocyte injury and apoptosis.^2,21^ In heart failure and hypertension, viral uptake through increased angiotensin-converting enzyme 2 (ACE2) receptors may increase the risk of severe outcomes.^22,25^, Direct cardiac injury, increased coagulation abnormalities, thrombotic events, and stress cardiomyopathy increase the risk of severe outcomes in individuals with heart failure or atrial fibrillation.^25,26^

### Respiratory Diseases

Individuals with COPD and other respiratory diseases (including acute respiratory distress syndrome, interstitial lung disease, and chronic lung diseases excluding asthma), had significantly higher risks of death, hospitalization and the combined outcome, consistent with a pre-Omicron meta-analysis on COPD.^27^

Micro-thrombosis, secondary bacterial infection and the effects of intrapulmonary shunting, have been proposed as potential mechanisms increasing the risk of severe outcomes from COVID-19 in COPD.^28^ For all chronic respiratory diseases, reduced lung function at baseline may decrease tolerance to further lung injury from SARS-CoV-2 infection, resulting in poorer outcomes.^29^ Additionally, individuals with chronic lung disease may experience impaired responses to vaccines, which may lead to increased risk of acquiring SARS-Cov-2 and developing severe complications.^30^

The lack of association between asthma and severe outcomes from COVID-19 may be attributed to down-regulated ACE2 receptors in those with T2-high asthma, reducing susceptibility to SARS-CoV-2 infection.^31^ A previous meta-analysis found a similar protective effect of asthma; it is possible that controlling asthma symptoms by routine asthma treatment reduces worsening of symptoms and risk of hospitalization.^32^

### Diabetes

Risk of severe outcomes from COVID-19 was significantly increased in individuals with diabetes, consistent with pre-Omicron studies that reported increased risk of mortality.^19,33^ Cytokine storm, pulmonary and endothelial dysfunction, and hypercoagulation have been attributed as potential mechanisms increasing the risk of severe outcomes.^34^

### Obesity

Risk of ICU admission and hospitalization were significantly increased in individuals with obesity, but there was no association with death and the combined outcome. A pre-Omicron SLR and meta-analysis found a similar pattern, with increased risk of COVID-19–related hospitalizations, but also death, unlike the current study.^35^ The increased risk of severe disease may be explained by a hyperinflammatory response to SARS-CoV-2 infection linked to high blood levels of saturated fatty acids in patients with obesity,^36^ as well as predisposition to immunopathological exaggeration of immuno-metabolic disorders.^37^ A possible explanation for the protective effect from death observed in our study is that where ICU occupancy was reaching capacity within a hospital trust, individuals with obesity were explicitly prioritized in ICU admission decisions. This prioritization could be due to the difficulty in maintaining adequate oxygen levels without mechanical ventilation on a normal hospital ward.^38^ Consequently, they may have received higher standard of care.^38^

### Quality of Evidence

To our knowledge, this is the first comprehensive SLR to assess severe outcomes from COVID-19 in people with comorbidities during the Omicron era. Most of the results included in the meta-analyses were adjusted for age and comorbidities, and main analyses were based on these most adjusted estimates, increasing confidence in the findings. In addition, several sensitivity and subgroup analyses were performed, and most sensitivity analyses showed no change in significance from the main analyses, indicating that the pooled estimates are robust.

This study had several limitations. First, there was high clinical and statistical heterogeneity between the studies, due, in part, to variation in interpretation of outcome definitions. This was especially pronounced for the combined outcome as multiple different outcomes were included in its definition. In addition, different adjustments were used between studies: while most studies were adjusted for demographic characteristics, there were variations in adjustments of vaccination rate and comorbidities, reducing comparability of the studies.

Second, while studies included in this review cover wide geographical areas, inter-country differences, such as country-level income, are important predictors of COVID-19 control measures, which in turn affect COVID-19 rates and severity.^39,40^ However, analysis by geographical region was not performed due to the high between-study heterogeneity and small numbers of studies by region. Finally, due to lack of data, high between-study heterogeneity, and overlap in Omicron subvariant time periods, no sub-analyses were performed by time period, subvariant, vaccine type, or vaccination status. These can be important effect modifiers and should be investigated in future research.

#### Implications for Practice and Further Research

This study synthesizes data to enhance healthcare providers’ understanding of COVID-19 disease burden in populations with comorbidities during the Omicron era and ensure targeted measures are implemented.

Vaccination is the most effective tool to reduce the risk and prevalence of COVID-19 disease, COVID-19-related hospitalization, ICU admission and death.^4,41^ Targeting vaccination and other public health measures for those patients at greatest risk due to comorbidity may help prevent or mitigate severe outcomes from COVID-19.

Due to lack of data for several comorbidities in the Omicron era, further high-quality prospective studies are needed to investigate severe outcomes from COVID-19 in the following populations: type 1 and type 2 diabetes, PVD, hypertension, and thrombosis. It would be valuable to investigate the effects of multi-morbidity and the severity of comorbidities on patient outcomes, as well as the reciprocal effect of COVID-19 on the underlying comorbidities.

Finally, further investigations into the impact of age, evolving variants, vaccination status and geographical region on comorbid individuals with COVID-19 in the Omicron era, are likely to provide more insight into disease epidemiology.

## Conclusions

This SLR and meta-analysis on the burden of COVID-19 in individuals with comorbidities during the Omicron era demonstrated that those with cerebrovascular disease, COPD, diabetes, respiratory diseases, heart disease, heart failure, and hypertension are at increased risk of death and hospitalization, while individuals with diabetes and obesity are at increased risk of ICU admission, in comparison with those without the respective comorbidity. Of the meta-analyzed comorbid conditions, heart failure was associated with the greatest increase in risk of severe outcomes from COVID-19. Clinicians and policy makers should consider targeting public health measures, such as seasonal vaccination, toward these most at-risk groups to protect vulnerable populations with comorbidities who are at higher risk for developing severe outcomes following COVID-19 disease.

## Data Availability

The datasets generated during and/or analyzed during the current study are available from the corresponding author on reasonable request.

## Acknowledgements

The authors would like to thank the individuals, their families and all investigators involved in this study.

Guidance through the review process as well as contributions to systematic review processes, such as screening, risk of bias assessment, and data extraction, were provided by Nick Pooley, PhD (Maverex Ltd), Masoumeh Kisomi, PhD (Maverex Ltd), and Megha Garg, MD (Maverex Ltd). Statistical support, including the design and running of the meta-analyses, was provided by Medha Shrivastava, MSc (Maverex Ltd). Kate Misso, MSc/MCLIP (Maverex Ltd), designed and performed the electronic searches in this systematic review.

Medical writing support, including assisting authors with development of the outline and initial draft, incorporation of comments, figure preparation, referencing, and data checking was provided by Rachel O’Meara, PhD, and editorial support, including formatting, proofreading, and submission was provided by Michelle Seddon, Dip Psychol, all of Paragon (a division of Prime, Knutsford, UK). The study was supported by BioNTech SE, Mainz, Germany, according to Good Publication Practice guidelines (<u>Link</u>). The sponsor was involved in the study design, analysis and interpretation of data in the manuscript as well as data checking of information provided in the manuscript. However, ultimate responsibility for opinions, conclusions, and data interpretation lies with the authors.

## Author contributions

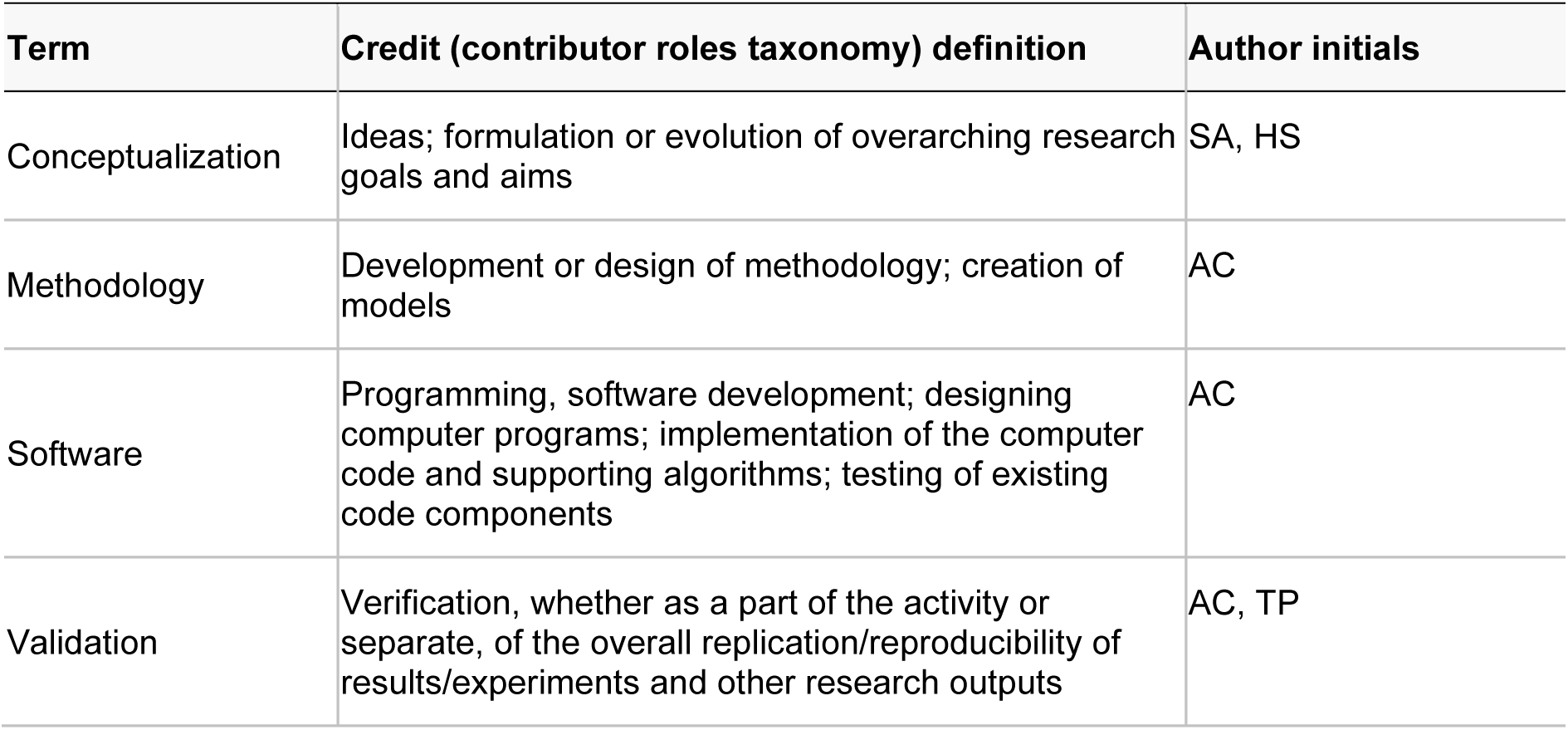
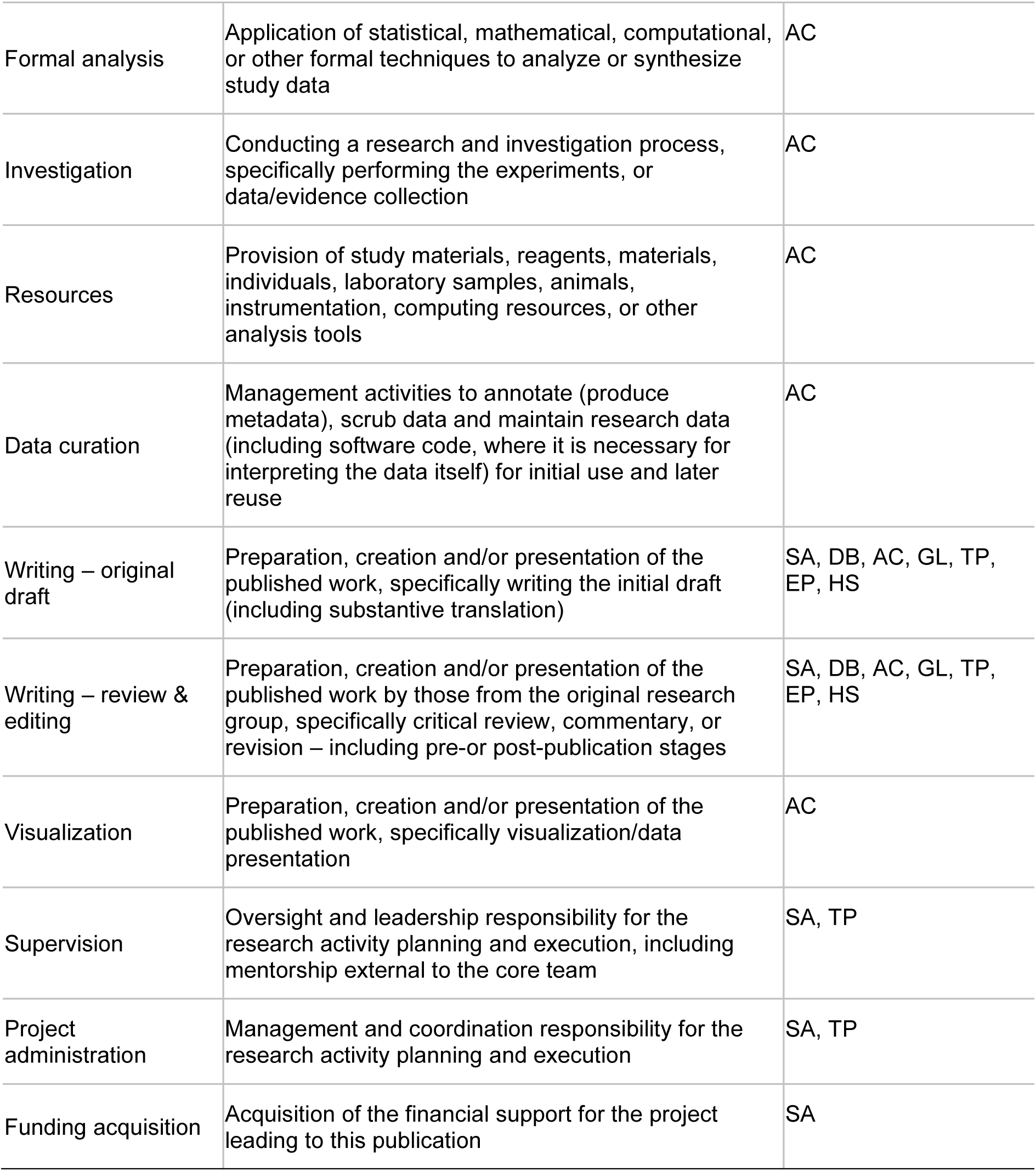

## References

1. Nab L, Parker EPK, Andrews CD, et al. Changes in COVID-19-related mortality across key demographic and clinical subgroups in England from 2020 to 2022: a retrospective cohort study using the OpenSAFELY platform. Lancet Public Health. 2023;8(5):e364–e377.

2. Harrison SL, Buckley BJR, Rivera-Caravaca JM, Zhang J, Lip GYH. Cardiovascular risk factors, cardiovascular disease, and COVID-19: an umbrella review of systematic reviews. Eur Heart J Qual Care Clin Outcomes. 2021;7(4):330–339.

3. Chatterjee S, Nalla LV, Sharma M, et al. Association of COVID-19 with comorbidities: an update. ACS Pharmacol Transl Sci. 2023;6(3):334–354.

4. Ellis RJ, Moffatt CR, Aaron LT, et al. Factors associated with hospitalisations and deaths of residential aged care residents with COVID-19 during the Omicron (BA.1) wave in Queensland. Med J Aust. 2023;218(4):174–179.

5. Veneti L, Boas H, Brathen Kristoffersen A, et al. Reduced risk of hospitalisation among reported COVID-19 cases infected with the SARS-CoV-2 Omicron BA.1 variant compared with the Delta variant, Norway, December 2021 to January 2022. Euro Surveill. 2022;27(4):pii=2200077.

6. Maslo C, Friedland R, Toubkin M, et al. Characteristics and outcomes of hospitalized patients in South Africa during the COVID-19 Omicron wave compared with previous waves. JAMA. 2022;327(6):583–584.

7. Sheikh A, Kerr S, Woolhouse M, et al. Severity of omicron variant of concern and effectiveness of vaccine boosters against symptomatic disease in Scotland (EAVE II): a national cohort study with nested test-negative design. Lancet Infect Dis. 2022:959–966.

8. World Health Organization. Classification of Omicron (B.1.1.529): SARS-CoV-2 variant of concern. Updated November 26, 2021.https://www.who.int/news/item/26-11-2021-classification-of-omicron-(b.1.1.529)-sars-cov-2-variant-of-concern. Accessed July 3, 2024.

9. Jose MM, Feng J, Nguyen HT, et al. The influence of comorbidities, general health status, and self-care self-efficacy on COVID-19 symptoms during the omicron wave. Cureus. 2023;15(11):e49176.

10. Ouzzani M, Hammady H, Fedorowicz Z, Elmagarmid A. Rayyan-a web and mobile app for systematic reviews. Syst Rev. 2016;5(1):210.

11. Viechtbauer W. Conducting meta-analyses in R with the metafor package. J Stat Softw. 2010;36(3):1–48.

12. Geng Y, Nie Q, Liu F, et al. Understanding clinical characteristics influencing adverse outcomes of Omicron infection: a retrospective study with propensity score matching from a Fangcang hospital. Front Cell Infect Microbiol. 2023;13:1115089.

13. Shakor ASaA, Samsudin EZ, Chen XW, Ghazali MH. Factors associated with COVID-19 brought-in deaths: a data-linkage comparative cross-sectional study. J Infect Public Health. 2023;16(12):2068–2078.

14. Morris CP, Eldesouki RE, Sachithanandham J, et al. Omicron subvariants: clinical, laboratory, and cell culture characterization. Clin Infect Dis. 2023;76(7):1276–1284.

15. Bao S, Lu G, Kang Y, et al. A diagnostic model for serious COVID-19 infection among older adults in Shanghai during the Omicron wave. Front Med (Lausanne). 2022;9:1018516.

16. Oelsner EC, Sun Y, Balte PP, et al. Epidemiologic Features of Recovery From SARS-CoV-2 Infection. JAMA Network Open. 2024;7(6):e2417440–e2417440.

17. Ferrone SR, Sanmartin MX, Ohara J, et al. Acute ischemic stroke outcomes in patients with COVID-19: a systematic review and meta-analysis. J Neurointerv Surg. 2024;16(4):333–341.

18. Chen R, Liang W, Jiang M, et al. Risk Factors of Fatal Outcome in Hospitalized Subjects With Coronavirus Disease 2019 From a Nationwide Analysis in China. Chest. 2020;158(1):97–105.

19. Williamson EJ, Walker AJ, Bhaskaran K, et al. Factors associated with COVID-19-related death using OpenSAFELY. Nature. 2020;584(7821):430–436.

20. Wang L, He W, Yu X, et al. Coronavirus disease 2019 in elderly patients: Characteristics and prognostic factors based on 4-week follow-up. J Infect. 2020;80(6):639–645.

21. Mishra P, Parveen R, Bajpai R, Samim M, Agarwal NB. Impact of cardiovascular diseases on severity of COVID-19 patients: a systematic review. Ann Acad Med Singap. 2021;50(1):52–60.

22. Pranata R, Lim MA, Huang I, Raharjo SB, Lukito AA. Hypertension is associated with increased mortality and severity of disease in COVID-19 pneumonia: a systematic review, meta-analysis and meta-regression. J Renin Angiotensin Aldosterone Syst. 2020;21(2):1470320320926899.

23. Katz JM, Libman RB, Wang JJ, et al. Cerebrovascular complications of COVID-19. Stroke. 2020;51(9):e227–e231.

24. Ntaios G, Michel P, Georgiopoulos G, et al. Characteristics and outcomes in patients with COVID-19 and acute ischemic stroke: the global COVID-19 stroke registry. Stroke. 2020;51(9):e254–e258.

25. Bader F, Manla Y, Atallah B, Starling RC. Heart failure and COVID-19. Heart Fail Rev. 2021;26(1):1–10.

26. Li Z, Shao W, Zhang J, et al. Prevalence of atrial fibrillation and associated mortality among hospitalized patients with COVID-19: a systematic review and meta-analysis. Front Cardiovasc Med. 2021;8:720129.

27. Gerayeli FV, Milne S, Cheung C, et al. COPD and the risk of poor outcomes in COVID-19: a systematic review and meta-analysis. EClinicalMedicine. 2021;33:100789.

28. Singh D, Mathioudakis AG, Higham A. Chronic obstructive pulmonary disease and COVID-19: interrelationships. Curr Opin Pulm Med. 2022;28(2):76–83.

29. Russell CD, Lone NI, Baillie JK. Comorbidities, multimorbidity and COVID-19. Nat Med. 2023;29(2):334–343.

30. Liu H, Aviszus K, Zelarney P, et al. Vaccine-elicited B- and T-cell immunity to SARS-CoV-2 is impaired in chronic lung disease patients. ERJ Open Res. 2023;9(5):00400–02023.

31. Sunjaya AP, Allida SM, Di Tanna GL, Jenkins C. Asthma and risk of infection, hospitalization, ICU admission and mortality from COVID-19: systematic review and meta-analysis. J Asthma. 2022;59(5):866–879.

32. Shi L, Han X, Wang Y, Xu J, Yang H. Significant association between asthma and a lower risk of mortality among COVID-19 patients in Spain: A meta-analysis. Qatar Med J. 2024;2024(3):34.

33. Javid FA, Waheed FA, Zainab N, et al. COVID-19 and diabetes in 2020: a systematic review. J Pharm Policy Pract. 2023;16(1):42.

34. Erener S. Diabetes, infection risk and COVID-19. Mol Metab. 2020;39:101044.

35. Sawadogo W, Tsegaye M, Gizaw A, Adera T. Overweight and obesity as risk factors for COVID-19-associated hospitalisations and death: systematic review and meta-analysis. BMJ Nutr Prev Health. 2022;5(1):10–18.

36. Moraes-Vieira P. Immuno-metabolic regulation of macrophages in health and diseases. Presented at: FAPESP Week; June 29, 2024; Shenzhen, China. https://fapesp.br/week/study-reveals-new-factor-associated-with-the-risk-of-severe-covid-19-in-people-with-obesity

37. Khwatenge CN, Pate M, Miller LC, Sang Y. Immunometabolic Dysregulation at the Intersection of Obesity and COVID-19. Review. Frontiers in Immunology. 2021;12

38. Dana R, Bannay A, Bourst P, et al. Obesity and mortality in critically ill COVID-19 patients with respiratory failure. Int J Obes (Lond). 2021;45(9):2028–2037.

39. Durmuş V. Is the country-level income an important factor to consider for COVID-19 control? An analysis of selected 100 countries. Int J Health Gov. 2021;26(2):100–113.

40. Hopman J, Mehtar S. Country level analysis of COVID-19 policies. EClinicalMedicine. 2020;25:100500.

41. Whitaker HJ, Tsang RSM, Byford R, et al. COVID-19 vaccine effectiveness against hospitalisation and death of people in clinical risk groups during the Delta variant period: English primary care network cohort study. Journal of Infection. 2023;87(4):315–327.

42. Agrawal U, Bedston S, McCowan C, et al. Severe COVID-19 outcomes after full vaccination of primary schedule and initial boosters: pooled analysis of national prospective cohort studies of 30 million individuals in England, Northern Ireland, Scotland, and Wales. Lancet. 2022;400(10360):1305–1320.

43. AlBahrani S, AlBarrak A, Al-Musawi T, et al. COVID-19 vaccine had a significant positive impact on patients with SARS-COV-2 during the third (Omicron) wave in Saudi Arabia. J Infect Public Health. 2022;15(11):1169–1174.

44. Arbel R, Peretz A, Sergienko R, et al. Effectiveness of a bivalent mRNA vaccine booster dose to prevent severe COVID-19 outcomes: a retrospective cohort study. Lancet Infect Dis. 2023;23(8):914–921.

45. Arbel R, Sergienko R, Friger M, et al. Effectiveness of a second BNT162b2 booster vaccine against hospitalization and death from COVID-19 in adults aged over 60 years. Nat Med. 2022;28(7):1486–1490.

46. Bedston S, Almaghrabi F, Patterson L, et al. Risk of severe COVID-19 outcomes after autumn 2022 COVID-19 booster vaccinations: a pooled analysis of national prospective cohort studies involving 7.4 million adults in England, Northern Ireland, Scotland and Wales. Lancet Reg Health Eur. 2024;37:100816.

47. Benites-Godínez V, Mendoza-Cano O, Trujillo X, et al. Survival analysis and contributing factors among PCR-confirmed adult inpatients during the endemic phase of COVID-19. Diseases. 2023;11(3):119.

48. Beraud G, Bouetard L, Civljak R, et al. Impact of vaccination on the presence and severity of symptoms in hospitalized patients with an infection of the Omicron variant (B.1.1.529) of the SARS-CoV-2 (subvariant BA.1). Clin Microbiol Infect. 2023;29(5):642–650.

49. Briciu V, Topan A, Calin M, et al. Comparison of COVID-19 severity in vaccinated and unvaccinated patients during the Delta and Omicron wave of the pandemic in a Romanian tertiary infectious diseases hospital. Healthcare. 2023;11(3):373.

50. Brosh-Nissimov T, Hussein K, Wiener-Well Y, et al. Hospitalized patients with severe coronavirus disease 2019 during the Omicron wave in Israel: benefits of a fourth vaccine dose. Clin Infect Dis. 2022;76(3):e234–e239.

51. Bulgaresi M, Rivasi G, Tarantini F, et al. Impact of SARS-CoV2 infection on mortality and hospitalization in nursing home residents during the “Omicron era”. Aging Clin Exp Res. 2023;35(6):1393–1399.

52. Chan M, Owens L, Gray ML, et al. Asthma and susceptibility to COVID-19 in Australian children during Alpha, Delta and Omicron waves of the COVID-19 pandemic. J Asthma Allergy. 2023;16(null):1139–1155.

53. Chen CL, Teng CK, Chen WC, et al. Clinical characteristics and treatment outcomes among the hospitalized elderly patients with COVID-19 during the late pandemic phase in central Taiwan. J Microbiol Immunol Infect. 2024;57(2):257–268.

54. Choi S-H, Choi JH, Lee JK, et al. Clinical characteristics and outcomes of children with SARS-CoV-2 infection during the Delta and Omicron variant-dominant periods in Korea. J Korean Med Sci. 2023;38(9):e65.

55. de Prost N, Audureau E, Heming N, et al. Clinical phenotypes and outcomes associated with SARS-CoV-2 variant Omicron in critically ill French patients with COVID-19. Nat Commun. 2022;13(1):6025.

56. Drummond PD, de Salles DB, de Souza NSH, et al. Profile and outcomes of hospitalized COVID-19 patients during the prevalence of the Omicron variant according to the Brazilian regions: a retrospective cohort study from 2022. Vaccines. 2023;11(10):1568.

57. Elamin MY, Maslamani YA, Alsheikh FA, et al. Impact of vaccination on morbidity and mortality in adults hospitalized with COVID-19 during the Omicron wave in the Jazan Region, Saudi Arabia. Saudi Med J. 2024;45(2):179–187.

58. Favia G, Barile G, Tempesta A, et al. Relationship between oral lesions and severe SARS-CoV-2 infection in intensive care unit patients. Oral Dis. 2024;30(3):1296–1303.

59. Finkas LK, Ramesh N, Block LS, et al. Asthma and COVID-19 outcomes: a prospective study in a large health care delivery system. J Asthma Allergy. 2023;16:1041–1051.

60. Flacco ME, Acuti Martellucci C, Soldato G, et al. Predictors of SARS-CoV-2 infection and severe and lethal COVID-19 after three years of follow-up: a population-wide study. Viruses. 2023;15(9):1794.

61. Flisiak R, Zarębska-Michaluk D, Dobrowolska K, et al. Change in the clinical picture of hospitalized patients with COVID-19 between the early and late period of dominance of the Omicron SARS-CoV-2 variant. J Clin Med. 2023;12(17):5572.

62. Gazit S, Saciuk Y, Perez G, et al. Short term, relative effectiveness of four doses versus three doses of BNT162b2 vaccine in people aged 60 years and older in Israel: retrospective, test negative, case-control study. BMJ. 2022;377:e071113.

63. Grannec F, Meddeb L, Tissot-Dupont H, Gentile S, Brouqui P. Pre-hospital management of patients with COVID-19 and the impact on hospitalization. Medicina. 2023;59(8):1440.

64. Griggs EP, Mitchell PK, Lazariu V, et al. Clinical epidemiology and risk factors for critical outcomes among vaccinated and unvaccinated adults hospitalized with COVID-19-VISION Network, 10 states, June 2021-March 2023. Clin Infect Dis. 2024;78(2):338–348.

65. Guo Y, Guo Y, Ying H, et al. In-hospital adverse outcomes and risk factors among chronic kidney disease patients infected with the omicron variant of SARS-CoV-2: a single-center retrospective study. BMC Infect Dis. 2023;23(1):698.

66. Helmy MA, Milad LM, Hasanin AM, et al. Parasternal intercostal thickening at hospital admission: a promising indicator for mechanical ventilation risk in subjects with severe COVID-19. J Clin Monit Comput. 2023;37(5):1287–1293.

67. Hippisley-Cox J, Khunti K, Sheikh A, Nguyen-Van-Tam JS, Coupland CAC. Risk prediction of covid-19 related death or hospital admission in adults testing positive for SARS-CoV-2 infection during the omicron wave in England (QCOVID4): cohort study. BMJ. 2023;381:e072976.

68. Jamaati H, Karimi S, Ghorbani F, et al. Effectiveness of different vaccine platforms in reducing mortality and length of ICU stay in severe and critical cases of COVID-19 in the Omicron variant era: a national cohort study in Iran. J Med Virol. 2023;95(3):e28607.

69. Karageorgou V, Papaioannou AI, Kallieri M, et al. Patients hospitalized for COVID-19 in the periods of Delta and Omicron variant dominance in Greece: determinants of severity and mortality. J Clin Med. 2023;12(18):5904.

70. Kim SH, Kim T, Choi H, Shin TR, Sim YS. Clinical outcome and prognosis of a nosocomial outbreak of COVID-19. J Clin Med. 2023;12(6):2279.

71. Klein EY, Fall A, Norton JM, et al. Severity outcomes associated with SARS-CoV-2 XBB variants, an observational analysis. J Clin Virol. 2023;165:105500.

72. Lee CM, Kim M, Park SW, et al. Clinical outcomes and immunological features of COVID-19 patients receiving B-cell depletion therapy during the Omicron era. Infect Dis (Lond). 2024;56(2):116–127.

73. Lewnard JA, Hong VX, Patel MM, et al. Clinical outcomes associated with SARS-CoV-2 Omicron (B.1.1.529) variant and BA.1/BA.1.1 or BA.2 subvariant infection in Southern California. Nat Med. 2022;28(9):1933–1943.

74. Li D-J, Zhou C-C, Huang F, Shen F-M, Li Y-C. Clinical features of omicron SARS-CoV-2 variants infection associated with co-infection and ICU-acquired infection in ICU patients. Front Public Health. 2024;11:1320340.

75. Li H, Jia X, Wang Y, et al. Differences in the severity and mortality risk factors for patients hospitalized for COVID-19 pneumonia between the early wave and the very late stage of the pandemic. Front Med (Lausanne). 2023;10:1238713.

76. Liu Y, Chen D, Li J, et al. Metabolic syndrome is associated with poor Omicron infection prognosis while inactivated vaccine improves the outcome of Coronavirus Disease 2019 among Chinese inhabitants: a retrospective observational study from a Chinese municipality. Vaccines. 2023;11(10):1554.

77. Liu Y, Qi Z, Bai M, et al. Combination of chest computed tomography value and clinical laboratory data for the prognostic risk evaluation of patients with COVID-19. Int J Gen Med. 2023;16:3829–3842.

78. Lu G, Zhang Y, Zhang H, et al. Geriatric risk and protective factors for serious COVID-19 outcomes among older adults in Shanghai Omicron wave. Emerg Microbes Infect. 2022;11(1):2045–2054.

79. Manchanda V, Mitra S, Rafique I, et al. Is Omicron really mild? – Comparative analysis of comorbidities and disease outcomes associated with SARS-CoV-2 Omicron (B.1.1.529) and Delta (B.1.617.2) variants. Indian J Med Microbiol. 2023;45:100391.

80. Mayer C, Woo MS, Brehm TT, et al. History of cerebrovascular disease but not dementia increases the risk for secondary vascular events during SARS-CoV-2 infection with presumed Omicron variant: a retrospective observational study. Eur J Neurol. 2023;30(8):2297–2304.

81. McNeil T, Zhang F, Moffatt S, Emeto TI, Tucker E. Nosocomial COVID-19 infection in the era of vaccination and antiviral therapy. Intern Med J. 2024;54(3):374–381.

82. Mendoza-Cano O, Trujillo X, Ríos-Silva M, et al. Association between vaccination status for COVID-19 and the risk of severe symptoms during the endemic phase of the disease. Vaccines. 2023;11(10):1512.

83. Mizrahi Reuveni M, Kertes J, Shapiro Ben David S, et al. Risk stratification model for severe COVID-19 disease: a retrospective cohort study. Biomedicines. 2023;11(3):767.

84. Mosallami Aghili SM, Khoshfetrat M, Asgari A, et al. Association of echocardiographic findings with in-hospital mortality of COVID-19 patients and their changes in one-month follow-up; a cohort study. Arch Acad Emerg Med. 2022;10(1):e85.

85. Mumtaz A, Rehman E, Rahaman MA, Rehman S. Inflammatory biomarkers and cardiac injury in COVID-19 patients. Front Public Health. 2022;10:1024535.

86. O’Leary AL, Wattengel BA, Carter MT, Drye AF, Mergenhagen KA. Risk factors associated with mortality in hospitalized patients with laboratory confirmed SARS-CoV-2 infection during the period of omicron (B.1.1.529) variant predominance. Am J Infect Control. 2023;51(6):603–606.

87. Parajuli P, Sabo R, Alsaadawi R, et al. Fibrosis-4 (FIB-4) index as a predictor for mechanical ventilation and 30-day mortality across COVID-19 variants. J Clin Transl Sci. 2023;7(1):e213.

88. Parra-Bracamonte GM, Lopez-Villalobos N, Velazquez MA, et al. Comparative analysis of risk factors for COVID-19 mortality before, during and after the vaccination programme in Mexico. Public Health. 2023;215:94–99.

89. Patton MJ, Orihuela CJ, Harrod KS, et al. COVID-19 bacteremic co-infection is a major risk factor for mortality, ICU admission, and mechanical ventilation. Crit Care. 2023;27(1):34.

90. Radhakrishnan N, Liu M, Idowu B, et al. Comparison of the clinical characteristics of SARS-CoV-2 Delta (B.1.617.2) and Omicron (B.1.1.529) infected patients from a single hospitalist service. BMC Infect Dis. 2023;23(1):747.

91. Russell SL, Klaver BRA, Harrigan SP, et al. Clinical severity of Omicron subvariants BA.1, BA.2, and BA.5 in a population-based cohort study in British Columbia, Canada. J Med Virol. 2023;95(1):e28423.

92. Sardinha DM, Ferreira ALdS, Guimarães RJdPSe, Lima KVB, Lima LNGC. Clinical characteristics and outcomes among vaccinated and unvaccinated patients with cardiovascular disease who were hospitalized for COVID-19 in Brazil: retrospective cohort. Vaccines. 2023;11(4):861.

93. Shi HJ, Yang J, Eom JS, et al. Clinical characteristics and risk factors for mortality in critical COVID-19 patients aged 50 years or younger during Omicron wave in Korea: comparison with patients older than 50 years of age. J Korean Med Sci. 2023;38(28):e217.

94. Simmons AE, Amoako A, Grima AA, et al. Vaccine effectiveness against hospitalization among adolescent and pediatric SARS-CoV-2 cases between May 2021 and January 2022 in Ontario, Canada: a retrospective cohort study. PLoS One. 2023;18(3):e0283715.

95. Skarbinski J, Wood MS, Chervo TC, et al. Risk of severe clinical outcomes among persons with SARS-CoV-2 infection with differing levels of vaccination during widespread Omicron (B.1.1.529) and Delta (B.1.617.2) variant circulation in Northern California: a retrospective cohort study. Lancet Reg Health Am. 2022;12:100297.

96. Sonaglioni A, Lombardo M, Albini A, et al. Charlson comorbidity index, neutrophil-to-lymphocyte ratio and undertreatment with renin-angiotensin-aldosterone system inhibitors predict in-hospital mortality of hospitalized COVID-19 patients during the omicron dominant period. Front Immunol. 2022;13:958418.

97. Stepanova M, Lam B, Younossi E, et al. The impact of variants and vaccination on the mortality and resource utilization of hospitalized patients with COVID-19. BMC Infect Dis. 2022;22(1):702.

98. Tsujimoto Y, Kobayashi M, Oku T, et al. Outcomes in novel hospital-at-home model for patients with COVID-19: a multicentre retrospective cohort study. Fam Pract. 2023;40(5-6):662–670.

99. Vo AD, La J, Wu JT, et al. Factors associated with severe COVID-19 among vaccinated adults treated in US veterans affairs hospitals. JAMA Netw Open. 2022;5(10):e2240037.

100. Wang X, Zein J, Ji X, Lin DY. Impact of vaccination, prior infection, and therapy on Omicron infection and mortality. J Infect Dis. 2023;227(8):970–976.

101. Ward IL, Robertson C, Agrawal U, et al. Risk of COVID-19 death in adults who received booster COVID-19 vaccinations in England. Nat Commun. 2024;15(1):398.

102. Xin S, Chen W, Yu Q, Gao L, Lu G. Effect of the number of coronavirus disease 2019 (COVID-19) vaccination shots on the occurrence of pneumonia, severe pneumonia, and death in SARS-CoV-2-infected patients. Front Public Health. 2024;11:1330106.

103. Yang H, Wang Z, Zhang Y, et al. Clinical characteristics and factors for serious outcomes among outpatients infected with the Omicron subvariant BF.7. J Med Virol. 2023;95(8):e28977.

104. Ying-Hao P, Yuan-Yuan G, Hai-Dong Z, et al. Clinical characteristics and analysis of risk factors for disease progression of patients with SARS-CoV-2 Omicron variant infection: a retrospective study of 25207 cases in a Fangcang hospital. Front Cell Infect Microbiol. 2022;12:1009894.

105. Zhang Y, Han J, Sun F, et al. A practical scoring model to predict the occurrence of critical illness in hospitalized patients with SARS-CoV-2 omicron infection. Front Microbiol. 2022;13:1031231.

106. Zhao Q, Zheng B, Han B, et al. Is azvudine comparable to nirmatrelvir-ritonavir in real-world efficacy and safety for hospitalized patients with COVID-19? a retrospective cohort study. Infect Dis Ther. 2023;12(8):2087–2102.

107. Zhu Z, Cai J, Tang Q, et al. Circulating eosinophils associated with responsiveness to COVID-19 vaccine and the disease severity in patients with SARS-CoV-2 omicron variant infection. BMC Pulm Med. 2023;23(1):177.

