## Supplementary material for "Risk of Severe Outcomes From COVID-19 in Comorbid Populations in the Omicron Era: A Meta-analysis"

Supplemental Material

### 1. Supplemental eMethods

#### 1.1. Search Strategy: Burden/Risk of COVID-19 Infection

The following databases were used for searches: Embase, MEDLINE, PubMed, Europe PMC (including MedRix and bioRxiv preprints), Latin American and Caribbean Health Sciences Literature (LILACS), Cochrane COVID-19 Study Register, and the WHO COVID-19 Database. Rolling searches were performed each month for studies available between 1 January 2022 and 13 March 2024, though the WHO COVID-19 database was no longer available before the end date (ceased June 2023). Hand searching was also performed, in which bibliographies of identified studies were checked, and for particularly relevant studies, citation tracking (Google Scholar) was performed.

Searching was carried out using two separate search approaches:

- Search 1: Burden/risk of COVID-19 infection
- Search 2: Protective effect of vaccination on burden/risk of COVID-19 infection

The following table includes the number of results for each database:

| **Database** | **Host** | **Date range** | **Date searched** | **S1: Burden results** | **S2: Vaccine results** |
| --- | --- | --- | --- | --- | --- |
| **January 2024 searches** |  |  |  |  |  |
| Embase | Ovid | 2022-2023/12/29 | 2.1.24 | 4172 | 1628 |
| Medline ALL | Ovid | 2022-2024/02/02 | 3.1.24 | 3385 | 825 |
| PubMed | NLM | 2022-2024/01/03 | 3.1.24 | 1047 | 281 |
| LILACS | www | 2022-2024/01/04 | 4.1.24 | 38 | 3 |
| Cochrane COVID Register | www | 2022-2024/01/04 | 4.1.24 | 3814 | 1792 |
| Europe PMC, includes BioRxiv & medRxiv | www | 2022-2024/01/04 | 4.1.24 | 10 | 10 |
| WHO COVID-19 database | www | 2022-2024/01/03 | 3.1.24 | 2268 | 477 |
| **January 2024 subtotal** |  |  |  | **14734** | **5016** |
| **February 2024 update searches** |  |  |  |  |  |
| Embase | Ovid | 2022-2024/02/01 | 1.2.24 | 460 | 110 |
| Medline ALL | Ovid | 2022-2024/02/01 | 1.2.24 | 363 | 51 |
| PubMed | NLM | 2022-2024/02/01 | 1.2.24 | 92 | 20 |
| LILACS | www | 2022-2024/02/01 | 2.2.24 | 3 | 0 |
| Cochrane COVID Register | www | 2022-2024/02/01 | 2.2.24 | 0 | 1 |
| Europe PMC, includes BioRxiv & medRxiv | www | 2022-2024/02/01 | 1.2.24 | 0 | 1 |
| WHO COVID-19 database* | www | Not updated | - | - | - |
| **February 2024 subtotal** |  |  |  | **918** | **183** |
| **March 2024 update searches** |  |  |  |  |  |
| Embase | Ovid | 2022-2024/03/01 | 1.3.24 | 404 | 86 |
| 68 | Ovid | 2022-2024/03/01 | 1.3.24 | 386 | 63 |
| PubMed | NLM | 2022-2024/03/01 | 12.3.24 | 68 | 17 |
| LILACS | www | 2022-2024/03/01 | 13.3.24 | 0 | 0 |
| Cochrane COVID Register | www | 2022-2024/03/01 | 13.3.24 | 59 | 0 |
| Europe PMC, includes BioRxiv & medRxiv | www | 2022-2024/03/01 | 1.3.24 | 3 | 0 |
| WHO COVID-19 database* | www | Not updated | - | - | - |
| **March 2024 update subtotal** |  |  |  | **920** | **166** |
| **Subtotal retrieved** |  |  |  | **16572** | **5365** |
| **Subtotal screened** |  |  |  | **9425** | **2167** |
| **Subtotal duplicates** |  |  |  | **7147** | **3198** |
| **Combined total retrieved** |  |  |  | **21937** | |
| **Combined total screened** |  |  |  | **11592** | |
| **Combined total duplicates** |  |  |  | **10345** | |

***** WHO COVID-19 database ceased in June 2023, therefore update searches were not necessary.

##### 1.1.1. Search 1: Burden/risk of COVID-19 Infection Search Strategies

**Embase (Ovid): 2022-2023/12/29**

Searched 2.1.24

1	exp coronavirus disease 2019/	374674

2	sars-related coronavirus/ or exp Severe acute respiratory syndrome coronavirus 2/	106902

3	(coronavirinae/ or betacoronavirus/ or coronavirus infection/) and (epidemic/ or pandemic/)	10615

4	(Coronavirus$ or "covid 19" or 2019-ncov).ti,ab,kw,kf,ot.	430070

5	(2019-ncov or 2019ncov or corona-virus$ or cov19 or cov-19 or 19nCoV or COVID19 or COVID2019 or "COVID-19 2019").ti,ab,kw,kf,ot.	15833

6	(ncov$ or "sars cov$" or sarscov$ or "sars coronavirus$" or coronovirus$ or corono$ virus$ or "19-nCoV$" or 19nCoV$).ti,ab,kw,kf,ot.	160670

7	(SARS2$ or "SARS-2$" or SARScoronavirus$ or SARS-coronavirus$ or SARScoronovirus$ or SARS-coronovirus$).ti,ab,kw,kf,ot.	2922

8	("HCoV-19$" or HCoV19$ or "HCoV-2019$" or HCoV2019$).ti,ab,kw,kf,ot.	70

9	("2019 novel$" or Ncov$).ti,ab,kw,kf,ot.	5843

10	("Severe Acute Respiratory Syndrome Coronavirus 2" or "Severe Acute Respiratory Syndrome Corona Virus 2").ti,ab,kw,kf,ot.	36797

11	or/1-10	496535

12	("2022" or "2023" or "2024").ti,ab,ot.	235450

13	11 and 12	26927

14	(strain or strains or variant$ or mutation$).ti,ab,ot,kf,kw.	2383716

15	11 and 14	36992

16	exp "SARS-CoV-2 (lineage B.1.1)"/	8191

17	exp "SARS-CoV-2 Omicron"/	7250

18	(omikron or Omicron or "B.1.1.529" or "B11529" or xbb$).af.	13566

19	or/16-18	14551

20	or/13,15,19	63058

21	Clinical study/	165138

22	Case control study/	211296

23	Family study/	25764

24	Longitudinal study/	203445

25	Retrospective study/	1544157

26	Prospective study/	899644

27	Randomized controlled trials/	267337

28	26 not 27	888668

29	Cohort analysis/	1096585

30	(Cohort adj (study or studies)).mp.	493516

31	(Case control adj (study or studies)).tw.	171865

32	(follow up adj (study or studies)).tw.	74874

33	(observational adj (study or studies)).tw.	264634

34	(epidemiologic$ adj (study or studies)).tw.	123898

35	(cross sectional adj (study or studies)).tw.	356386

36	or/21-25,28-35	4072059

37	animal/ or animal experiment/	4720382

38	(rat or rats or mouse or mice or murine or rodent or rodents or hamster or hamsters or pig or pigs or porcine or rabbit or rabbits or animal or animals or dogs or dog or cats or cow or bovine or sheep or ovine or monkey or monkeys).ti,ab,ot,hw.	7724982

39	or/37-38	7724982

40	human experiment/ or exp humans/	25973725

41	39 not (39 and 40)	5781960

42	36 not 41	4009600

43	limit 42 to yr="2022 -Current"	754329

44	43 not (letter or editorial or conference or "conference abstract" or "conference paper" or "conference review").pt.	555478

45	mortality/ or death/ or "cause of death"/ or survival rate/	1478645

46	(mortalit$ or death$ or fatal$ or survival).ti,ab,ot,kf,kw.	4124730

47	exp intensive care/ or exp intensive care unit/ or high dependency unit/	1066416

48	(close attention unit$ or intensive care or respiratory care unit or respiratory care units or special care unit or special care units or high dependency unit or high dependency units).ti,ab,ot,kf,kw.	298895

49	(ICU or ICUs or CCU or CCus or GICU or GICUs or HDU or HDUs or ITU or ITUs).ti,ab,ot,kf,kw.	187026

50	(Hospitaliz$ or Hospitalis$ or (hospital adj2 Admission$) or Critical Care).ti,ab,ot,kf,kw.	716741

51	hospitalization/	546684

52	("length of stay" or "duration of stay" or "extended stay" or "prolonged stay" or "hospital stay").ti,ab,ot,hw.	412206

53	("in-patient stay" or "inpatient stay" or "in-patient stays" or "inpatient stays").ti,ab,ot,hw.	7835

54	or/45-53	5826795

55	exp risk/ or exp risk factor/ or exp risk assessment/	3170934

56	hazard ratio/ or odds ratio/	99395

57	mortality risk/	46876

58	life table/	5253

59	(risk$ or danger$ or association$ or peril or jeopard$ or threat$ or chance or chances or hazard$ or gamble$ or probabilit$ or "at stake" or endanger$ or associat$ or likelihood$ or possibilit$ or correlation$ or odds).ti,ab,ot,kf,kw.	12507127

60	(morbidit$ or comorbid$ or co-morbid$).ti,ab,ot,kf,kw.	1138678

61	(incidence or prevalence or predict$ or prognosis).ti,ab,ot,kf,kw.	5507851

62	(HR or RR or aiRR or aOR or "adjusted OR").ti,ab,ot,kf,kw.	753007

63	or/55-62	15711755

**64	20 and 54 and 63 and 44	4172**

*The Embase strategy was updated on 1.2.24 (460 records) and 1.3.24 (404 records).*

COVID facet based on terms from:

World Health Organization (26 May 2021) WHO COVID-19 Database Search Strategy. Systematic search of the COVID-19 literature performed Monday through Friday for the WHO Database. Search strategy as of 26 May 2021. Searches performed by Tomas Allen, Kavita Kothari, and Martha Knuth. Available from: <https://www.who.int/docs/default-source/coronaviruse/who-covid-19-database/who-covid-19_sources_searchstrategy_20210526.pdf?sfvrsn=65209cc2_5>

Canadian Agency for Drugs and Technologies in Health (2.9.21) CADTH COVID-19 Search Strings: COVID-19 — EMBASE (Internet). Available from: <https://covid.cadth.ca/literature-searching-tools/cadth-covid-19-search-strings/>

NICE (18 December 2020) [accessed 17.8.21] COVID-19 rapid guideline: managing the long-term effects of COVID-19 [NG188]. Search history record [PDF]. NICE: London. Available from: <https://www.nice.org.uk/guidance/ng188/evidence/search-strategies-pdf-8957634445>

Observational study design filter adapted from:

Scottish Intercollegiate Guidelines Network (SIGN). Search filters: observational studies. Embase. Edinburgh: SIGN, Last Modified 24/04/17 Available from: <https://www.sign.ac.uk/what-we-do/methodology/search-filters>

Observational Studies — Embase. In: CADTH Search Filters Database. Ottawa: CADTH; 2023 [accessed 7.11.23]: <https://searchfilters.cadth.ca/link/37>

**Medline ALL (Ovid): 2022-2024/01/02**

Searched 3.1.24

1	exp COVID-19/	250794

2	exp Severe acute respiratory syndrome-related coronavirus/	166686

3	coronaviridae/ or exp coronavirus/	178833

4	Betacoronavirus/	33277

5	Coronavirus Infections/	46094

6	or/3-5	185838

7	epidemics/ or pandemics/	139078

8	Disease Outbreaks/	92374

9	or/7-8	226884

10	6 and 9	83151

11	(Coronavirus$ or "covid 19" or 2019-ncov).ti,ab,kw,kf,ot.	385791

12	(2019-ncov or 2019ncov or corona-virus$ or cov19 or cov-19 or 19nCoV or COVID19 or COVID2019 or "COVID-19 2019").ti,ab,kw,kf,ot.	10202

13	(ncov$ or "sars cov$" or sarscov$ or "sars coronavirus$" or coronovirus$ or corono$ virus$ or "19-nCoV$" or 19nCoV$).ti,ab,kw,kf,ot.	140222

14	(SARS2$ or "SARS-2$" or SARScoronavirus$ or SARS-coronavirus$ or SARScoronovirus$ or SARS-coronovirus$).ti,ab,kw,kf,ot.	2635

15	("HCoV-19$" or HCoV19$ or "HCoV-2019$" or HCoV2019$).ti,ab,kw,kf,ot.	66

16	("2019 novel$" or Ncov$).ti,ab,kw,kf,ot.	5236

17	("Severe Acute Respiratory Syndrome Coronavirus 2" or "Severe Acute Respiratory Syndrome Corona Virus 2").ti,ab,kw,kf,ot.	36907

18	or/1-2,10-17	416995

19	("2022" or "2023" or "2024").ti,ab,ot.	153808

20	18 and 19	17876

21	(strain or strains or variant$ or mutation$).ti,ab,ot,kf,kw.	1916135

22	18 and 21	32095

23	(omikron or Omicron or "B.1.1.529" or "B11529" or xbb$).af.	9846

24	or/20,22-23	48695

25	mortality/ or "cause of death"/ or fatal outcome/ or hospital mortality/ or survival rate/	388730

26	Death/	20522

27	(mortalit$ or death$ or fatal$ or survival).ti,ab,ot,kf,kw.	2881687

28	exp Critical Care/	67547

29	exp Intensive Care Units/	107853

30	(close attention unit$ or intensive care or respiratory care unit or respiratory care units or special care unit or special care units or high dependency unit or high dependency units).ti,ab,ot,kf,kw.	202827

31	(ICU or ICUs or CCU or CCus or GICU or GICUs or HDU or HDUs or ITU or ITUs).ti,ab,ot,kf,kw.	95415

32	(Hospitaliz$ or Hospitalis$ or (hospital adj2 Admission$) or Critical Care).ti,ab,ot,kf,kw.	436800

33	Hospitalization/	137866

34	("length of stay" or "duration of stay" or "extended stay" or "prolonged stay" or "hospital stay").ti,ab,ot,hw.	220112

35	"Length of Stay"/	103573

36	("in-patient stay" or "inpatient stay" or "in-patient stays" or "inpatient stays").ti,ab,ot,hw.	3851

37	or/25-36	3560422

38	exp risk/	1397708

39	odds ratio/ or proportional hazards models/	185114

40	life tables/	6619

41	(risk$ or danger$ or association$ or peril or jeopard$ or threat$ or chance or chances or hazard$ or gamble$ or probabilit$ or "at stake" or endanger$ or associat$ or likelihood$ or possibilit$ or correlation$ or odds).ti,ab,ot,kf,kw.	9312430

42	(morbidit$ or comorbid$ or co-morbid$).ti,ab,ot,kf,kw.	716968

43	(incidence or prevalence or predict$ or prognosis).ti,ab,ot,kf,kw.	3993128

44	(HR or RR or aiRR or aOR or "adjusted OR").ti,ab,ot,kf,kw.	471257

45	or/38-44	11628532

46	Epidemiologic studies/	9457

47	exp case control studies/	1470108

48	exp cohort studies/	2556574

49	Case control.tw.	158730

50	Longitudinal.tw.	335439

51	Retrospective.tw.	782068

52	Cross sectional.tw.	540774

53	Cross-sectional studies/	487981

54	Observational Studies as Topic/	9294

55	(observational adj3 (study or studies or design or analysis or analyses)).ti,ab,kf.	234202

56	((follow up or followup) adj7 (study or studies or design or analysis or analyses)).ti,ab,kf.	175238

57	(epidemiologic$ adj3 (study or studies or design or analysis or analyses)).ti,ab,kf.	119921

58	((longterm or (long adj term)) adj7 (study or studies or design or analysis or analyses or data)).ti,ab,kf.	160631

59	(cohort$ adj3 (study or studies or design or analysis or analyses or data)).ti,ab,kf.	420054

60	(prospective adj7 (study or studies or design or analysis or analyses)).ti,ab,kf.	551875

61	or/46-60	4243864

62	animals/ not (animals/ and humans/)	5148855

63	61 not 62	4139882

64	63 not (case reports or clinical conference or comment or editorial or letter).pt.	3929840

65	limit 64 to yr="2022 -Current"	538850

**66	24 and 37 and 45 and 65	3385**

*The Medline ALL strategy was updated on 1.2.24 (363 records) and 1.3.24 (363 records).*

COVID facet based on terms from:

World Health Organization (26 May 2021) WHO COVID-19 Database Search Strategy. Systematic search of the COVID-19 literature performed Monday through Friday for the WHO Database. Search strategy as of 26 May 2021. Searches performed by Tomas Allen, Kavita Kothari, and Martha Knuth. Available from: <https://www.who.int/docs/default-source/coronaviruse/who-covid-19-database/who-covid-19_sources_searchstrategy_20210526.pdf?sfvrsn=65209cc2_5>

Canadian Agency for Drugs and Technologies in Health (2.9.21) CADTH COVID-19 Search Strings: COVID-19 — EMBASE (Internet). Available from: <https://covid.cadth.ca/literature-searching-tools/cadth-covid-19-search-strings/>

NICE (18 December 2020) [accessed 17.8.21] COVID-19 rapid guideline: managing the long-term effects of COVID-19 [NG188]. Search history record [PDF]. NICE: London. Available from: <https://www.nice.org.uk/guidance/ng188/evidence/search-strategies-pdf-8957634445>

Observational study design filter adapted from:

Scottish Intercollegiate Guidelines Network (SIGN). Search filters: observational studies. Medline. Edinburgh: SIGN, Last Modified 24/04/17 Available from: <https://www.sign.ac.uk/what-we-do/methodology/search-filters/>

Observational Studies — Medline. In: CADTH Search Filters Database. Ottawa: CADTH; 2023 [accessed 7.11.23]: <https://searchfilters.cadth.ca/list?q=topic%3A%22Observational%20studies%22&p=1&ps=&sort=title_sort%20asc>

**PubMed (NLM): 2022-2024/01/03**

Searched 3.1.24

[**https://pubmed.ncbi.nlm.nih.gov/**](https://pubmed.ncbi.nlm.nih.gov/)

**35	#13 AND #30 AND #34 AND #24 AND #25	1,047**

34	#31 OR #32 OR #33	9,159,936

33	morbidity[Title/Abstract] OR comorbidity[Title/Abstract] OR "co-morbidity"[Title/Abstract] OR morbidities[Title/Abstract] OR comorbidities[Title/Abstract] OR "co-morbidities"[Title/Abstract] OR incidence[Title/Abstract] OR prevalence[Title/Abstract] OR predict[Title/Abstract] OR predicted[Title/Abstract] OR predictive[Title/Abstract] OR prognosis[Title/Abstract] OR prediction[Title/Abstract] OR predictions[Title/Abstract] OR HR[Title/Abstract] OR RR[Title/Abstract] OR aiRR[Title/Abstract] OR aOR[Title/Abstract] OR "adjusted OR"[Title/Abstract]	4,362,222

32	risk[Title/Abstract] OR risks[Title/Abstract] OR risky[Title/Abstract] OR danger[Title/Abstract] OR dangers[Title/Abstract] OR dangerous[Title/Abstract] OR association[Title/Abstract] OR associations[Title/Abstract] OR peril[Title/Abstract] OR jeopardy[Title/Abstract] OR jeopardise[Title/Abstract] OR jeopardize[Title/Abstract] OR threat[Title/Abstract] OR threats[Title/Abstract] OR chance[Title/Abstract] OR chances[Title/Abstract] OR hazard[Title/Abstract] OR hazards[Title/Abstract] OR hazardous[Title/Abstract] OR gamble[Title/Abstract] OR gambles[Title/Abstract] OR gambling[Title/Abstract] OR probability[Title/Abstract] OR probabilities[Title/Abstract] OR "at stake"[Title/Abstract] OR endanger[Title/Abstract] OR endangers[Title/Abstract] OR endangered[Title/Abstract] OR endangering[Title/Abstract] OR associate[Title/Abstract] OR associates[Title/Abstract] OR associating[Title/Abstract] OR association[Title/Abstract] OR likelihood[Title/Abstract] OR likelihoods[Title/Abstract] OR possibility[Title/Abstract] OR possibilities[Title/Abstract] OR correlation[Title/Abstract] OR correlations[Title/Abstract] OR odds[Title/Abstract]	6,440,552

31	((("Risk"[Mesh]) OR "Odds Ratio"[Mesh:NoExp]) OR "Proportional Hazards Models"[Mesh:NoExp]) OR "Life Tables"[Mesh:NoExp]	1,481,626

30	#26 OR #27 OR #28 OR #29	3,581,605

29	Hospitalized[Title/Abstract] OR Hospitalised[Title/Abstract] OR Hospitalization[Title/Abstract] OR Hospitalisation[Title/Abstract] OR Hospitalizations[Title/Abstract] OR Hospitalisations[Title/Abstract] OR "hospital Admission"[Title/Abstract] OR "hospital Admissions"[Title/Abstract] OR "Critical Care"[Title/Abstract] OR "length of stay"[Title/Abstract] OR "duration of stay"[Title/Abstract] OR "extended stay"[Title/Abstract] OR "prolonged stay"[Title/Abstract] OR "hospital stay"[Title/Abstract] OR "in-patient stay"[Title/Abstract] OR "inpatient stay"[Title/Abstract] OR "in-patient stays"[Title/Abstract] OR "inpatient stays"[Title/Abstract]	565,867

28	mortality[Title/Abstract] OR death[Title/Abstract] OR deaths[Title/Abstract] OR fatal[Title/Abstract] OR fatality[Title/Abstract] OR fatalities[Title/Abstract] OR survival[Title/Abstract] OR "close attention unit"[Title/Abstract] OR "close attention units"[Title/Abstract] OR "intensive care"[Title/Abstract] OR "respiratory care unit"[Title/Abstract] OR "respiratory care units"[Title/Abstract] OR "special care unit"[Title/Abstract] OR "special care units"[Title/Abstract] OR "high dependency unit"[Title/Abstract] OR "high dependency units"[Title/Abstract] OR ICU[Title/Abstract] OR ICUs[Title/Abstract] OR CCU[Title/Abstract] OR CCus[Title/Abstract] OR GICU[Title/Abstract] OR GICUs[Title/Abstract] OR HDU[Title/Abstract] OR HDUs[Title/Abstract] OR ITU[Title/Abstract] OR ITUs[Title/Abstract]	3,013,975

27	((("Length of Stay"[Mesh:NoExp]) OR "Critical Care"[Mesh]) OR "Intensive Care Units"[Mesh]) OR "Hospitalization"[Mesh]	429,629

26	((((("Mortality"[Mesh:NoExp]) OR "Cause of Death"[Mesh:NoExp]) OR "Fatal Outcome"[Mesh:NoExp]) OR "Hospital Mortality"[Mesh:NoExp]) OR "Survival Rate"[Mesh:NoExp]) OR "Death"[Mesh:NoExp]	408,041

25	pubstatusaheadofprint OR publisher[sb] OR pubmednotmedline[sb]	5,627,356

24	#22 AND #23	493,476

23	("2022/01/01"[Date - Publication] : "3000"[Date - Publication])	3,253,097

22	#20 NOT #21	3,739,291

21	LETTER[Publication Type] OR EDITORIAL[Publication Type] OR COMMENT[Publication Type]	2,213,615

20	#16 NOT #19	3,825,199

19	#18 NOT (#18 AND #17)	3,757,858

18	rat[tiab] or rats[tiab] or mouse[tiab] or mice[tiab] or murine[tiab] or rodent[tiab] or rodents[tiab] or hamster[tiab] or hamsters[tiab] or pig[tiab] or pigs[tiab] or porcine[tiab] or rabbit[tiab] or rabbits[tiab] or animal[tiab] or animals[tiab] or dogs[tiab] or dog[tiab] or cats[tiab] or cow[tiab] or bovine[tiab] or sheep[tiab] or ovine[tiab] or monkey[tiab] or monkeys[tiab]	4,715,684

17	Human*[tiab]	3,295,660

16	#14 OR #15	3,914,514

15	((("Epidemiologic Studies"[Mesh:NoExp]) OR "Case-Control Studies"[Mesh]) OR "Cohort Studies"[Mesh]) OR "Cross-Sectional Studies"[Mesh:NoExp]	3,195,884

14	"Case control"[Title/Abstract] OR "cohort study"[Title/Abstract] OR "cohort studies"[Title/Abstract] OR "cohort analysis"[Title/Abstract] OR "cohort analyses"[Title/Abstract] OR "follow up study"[Title/Abstract] OR "follow up studies"[Title/Abstract] OR "observational study"[Title/Abstract] OR "observational studies"[Title/Abstract] OR Longitudinal[Title/Abstract] OR Retrospective[Title/Abstract] OR "Cross sectional"[Title/Abstract]	2,068,573

13	#9 OR #11 OR #12	47,832

12	omikron[Title/Abstract] OR Omicron[Title/Abstract] OR "B.1.1.529"[Title/Abstract] OR "B11529"[Title/Abstract] OR xbb[Title/Abstract]	9,468

11	#7 AND #10	31,553

10	strain[Title/Abstract] OR strains[Title/Abstract] OR variant[Title/Abstract] OR variants[Title/Abstract] OR mutation[Title/Abstract] OR mutations[Title/Abstract]	1,893,774

9	#7 AND #8	17,832

8	"2022"[Title/Abstract] OR "2023"[Title/Abstract]	158,464

7	#1 OR #4 OR #5 OR #6	412,676

6	"COVID-19 2019"[Title/Abstract] OR ncov[Title/Abstract] OR "sars cov"[Title/Abstract] OR sarscov[Title/Abstract] OR "sars coronavirus"[Title/Abstract] OR coronovirus[Title/Abstract] OR "corono virus"[Title/Abstract] OR "19-nCoV"[Title/Abstract] OR "19nCoV"[Title/Abstract] OR "SARS2"[Title/Abstract] OR "SARS-2"[Title/Abstract] OR SARScoronavirus[Title/Abstract] OR "SARS-coronavirus"[Title/Abstract] OR SARScoronovirus[Title/Abstract] OR "SARS-coronovirus"[Title/Abstract] OR "HCoV-19"[Title/Abstract] OR "HCoV19"[Title/Abstract] OR "HCoV-2019"[Title/Abstract] OR "HCoV2019"[Title/Abstract] OR "2019 novel"[Title/Abstract] OR "Severe Acute Respiratory Syndrome Coronavirus 2"[Title/Abstract] OR "Severe Acute Respiratory Syndrome Corona Virus 2"[Title/Abstract]	141,123

5	"Coronavirus"[Title/Abstract] OR "covid 19"[Title/Abstract] OR "2019-ncov"[Title/Abstract] OR "2019-ncov"[Title/Abstract] OR "2019ncov"[Title/Abstract] OR "corona-virus"[Title/Abstract] OR "cov19"[Title/Abstract] OR "cov-19"[Title/Abstract] OR "19nCoV"[Title/Abstract] OR "COVID19"[Title/Abstract] OR "COVID2019"[Title/Abstract]	382,642

4	#2 AND #3	83,020

3	(("Epidemics"[Mesh:NoExp]) OR "Pandemics"[Mesh:NoExp]) OR "Disease Outbreaks"[Mesh:NoExp]	226,548

2	((("Coronaviridae"[Mesh:NoExp]) OR "Coronavirus"[Mesh]) OR "Betacoronavirus"[Mesh:NoExp]) OR "Coronavirus Infections"[Mesh:NoExp]	185,366

1	("COVID-19"[Mesh]) OR "Severe acute respiratory syndrome-related coronavirus"[Mesh]	258,558

*The PubMed strategy was updated on 1.2.24 (92 records) and 1.3.24 (68 records).*

COVID facet based on terms from:

World Health Organization (26 May 2021) WHO COVID-19 Database Search Strategy. Systematic search of the COVID-19 literature performed Monday through Friday for the WHO Database. Search strategy as of 26 May 2021. Searches performed by Tomas Allen, Kavita Kothari, and Martha Knuth. Available from: <https://www.who.int/docs/default-source/coronaviruse/who-covid-19-database/who-covid-19_sources_searchstrategy_20210526.pdf?sfvrsn=65209cc2_5>

Canadian Agency for Drugs and Technologies in Health (2.9.21) CADTH COVID-19 Search Strings: COVID-19 — EMBASE (Internet). Available from: <https://covid.cadth.ca/literature-searching-tools/cadth-covid-19-search-strings/>

NICE (18 December 2020) [accessed 17.8.21] COVID-19 rapid guideline: managing the long-term effects of COVID-19 [NG188]. Search history record [PDF]. NICE: London. Available from: <https://www.nice.org.uk/guidance/ng188/evidence/search-strategies-pdf-8957634445>

*Observational study design filter adapted from:*

Scottish Intercollegiate Guidelines Network (SIGN). Search filters: observational studies. Medline. Edinburgh: SIGN, Last Modified 24/04/17 Available from: <https://www.sign.ac.uk/what-we-do/methodology/search-filters/>

*PubMed limit:*

Duffy S, de Kock S, Misso K, Noake C, Ross J, Stirk L. Supplementary searches of PubMed to improve currency of MEDLINE and MEDLINE In-Process searches via Ovid. J Med Libr Assoc. 2016 Oct;104(4):309-312. doi: 10.3163/1536-5050.104.4.011. <https://www.ncbi.nlm.nih.gov/pmc/articles/PMC5079494/>

**Europe PMC, including medRxiv and bioRxiv preprints (Internet): up to 2024/01/04**

Searched 4.1.24

**<https://europepmc.org/>**

| **Search terms** | **Results** |
| --- | --- |
| (TITLE:"COVID-19" OR TITLE:"coronavirus" OR TITLE:"COVID" OR TITLE:"NCOV" OR TITLE:"SARS-CoV-2") AND (TITLE:"risk" OR TITLE:"risks" OR TITLE:"hazard" OR TITLE:"hazards" OR TITLE:"probability" OR TITLE:" probabilities" OR TITLE:"likelihood" OR TITLE:"morbidity" OR TITLE:"comorbidity" OR TITLE:"predict" OR TITLE:"predictive" OR TITLE:"HR" OR TITLE:"RR" OR TITLE:"aiRR" OR TITLE:"aOR" OR TITLE:"adjusted OR") AND (TITLE:"2022" OR TITLE:"2023" OR TITLE:"strain" OR TITLE:"strains" OR TITLE:"variant" OR TITLE:"variants" OR TITLE:"mutation" OR TITLE:"mutations" OR TITLE:"omicron" OR TITLE:"omicron") | **89** |

*The Europe PMC strategy was updated on 1.2.24 (0 records) and 1.3.24 (3 records).*

**Latin American and Caribbean Health Sciences Literature (LILACS) (Internet): 2022-2024/01/07**

Searched 4.1.24

<https://search.bvsalud.org/portal/?lang=en>

Searched Title/Abstract/Subject

Limited to Observational studies

Limited to 2022-2024/01/04

Limited to LILACS only

((COVID OR NCOV OR coronavirus OR COVID19 OR "SARs-COV-2" OR Omicron OR omicron)) AND ((mortality OR death OR deaths OR fatal OR fatality OR fatalities OR survival OR "close attention unit" OR "close attention units" OR "intensive care" OR "respiratory care unit" OR "respiratory care units" OR "special care unit" OR "special care units" OR "high dependency unit" OR "high dependency units" OR ICU OR ICUs OR CCU OR CCus OR GICU OR GICUs OR HDU OR HDUs OR ITU OR ITUs OR Hospitalized OR Hospitalised OR Hospitalization OR Hospitalisation OR Hospitalizations OR Hospitalisations OR "hospital Admission" OR "hospital Admissions" OR "Critical Care" OR "length of stay" OR "duration of stay" OR "extended stay" OR "prolonged stay" OR "hospital stay" OR "in-patient stay" OR "inpatient stay" OR "in-patient stays" OR "inpatient stays") ) AND ((risk OR risks OR risky OR danger OR dangers OR dangerous OR association OR associations OR peril OR jeopardy OR jeopardise OR jeopardize OR threat OR threats OR chance OR chances OR hazard OR hazards OR hazardous OR gamble OR gambles OR gambling OR probability OR probabilities OR "at stake" OR endanger OR endangers OR endangered OR endangering OR associate OR associates OR associating OR association OR likelihood OR likelihoods OR possibility OR possibilities OR correlation OR correlations OR odds OR morbidity OR comorbidity OR "co-morbidity" OR morbidities OR comorbidities OR "co-morbidities" OR incidence OR prevalence OR predict OR predicted OR predictive OR prognosis OR prediction OR predictions OR HR OR RR OR aiRR OR aOR OR "adjusted OR")) AND ((2022 OR 2023 OR 2024 OR strain OR strains OR variant OR variants OR mutations OR mutation OR omikron OR Omicron OR "B.1.1.529" OR "B11529" OR xbb) )

**N = 38**

*The LILACS strategy was updated on 1.2.24 (3 records) and 1.3.24 (0 records).*

**Cochrane COVID-19 Study Register (www): 2022-2024/01/04**

Searched 4.1.24

<https://covid-19.cochrane.org/>

Limited to Observational cohort studies.

Limited 2022-2024/01/04

(mortality OR death OR deaths OR fatal OR fatality OR fatalities OR survival OR "close attention unit" OR "close attention units" OR "intensive care" OR "respiratory care unit" OR "respiratory care units" OR "special care unit" OR "special care units" OR "high dependency unit" OR "high dependency units" OR ICU OR ICUs OR CCU OR CCus OR GICU OR GICUs OR HDU OR HDUs OR ITU OR ITUs OR Hospitalized OR Hospitalised OR Hospitalization OR Hospitalisation OR Hospitalizations OR Hospitalisations OR "hospital Admission" OR "hospital Admissions" OR "Critical Care" OR "length of stay" OR "duration of stay" OR "extended stay" OR "prolonged stay" OR "hospital stay" OR "in-patient stay" OR "inpatient stay" OR "in-patient stays" OR "inpatient stays") AND (risk OR risks OR risky OR danger OR dangers OR dangerous OR association OR associations OR peril OR jeopardy OR jeopardise OR jeopardize OR threat OR threats OR chance OR chances OR hazard OR hazards OR hazardous OR gamble OR gambles OR gambling OR probability OR probabilities OR "at stake" OR endanger OR endangers OR endangered OR endangering OR associate OR associates OR associating OR association OR likelihood OR likelihoods OR possibility OR possibilities OR correlation OR correlations OR odds OR morbidity OR comorbidity OR "co-morbidity" OR morbidities OR comorbidities OR "co-morbidities" OR incidence OR prevalence OR predict OR predicted OR predictive OR prognosis OR prediction OR predictions OR HR OR RR OR aiRR OR aOR OR "adjusted OR")

AND (2022 OR 2023 OR 2024 OR strain OR strains OR variant OR variants OR mutations OR mutation OR omikron OR Omicron OR "B.1.1.529" OR "B11529" OR xbb) AND (cohort OR cohorts OR observational OR "follow up" OR followup OR epidemiologic OR epidemiological OR longterm OR "long term" OR longitudinal OR prospective)

**N = 3814**

*The Cochrane COVID-19 Study Register strategy was updated on 1.2.24 (0 records) and 1.3.24 (59 records****).***

**WHO COVID-19 (Internet): 2022-2024/01/03**

Searched 3.1.24

<https://search.bvsalud.org/global-literature-on-novel-coronavirus-2019-ncov/?lang=en>

Advanced search (title only)

(ti:(mortality OR death OR deaths OR fatal OR fatality OR fatalities OR survival OR "close attention unit" OR "close attention units" OR "intensive care" OR "respiratory care unit" OR "respiratory care units" OR "special care unit" OR "special care units" OR "high dependency unit" OR "high dependency units" OR ICU OR ICUs OR CCU OR CCus OR GICU OR GICUs OR HDU OR HDUs OR ITU OR ITUs OR Hospitalized OR Hospitalised OR Hospitalization OR Hospitalisation OR Hospitalizations OR Hospitalisations OR "hospital Admission" OR "hospital Admissions" OR "Critical Care" OR "length of stay" OR "duration of stay" OR "extended stay" OR "prolonged stay" OR "hospital stay" OR "in-patient stay" OR "inpatient stay" OR "in-patient stays" OR "inpatient stays")) AND (ti:(risk OR risks OR risky OR danger OR dangers OR dangerous OR association OR associations OR peril OR jeopardy OR jeopardise OR jeopardize OR threat OR threats OR chance OR chances OR hazard OR hazards OR hazardous OR gamble OR gambles OR gambling OR probability OR probabilities OR "at stake" OR endanger OR endangers OR endangered OR endangering OR associate OR associates OR associating OR association OR likelihood OR likelihoods OR possibility OR possibilities OR correlation OR correlations OR odds OR morbidity OR comorbidity OR "co-morbidity" OR morbidities OR comorbidities OR "co-morbidities" OR incidence OR prevalence OR predict OR predicted OR predictive OR prognosis OR prediction OR predictions OR HR OR RR OR aiRR OR aOR OR "adjusted OR"))

Limited to Observational and cohort studies.

Limited 2022-2023/06.

**N = 2268**

***** WHO COVID-19 database ceased on June 2023, therefore update searches were not necessary.

##### 1.1.2. Search 2: Protective Effect of Vaccination on Burden/Risk of COVID-19 Infection Search Strategies

**Embase (Ovid): 2022-2023/12/29**

Searched 2.1.24

1	exp coronavirus disease 2019/	374674

2	sars-related coronavirus/ or exp Severe acute respiratory syndrome coronavirus 2/	106902

3	(coronavirinae/ or betacoronavirus/ or coronavirus infection/) and (epidemic/ or pandemic/)	10615

4	(Coronavirus$ or "covid 19" or 2019-ncov).ti,ab,kw,kf,ot.	430070

5	(2019-ncov or 2019ncov or corona-virus$ or cov19 or cov-19 or 19nCoV or COVID19 or COVID2019 or "COVID-19 2019").ti,ab,kw,kf,ot.	15833

6	(ncov$ or "sars cov$" or sarscov$ or "sars coronavirus$" or coronovirus$ or corono$ virus$ or "19-nCoV$" or 19nCoV$).ti,ab,kw,kf,ot.	160670

7	(SARS2$ or "SARS-2$" or SARScoronavirus$ or SARS-coronavirus$ or SARScoronovirus$ or SARS-coronovirus$).ti,ab,kw,kf,ot.	2922

8	("HCoV-19$" or HCoV19$ or "HCoV-2019$" or HCoV2019$).ti,ab,kw,kf,ot.	70

9	("2019 novel$" or Ncov$).ti,ab,kw,kf,ot.	5843

10	("Severe Acute Respiratory Syndrome Coronavirus 2" or "Severe Acute Respiratory Syndrome Corona Virus 2").ti,ab,kw,kf,ot.	36797

11	or/1-10	496535

12	("2022" or "2023").ti,ab,ot.	235450

13	11 and 12	26927

14	(strain or strains or variant$ or mutation$).ti,ab,ot,kf,kw.	2383716

15	11 and 14	36992

16	exp "SARS-CoV-2 (lineage B.1.1)"/	8191

17	exp "SARS-CoV-2 Omicron"/	7250

18	(omikron or Omicron or "B.1.1.529" or "B11529" or xbb$).af.	13566

19	or/16-18	14551

20	or/13,15,19	63058

21	Clinical study/	165138

22	Case control study/	211296

23	Family study/	25764

24	Longitudinal study/	203445

25	Retrospective study/	1544157

26	Prospective study/	899644

27	Randomized controlled trials/	267337

28	26 not 27	888668

29	Cohort analysis/	1096585

30	(Cohort adj (study or studies)).mp.	493516

31	(Case control adj (study or studies)).tw.	171865

32	(follow up adj (study or studies)).tw.	74874

33	(observational adj (study or studies)).tw.	264634

34	(epidemiologic$ adj (study or studies)).tw.	123898

35	(cross sectional adj (study or studies)).tw.	356386

36	or/21-25,28-35	4072059

37	animal/ or animal experiment/	4720382

38	(rat or rats or mouse or mice or murine or rodent or rodents or hamster or hamsters or pig or pigs or porcine or rabbit or rabbits or animal or animals or dogs or dog or cats or cow or bovine or sheep or ovine or monkey or monkeys).ti,ab,ot,hw.	7724982

39	or/37-38	7724982

40	human experiment/ or exp humans/	25973725

41	39 not (39 and 40)	5781960

42	36 not 41	4009600

43	limit 42 to yr="2022 -Current"	754329

44	43 not (letter or editorial or conference or "conference abstract" or "conference paper" or "conference review").pt.	555478

45	mortality/ or death/ or "cause of death"/ or survival rate/	1478645

46	(mortalit$ or death$ or fatal$ or survival).ti,ab,ot,kf,kw.	4124730

47	exp intensive care/ or exp intensive care unit/ or high dependency unit/	1066416

48	(close attention unit$ or intensive care or respiratory care unit or respiratory care units or special care unit or special care units or high dependency unit or high dependency units).ti,ab,ot,kf,kw.	298895

49	(ICU or ICUs or CCU or CCus or GICU or GICUs or HDU or HDUs or ITU or ITUs).ti,ab,ot,kf,kw.	187026

50	(Hospitaliz$ or Hospitalis$ or (hospital adj2 Admission$) or Critical Care).ti,ab,ot,kf,kw.	716741

51	hospitalization/	546684

52	("length of stay" or "duration of stay" or "extended stay" or "prolonged stay" or "hospital stay").ti,ab,ot,hw.	412206

53	("in-patient stay" or "inpatient stay" or "in-patient stays" or "inpatient stays").ti,ab,ot,hw.	7835

54	exp risk/ or exp risk factor/ or exp risk assessment/	3170934

55	hazard ratio/ or odds ratio/	99395

56	mortality risk/	46876

57	(risk$ or danger$ or association$ or peril or jeopard$ or threat$ or chance or chances or hazard$ or gamble$ or probabilit$ or "at stake" or endanger$ or associat$ or likelihood$ or possibilit$ or correlation$ or odds or "vaccine effectiveness" or "immune evasion" or "immunity evasion").ti,ab,ot,kf,kw.	12518691

58	(morbidit$ or comorbid$ or co-morbid$).ti,ab,ot,kf,kw.	1138678

59	(incidence or prevalence or predict$ or prognosis).ti,ab,ot,kf,kw.	5507851

60	(HR or RR or aiRR or aOR or "adjusted OR").ti,ab,ot,kf,kw.	753007

61	disease severity assessment/ or covid-19 severity score/ or global severity index/ or quick covid-19 severity index/ or "severity of illness index"/	32588

62	morbidity/	416716

63	disease severity/ or critical illness/ or emergency status/ or urgency status/	772357

64	((severe or severity) adj3 (disease$ or infection$ or illness$ or complication$ or condition$ or illhealth$ or ill-health$ or sickness or virus or infirmity or affliction$)).ti,ab,ot,kf,kw.	423477

65	morbidit$.ti,ab,ot,kf,kw.	754095

66	or/45-65	18057098

67	Tozinameran/	11540

68	((pfizer$ or biontech) adj5 vaccin$).ti,ab,ot,kf,kw,hw,du,dy,tn,rn.	4443

69	(tozinameran or Comirnaty or "Pfizer-BioNTech" or "pf07302048" or "pf-07302048" or " BNT-162b2" or "BNT162b2" or Pidacmeran or "BNT162C2" or "BNT-162C2" or Abdavomeran or "BNT-162B1" or "BNT162B1" or "BNT162A1" or "BNT-162A1").ti,ab,ot,kf,kw,hw,du,dy,tn,rn.	15132

70	"Pfizer-BioNTech".mf.	947

71	"2417899-77-3".af.	12357

72	elasomeran/	6806

73	(Elasomeran or moderna or Spikevax or davesomeran or imelasomeran or andusomeran or "mRNA 1273" or "mRNA1273" or "mRNA-1273.211" or "mRNA 1273.211" or Spikevax or "M-1273" or "M1273" or "CX-024414" or "CX024414" or "TAK-919" or "TAK919").ti,ab,ot,kf,kw,hw,du,dy,tn,rn.	9771

74	("2430046-03-8" or "2457298-05-2").af.	6468

75	RNA vaccine/	5844

76	(("messenger RNA" or mrna or "RNA based") adj4 (vaccin$ or jab or jabs or shot or shots or immunis$ or immuniz$)).ti,ab,ot,kf,kw.	12887

77	or/67-76	25983

**78	and/20,44,66,77	1628**

*The Embase strategy was updated on 1.2.24 (110 records) and 1.3.24 (86 records).*

COVID facet based on terms from:

World Health Organization (26 May 2021) WHO COVID-19 Database Search Strategy. Systematic search of the COVID-19 literature performed Monday through Friday for the WHO Database. Search strategy as of 26 May 2021. Searches performed by Tomas Allen, Kavita Kothari, and Martha Knuth. Available from: <https://www.who.int/docs/default-source/coronaviruse/who-covid-19-database/who-covid-19_sources_searchstrategy_20210526.pdf?sfvrsn=65209cc2_5>

Canadian Agency for Drugs and Technologies in Health (2.9.21) CADTH COVID-19 Search Strings: COVID-19 — EMBASE (Internet). Available from: <https://covid.cadth.ca/literature-searching-tools/cadth-covid-19-search-strings/>

NICE (18 December 2020) [accessed 17.8.21] COVID-19 rapid guideline: managing the long-term effects of COVID-19 [NG188]. Search history record [PDF]. NICE: London. Available from: <https://www.nice.org.uk/guidance/ng188/evidence/search-strategies-pdf-8957634445>

*Observational study design filter adapted from:*

Scottish Intercollegiate Guidelines Network (SIGN). Search filters: observational studies. Embase. Edinburgh: SIGN, Last Modified 24/04/17 Available from: <https://www.sign.ac.uk/what-we-do/methodology/search-filters/>

Observational Studies - Embase. In: CADTH Search Filters Database. Ottawa: CADTH; 2023 [accessed 7.11.23]: <https://searchfilters.cadth.ca/link/37>

**Medline ALL (Ovid): 2022-2024/01/02**

Searched 2.1.24

1	exp COVID-19/	250794

2	exp Severe acute respiratory syndrome-related coronavirus/	166686

3	coronaviridae/ or exp coronavirus/	178833

4	Betacoronavirus/	33277

5	Coronavirus Infections/	46094

6	or/3-5	185838

7	epidemics/ or pandemics/	139078

8	Disease Outbreaks/	92374

9	or/7-8	226884

10	6 and 9	83151

11	(Coronavirus$ or "covid 19" or 2019-ncov).ti,ab,kw,kf,ot.	385791

12	(2019-ncov or 2019ncov or corona-virus$ or cov19 or cov-19 or 19nCoV or COVID19 or COVID2019 or "COVID-19 2019").ti,ab,kw,kf,ot.	10202

13	(ncov$ or "sars cov$" or sarscov$ or "sars coronavirus$" or coronovirus$ or corono$ virus$ or "19-nCoV$" or 19nCoV$).ti,ab,kw,kf,ot.	140222

14	(SARS2$ or "SARS-2$" or SARScoronavirus$ or SARS-coronavirus$ or SARScoronovirus$ or SARS-coronovirus$).ti,ab,kw,kf,ot.	2635

15	("HCoV-19$" or HCoV19$ or "HCoV-2019$" or HCoV2019$).ti,ab,kw,kf,ot.	66

16	("2019 novel$" or Ncov$).ti,ab,kw,kf,ot.	5236

17	("Severe Acute Respiratory Syndrome Coronavirus 2" or "Severe Acute Respiratory Syndrome Corona Virus 2").ti,ab,kw,kf,ot.	36907

18	or/1-2,10-17	416995

19	("2022" or "2023").ti,ab,ot.	153808

20	18 and 19	17876

21	(strain or strains or variant$ or mutation$).ti,ab,ot,kf,kw.	1916135

22	18 and 21	32095

23	(omikron or Omicron or "B.1.1.529" or "B11529" or xbb$).af.	9846

24	or/20,22-23	48695

25	Epidemiologic studies/	9457

26	exp case control studies/	1470108

27	exp cohort studies/	2556574

28	Case control.tw.	158730

29	Longitudinal.tw.	335439

30	Retrospective.tw.	782068

31	Cross sectional.tw.	540774

32	Cross-sectional studies/	487981

33	Observational Studies as Topic/	9294

34	(observational adj3 (study or studies or design or analysis or analyses)).ti,ab,kf.	234202

35	((follow up or followup) adj7 (study or studies or design or analysis or analyses)).ti,ab,kf.	175238

36	(epidemiologic$ adj3 (study or studies or design or analysis or analyses)).ti,ab,kf.	119921

37	((longterm or (long adj term)) adj7 (study or studies or design or analysis or analyses or data)).ti,ab,kf.	160631

38	(cohort$ adj3 (study or studies or design or analysis or analyses or data)).ti,ab,kf.	420054

39	(prospective adj7 (study or studies or design or analysis or analyses)).ti,ab,kf.	551875

40	or/25-39	4243864

41	animals/ not (animals/ and humans/)	5148855

42	40 not 41	4139882

43	42 not (case reports or clinical conference or comment or editorial or letter).pt.	3929840

44	limit 43 to yr="2022 -Current"	538850

45	mortality/ or "cause of death"/ or fatal outcome/ or hospital mortality/ or survival rate/ or Death/	408166

46	(mortalit$ or death$ or fatal$ or survival).ti,ab,ot,kf,kw.	2881687

47	exp Critical Care/ or exp Intensive Care Units/	159166

48	(close attention unit$ or intensive care or respiratory care unit or respiratory care units or special care unit or special care units or high dependency unit or high dependency units).ti,ab,ot,kf,kw.	202827

49	(ICU or ICUs or CCU or CCus or GICU or GICUs or HDU or HDUs or ITU or ITUs).ti,ab,ot,kf,kw.	95415

50	(Hospitaliz$ or Hospitalis$ or (hospital adj2 Admission$) or Critical Care).ti,ab,ot,kf,kw.	436800

51	Hospitalization/ or "Length of Stay"/	228750

52	("length of stay" or "duration of stay" or "extended stay" or "prolonged stay" or "hospital stay").ti,ab,ot,hw.	220112

53	("in-patient stay" or "inpatient stay" or "in-patient stays" or "inpatient stays").ti,ab,ot,hw.	3851

54	exp risk/ or odds ratio/ or proportional hazards models/ or life tables/	1482224

55	(risk$ or danger$ or association$ or peril or jeopard$ or threat$ or chance or chances or hazard$ or gamble$ or probabilit$ or "at stake" or endanger$ or associat$ or likelihood$ or possibilit$ or correlation$ or odds or "vaccine effectiveness" or "immune evasion" or "immunity evasion").ti,ab,ot,kf,kw.	9321739

56	(morbidit$ or comorbid$ or co-morbid$).ti,ab,ot,kf,kw.	716968

57	(incidence or prevalence or predict$ or prognosis).ti,ab,ot,kf,kw.	3993128

58	(HR or RR or aiRR or aOR or "adjusted OR").ti,ab,ot,kf,kw.	471257

59	"severity of illness index"/ or sickness impact profile/	277780

60	patient acuity/ or early warning score/	3345

61	Morbidity/	34586

62	Critical Illness/	39796

63	Emergencies/	43459

64	((severe or severity) adj3 (disease$ or infection$ or illness$ or complication$ or condition$ or illhealth$ or ill-health$ or sickness or virus or infirmity or affliction$)).ti,ab,ot,kf,kw.	285081

65	or/45-64	13120377

66	exp mRNA Vaccines/	5326

67	((pfizer$ or biontech) adj5 vaccin$).ti,ab,ot,kf,kw,hw,nm,rn.	3046

68	(tozinameran or Comirnaty or "Pfizer-BioNTech" or "pf07302048" or "pf-07302048" or " BNT-162b2" or "BNT162b2" or Pidacmeran or "BNT162C2" or "BNT-162C2" or Abdavomeran or "BNT-162B1" or "BNT162B1" or "BNT162A1" or "BNT-162A1").ti,ab,ot,kf,kw,hw,nm,rn.	6421

69	"Pfizer-BioNTech".af.	2384

70	"2417899-77-3".af.	0

71	(Elasomeran or moderna or Spikevax or davesomeran or imelasomeran or andusomeran or "mRNA 1273" or "mRNA1273" or "mRNA-1273.211" or "mRNA 1273.211" or Spikevax or "M-1273" or "M1273" or "CX-024414" or "CX024414" or "TAK-919" or "TAK919").ti,ab,ot,kf,kw,hw,nm,rn.	4118

72	("2430046-03-8" or "2457298-05-2").af.	0

73	(("messenger RNA" or mrna or "RNA based") adj4 (vaccin$ or jab or jabs or shot or shots or immunis$ or immuniz$)).ti,ab,ot,kf,kw.	10148

74	or/66-73	15059

**75	24 and 44 and 74 and 65	825**

*The Medline ALL strategy was updated on 1.2.24 (51 records) and 1.3.24 (63 records).*

COVID facet based on terms from:

World Health Organization (26 May 2021) WHO COVID-19 Database Search Strategy. Systematic search of the COVID-19 literature performed Monday through Friday for the WHO Database. Search strategy as of 26 May 2021. Searches performed by Tomas Allen, Kavita Kothari, and Martha Knuth. Available from: <https://www.who.int/docs/default-source/coronaviruse/who-covid-19-database/who-covid-19_sources_searchstrategy_20210526.pdf?sfvrsn=65209cc2_5>

Canadian Agency for Drugs and Technologies in Health (2.9.21) CADTH COVID-19 Search Strings: COVID-19 — EMBASE (Internet). Available from: <https://covid.cadth.ca/literature-searching-tools/cadth-covid-19-search-strings/>

NICE (18 December 2020) [accessed 17.8.21] COVID-19 rapid guideline: managing the long-term effects of COVID-19 [NG188]. Search history record [PDF]. NICE: London. Available from: <https://www.nice.org.uk/guidance/ng188/evidence/search-strategies-pdf-8957634445>

*Observational study design filter adapted from:*

Observational study design filter adapted from:

Scottish Intercollegiate Guidelines Network (SIGN). Search filters: observational studies. Medline. Edinburgh: SIGN, Last Modified 24/04/17 Available from: <https://www.sign.ac.uk/what-we-do/methodology/search-filters/>

Observational Studies - Medline. In: CADTH Search Filters Database. Ottawa: CADTH; 2023 [accessed 7.11.23]: <https://searchfilters.cadth.ca/list?q=topic%3A%22Observational%20studies%22&p=1&ps=&sort=title_sort%20asc>

**PubMed (NLM): 2022-2024/01/03**

Searched 3.1.24

[**https://pubmed.ncbi.nlm.nih.gov/**](https://pubmed.ncbi.nlm.nih.gov/)

**32	#13 AND #24 AND #31 AND #25	281**

31	#26 OR #27 OR #28 OR #29 OR #30	19,625

30	(("messenger RNA"[Title/Abstract] OR mrna[Title/Abstract] OR "RNA based"[Title/Abstract]) AND (vaccine[Title/Abstract] OR vaccines[Title/Abstract] OR vaccination[Title/Abstract] OR jab[Title/Abstract] OR jabs[Title/Abstract] OR shot[Title/Abstract] OR shots[Title/Abstract] OR immunise[Title/Abstract] OR immunised[Title/Abstract] OR immunisation[Title/Abstract] OR immunize[Title/Abstract] OR immunized[Title/Abstract] OR immunization[Title/Abstract]))	15,979

29	Elasomeran[Title/Abstract] OR moderna[Title/Abstract] OR Spikevax[Title/Abstract] OR davesomeran[Title/Abstract] OR imelasomeran[Title/Abstract] OR andusomeran[Title/Abstract] OR "mRNA 1273"[Title/Abstract] OR "mRNA1273"[Title/Abstract] OR "mRNA-1273.211"[Title/Abstract] OR "mRNA 1273.211"[Title/Abstract] OR Spikevax[Title/Abstract] OR "M-1273"[Title/Abstract] OR "M1273"[Title/Abstract] OR "CX-024414"[Title/Abstract] OR "CX024414"[Title/Abstract] OR "TAK-919"[Title/Abstract] OR "TAK919"[Title/Abstract] OR "2430046-03-8"[Title/Abstract] OR "2457298-05-2"[Title/Abstract]	3,072

28	tozinameran[Title/Abstract] OR Comirnaty[Title/Abstract] OR "Pfizer-BioNTech"[Title/Abstract] OR "pf07302048"[Title/Abstract] OR "pf-07302048"[Title/Abstract] OR " BNT-162b2"[Title/Abstract] OR "BNT162b2"[Title/Abstract] OR Pidacmeran[Title/Abstract] OR "BNT162C2"[Title/Abstract] OR "BNT-162C2"[Title/Abstract] OR Abdavomeran[Title/Abstract] OR "BNT-162B1"[Title/Abstract] OR "BNT162B1"[Title/Abstract] OR "BNT162A1"[Title/Abstract] OR "BNT-162A1"[Title/Abstract] OR "Pfizer-BioNTech"[Title/Abstract] OR "2417899-77-3"[Title/Abstract]	6,392

27	((pfizer[Title/Abstract] OR biontech[Title/Abstract]) AND (vaccine[Title/Abstract] OR vaccines[Title/Abstract]))	3,668

26	"mRNA Vaccines"[Mesh]	5,307

25	pubstatusaheadofprint OR publisher[sb] OR pubmednotmedline[sb]	5,627,356

24	#22 AND #23	493,476

23	("2022/01/01"[Date - Publication] : "3000"[Date - Publication])	3,253,097

22	#20 NOT #21	3,739,291

21	LETTER[Publication Type] OR EDITORIAL[Publication Type] OR COMMENT[Publication Type]	2,213,615

20	#16 NOT #19	3,825,199

19	#18 NOT (#18 AND #17)	3,757,858

18	rat[tiab] or rats[tiab] or mouse[tiab] or mice[tiab] or murine[tiab] or rodent[tiab] or rodents[tiab] or hamster[tiab] or hamsters[tiab] or pig[tiab] or pigs[tiab] or porcine[tiab] or rabbit[tiab] or rabbits[tiab] or animal[tiab] or animals[tiab] or dogs[tiab] or dog[tiab] or cats[tiab] or cow[tiab] or bovine[tiab] or sheep[tiab] or ovine[tiab] or monkey[tiab] or monkeys[tiab]	4,715,684

17	Human*[tiab]	3,295,660

16	#14 OR #15	3,914,514

15	((("Epidemiologic Studies"[Mesh:NoExp]) OR "Case-Control Studies"[Mesh]) OR "Cohort Studies"[Mesh]) OR "Cross-Sectional Studies"[Mesh:NoExp]	3,195,884

14	"Case control"[Title/Abstract] OR "cohort study"[Title/Abstract] OR "cohort studies"[Title/Abstract] OR "cohort analysis"[Title/Abstract] OR "cohort analyses"[Title/Abstract] OR "follow up study"[Title/Abstract] OR "follow up studies"[Title/Abstract] OR "observational study"[Title/Abstract] OR "observational studies"[Title/Abstract] OR Longitudinal[Title/Abstract] OR Retrospective[Title/Abstract] OR "Cross sectional"[Title/Abstract]	2,068,573

13	#9 OR #11 OR #12	47,832

12	omikron[Title/Abstract] OR Omicron[Title/Abstract] OR "B.1.1.529"[Title/Abstract] OR "B11529"[Title/Abstract] OR xbb[Title/Abstract]	9,468

11	#7 AND #10	31,553

10	strain[Title/Abstract] OR strains[Title/Abstract] OR variant[Title/Abstract] OR variants[Title/Abstract] OR mutation[Title/Abstract] OR mutations[Title/Abstract]	1,893,774

9	#7 AND #8	17,832

8	"2022"[Title/Abstract] OR "2023"[Title/Abstract]	158,464

7	#1 OR #4 OR #5 OR #6	412,676

6	"COVID-19 2019"[Title/Abstract] OR ncov[Title/Abstract] OR "sars cov"[Title/Abstract] OR sarscov[Title/Abstract] OR "sars coronavirus"[Title/Abstract] OR coronovirus[Title/Abstract] OR "corono virus"[Title/Abstract] OR "19-nCoV"[Title/Abstract] OR "19nCoV"[Title/Abstract] OR "SARS2"[Title/Abstract] OR "SARS-2"[Title/Abstract] OR SARScoronavirus[Title/Abstract] OR "SARS-coronavirus"[Title/Abstract] OR SARScoronovirus[Title/Abstract] OR "SARS-coronovirus"[Title/Abstract] OR "HCoV-19"[Title/Abstract] OR "HCoV19"[Title/Abstract] OR "HCoV-2019"[Title/Abstract] OR "HCoV2019"[Title/Abstract] OR "2019 novel"[Title/Abstract] OR "Severe Acute Respiratory Syndrome Coronavirus 2"[Title/Abstract] OR "Severe Acute Respiratory Syndrome Corona Virus 2"[Title/Abstract]	141,123

5	"Coronavirus"[Title/Abstract] OR "covid 19"[Title/Abstract] OR "2019-ncov"[Title/Abstract] OR "2019-ncov"[Title/Abstract] OR "2019ncov"[Title/Abstract] OR "corona-virus"[Title/Abstract] OR "cov19"[Title/Abstract] OR "cov-19"[Title/Abstract] OR "19nCoV"[Title/Abstract] OR "COVID19"[Title/Abstract] OR "COVID2019"[Title/Abstract]	382,642

4	#2 AND #3	83,020

3	(("Epidemics"[Mesh:NoExp]) OR "Pandemics"[Mesh:NoExp]) OR "Disease Outbreaks"[Mesh:NoExp]	226,548

2	((("Coronaviridae"[Mesh:NoExp]) OR "Coronavirus"[Mesh]) OR "Betacoronavirus"[Mesh:NoExp]) OR "Coronavirus Infections"[Mesh:NoExp]	185,366

1	("COVID-19"[Mesh]) OR "Severe acute respiratory syndrome-related coronavirus"[Mesh]	258,558

*The PubMed strategy was updated on 1.2.24 (20 records) and 1.3.24 (17 records).*

COVID facet based on terms from:

World Health Organization (26 May 2021) WHO COVID-19 Database Search Strategy. Systematic search of the COVID-19 literature performed Monday through Friday for the WHO Database. Search strategy as of 26 May 2021. Searches performed by Tomas Allen, Kavita Kothari, and Martha Knuth. Available from: <https://www.who.int/docs/default-source/coronaviruse/who-covid-19-database/who-covid-19_sources_searchstrategy_20210526.pdf?sfvrsn=65209cc2_5>

Canadian Agency for Drugs and Technologies in Health (2.9.21) CADTH COVID-19 Search Strings: COVID-19 — EMBASE (Internet). Available from: <https://covid.cadth.ca/literature-searching-tools/cadth-covid-19-search-strings/>

NICE (18 December 2020) [accessed 17.8.21] COVID-19 rapid guideline: managing the long-term effects of COVID-19 [NG188]. Search history record [PDF]. NICE: London. Available from: <https://www.nice.org.uk/guidance/ng188/evidence/search-strategies-pdf-8957634445>

*Observational study design filter adapted from:*

Scottish Intercollegiate Guidelines Network (SIGN). Search filters: observational studies. Medline. Edinburgh: SIGN, Last Modified 24/04/17 Available from: <https://www.sign.ac.uk/what-we-do/methodology/search-filters/>

*PubMed limit:*

Duffy S, de Kock S, Misso K, Noake C, Ross J, Stirk L. Supplementary searches of PubMed to improve currency of MEDLINE and MEDLINE In-Process searches via Ovid. J Med Libr Assoc. 2016 Oct;104(4):309-312. doi: 10.3163/1536-5050.104.4.011. <https://www.ncbi.nlm.nih.gov/pmc/articles/PMC5079494/>

**Europe PMC, including MedRxiv and bioRxiv preprints (Internet): up to 2024/01/04**

Searched 4.1.24

[**https://europepmc.org/**](https://europepmc.org/)

| **Search terms** | **Results** |
| --- | --- |
| (TITLE:"COVID-19" OR TITLE:"coronavirus" OR TITLE:"COVID" OR TITLE:"NCOV" OR TITLE:"SARS-CoV-2") AND (TITLE:"2022" OR TITLE:"2023" OR TITLE:"strain" OR TITLE:"strains" OR TITLE:"variant" OR TITLE:"variants" OR TITLE:"mutation" OR TITLE:"mutations" OR TITLE:"omicron" OR TITLE:"omicron") AND (TITLE:"pfizer" OR TITLE:"biontech" OR TITLE:" tozinameran" OR TITLE:" Comirnaty" OR TITLE:" Pfizer-BioNTech" OR TITLE:" BNT-162b2" OR TITLE:" BNT162b2" OR TITLE:"BNT162C2" OR TITLE:"BNT-162C2" OR TITLE:"Abdavomeran" OR TITLE:"Elasomeran" OR TITLE:"moderna" OR TITLE:"spikevax" OR TITLE:"TAK-919" OR TITLE:"TAK919" OR TITLE:"RNA vaccine" OR TITLE:"RNA vaccines" OR TITLE:"mrna vaccine" OR TITLE:"mrna vaccines") AND (TITLE:"cohort" OR TITLE:"observational" OR TITLE:"longitudinal" OR TITLE:"followup" OR TITLE:"follow up" OR TITLE:"epidemiologic" OR TITLE:"epidemiological" OR TITLE:"longterm" OR TITLE:"long term" OR TITLE:"prospective") | **10** |

*The Europe PMC strategy was updated on 1.2.24 (1 record) and 1.3.24 (0 records).*

**Latin American and Caribbean Health Sciences Literature (LILACS) (Internet): 2022-2024/01/04**

Searched 4.1.24

<https://search.bvsalud.org/portal/?lang=en>

Searched Title/Abstract/Subject

Limited to Observational studies

Limited to 2022-2024/01/04

Limited to LILACS only

((COVID OR NCOV OR coronavirus OR COVID19 OR "SARs-COV-2" OR Omicron OR omicron)) AND ((mortality OR death OR deaths OR fatal OR fatality OR fatalities OR survival OR "close attention unit" OR "close attention units" OR "intensive care" OR "respiratory care unit" OR "respiratory care units" OR "special care unit" OR "special care units" OR "high dependency unit" OR "high dependency units" OR ICU OR ICUs OR CCU OR CCus OR GICU OR GICUs OR HDU OR HDUs OR ITU OR ITUs OR Hospitalized OR Hospitalised OR Hospitalization OR Hospitalisation OR Hospitalizations OR Hospitalisations OR "hospital Admission" OR "hospital Admissions" OR "Critical Care" OR "length of stay" OR "duration of stay" OR "extended stay" OR "prolonged stay" OR "hospital stay" OR "in-patient stay" OR "inpatient stay" OR "in-patient stays" OR "inpatient stays" OR risk OR risks OR risky OR danger OR dangers OR dangerous OR association OR associations OR peril OR jeopardy OR jeopardise OR jeopardize OR threat OR threats OR chance OR chances OR hazard OR hazards OR hazardous OR gamble OR gambles OR gambling OR probability OR probabilities OR "at stake" OR endanger OR endangers OR endangered OR endangering OR associate OR associates OR associating OR association OR likelihood OR likelihoods OR possibility OR possibilities OR correlation OR correlations OR odds OR morbidity OR comorbidity OR "co-morbidity" OR morbidities OR comorbidities OR "co-morbidities" OR incidence OR prevalence OR predict OR predicted OR predictive OR prognosis OR prediction OR predictions OR HR OR RR OR aiRR OR aOR OR "adjusted OR" OR "disease severity" OR "severity score" OR "severity index" OR "severity of illness" OR "critical illness" OR "emergency status" OR "urgency status" OR "severe complication" OR "severe complications") ) AND ((pfizer OR biontech OR tozinameran OR Comirnaty OR "Pfizer-BioNTech" OR "pf07302048" OR "pf-07302048" OR " BNT-162b2" OR "BNT162b2" OR Pidacmeran OR "BNT162C2" OR "BNT-162C2" OR Abdavomeran OR "BNT-162B1" OR "BNT162B1" OR "BNT162A1" OR "BNT-162A1" OR "Pfizer-BioNTech" OR "2417899-77-3" OR Elasomeran OR moderna OR Spikevax OR davesomeran OR imelasomeran OR andusomeran OR "mRNA 1273" OR "mRNA1273" OR "mRNA-1273.211" OR "mRNA 1273.211" OR Spikevax OR "M-1273" OR "M1273" OR "CX-024414" OR "CX024414" OR "TAK-919" OR "TAK919" OR "2430046-03-8" OR "2457298-05-2" OR "RNA vaccine" OR "RNA vaccines" OR "RNA vaccination" OR "RNA shot" OR "RNA shots" OR "mRNA vaccine" OR "mRNA vaccines" OR "mRNA vaccination" OR "mRNA shot" OR "mRNA shots") ) AND ((2022 OR 2023 OR 2024 OR strain OR strains OR variant OR variants OR mutations OR mutation OR omikron OR Omicron OR "B.1.1.529" OR "B11529" OR xbb) )

**N = 3**

*The LILACS strategy was updated on 1.2.24 (0 records) and 1.3.24 (0 records).*

**Cochrane COVID-19 Study Register (www): 2022-2024/01/04**

Searched 4.1.24

<https://covid-19.cochrane.org/>

Limited to Observational cohort studies.

Limited 2022-2024/01/04

(mortality OR death OR deaths OR fatal OR fatality OR fatalities OR survival OR "close attention unit" OR "close attention units" OR "intensive care" OR "respiratory care unit" OR "respiratory care units" OR "special care unit" OR "special care units" OR "high dependency unit" OR "high dependency units" OR ICU OR ICUs OR CCU OR CCus OR GICU OR GICUs OR HDU OR HDUs OR ITU OR ITUs OR Hospitalized OR Hospitalised OR Hospitalization OR Hospitalisation OR Hospitalizations OR Hospitalisations OR "hospital Admission" OR "hospital Admissions" OR "Critical Care" OR "length of stay" OR "duration of stay" OR "extended stay" OR "prolonged stay" OR "hospital stay" OR "in-patient stay" OR "inpatient stay" OR "in-patient stays" OR "inpatient stays" OR risk OR risks OR risky OR danger OR dangers OR dangerous OR association OR associations OR peril OR jeopardy OR jeopardise OR jeopardize OR threat OR threats OR chance OR chances OR hazard OR hazards OR hazardous OR gamble OR gambles OR gambling OR probability OR probabilities OR "at stake" OR endanger OR endangers OR endangered OR endangering OR associate OR associates OR associating OR association OR likelihood OR likelihoods OR possibility OR possibilities OR correlation OR correlations OR odds OR morbidity OR comorbidity OR "co-morbidity" OR morbidities OR comorbidities OR "co-morbidities" OR incidence OR prevalence OR predict OR predicted OR predictive OR prognosis OR prediction OR predictions OR HR OR RR OR aiRR OR aOR OR "adjusted OR" OR "disease severity" OR "severity score" OR "severity index" OR "severity of illness" OR "critical illness" OR "emergency status" OR "urgency status" OR "severe complication" OR "severe complications") AND (pfizer OR biontech OR tozinameran OR Comirnaty OR "Pfizer-BioNTech" OR "pf07302048" OR "pf-07302048" OR " BNT-162b2" OR "BNT162b2" OR Pidacmeran OR "BNT162C2" OR "BNT-162C2" OR Abdavomeran OR "BNT-162B1" OR "BNT162B1" OR "BNT162A1" OR "BNT-162A1" OR "Pfizer-BioNTech" OR "2417899-77-3" OR Elasomeran OR moderna OR Spikevax OR davesomeran OR imelasomeran OR andusomeran OR "mRNA 1273" OR "mRNA1273" OR "mRNA-1273.211" OR "mRNA 1273.211" OR Spikevax OR "M-1273" OR "M1273" OR "CX-024414" OR "CX024414" OR "TAK-919" OR "TAK919" OR "2430046-03-8" OR "2457298-05-2" OR "RNA vaccine" OR "RNA vaccines" OR "RNA vaccination" OR "RNA shot" OR "RNA shots" OR "mRNA vaccine" OR "mRNA vaccines" OR "mRNA vaccination" OR "mRNA shot" OR "mRNA shots") AND (2022 OR 2023 OR 2024 OR strain OR strains OR variant OR variants OR mutations OR mutation OR omikron OR Omicron OR "B.1.1.529" OR "B11529" OR xbb) AND (cohort OR cohorts OR observational OR "follow up" OR followup OR epidemiologic OR epidemiological OR longterm OR "long term" OR longitudinal OR prospective)

**N = (1551 studies) 1792 references**

*The Cochrane COVID-19 Study Register strategy was updated on 1.2.24 (1 record) and 1.3.24 (0 records).*

**WHO COVID-19 (Internet): 2022-2024/01/03**

Searched 3.1.24

[**https://search.bvsalud.org/global-literature-on-novel-coronavirus-2019-ncov/?lang=en**](https://search.bvsalud.org/global-literature-on-novel-coronavirus-2019-ncov/?lang=en)

Limited to Observational and cohort studies.

Limited 2022-2023.

(ti:(mortality OR death OR deaths OR fatal OR fatality OR fatalities OR survival OR "close attention unit" OR "close attention units" OR "intensive care" OR "respiratory care unit" OR "respiratory care units" OR "special care unit" OR "special care units" OR "high dependency unit" OR "high dependency units" OR ICU OR ICUs OR CCU OR CCus OR GICU OR GICUs OR HDU OR HDUs OR ITU OR ITUs OR Hospitalized OR Hospitalised OR Hospitalization OR Hospitalisation OR Hospitalizations OR Hospitalisations OR "hospital Admission" OR "hospital Admissions" OR "Critical Care" OR "length of stay" OR "duration of stay" OR "extended stay" OR "prolonged stay" OR "hospital stay" OR "in-patient stay" OR "inpatient stay" OR "in-patient stays" OR "inpatient stays" OR risk OR risks OR risky OR danger OR dangers OR dangerous OR association OR associations OR peril OR jeopardy OR jeopardise OR jeopardize OR threat OR threats OR chance OR chances OR hazard OR hazards OR hazardous OR gamble OR gambles OR gambling OR probability OR probabilities OR "at stake" OR endanger OR endangers OR endangered OR endangering OR associate OR associates OR associating OR association OR likelihood OR likelihoods OR possibility OR possibilities OR correlation OR correlations OR odds OR morbidity OR comorbidity OR "co-morbidity" OR morbidities OR comorbidities OR "co-morbidities" OR incidence OR prevalence OR predict OR predicted OR predictive OR prognosis OR prediction OR predictions OR HR OR RR OR aiRR OR aOR OR "adjusted OR" OR "disease severity" OR "severity score" OR "severity index" OR "severity of illness" OR "critical illness" OR "emergency status" OR "urgency status" OR "severe complication" OR "severe complications")) AND (pfizer$ OR biontech OR tozinameran OR Comirnaty OR "Pfizer-BioNTech" OR "pf07302048" OR "pf-07302048" OR " BNT-162b2" OR "BNT162b2" OR Pidacmeran OR "BNT162C2" OR "BNT-162C2" OR Abdavomeran OR "BNT-162B1" OR "BNT162B1" OR "BNT162A1" OR "BNT-162A1" OR "Pfizer-BioNTech" OR "2417899-77-3" OR Elasomeran OR moderna OR Spikevax OR davesomeran OR imelasomeran OR andusomeran OR "mRNA 1273" OR "mRNA1273" OR "mRNA-1273.211" OR "mRNA 1273.211" OR Spikevax OR "M-1273" OR "M1273" OR "CX-024414" OR "CX024414" OR "TAK-919" OR "TAK919" OR "2430046-03-8" OR "2457298-05-2" OR "RNA vaccine" OR "RNA vaccines" OR "RNA vaccination" OR "RNA shot" OR "RNA shots" OR "mRNA vaccine" OR "mRNA vaccines" OR "mRNA vaccination" OR "mRNA shot" OR "mRNA shots")

**N = 477**

***** WHO COVID-19 database ceased in June 2023, therefore update searches were not necessary.

#### 1.2. Data Extraction

Data were extracted into a predefined table. One reviewer extracted data on study and patient characteristics, and outcomes of interest, and a second reviewer verified the accuracy of extraction. Only data relating to the Omicron variant were extracted for studies which included other COVID-19 variants.

#### 1.3. Quality Assessment

Risk of bias was assessed using the Newcastle–Ottawa scale for cohort and case-control studies^1^ and the Joanna Briggs Institute checklist for cross-sectional studies.^2^ A single reviewer conducted the assessments and a second reviewer verified the results. Discrepancies were resolved by discussion.

#### 1.4. Statistical Analysis

Odds ratios, hazard ratios, and rate ratios, are considered equal estimates and were therefore combined. Weights were calculated using the inverse variance method (weight=1/variance). Random-effects DerSimonian and Laird models^3^ were fitted to calculate pooled risk ratios (RR) and 95% confidence intervals (CI) for all outcomes. Population analyses that included fewer than 5 subgroups were not conducted due to the challenges associated with combining few studies.

Heterogeneity was measured using Cochran’s Q statistic, with statistical significance set as *P < .*05, and quantified by the I^2^ test.^4^The I^2^ statistic as defined by the Cochrane Handbook for Systematic Reviews of Interventions was used for thresholds for interpretation: 0% to 40% might not be important; 40% to 60% may represent moderate heterogeneity; 60% to 90% may represent substantial heterogeneity; 90% to 100% represents considerable heterogeneity.

Publication bias was assessed using funnel plots and the Egger’s test.^5^

##### 1.4.1.  Sensitivity Analyses

The robustness of the results was assessed using the following sensitivity analyses:

- ‘Leave-1-out’ sensitivity analysis^6^ assessed the effect of removing individual studies on pooled estimates
- ‘Least adjusted’ sensitivity analysis included the least adjusted outcomes reported
- ‘Only adjusted’ sensitivity analysis excluded subgroup results that were not adjusted
- ‘Excluding studies for population overlap’ sensitivity analysis aimed to exclude overlapping studies
- Studies that exclusively included children were excluded from all main analyses. For analyses for which those studies were eligible, an additional sensitivity analysis called ‘Including children’ was conducted

##### 1.4.2.  Additional Subgroup Analyses

The ‘Overweight+’ subgroup included populations that have body mass index ≥25 kg/m^2^ and was analyzed in the comorbid population with obesity.

The ‘Diabetes type 1’ and ‘Diabetes type 2’ subgroups were analyzed in the comorbid population with diabetes.

### 2. Supplemental eResults

#### 2.1. Risk of ICU Admission: Subgroup Analyses

Additionally, there was loss of significance in the ‘COVID-19-related only’ and ‘Hospitalized’ subgroups in patients with diabetes (**eTable 9**).

#### 2.2. Risk of the Combined Outcome (Death, Hospitalization, or ICU Admission)

For all comorbid conditions, except obesity, individuals with the comorbidity had an increased risk of the combined outcome in comparison with individuals without the comorbidity. The increased risk was significant for all comorbidities (*P < .*05) except asthma and hypertension (**Figure 2**). Individuals with obesity had a lower (but non-significant) risk of the combined outcome in comparison with individuals without obesity.

Statistical heterogeneity was considerable for most comorbidities (6/10 applicable comorbid populations). Publication bias was not assessed in most of the analyses due to the small number of studies. Bias was found to be present in the comorbid group with respiratory disease, and absent in the groups with diabetes, heart disease, and hypertension.

Nearly all sensitivity analyses had the same direction of effect and degree of statistical significance as the main analysis for the combined outcome (**Supplementary Table 10**), indicating the robustness of the main analysis results. ‘Leave-1-out’ sensitivity analyses for the comorbid group with respiratory disease resulted in different degrees of significance compared with the main analysis for 2 studies (Vo 2022^7^ and Agrawal 2022^8^).

In subgroup analyses, asthma, respiratory diseases and hypertension resulted in different (although non-significant) directions of effect compared with the main analyses within hospitalized populations. In the general population and ‘COVID-19 related only’ subgroups of individuals, the increased risk of the combined outcome was significant in individuals with asthma and hypertension, in contrast with the main analysis, where the associations were non-significant (**Supplementary Table 11**). As in the main analysis, among the subgroup of hospitalized individuals, those with chronic obstructive pulmonary disease (COPD) and diabetes had an increased risk of the combined outcome compared with individuals without the comorbidity; however, the associations were non-significant in this subgroup (**Supplementary Table 11**).

#### 2.3. Supplemental Tables and Figures

##### 2.3.1. Supplemental eTable 1. Eligibility Criteria

| **Characteristic** | **Inclusion criteria** | **Exclusion criteria** |
| --- | --- | --- |
| Population | - Cardiovascular/cerebrovascular disease:   - Acute and chronic cardiovascular/cerebrovascular groups   - Heart disease   - Atrial fibrillation   - Congestive heart failure   - Peripheral vascular disease   - Myocardial infarction (current or historic)   - Stroke (any type, current or historic)   - Vertebral stenosis   - Intracranial stenosis   - Vascular malformations   - Hypertension, including pulmonary hypertension - Chronic lung condition:   - Lung/respiratory disease groups   - Chronic obstructive pulmonary disease   - Asthma   - Interstitial lung disease - Diabetes type 2 - Diabetes type 1 - Obesity/BMI >30 kg/m^2^ | - Patients with unspecified comorbidities - All patients with conditions of interest to prevent synergizing effects - High-risk patient groups, i.e. patients who are eligible to take antivirals to prevent severe COVID-19 - Highly vulnerable populations, such as the elderly in nursing homes |
| Comparators | - People without respective comorbidities - General population | - Non-comparative studies |
| Outcomes | - Risk of:   - Hospitalization   - ICU admission   - Death   - Combined: other potential COVID-19 severity outcomes, where severity was measured by any of the outcomes outlined above   Outcomes that were not explicit ‘Hospitalization’ or ‘ICU’, such as mechanical ventilation, were also included in this review as part of ‘ICU’ or ‘Hospitalization’ outcomes, depending on the definitions. | - Studies which did not report at least 1 relevant outcome of interest |
| Outcome measures | - Risk ratio - Rate ratio - Hazard ratio - Odds ratio - Incidence rate ratios | - |
| Study design | - Observational (cohort, case-control, cross-sectional) | - Interventional studies, such as randomized/non-randomized controlled trials |
| Language | - Studies with full text published in English were included. All potentially relevant publications without English language full text will be listed | - Studies without a full text published in English |
| Timeframe | - Majority of cases contracted during Omicron variant, as defined by study authors. When the study period was not described as predominantly Omicron, the cut-off date of December 2021, when WHO declared Omicron as a new variant of concern,^18^ was chosen | - Studies not reporting data on the Omicron variant |
| Publication date | - Studies published after 1 Jan 2022 | - Studies published prior to 1 Jan 2022 |
| Publication type | - Full-text articles | - Conference abstracts - Letters - Case reports - Editorials |
| Countries | - Any |  |

BMI, body mass index; COVID-19, coronavirus disease 2019; ICU, intensive care unit; WHO, World Health Organization.

##### 2.3.3. Supplemental eTable 2. Omicron Period in the Included Studies

| **Year** | **2021** | | **2022** | | | | | | | | | | | | **2023** | | | | | | | | | | | |
| --- | --- | --- | --- | --- | --- | --- | --- | --- | --- | --- | --- | --- | --- | --- | --- | --- | --- | --- | --- | --- | --- | --- | --- | --- | --- | --- |
| **Study/ Month** | **11** | **12** | **1** | **2** | **3** | **4** | **5** | **6** | **7** | **8** | **9** | **10** | **11** | **12** | **1** | **2** | **3** | **4** | **5** | **6** | **7** | **8** | **9** | **10** | **11** | **12** |
| Agrawal 2022^8^ |  |  |  |  |  |  |  |  |  |  |  |  |  |  |  |  |  |  |  |  |  |  |  |  |  |  |
| AlBahrani 2022^9^ |  |  |  |  |  |  |  |  |  |  |  |  |  |  |  |  |  |  |  |  |  |  |  |  |  |  |
| Arbel 2023^10^ |  |  |  |  |  |  |  |  |  |  |  |  |  |  |  |  |  |  |  |  |  |  |  |  |  |  |
| Arbel 2022^11^ |  |  |  |  |  |  |  |  |  |  |  |  |  |  |  |  |  |  |  |  |  |  |  |  |  |  |
| Bao 2022^12^ |  |  |  |  |  |  |  |  |  |  |  |  |  |  |  |  |  |  |  |  |  |  |  |  |  |  |
| Bedston 2024^13^ |  |  |  |  |  |  |  |  |  |  |  |  |  |  |  |  |  |  |  |  |  |  |  |  |  |  |
| Benites-Godínez 2023^14^ |  |  |  |  |  |  |  |  |  |  |  |  |  |  |  |  |  |  |  |  |  |  |  |  |  |  |
| Beraud 2023^15^ |  |  |  |  |  |  |  |  |  |  |  |  |  |  |  |  |  |  |  |  |  |  |  |  |  |  |
| Briciu 2023^16^ |  |  |  |  |  |  |  |  |  |  |  |  |  |  |  |  |  |  |  |  |  |  |  |  |  |  |
| Brosh-Nissimov 2023^17^ |  |  |  |  |  |  |  |  |  |  |  |  |  |  |  |  |  |  |  |  |  |  |  |  |  |  |
| Bulgaresi 2023^18^ |  |  |  |  |  |  |  |  |  |  |  |  |  |  |  |  |  |  |  |  |  |  |  |  |  |  |
| Chan 2023^19^ |  |  |  |  |  |  |  |  |  |  |  |  |  |  |  |  |  |  |  |  |  |  |  |  |  |  |
| Chen 2024^20^ |  |  |  |  |  |  |  |  |  |  |  |  |  |  |  |  |  |  |  |  |  |  |  |  |  |  |
| Choi 2023^21^ |  |  |  |  |  |  |  |  |  |  |  |  |  |  |  |  |  |  |  |  |  |  |  |  |  |  |
| de Prost 2022^22^ |  |  |  |  |  |  |  |  |  |  |  |  |  |  |  |  |  |  |  |  |  |  |  |  |  |  |
| Drummond 2023^23^ |  |  |  |  |  |  |  |  |  |  |  |  |  |  |  |  |  |  |  |  |  |  |  |  |  |  |
| Elamin 2024^24^ |  |  |  |  |  |  |  |  |  |  |  |  |  |  |  |  |  |  |  |  |  |  |  |  |  |  |
| Ellis 2023^25^ |  |  |  |  |  |  |  |  |  |  |  |  |  |  |  |  |  |  |  |  |  |  |  |  |  |  |
| Favia 2023^26^ |  |  |  |  |  |  |  |  |  |  |  |  |  |  |  |  |  |  |  |  |  |  |  |  |  |  |
| Finkas 2023^27^ |  |  |  |  |  |  |  |  |  |  |  |  |  |  |  |  |  |  |  |  |  |  |  |  |  |  |
| Flacco 2023^28^ |  |  |  |  |  |  |  |  |  |  |  |  |  |  |  |  |  |  |  |  |  |  |  |  |  |  |
| Flisiak 2023^29^ |  |  |  |  |  |  |  |  |  |  |  |  |  |  |  |  |  |  |  |  |  |  |  |  |  |  |
| Gazit 2022^30^ |  |  |  |  |  |  |  |  |  |  |  |  |  |  |  |  |  |  |  |  |  |  |  |  |  |  |
| Geng 2023^31^ |  |  |  |  |  |  |  |  |  |  |  |  |  |  |  |  |  |  |  |  |  |  |  |  |  |  |
| Grannec 2023^32^ |  |  |  |  |  |  |  |  |  |  |  |  |  |  |  |  |  |  |  |  |  |  |  |  |  |  |
| Griggs 2024^33^ |  |  |  |  |  |  |  |  |  |  |  |  |  |  |  |  |  |  |  |  |  |  |  |  |  |  |
| Guo 2023^34^ |  |  |  |  |  |  |  |  |  |  |  |  |  |  |  |  |  |  |  |  |  |  |  |  |  |  |
| Helmy 2023^35a^ |  |  |  |  |  |  |  |  |  |  |  |  |  |  |  |  |  |  |  |  |  |  |  |  |  |  |
| Hippisley-Cox 2023^36^ |  |  |  |  |  |  |  |  |  |  |  |  |  |  |  |  |  |  |  |  |  |  |  |  |  |  |
| Jamaati 2023^37^ |  |  |  |  |  |  |  |  |  |  |  |  |  |  |  |  |  |  |  |  |  |  |  |  |  |  |
| Karageorgou 2023^38^ |  |  |  |  |  |  |  |  |  |  |  |  |  |  |  |  |  |  |  |  |  |  |  |  |  |  |
| Kim 2023^39^ |  |  |  |  |  |  |  |  |  |  |  |  |  |  |  |  |  |  |  |  |  |  |  |  |  |  |
| Klein 2023^40^ |  |  |  |  |  |  |  |  |  |  |  |  |  |  |  |  |  |  |  |  |  |  |  |  |  |  |
| Lee 2023^41^ |  |  |  |  |  |  |  |  |  |  |  |  |  |  |  |  |  |  |  |  |  |  |  |  |  |  |
| Lewnard 2022^42^ |  |  |  |  |  |  |  |  |  |  |  |  |  |  |  |  |  |  |  |  |  |  |  |  |  |  |
| Li 2024^43^ |  |  |  |  |  |  |  |  |  |  |  |  |  |  |  |  |  |  |  |  |  |  |  |  |  |  |
| Li 2023^44^ |  |  |  |  |  |  |  |  |  |  |  |  |  |  |  |  |  |  |  |  |  |  |  |  |  |  |
| Liu 2023^45^ |  |  |  |  |  |  |  |  |  |  |  |  |  |  |  |  |  |  |  |  |  |  |  |  |  |  |
| Liu 2023a^46^ |  |  |  |  |  |  |  |  |  |  |  |  |  |  |  |  |  |  |  |  |  |  |  |  |  |  |
| Lu 2022^47^ |  |  |  |  |  |  |  |  |  |  |  |  |  |  |  |  |  |  |  |  |  |  |  |  |  |  |
| Manchanda 2023^48^ |  |  |  |  |  |  |  |  |  |  |  |  |  |  |  |  |  |  |  |  |  |  |  |  |  |  |
| Mayer 2023^49^ |  |  |  |  |  |  |  |  |  |  |  |  |  |  |  |  |  |  |  |  |  |  |  |  |  |  |
| McNeil 2023^50^ |  |  |  |  |  |  |  |  |  |  |  |  |  |  |  |  |  |  |  |  |  |  |  |  |  |  |
| Mendoza-Cano 2023 ^51^ |  |  |  |  |  |  |  |  |  |  |  |  |  |  |  |  |  |  |  |  |  |  |  |  |  |  |
| Mizrahi Reuveni 2023^52^ |  |  |  |  |  |  |  |  |  |  |  |  |  |  |  |  |  |  |  |  |  |  |  |  |  |  |
| Morris 2023^53^ |  |  |  |  |  |  |  |  |  |  |  |  |  |  |  |  |  |  |  |  |  |  |  |  |  |  |
| Mosallami Aghili 2022^54^ |  |  |  |  |  |  |  |  |  |  |  |  |  |  |  |  |  |  |  |  |  |  |  |  |  |  |
| Mumtaz 2022^55^ |  |  |  |  |  |  |  |  |  |  |  |  |  |  |  |  |  |  |  |  |  |  |  |  |  |  |
| Nab 2023^56^ |  |  |  |  |  |  |  |  |  |  |  |  |  |  |  |  |  |  |  |  |  |  |  |  |  |  |
| O'Leary 2023^57^ |  |  |  |  |  |  |  |  |  |  |  |  |  |  |  |  |  |  |  |  |  |  |  |  |  |  |
| Parajuli 2023^58^ |  |  |  |  |  |  |  |  |  |  |  |  |  |  |  |  |  |  |  |  |  |  |  |  |  |  |
| Parra-Bracamonte 2023^59^ |  |  |  |  |  |  |  |  |  |  |  |  |  |  |  |  |  |  |  |  |  |  |  |  |  |  |
| Patton 2023^60^ |  |  |  |  |  |  |  |  |  |  |  |  |  |  |  |  |  |  |  |  |  |  |  |  |  |  |
| Radhakrishnan 2023^61^ |  |  |  |  |  |  |  |  |  |  |  |  |  |  |  |  |  |  |  |  |  |  |  |  |  |  |
| Russell 2022^62^ |  |  |  |  |  |  |  |  |  |  |  |  |  |  |  |  |  |  |  |  |  |  |  |  |  |  |
| Sardinha 2023^63^ |  |  |  |  |  |  |  |  |  |  |  |  |  |  |  |  |  |  |  |  |  |  |  |  |  |  |
| Shakor 2023^64^ |  |  |  |  |  |  |  |  |  |  |  |  |  |  |  |  |  |  |  |  |  |  |  |  |  |  |
| Shi 2023^65^ |  |  |  |  |  |  |  |  |  |  |  |  |  |  |  |  |  |  |  |  |  |  |  |  |  |  |
| Simmons 2023^66^ |  |  |  |  |  |  |  |  |  |  |  |  |  |  |  |  |  |  |  |  |  |  |  |  |  |  |
| Skarbinski 2022^67^ |  |  |  |  |  |  |  |  |  |  |  |  |  |  |  |  |  |  |  |  |  |  |  |  |  |  |
| Sonaglioni 2022^68^ |  |  |  |  |  |  |  |  |  |  |  |  |  |  |  |  |  |  |  |  |  |  |  |  |  |  |
| Stepanova 2022^69^ |  |  |  |  |  |  |  |  |  |  |  |  |  |  |  |  |  |  |  |  |  |  |  |  |  |  |
| Tsujimoto 2023^70^ |  |  |  |  |  |  |  |  |  |  |  |  |  |  |  |  |  |  |  |  |  |  |  |  |  |  |
| Vo 2022^7^ |  |  |  |  |  |  |  |  |  |  |  |  |  |  |  |  |  |  |  |  |  |  |  |  |  |  |
| Wang 2023^71^ |  |  |  |  |  |  |  |  |  |  |  |  |  |  |  |  |  |  |  |  |  |  |  |  |  |  |
| Ward 2024^72^ |  |  |  |  |  |  |  |  |  |  |  |  |  |  |  |  |  |  |  |  |  |  |  |  |  |  |
| Xin 2024^73^ |  |  |  |  |  |  |  |  |  |  |  |  |  |  |  |  |  |  |  |  |  |  |  |  |  |  |
| Yang 2023^74^ |  |  |  |  |  |  |  |  |  |  |  |  |  |  |  |  |  |  |  |  |  |  |  |  |  |  |
| Ying-hao 2022^75^ |  |  |  |  |  |  |  |  |  |  |  |  |  |  |  |  |  |  |  |  |  |  |  |  |  |  |
| Zhang 2022^76^ |  |  |  |  |  |  |  |  |  |  |  |  |  |  |  |  |  |  |  |  |  |  |  |  |  |  |
| Zhao 2023^77^ |  |  |  |  |  |  |  |  |  |  |  |  |  |  |  |  |  |  |  |  |  |  |  |  |  |  |
| Zhu 2023^78^ |  |  |  |  |  |  |  |  |  |  |  |  |  |  |  |  |  |  |  |  |  |  |  |  |  |  |

^a^ Helmy 2023 did not explicitly mention the Omicron variant and study period; however, it was included based on the following sentence: “This prospective observational study was conducted in a university Hospital after the institutional research ethics board approval (N-24-2022)”

Green indicates Omicron study period of the studies included in the analyses; Orange indicates Omicron study period of the studies excluded from the analyses.

##### 2.3.4. Supplemental eTable 3. Risk of Bias Assessment

| **Study** | **Low** | **Medium** | **High** |
| --- | --- | --- | --- |
| **Agrawal 2022^8^** | 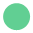 |  |  |
| **AlBahrani 2022^9^** | 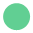 |  |  |
| **Arbel 2023^10^** | 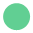 |  |  |
| **Arbel 2022^11^** | 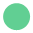 |  |  |
| **Bao 2022^12^**^a^ | 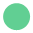 |  |  |
| **Bedston 2024^13^** | 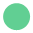 |  |  |
| **Benites-Godínez 2023^14^** | 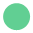 |  |  |
| **Beraud 2023^15^** | 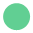 |  |  |
| **Briciu 2023^16^** | 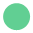 |  |  |
| **Brosh-Nissimov 2023^17^** | 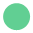 |  |  |
| **Bulgaresi 2023^18^** | 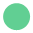 |  |  |
| **Chan 2023^19^**^a^ | 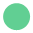 |  |  |
| **Chen 2024^20^** | 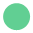 |  |  |
| **Choi 2023^21^** | 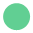 |  |  |
| **de Prost 2022^22^** | 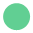 |  |  |
| **Drummond 2023^23^** | 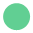 |  |  |
| **Elamin 2024^24^** | 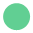 |  |  |
| **Ellis 2023^25^** | 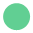 |  |  |
| **Favia 2023^26^** | 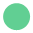 |  |  |
| **Finkas 2023^27^** |  | 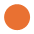 |  |
| **Flacco 2023^28^** | 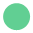 |  |  |
| **Flisiak 2023^29^** | 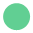 |  |  |
| **Gazit 2022^30^** |  | 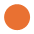 |  |
| **Geng 2023^31^** |  |  | 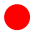 |
| **Grannec 2023^32^** | 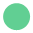 |  |  |
| **Griggs 2024^33^** |  | 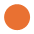 |  |
| **Guo 2023^34^** | 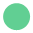 |  |  |
| **Helmy 2023^35^** |  | 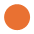 |  |
| **Hippisley-Cox 2023^36^** |  | 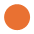 |  |
| **Jamaati 2023^37^** |  | 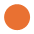 |  |
| **Karageorgou 2023^38^** | 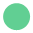 |  |  |
| **Kim 2023^39^** | 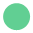 |  |  |
| **Klein 2023^40^** | 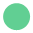 |  |  |
| **Lee 2023^41^** | 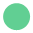 |  |  |
| **Lewnard 2022^42^** |  | 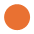 |  |
| **Li 2024^43^** |  | 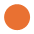 |  |
| **Li 2023^44a^** | 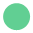 |  |  |
| **Liu 2023^45^** | 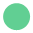 |  |  |
| **Liu 2023a^46^** | 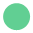 |  |  |
| **Lu 2022^47a^** | 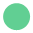 |  |  |
| **Manchanda 2023^48^** | 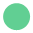 |  |  |
| **Mayer 2023^49^** |  | 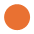 |  |
| **McNeil 2023^50^** |  | 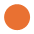 |  |
| **Mendoza-Cano 2023^51^** | 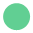 |  |  |
| **Mizrahi Reuveni 2023^52^** | 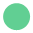 |  |  |
| **Morris 2023^53^** | 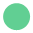 |  |  |
| **Mosallami Aghili 2022^54^** | 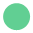 |  |  |
| **Mumtaz 2022^55^** | 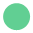 |  |  |
| **Nab 2023^56^** | 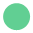 |  |  |
| **O'Leary 2023^57^** | 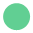 |  |  |
| **Parajuli 2023^58^** | 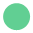 |  |  |
| **Parra-Bracamonte 2023^59^** | 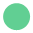 |  |  |
| **Patton 2023^60^** |  | 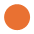 |  |
| **Radhakrishnan 2023^61^** |  | 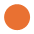 |  |
| **Russell 2022^62^** |  | 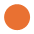 |  |
| **Sardinha 2023^63^** | 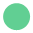 |  |  |
| **Shakor 2023^64a^** |  |  | 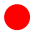 |
| **Shi 2023^65^** | 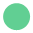 |  |  |
| **Simmons 2023^66^** | 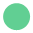 |  |  |
| **Skarbinski 2022^67^** | 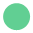 |  |  |
| **Sonaglioni 2022^68^** | 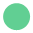 |  |  |
| **Stepanova 2022^69^** |  | 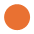 |  |
| **Tsujimoto 2023^70^** | 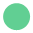 |  |  |
| **Vo 2022^7^** | 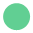 |  |  |
| **Wang 2023^71^** | 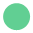 |  |  |
| **Ward 2024^72^** | 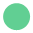 |  |  |
| **Xin 2024^73^** | 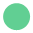 |  |  |
| **Yang 2023^74^** | 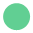 |  |  |
| **Ying-hao 2022^75^** | 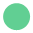 |  |  |
| **Zhang 2022^76^** | 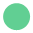 |  |  |
| **Zhao 2023^77^** | 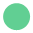 |  |  |
| **Zhu 2023^78a^** | 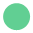 |  |  |

^a^ Risk of bias was assessed using the Joanna Briggs Institute critical appraisal checklist for analytical cross-sectional studies. Green indicates low risk of bias, amber – medium, and red – high.

##### 2.3.5. Supplemental eTable 4. Main and Sensitivity Analyses Results of the ‘Death’ Outcome for All Comorbid Populations of Interest

| **Comorbidity** | **Main analysis (RR [95% CI], *P* value, I^2^)** | **Statistical evidence of publication bias (yes/no)** | **Least adjusted**  **(RR [95% CI], *P* value, I^2^)** | **Only adjusted**  **(RR [95% CI], *P* value, I^2^)** | **Excluding studies for population overlap**  **(RR [95% CI], *P* value, I^2^)** | **Including ‘Overweight+’ subgroups^a^**  **(RR [95% CI], *P* value, I^2^)** | **Leave-1-out (yes/ no for significant difference of removing a subgroup)** |
| --- | --- | --- | --- | --- | --- | --- | --- |
| **Asthma** | 1.10 (0.92, 1.33), *P = .*3, I^2^=93% | No | 1.16 (0.95, 1.4), *P = .*138, I^2^=94% | 1.1 (0.96, 1.33), *P = .*346, I^2^=94% | 1.1 (0.92,1. 33), *P* =0.302, I^2^=93% | NA | No |
| **Cerebrovascular disease** | 1.47 (1.25, 1.73), *P < .*05, I^2^=90% | No | 1.51 (1.23, 1.76), *P < .*001, I^2^=88% | 1.5 (1.23, 1.78), *P < .*001, I^2^=91% | 1.47 (1.25,1.73), *P < .*001, I^2^=89% | NA | No |
| **COPD** | 1.43 (1.11, 1.84), *P < .*05, I^2^=94% | No | 1.47 (1.1, 2.04), *P < .*05, I^2^=97% | 1.48 (1.15, 1.91), *P < .*05, I^2^=95% | 1.43 (1.11, 1.84), *P < .*05, I^2^=94% | NA | No |
| **Diabetes** | 1.37 (1.26, 1.48),  *P < .*05, I^2^=91% | Yes | 1.44 (1.32, 1.57), *P < .*001, I^2^= 93% | 1.38 (1.27, 1.51), *P < .*001, I^2^= 92% | 1.39 (1.28, 1.51), *P < .*001, I^2^= 91% | NA | No |
| **Obesity** | 1.07 (0.95, 1.20), *P > .*05, I^2^=92% | No | 1.08 (0.96, 1.2), *P = .*201, I^2^= 91% | 1.08 (0.96, 1.22), *P = .*225, I^2^ =92% | 1.07 (0.95, 1.2), *P = .*268, I^2^=92% | 1.04 (0.93, 1.16), *P = .*481, I^2^=91% | No |
| **PVD** | NA | NA | NA | NA | NA | NA | NA |
| **Respiratory diseases** | 1.43 (1.14, 1.79), *P < .*05, I^2^=98% | No | 1.53 (1.21, 1.92), *P < .*001, I^2^= 98% | 1.53 (1.2, 1.95), *P*=<.001, I^2^=98% | 1.46 (1.15, 1.84), *P < .*05, I^2^= 98% | NA | No |
| **Atrial fibrillation** | NA | NA | NA | NA | NA | NA | NA |
| **Heart disease** | 1.35 (1.20, 1.52) *P < .*05, I^2^= 98% | No | 1.43 (1.27, 1.61), *P < .*001, I^2^=98% | 1.37 (1.21, 1.54), *P < .*001, I^2^= 98% | 1.35 (1.2, 1.52), *P < .*001, I^2^=98% | NA | No |
| **Heart failure** | 1.78 (1.46, 2.16), *P < .*05, I^2^=87% | No | 1.84 (1.46, 2.33), *P < .*001, I^2^= 91% | 1.83 (1.47, 2.27), *P*=<.001, I^2^=89% | 1.78 (1.46, 2.16), *P*=<.001, I^2^=87% | NA | No |
| **Hypertension** | 1.20 (1.09, 1.32), *P < .*05, I^2^=78% | No | 1.23 (1.12, 1.36), *P < .*001, I^2^=81% | 1.22 (1.01, 1.36), *P < .*001, I^2^=84% | 1.25 (1.13, 1.38), *P < .*001, I^2^=79% | NA | No |
| **Thrombosis** | NA | NA | NA | NA | NA | NA | NA |

BMI, body mass index; CI, confidence interval; COPD, chronic obstructive pulmonary disease; NA, not applicable; PVD, peripheral vascular disease; RR, risk ratio.

^a^ ’Overweight+’ subgroups include populations who have BMI ≥25 kg/m^2^, but the upper limit is unclear.

Green indicates that the significance of the result is the same as of the ‘Main’ analysis. All estimates are rounded to 2 decimal places.

##### 2.3.6. Supplemental eTable 5. Main and Population Subgroup Analyses Results of the ‘Death’ Outcome for all Comorbid Populations of Interest

| **Comorbidity** | **Main analysis (RR [95% CI], *P* value, I^2^)** | **General population**  **(RR [95% CI], *P* value, I^2^)** | **Hospitalized population**  **(RR [95% CI], *P* value, I^2^)** | **COVID-19 related only (RR [95% CI], *P* value, I^2^)** | **Subgroup of a population older than 50 years (RR [95% CI], *P* value, I^2^)** | **Type 1 diabetes (RR [95% CI], *P* value, I^2^)** | **Type 2 diabetes (RR [95% CI], *P* value, I^2^)** |
| --- | --- | --- | --- | --- | --- | --- | --- |
| **Asthma** | 1.10 (0.92, 1.33), *P = .*3, I^2^=93% | 1.13 (0.91, 1.41), *P = .*294, I^2^=93% | 0.84 (0.75, 0.95), *P < .*05, I^2^=1% | 1.18 (0.92, 1.52), *P = .*194, I^2^=93% | 0.86 (0.77, 0.95), *P < .*05, I^2^=0% | NA | NA |
| **Cerebrovascular disease** | 1.47 (1.25, 1.73), *P < .*05, I^2^=90% | 1.49 (1.23, 1.77), *P < .*001, I^2^=92% | 1.4 (0.73, 2.69), *P = .*308, I^2^=44% | 1.49 (1.26, 1.77), *P < .*001, I^2^=92% | 1.4 (1.13, 1.74), *P < .*05, I^2^=63% | NA | NA |
| **COPD** | 1.43 (1.11, 1.84), *P < .*05, I^2^= 94% | 1.59 (1.22, 2.08), *P < .*001, I^2^=95% | 0.71 (0.44, 1.13), *P = .*149, I^2^=0% | 1.74 (1.6, 1.9), *P*=<.001, I^2^=31% | 1.75 (1.48, 2.07), *P*=<.001, I^2^=30% | NA | NA |
| **Diabetes** | 1.37 (1.26, 1.48), *P < .*05, I^2^=91% | 1.47 (1.34, 1.62), *P < .*001, I^2^=91% | 1.18 (1.06, 1.33), *P < .*05, I^2^=77% | 1.51 (1.37, 1.66), *P < .*001, I^2^=81% | 1.34 (1.15, 1.56), *P < .*001, I^2^=71% | 2.44 (1.07, 5.56), *P < .*05, I^2^=91% | 1.36 (1.19, 1.55), *P*=<.001, I^2^=71% |
| **Obesity** | 1.07 (0.95, 1.20), *P > .*05, I^2^= 92%) | 1.16 (0.96, 1.41), *P = .*119, I^2^=94% | 0.88 (0.77, 1.01), *P = .*0654, I^2^=69% | 1.27 (0.97, 1.67), *P = .*0777, I^2^=95% | NA | NA | NA |
| **PVD** | NA | NA | NA | NA | NA | NA | NA |
| **Respiratory diseases** | 1.43 (1.14, 1.79), *P < .*05, I^2^= 98% | 1.88 (1.45, 2.44), *P < .*001, I^2^=95% | 1.16 (0.98, 1.38), *P = .*0869, I^2^=89% | 1.92 (1.46, 2.51), *P < .*001, I^2^=96% | 2.38 (1.07, 5.28), *P = .*0332, I^2^=97% | NA | NA |
| **Atrial fibrillation** | NA | NA | NA | NA | NA | NA | NA |
| **Heart disease** | 1.35 (1.20, 1.52) *P < .*05, I^2^=98% | 1.32 (1.03, 1.7), *P < .*05, I^2^=98% | 1.32 (1.16, 1.49), *P < .*001, I^2^=96% | 1.37 (1.1, 1.7), *P < .*05, I^2^=96% | 1.08 (1.07, 1.16), *P = .*034, I^2^=0% | NA | NA |
| **Heart failure** | 1.78 (1.46, 2.16), *P < .*05, I^2^=87% | 1.99 (1.56, 2.54), *P < .*001, I^2^=92% | 1.41 (0.1, 1.99), *P = .*0504, I^2^=55% | 1.99 (1.56, 2.54), *P < .*001, I^2^=92% | 2.15 (1.3, 3.56), *P = .*003, I^2^=93% | NA | NA |
| **Hypertension** | 1.20 (1.09, 1.32), *P < .*05, I^2^=78% | 1.21 (1.08, 1.37), *P < .*05, I^2^=86% | 1.2 (0.98, 1.47), *P = .*0705, I^2^=67% | 1.2 (0.97, 1.49), *P = .*101, I^2^=84% | 1.22 (0.83, 1.79), *P = .*32, I^2^=63% | NA | NA |
| **Thrombosis** | NA | NA | NA | NA | NA | NA | NA |

CI, confidence interval; COPD, chronic obstructive pulmonary disease; NA, not applicable; PVD, peripheral vascular disease; RR, risk ratio.

Green indicates that the significance of the result is the same as of the ‘Main’ analysis; Orange indicates that the significance of the result is different from the main analysis. All estimates are rounded to 2 decimal places.

##### 2.3.7. Supplemental eTable 6. Main and Sensitivity Analyses Results of the ‘Hospitalization’ Outcome for all Comorbid Populations of Interest

| **Comorbidity** | **Main analysis (RR [95% CI], *P* value, I^2^)** | **Statistical evidence of publication bias (yes/no)** | **Least adjusted**  **(RR [95% CI], *P* value, I^2^)** | **Only adjusted**  **(RR [95% CI], *P* value, I^2^)** | **Excluding studies for population overlap**  **(RR [95% CI], *P* value, I^2^)** | **Including children**  **(RR [95% CI], *P* value, I^2^)** | **Including ‘Overweight+’ subgroups^a^**  **(RR [95% CI], *P* value, I^2^)** | **Leave-1-out (yes/no for significant difference of removing a subgroup)** |
| --- | --- | --- | --- | --- | --- | --- | --- | --- |
| **Asthma** | 1.03 (0.90, 1.17), *P > .*05, I^2^=65% | NA | 1.02 (0.96, 1.07), *P = .*556, I^2^=60% | 1.03 (0.9, 1.17), *P = .*693, I^2^=65% | 1.03 (0.9, 1.17), *P = .*693, I^2^=65% | 1.09 (0.9,1.32), *P = .*373, I^2^=78% | NA | No |
| **Cerebrovascular disease** | 1.37 (1.28, 1.47), *P < .*05, I^2^=21% | NA | 1.37 (1.29, 1.46), *P < .*001, I^2^=12% | 1.37 (1.28, 1.47), *P < .*001, I^2^=21% | 1.37 (1.28, 1.47), *P < .*001, I^2^=21% | NA | NA | No |
| **COPD** | 1.49 (1.31, 1.69), *P < .*05, I^2^=91% | NA | 1.51 (1.34, 1.7), *P < .*001, I^2^=90% | 1.49 (1.31, 1.7), *P < .*001, I^2^=91% | 1.49 (1.31, 1.7), *P < .*001, I^2^=91% | NA | NA | No |
| **Diabetes** | 1.46 (1.27, 1.67), *P < .*05, I^2^=96% | No | 1.48 (1.3, 1.67), *P < .*001, I^2^=95% | 1.46 (1.27, 1.67), *P < .*001, I^2^=96% | 1.46 (1.27, 1.67), *P < .*001, I^2^=96% | NA | NA | No |
| **Obesity** | 1.39 (1.13, 1.70), *P < .*05, I^2^=89% | NA | 1.39 (1.14, 1.7), *P < .*05, I^2^=88% | 1.39 (1.13, 1.7), *P < .*05, I^2^=89% | 1.39 (1.13, 1.7), *P < .*05, I^2^=89% | NA | 1.34 (1.11, 1.61), *P < .*05, I^2^=89% | No |
| **PVD** | NA | NA | NA | NA | NA | NA | NA | NA |
| **Respiratory diseases** | 1.50 (1.41, 1.60), *P < .*05, I^2^=0% | NA | 1.5 (1.41, 1.61), *P < .*001, I^2^=1% | 1.51 (1.41, 1.6), *P < .*001, I^2^=0% | 1.51 (1.4, 1.6), *P < .*001, I^2^=0% | NA | NA | No |
| **Atrial fibrillation** | NA | NA | NA | NA | NA | NA | NA | NA |
| **Heart disease** | 1.27 (1.17, 1.38), *P < .*05, I^2^=74% | NA | 1.3 (1.19, 1.41), *P < .*001, I^2^=75% | 1.27 (1.17, 1.38), *P < .*001, I^2^=74% | 1.27 (1.17, 1.38), *P < .*001, I^2^=74% | NA | NA | No |
| **Heart failure** | 1.61 (1.36, 1.91), *P < .*05, I^2^=91% | NA | 1.62 (1.37, 1.92), *P < .*001, I^2^=91% | 1.61 (1.36, 1.91), *P < .*001, I^2^=91% | 1.61 (1.36, 1.91), *P < .*001, I^2^=91% | NA | NA | No |
| **Hypertension** | 1.48 (1.03, 2.14), *P < .*05, I^2^=90% | NA | 1.48 (1.03, 2.14), *P < .*05, I^2^=91% | 1.49 (1.03, 2.14), *P < .*05, I^2^=91% | 1.49 (1.03, 2.14), *P < .*05, I^2^=91% | NA | NA | Yes |
| **Thrombosis** | NA | NA | NA | NA | NA | NA | NA | NA |

BMI, body mass index; CI, confidence interval; COPD, chronic obstructive pulmonary disease; NA, not applicable; PVD, peripheral vascular disease; RR, risk ratio.

^a^ ’Overweight+’ subgroups include populations who have BMI ≥25 kg/m^2^, but the upper limit is unclear.

Green indicates that the significance of the result is the same as of the ‘Main’ analysis; Orange indicates that the significance of the result is different from the main analysis. All estimates are rounded to 2 decimal places.

##### 2.3.8. Supplemental eTable 7. Main and Population Subgroup Analyses Results of the ‘Hospitalization’ Outcome for all Comorbid Populations of Interest

| **Comorbidity** | **Main analysis (RR [95% CI], *P* value, I^2^)** | **Subgroup of a population older than 50 years (RR [95% CI], *P* value, I^2^)** | **COVID-19 related only (RR [95% CI], *P* value, I^2^)** | **Type 1 diabetes (RR [95% CI], *P* value, I^2^)** | **Type 2 diabetes (RR [95% CI], *P* value, I^2^)** |
| --- | --- | --- | --- | --- | --- |
| **Asthma** | 1.03 (0.90, 1.17), *P > .*05, I^2^=65% | NA | 0.97 (0.8, 1.17), *P > .*05, I^2^=55% | NA | NA |
| **Cerebrovascular disease** | 1.37 (1.28, 1.47), *P < .*05, I^2^=21% | 1.48 (1.3, 1.7), *P < .*001, I^2^=0% | 1.38 (1.28, 1.5), *P < .*001, I^2^=43% | NA | NA |
| **COPD** | 1.49 (1.31, 1.69), *P < .*05, I^2^=91% | 1.9 (1.34, 2.68), *P < .*001, I^2^=80% | 1.6 (1.35, 1.9), *P < .*001, I^2^=91% | NA | NA |
| **Diabetes** | 1.46 (1.27, 1.67), *P < .*05, I^2^=96% | 1.47 (1.32, 1.65), *P < .*001, I^2^=0% | 1.65 (1.33, 2.03), *P < .*001, I^2^=98% | 2.97 (2.37, 3.71), *P < .*001, I^2^=68% | 1.3 (1.11, 1.53), *P < .*05, I^2^=98% |
| **Obesity** | 1.39 (1.13, 1.70), *P < .*05, I^2^=89% | NA | 1.05 (0.99, 1.1), *P > .*05, I^2^=0% | NA | NA |
| **PVD** | NA | NA | NA | NA | NA |
| **Respiratory diseases** | 1.50 (1.41, 1.60), *P < .*05, I^2^=0% | NA | 1.5 (1.25, 1.81), *P < .*001, I^2^=51% | NA | NA |
| **Atrial fibrillation** | NA | NA | NA | NA | NA |
| **Heart disease** | 1.27 (1.17, 1.38), *P < .*05, I^2^=74% | 1.19 (1.05, 1.34), *P < .*05, I^2^=0% | 1.23 (1.14, 1.34), *P < .*001, I^2^=73% | NA | NA |
| **Heart failure** | 1.61 (1.36, 1.91), *P < .*05, I^2^=91% | 2.13 (1.85, 2.44), *P < .*001, I^2^=0% | 1.66 (1.31, 2.1), *P < .*001, I^2^=91% | NA | NA |
| **Hypertension** | 1.48 (1.03, 2.14), *P < .*05, I^2^=90% | 1.34 (1.16,1.55), *P < .*001, I^2^=0% | 1.61 (1.07, 2.42), *P < .*05, I^2^=94% | NA | NA |
| **Thrombosis** | NA | NA | NA | NA | NA |

CI, confidence interval; COPD, chronic obstructive pulmonary disease; NA, not applicable; PVD, peripheral vascular disease; RR, risk ratio.

Green indicates that the significance of the result is the same as of the ‘Main’ analysis; Orange indicates that the significance of the result is different from the main analysis. All estimates are rounded to 2 decimal places.

##### 2.3.9. Supplemental eTable 8. Main and Sensitivity Analyses Results of the ‘ICU’ Outcome for all Comorbid Populations of Interest

| **Comorbidity** | **Main analysis (RR [95% CI], *P* value, I^2^)** | **Statistical evidence of publication bias (yes/no)** | **Least adjusted**  **(RR [95% CI], *P* value, I^2^)** | **Only adjusted**  **(RR [95% CI], *P* value, I^2^)** | **Excluding studies for population overlap**  **(RR [95% CI], *P* value, I^2^)** | **Including children**  **(RR [95% CI], *P* value, I^2^)** | **Including ‘Overweight+’^a^**  **(RR [95% CI], *P* value, I^2^)** | **Leave-1-out (yes/no for significant difference of removing a subgroup)** |
| --- | --- | --- | --- | --- | --- | --- | --- | --- |
| **Asthma** | NA | NA | NA | NA | NA | NA | NA | NA |
| **Cerebrovascular disease** | NA | NA | NA | NA | NA | NA | NA | NA |
| **COPD** | NA | NA | NA | NA | NA | NA | NA | NA |
| **Diabetes** | 1.20 (1.04, 1.38), *P < .*05, I^2^=72% | No | 1.43 (1.22, 1.67), *P < .*001, I^2^=88% | 1.2 (1.04, 1.39), *P < .*05, I^2^=75% | 1.2 (1.04, 1.38), *P < .*05, I^2^=72% | NA | NA | Yes |
| **Obesity** | 1.32 (1.11, 1.57), *P < .*05, I^2^=37% | NA | 1.33 (1.02, 1.75), *P < .*05, I^2^=74% | 1.32 (1.11, 1.57), *P < .*05, I^2^=37% | 1.32 (1.11, 1.57), *P < .*05, I^2^=37% | NA | 1.3 (1.12, 1.51), *P < .*001, I^2^=57% | Yes |
| **PVD** | NA | NA | NA | NA | NA | NA | NA | NA |
| **Respiratory diseases** | 1.15 (0.94, 1.41), *P > .*05, I^2^=84% | NA | 1.12 (0.92, 1.36), *P = .*263, I^2^=88% | 1.15 (0.94, 1.41), *P = .*162, I^2^=84% | 1.15 (0.94, 1.41), *P = .*162, I^2^=84% | NA | NA | No |
| **Atrial fibrillation** | NA | NA | NA | NA | NA | NA | NA | NA |
| **Heart disease** | 1.09 (0.90, 1.33), *P > .*05, I^2^=94% | No | 1.32 (1.02, 1.7), *P < .*05, I^2^=99% | 1.04 (0.85, 1.27), *P = .*714, I^2^=95% | 1.09 (0.9, 1.33), *P = .*37, I^2^=94% | NA | NA | No |
| **Heart failure** | NA | NA | NA | NA | NA | NA | NA | NA |
| **Hypertension** | 1.17 (0.90, 1.51), *P > .*05, I^2^=0% | NA | 1.36 (1.08, 1.72), *P < .*05, I^2^=0% | 1.33 (0.88, 2.01), *P = .*175, I^2^=0% | 1.17 (0.9, 1.51), *P = .*235, I^2^=0% | NA | NA | No |
| **Thrombosis** | NA | NA | NA | NA | NA | NA | NA | NA |

BMI, body mass index; CI, confidence interval; COPD, chronic obstructive pulmonary disease; ICU, intensive care unit; NA, not applicable; PVD, peripheral vascular disease; RR, risk ratio.

^a^ ’Overweight+’ subgroups include populations who have BMI ≥25 kg/m^2^, but the upper limit is unclear.

Green indicates that the significance of the result is the same as of the ‘Main’ analysis; Orange indicates that the significance of the result is different from the main analysis. All estimates are rounded to 2 decimal places.

##### 2.3.10. Supplemental eTable 9. Main and Population Subgroup Analyses Results of the ‘ICU’ Outcome for all Comorbid Populations of Interest

| **Comorbidity** | **Main analysis (RR [95% CI], *P* value, I^2^)** | **‘General’ population (RR [95% CI], *P* value, I^2^)** | **‘Hospitalized’ population (RR [95% CI], *P* value, I^2^)** | **COVID-19 related only (RR [95% CI], *P* value, I^2^)** | **Subgroup of a population older than 50 years (RR [95% CI], *P* value, I^2^)** | **Type 1 diabetes (RR [95% CI], *P* value, I^2^)** | **Type 2 diabetes (RR [95% CI], *P* value, I^2^)** |
| --- | --- | --- | --- | --- | --- | --- | --- |
| **Asthma** | NA | NA | NA | NA | NA | NA | NA |
| **Cerebrovascular disease** | NA | NA | NA | NA | NA | NA | NA |
| **COPD** | NA | NA | NA | NA | NA | NA | NA |
| **Diabetes** | 1.20 (1.04, 1.38), *P < .*05, I^2^= 72% | 1.43 (1.28, 1.59), *P < .*001, I^2^=0% | 1.1 (0.98, 1.21), *P = .*117, I^2^=36% | 1.21 (0.916, 1.59), *P > .*05, I^2^=71% | NA | NA | NA |
| **Obesity** | 1.32 (1.11, 1.57), *P < .*05, I^2^=37% | NA | 1.3 (1.09, 1.54), *P < .*05, I^2^=39% | NA | NA | NA | NA |
| **PVD** | NA | NA | NA | NA | NA | NA | NA |
| **Respiratory diseases** | 1.15 (0.94, 1.41), *P > .*05, I^2^=84% | NA | 1.03 (0.89, 1.2), *P = .*677, I^2^=56% | 1.46 (1.27, 1.68), *P < .*001, I^2^=0% | NA | NA | NA |
| **Atrial fibrillation** | NA | NA | NA | NA | NA | NA | NA |
| **Heart disease** | 1.09 (0.90, 1.33), *P > .*05, I^2^=94% | NA | 1.09 (0.9, 1.33), *P = .*369, I^2^=95% | 3.29 (2.13, 5.07), *P < .*001, I^2^=0% | NA | NA | NA |
| **Heart failure** | NA | NA | NA | NA | NA | NA | NA |
| **Hypertension** | 1.17 (0.90, 1.51), *P > .*05, I^2^=0% | NA | 1.15 (0.89, 1.5), *P = .*278, I^2^=0% | 1.03 (0.66, 1.63), *P = .*891, I^2^=0% | NA | NA | NA |
| **Thrombosis** | NA | NA | NA | NA | NA | NA | NA |

CI, confidence interval; COPD, chronic obstructive pulmonary disease; ICU, intensive care unit; NA, not applicable; PVD, peripheral vascular disease; RR, risk ratio.

Green indicates that the significance of the result is the same as of the ‘Main’ analysis; Orange indicates that the significance of the result is different from the main analysis. All estimates are rounded to 2 decimal places.

##### 2.3.11. Supplemental eTable 10. Main and Sensitivity Analyses Results of the ‘Combined’ Outcome for all Comorbid Populations of Interest

| **Comorbidity** | **Main analysis (RR [95% CI], *P* value, I^2^)** | **Statistical evidence of publication bias (yes/no)** | **Least adjusted**  **(RR [95% CI], *P* value, I^2^)** | **Only adjusted**  **(RR [95% CI], *P* value, I^2^)** | **Excluding studies for population overlap**  **(RR [95% CI], *P* value, I^2^)** | **Including children**  **(RR [95% CI], *P* value, I^2^)** | **Including ‘Overweight+’ subgroups^a^**  **(RR [95% CI], *P* value, I^2^)** | **Leave-1-out (yes/no for significant difference of removing a subgroup)** |
| --- | --- | --- | --- | --- | --- | --- | --- | --- |
| **Asthma** | 1.09 (0.93; 1.28), *P > .*05, I^2^=93% | NA | 1.09 (0.93, 1.28), *P = .*291, I^2^=93% | 1.06 (0.91, 1.24), *P*= 0.472, I^2^=94% | 1.09 (0.93, 1.28), *P = .*291, I^2^=93% | NA | NA | No |
| **Cerebrovascular disease** | 1.58 (1.43, 1.75), *P < .*05, I^2^=84% | NA | 1.6 (1.43, 1.78), *P < .*001, I^2^=85% | 1.58 (1.43, 1.76), *P < .*001, I^2^=86% | 1.58 (1.43, 1.75), *P < .*001, I^2^=84% | NA | NA | No |
| **COPD** | 1.58 (1.18, 2.11), *P < .*05, I^2^=99% | NA | 1.54 (1.15, 2.08), *P < .*05, I^2^=99% | 1.54 (1.13, 2.11), *P < .*05, I^2^=99% | 1.58 (1.18, 2.11), *P < .*05, I^2^=99% | NA | NA | No |
| **Diabetes** | 1.28 (1.15, 1.43), *P < .*05, I^2^=84% | No | 1.29 (1.16, 1.44), *P < .*001, I^2^=84% | 1.25 (1.12, 1.4), *P < .*001, I^2^=87% | 1.28 (1.15, 1.43), *P < .*001, I^2^=84% | NA | NA | No |
| **Obesity** | 0.97 (0.80, 1.17), *P > .*05, I^2^=84% | NA | 0.99 (0.81, 1.2), *P = .*884, I^2^=83% | 0.95 (0.76, 1.19), *P = .*664, I^2^=89% | 0.97 (0.8, 1.17), *P = .*714, I^2^=83% | 0.96 (0.8, 1.15), *P = .*657, I^2^=81% | 0.97 (0.8, 1.17), *P = .*714, I^2^=84% | No |
| **PVD** | NA | NA | NA | NA | NA | NA | NA | NA |
| **Respiratory diseases** | 1.48 (1.23, 1.79), *P < .*05, I^2^=78% | Yes | 1.51 (1.25, 1.82), *P < .*001, I^2^=78% | 1.6 (1.33, 1.93), *P < .*001, I^2^=81% | 1.5 (1.24, 1.81), *P < .*001, I^2^=79% | 1.49 (1.24, 1.79), *P < .*001, I^2^=75% | NA | Yes |
| **Atrial fibrillation** | 1.59 (1.34, 1.90), *P < .*05, I^2^=91% | NA | 1.6 (1.34, 1.9), *P < .*001, I^2^=91% | 1.59 (1.33, 1.89), *P < .*001, I^2^=93% | 1.6 (1.34, 1.9), *P < .*001, I^2^=91% | NA | NA | Yes |
| **Heart disease** | 1.43 (1.28, 1.60), *P < .*05, I^2^=92% | No | 1.4 (1.25, 1.58), *P < .*001, I^2^=93% | 1.42 (1.26, 1.6), *P < .*001, I^2^=94% | 1.37 (1.23, 1.53), *P < .*001, I^2^=92% | NA | NA | No |
| **Heart failure** | 1.63 (1.29, 2.06), *P < .*05, I^2^=98% | NA | 1.63 (1.29, 2.06), *P < .*001, I^2^=98% | 1.63 (1.29, 2.06), *P < .*001, I^2^=98% | 1.63 (1.29, 2.06), *P < .*001, I^2^=98% | NA | NA | No |
| **Hypertension** | 1.27 (0.96, 1.67), *P > .*05, I^2^=98% | No | 1.29 (0.98, 1.7), *P > .*05, I^2^=98% | 1.17 (0.83, 1.64), *P > .*05, I^2^=99% | 1.27 (0.95, 1.69), *P > .*05, I^2^=98% | NA | NA | No |
| **Thrombosis** | NA | NA | NA | NA | NA | NA | NA | NA |

BMI, body mass index; CI, confidence interval; COPD, chronic obstructive pulmonary disease; NA, not applicable; PVD, peripheral vascular disease; RR, risk ratio.

^a^ ’Overweight+’ subgroups include populations who have BMI ≥25 kg/m^2^, but the upper limit is unclear.

Green indicates that the significance of the result is the same as of the ‘Main’ analysis; Orange indicates that the significance of the result is different from the main analysis. All estimates are rounded to 2 decimal places.

##### 2.3.12. Supplemental eTable 11. Main and Population Subgroup Analyses Results of the ‘Combined’ Outcome for all Comorbid Populations of Interest

| **Comorbidity** | **Main analysis (RR [95% CI], *P* value, I^2^)** | **‘General’ population**  **(RR [95% CI, *P* value, I^2^)** | **‘Hospitalized’ population (RR [95% CI], *P* value, I^2^)** | **COVID-19 related only (RR [95% CI], *P* value, I^2^)** | **Subgroup of a population older than 50 years (RR [95% CI], *P* value, I^2^)** | **Type 1 diabetes (RR [95% CI], *P* value, I^2^)** | **Type 2 diabetes (RR [95% CI], *P* value, I^2^)** |
| --- | --- | --- | --- | --- | --- | --- | --- |
| **Asthma** | 1.09 (0.93, 1.28), *P > .*05, I^2^=93% | 1.26 (1.11, 1.43), *P < .*001, I^2^=87% | 0.97 (0.84, 1.13), *P = .*686, I^2^=66% | 1.28 (1.1, 1.49), *P < .*05, I^2^=88% | NA | NA | NA |
| **Cerebrovascular disease** | 1.58 (1.43, 1.75), *P < .*05, I^2^=84% | 1.77 (1.71, 1.83), *P < .*001, I^2^=0% | 1.45 (1.33, 1.59), *P < .*001, I^2^=40% | 1.77 (1.71, 1.83), *P < .*001, I^2^=0% | NA | NA | NA |
| **COPD** | 1.58 (1.18, 2.11), *P < .*05, I^2^=99% | 2.21 (1.95, 2.49), *P < .*001, I^2^=76% | 1.04 (0.96, 1.13), *P*= 0.302, I^2^=60% | 2.2 (1.94, 2.5), *P < .*001, I^2^=85% | NA | NA | NA |
| **Diabetes** | 1.28 (1.15, 1.43), *P < .*05, I^2^=84% | 1.43 (1.26, 1.62), *P < .*001, I^2^=76% | 1.1 (0.98, 1.19), *P*= 0.129, I^2^=58% | NA | NA | 2.09 (1.57, 2.76), *P < .*001, I^2^=0% | 1.47 (1.37, 1.58), *P < .*001, I^2^=0% |
| **Obesity** | 0.97 (0.80, 1.17), *P > .*05, I^2^=84% | 0.96 (0.79, 1.16), *P = .*656, I^2^=86% | NA | 0.92 (0.74, 1.14), *P = .*44, I^2^=90% | NA | NA | NA |
| **PVD** | NA | NA | NA | NA | NA | NA | NA |
| **Respiratory diseases** | 1.48 (1.23, 1.79), *P < .*05, I^2^=78% | 1.88 (1.74, 2.02), *P < .*001, I^2^=35% | 0.79 (0.59, 1.07), *P = .*132, I^2^=0% | 1.85 (1.7, 2.02), *P < .*001, I^2^=38% | NA | NA | NA |
| **Atrial fibrillation** | 1.59 (1.34, 1.90), *P < .*05, I^2^=91% | 1.59 (1.33, 1.89), *P < .*001, I^2^=93% | NA | 1.59 (1.33, 1.9), *P < .*001, I^2^=95% | NA | NA | NA |
| **Heart disease** | 1.43 (1.28, 1.60), *P < .*05, I^2^=92% | 1.48 (1.28, 1.71), *P < .*001, I^2^=94% | 1.32 (1.17, 1.48), *P < .*001, I^2^=71% | 1.56 (1.34, 1.81), *P < .*001, I^2^=93% | 1.75 (1.47, 2.09), *P*=<.001, I^2^=0% | NA | NA |
| **Heart failure** | 1.63 (1.29, 2.06), *P < .*05, I^2^=98% | 2.13 (1.84, 2.45), *P < .*001, I^2^=90% | 1.34 (1.23, 1.46), *P < .*001, I^2^=67% | 2.13 (1.84, 2.45), *P < .*001, I^2^=90% | NA | NA | NA |
| **Hypertension** | 1.29 (0.98, 1.7), *P > .*05, I^2^=98% | 1.79 (1.11, 2.88), *P < .*05, I^2^=98% | 0.92 (0.82, 1.03), *P = .*15, I^2^=65% | 1.63 (1.07, 2.47), *P < .*05, I^2^=98% | 1.01 (0.49, 2.1), *P > .*05, I^2^=82% | NA | NA |
| **Thrombosis** | NA | NA | NA | NA | NA | NA | NA |

CI, confidence interval; COPD, chronic obstructive pulmonary disease; NA, not applicable; PVD, peripheral vascular disease; RR, risk ratio.

Green indicates that the significance of the result is the same as of the ‘Main’ analysis; Orange indicates that the significance of the result is different from the main analysis. All estimates are rounded to 2 decimal places.

#### 2.4. Meta-analyses for Each Comorbidity: Forest Plots of Pooled RR for Each Comorbidity and Outcome

##### 2.4.1. Asthma

###### 2.4.1.1. Supplemental eFigure 1. Asthma, Death Outcome (No. of Subgroups: 12)

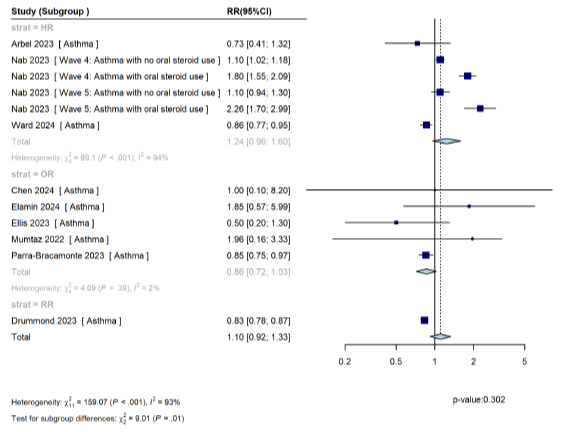

CI, confidence interval; HR, hazard ratio; OR, odds ratio; RR, risk ratio.

###### 2.4.1.2. Supplemental eFigure 2. Asthma, Hospitalization Outcome (No. of Subgroups: 5)

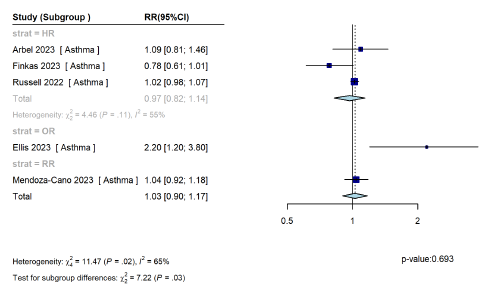

CI, confidence interval; HR, hazard ratio; OR, odds ratio; RR, risk ratio.

###### 2.4.1.3. Asthma, ICU Admission Outcome (No. of Studies: 4; No Subgroups Included in ‘Main’ Analysis: 3)

Three studies reported the following results for the ‘ICU’ outcome in the ‘Asthma’ populations, none of which were statistically significant:

- Elamin 2024^24^ (Asthma): odds ratio (OR) (95% confidence interval [CI]): 0.87 (0.26, 2.90)
- Helmy 2023^35^ (Bronchial asthma): OR (95% CI): 3.64 (0.04, 344.66)
- Russell 2022^62^ (Asthma): hazard ratio (HR) (95% CI): 0.99 (0.87, 1.12)

An additional study (Chan 2023^19^) that only included children (and thus would only be included in a ‘Including children’ sensitivity analysis) reported the following results:

- Asthma-Omicron BA.1 wave: OR (95% CI): 1.10 (0.17, 4.08)
- Asthma-Omicron BA.2 wave: OR (95% CI): 0.75 (0.11, 2.87)

###### 2.4.1.4. Supplemental eFigure 3. Asthma, Combined Outcome (No. of Subgroups: 9)

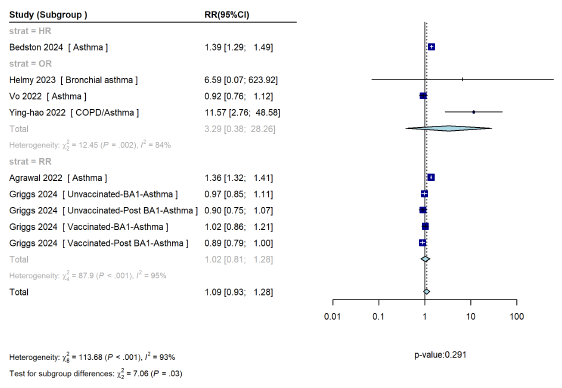

CI, confidence interval; HR, hazard ratio; OR, odds ratio; RR, risk ratio.

##### 2.4.2. Cerebrovascular Disease

###### 2.4.2.1. Supplemental eFigure 4. Cerebrovascular Disease, Death Outcome (No. of Subgroups: 11)

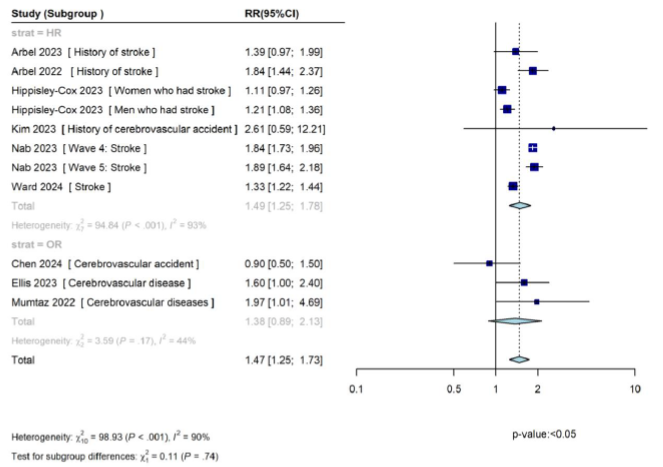

CI, confidence interval; HR, hazard ratio; OR, odds ratio; RR, risk ratio.

###### 2.4.2.2. Supplemental eFigure 5. Cerebrovascular Disease, Hospitalization Outcome (No. of Subgroups: 6)

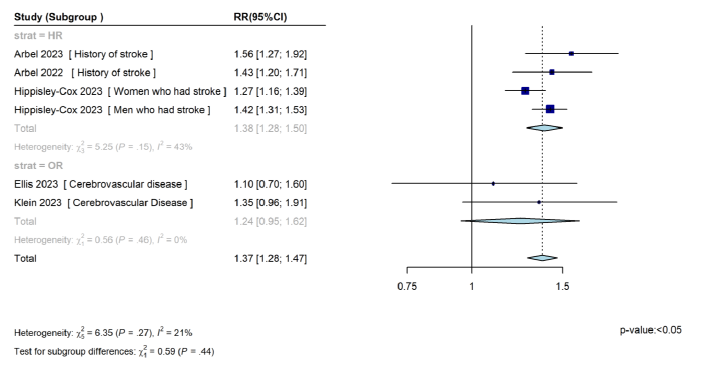

CI, confidence interval; HR, hazard ratio; OR, odds ratio; RR, risk ratio.

###### 2.4.2.3. Cerebrovascular Disease, ICU Admission Outcome (No. of Studies: 4; No. of Subgroups Included in ‘Main’ Analysis: 3)

The studies reported the following results for the ‘ICU’ outcome in the ‘Cerebrovascular disease’ populations:

- Helmy 2023^35^ (Stroke): OR (95% CI): 2.67 (0.03, 256.4)
- Mayer 2023^49^ (History of cerebrovascular disease): OR (95% CI): 1.02 (0.95, 1.1)
- Zhang 2022^76^ (Cerebrovascular disease): OR (95% CI): 2.04 (1.09, 3.77)

While all of the results showed an increase in the risk of ICU admission, only the Zhang 2022 estimate was statistically significant, based on the CI. Bao 2022^12^ provided a conflicting interpretation of the results, and thus, is not discussed in this section.

###### 2.4.2.4. Supplemental eFigure 6. Cerebrovascular Disease, Combined Outcome (No. of Subgroups: 8)

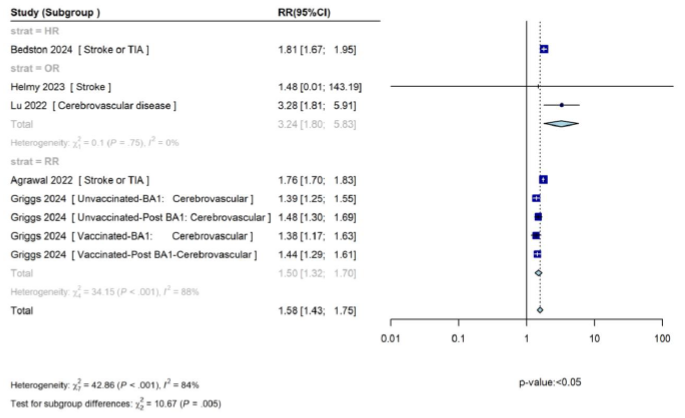

CI, confidence interval; HR, hazard ratio; OR, odds ratio; RR, risk ratio; TIA, transient ischemic attack.

##### 2.4.3. COPD

###### 2.4.3.1. Supplemental eFigure 7. COPD, Death Outcome (No. of Subgroups: 12)

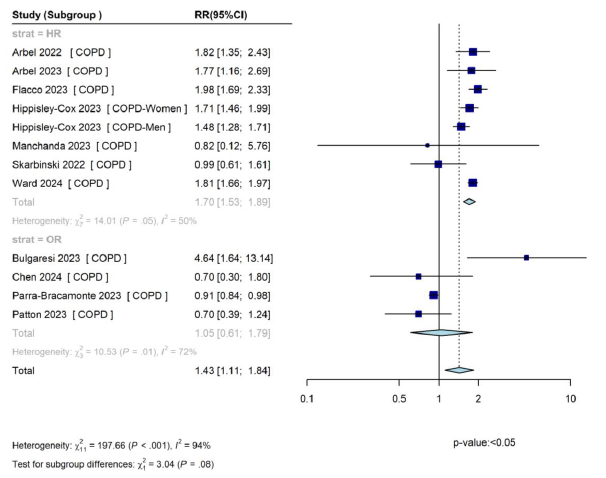

CI, confidence interval; COPD, chronic obstructive pulmonary disease; HR, hazard ratio; OR, odds ratio; RR, risk ratio.

###### 2.4.3.2. Supplemental eFigure 8: COPD, Hospitalization Outcome (No. of Subgroups: 8)

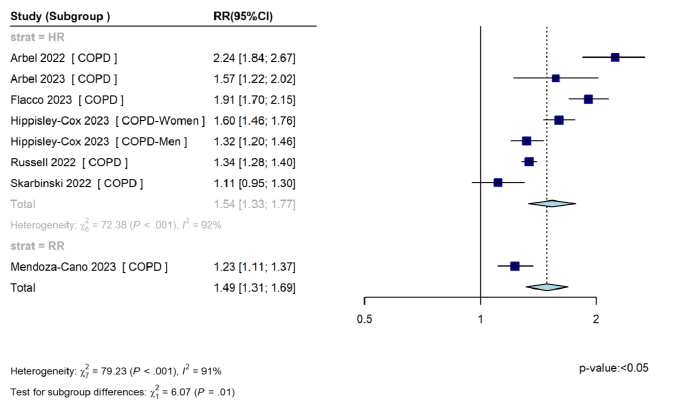

CI, confidence interval; COPD, chronic obstructive pulmonary disease; HR, hazard ratio; RR, risk ratio.

###### 2.4.3.3. COPD, ICU Admission Outcome (No. of Studies: 3)

The studies reported the following results for the ‘ICU’ outcome in the ‘COPD’ populations:

- Russell 2022^62^ (COPD): HR (95% CI): 1.06 (0.91, 1.22)
- Zhang 2022^76^ (COPD): OR (95% CI): 3.04 (1.21, 7.64)
- Zhu 2023^78^ (COPD): OR 1.86 (95% CI): (0.12, 2.69)

While the results of Russell 2022 and Zhu 2023 show an increased risk of the ‘ICU’ outcome in ‘COPD’ populations, the CI crossed the line of no effect, indicating that the results are not statistically significant. The results of Zhang 2022 showed a significant increase in the risk of the ‘ICU’ outcome in the ‘COPD’ populations. This might be due to the population of Zhang 2022 consisting of less vaccinated and generally older patients (the study did not provide the adjustments; thus, it is likely that these confounding factors were not adjusted for).

###### 2.4.3.4. Supplemental eFigure 9. COPD, Combined Outcome (No. of Subgroups: 9)

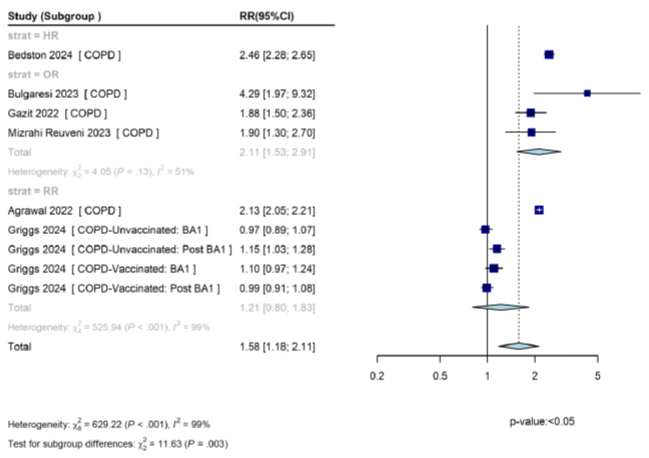

CI, confidence interval; COPD, chronic obstructive pulmonary disease; HR, hazard ratio; OR, odds ratio; RR, risk ratio.

##### 2.4.4. Diabetes

###### 2.4.4.1. Supplemental eFigure 10. Diabetes, Death Outcome (No. of Subgroups: 40)

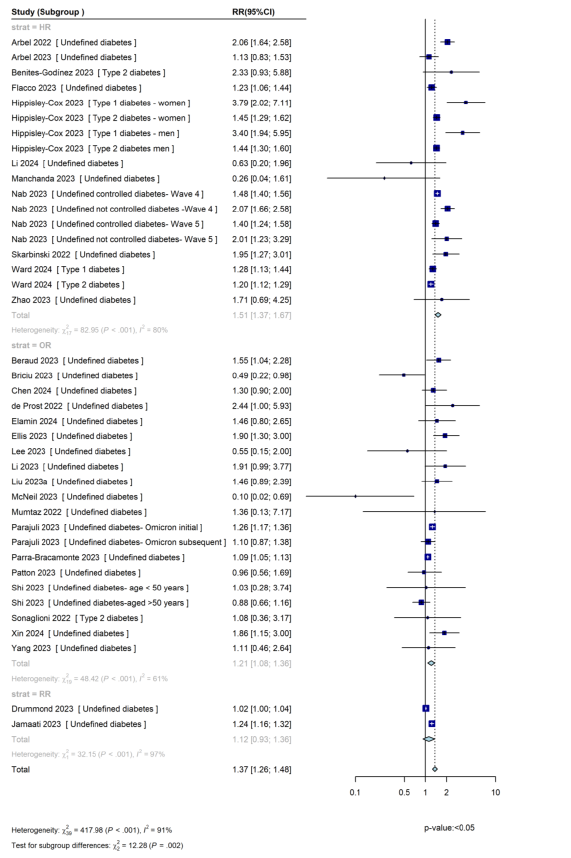

CI, confidence interval; HR, hazard ratio; OR, odds ratio; RR, risk ratio.

###### 2.4.4.2. Supplemental eFigure 11. Diabetes, Hospitalization Outcome (No. of Subgroups: 13)

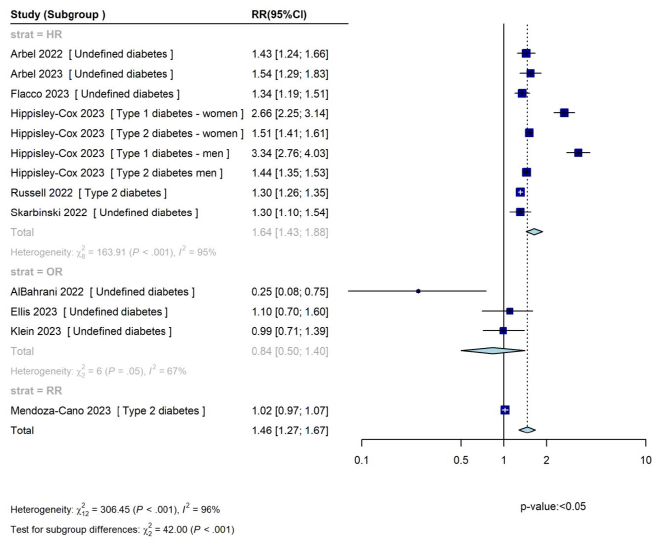

CI, confidence interval; HR, hazard ratio; OR, odds ratio; RR, risk ratio.

###### 2.4.4.3. Supplemental eFigure 12. Diabetes, ICU Admission Outcome (No. of Subgroups: 13)

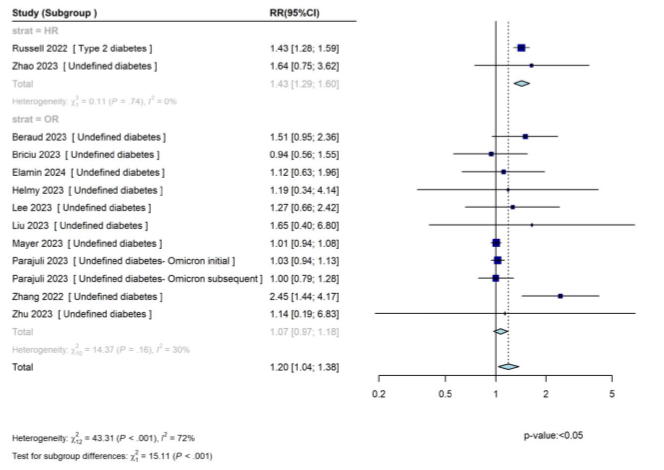

CI, confidence interval; HR, hazard ratio; OR, odds ratio; RR, risk ratio.

###### 2.4.4.4. Supplemental eFigure 13. Diabetes, Combined Outcome (No. of Subgroups: 19)

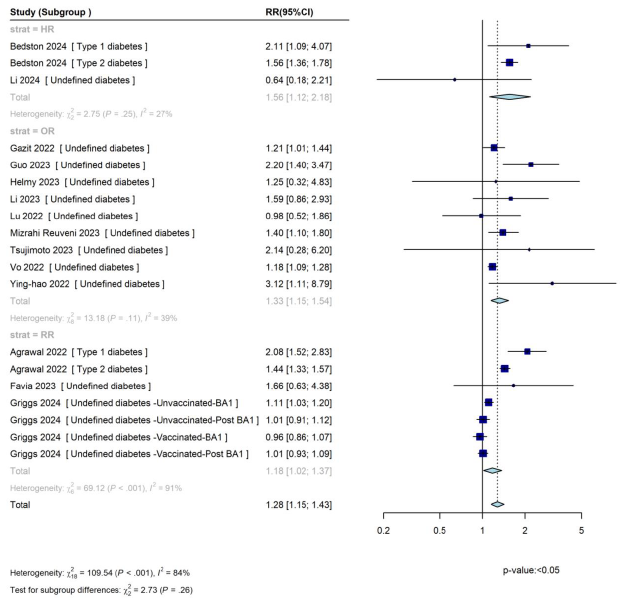

CI, confidence interval; HR, hazard ratio; OR, odds ratio; RR, risk ratio.

##### 2.4.5. Obesity

###### 2.4.5.1. Supplemental eFigure 14. Obesity, Death Outcome (No. of Subgroups: 21)

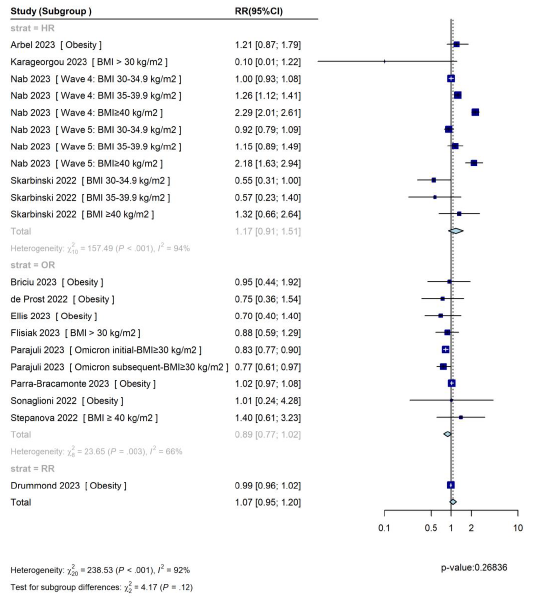

BMI, body mass index; CI, confidence interval; HR, hazard ratio; OR, odds ratio; RR, risk ratio.

###### 2.4.5.2. Supplemental Figure 15. Obesity, Hospitalization Outcome (No. of Subgroups: 9)

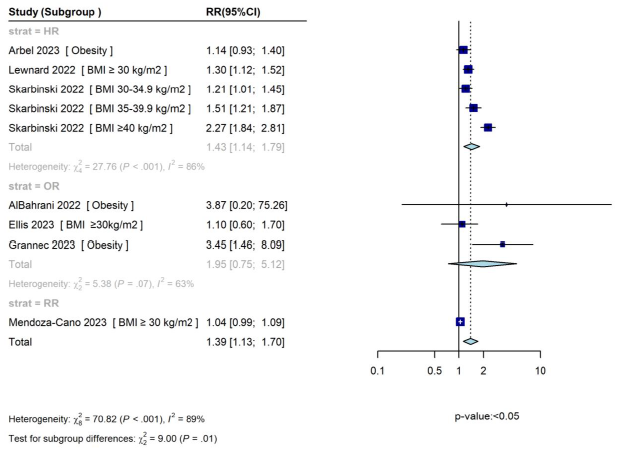

BMI, body mass index; CI, confidence interval; HR, hazard ratio; OR, odds ratio; RR, risk ratio.

###### 2.4.5.3. Supplemental eFigure 16. Obesity, ICU Admission Outcome (No. of Subgroups: 6)

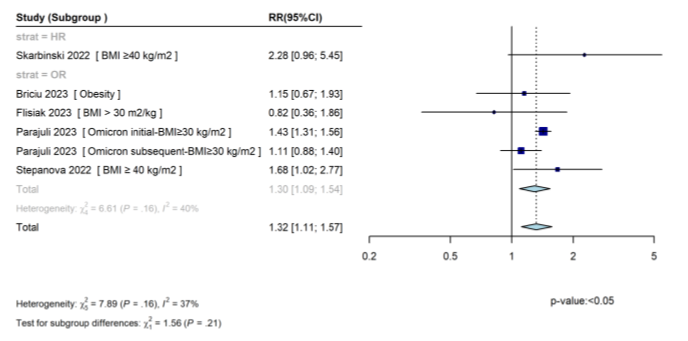

BMI, body mass index; CI, confidence interval; HR, hazard ratio; OR, odds ratio; RR, risk ratio.

###### 2.4.5.4. Supplemental eFigure 17. Obesity, Combined Outcome (No. of Subgroups: 7)

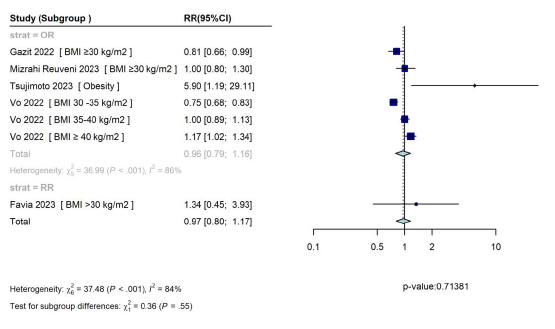

BMI, body mass index; CI, confidence interval; OR, odds ratio; RR, risk ratio.

##### 2.4.6. Peripheral Vascular Disease

###### 2.4.6.1. Peripheral Vascular Disease, Death Outcome (No. of Studies: 2; No. of Subgroups Included in ‘Main’ Analysis: 3)

The studies reported the following results for the ‘Death’ outcome in the ‘PVD’ populations:

- Hippisley-Cox 2023^36^ (Women with PVD): HR (95% CI): 1.51 (1.23, 1.86)
- Hippisley-Cox 2023^36^ (Men with PVD): HR (95% CI): 1.07 (0.90, 1.28)
- Ward 2024^72^ (PVD): HR (95% CI): 1.39 (1.21, 1.60)

Hippisley-Cox 2023 found a significant increase in the risk of death for women with PVD, but not for men with PVD, while Ward 2024 found a significant increase in death risk overall (significance level is based on the CIs).

###### 2.4.6.2. Peripheral Vascular Disease, Hospitalization Outcome (No. of Studies: 1; No. of Subgroups Included in ‘Main’ Analysis: 2)

The studies reported the following results for the ‘Hospitalization’ outcome in the ‘PVD’ populations:

- Hippisley-Cox 2023^36^ (Women with PVD): HR (95% CI): 1.34 (1.14, 1.57)
- Hippisley-Cox 2023^36^ (Men with PVD): HR (95% CI): HR: 1.11 (0.98, 1.25)

Hippisley-Cox 2023 reported a significant increase in the risk of death for women with PVD, but not for men with PVD (significance level is based on the CIs).

###### 2.4.6.3. Peripheral Vascular Disease, ICU Admission Outcome (No. of Studies: 0)

ICU analysis for the ‘PVD’ population was not feasible as there were no studies included.

###### 2.4.6.4. Peripheral Vascular Disease, Combined Outcome (No. of Studies: 3)

The studies reported the following results for the ‘Combined’ outcome in the ‘PVD’ populations:

- Agrawal 2022^8^ (PVD): OR (95% CI): 1.80 (1.69, 1.91)
- Bedston 2024^13^ (PVD): HR: (95% CI): 2.06 (1.81, 2.34)
- Vo 2022^7^ (PVD): OR (95% CI): 1.25 (1.09, 1.44)

All studies showed a significant association of the increased risk for the ‘Combined’ outcome in the ‘PVD’ populations (significance level is based on the CIs).

##### 2.4.7. Respiratory Diseases

###### 2.4.7.1. Supplemental eFigure 18. Respiratory Diseases, Death Outcome (No. of Subgroups: 23)

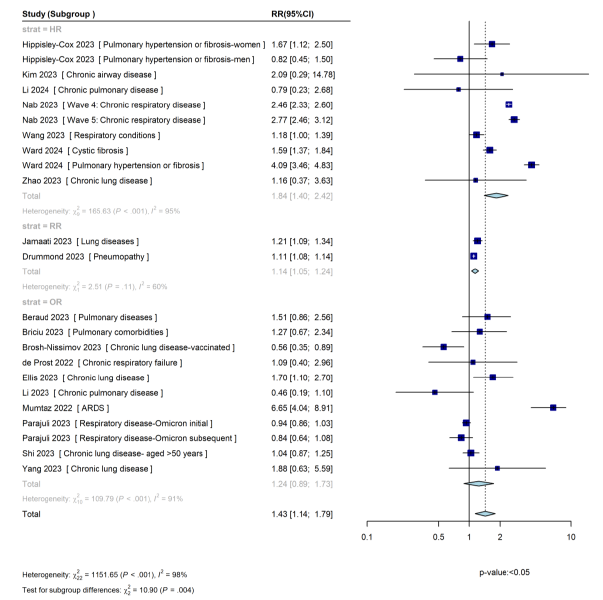

ARDS, Acute respiratory distress syndrome; CI, confidence interval; HR, hazard ratio; OR, odds ratio; RR, risk ratio.

###### 2.4.7.2. Supplemental eFigure 19. Respiratory Diseases, Hospitalization Outcome (No. of Subgroups: 6)

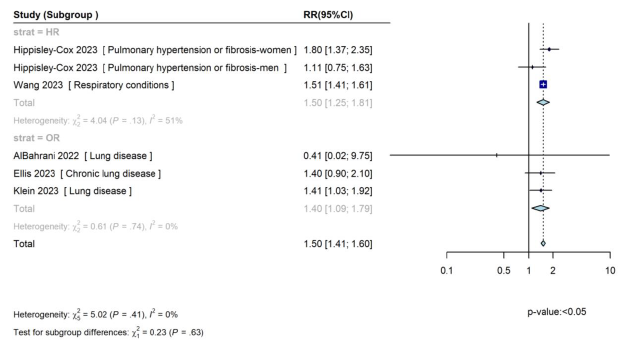

CI, confidence interval; HR, hazard ratio; OR, odds ratio; RR, risk ratio.

###### 2.4.7.3. Supplemental eFigure 20. Respiratory Diseases, ICU Admission Outcome (No. of Subgroups: 7)

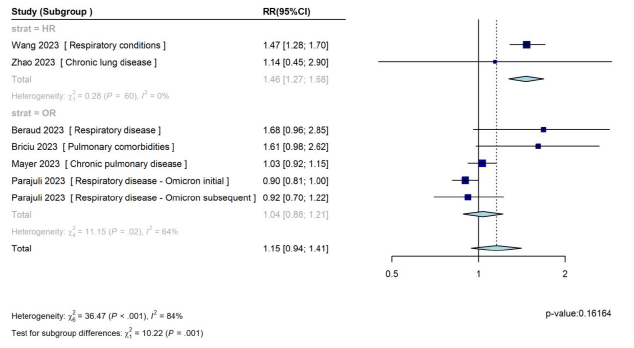

CI, confidence interval; HR, hazard ratio; OR, odds ratio; RR, risk ratio.

###### 2.4.7.4. Supplemental eFigure 21. Respiratory Diseases, Combined Outcome (No. of Subgroups: 10)

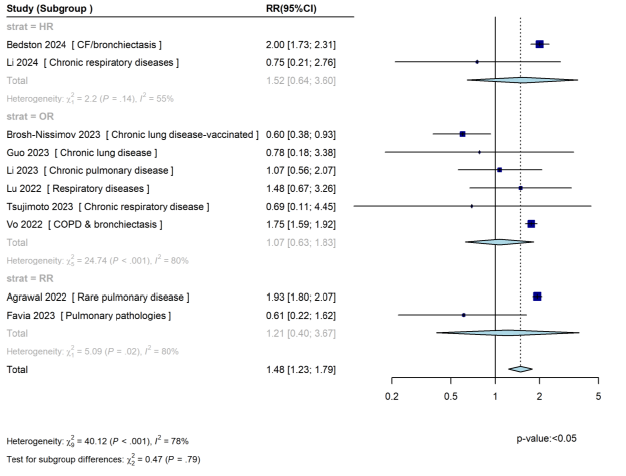

CF, cystic fibrosis; CI, confidence interval; COPD, chronic obstructive pulmonary disorder; HR, hazard ratio; OR, odds ratio; RR, risk ratio.

##### 2.4.8. Atrial Fibrillation

###### 2.4.8.1. Atrial Fibrillation, Death Outcome (No. of Studies: 3; No. of Subgroups Included in ‘Main’ Analysis: 4)

The studies reported the following results for the ‘Death’ outcome in the ‘Atrial fibrillation’ populations:

- Ellis 2023^25^ (Atrial fibrillation): OR (95% CI): 1.2 (0.7, 1.9)
- Hippisley-Cox 2023^36^ (Women with atrial fibrillation): HR (95% CI): 1.09 (0.96, 1.24)
- Hippisley-Cox 2023^36^ (Men with atrial fibrillation): HR (95% CI): 1.18 (1.06, 1.32)
- Ward 2024^72^ (Atrial fibrillation): HR (95% CI): 1.26 (1.18, 1.34)

Based on the CIs, all except 1 study (Ellis 2023) showed a significantly increased association between ‘Atrial fibrillation’ and the ‘Death’ outcome.

###### 2.4.8.2. Atrial Fibrillation, Hospitalization Outcome (No. of Studies: 4; No. of Subgroups Included in ‘Main’ Analysis: 4)

The studies reported the following results for the ‘Hospitalization’ outcome in the ‘Atrial fibrillation’ populations:

- Ellis 2023^25^ (Atrial fibrillation): OR (95% CI): 1.1 (0.7, 1.6)
- Hippisley-Cox 2023^36^ (Women with atrial fibrillation): HR (95% CI): 1.19 (1.08, 1.30)
- Hippisley-Cox 2023^36^ (Men with atrial fibrillation): HR (95% CI): 1.11 (1.03, 1.20)
- Klein 2023^40^ (Atrial fibrillation): OR (95% CI): 0.58 (0.38, 0.88)
- Morris 2023^53^ (Atrial fibrillation): OR: 5.83

Based on the CIs, only Hippisley-Cox 2023 reported a significantly increased association between ‘Hospitalization’ outcome and ‘Atrial fibrillation’ (for both men and women). Morris 2023 did not provide a CI; therefore, the precision and significance of the result are unclear. The study also did not report the number of the participants, rendering the interpretation difficult.

###### 2.4.8.3. Atrial Fibrillation, ICU Admission Outcome (No. of Studies: 2; No. of Studies Included in ‘Main’ Analysis: 2)

The studies reported the following results for the ‘ICU’ outcome in the ‘Atrial fibrillation’ populations:

- Helmy 2023^35^ (Atrial fibrillation): OR (95% CI): 6.40 (0.15, 275.62)
- Mayer 2023^49^ (Atrial fibrillation): OR (95% CI): 1.02 (0.95, 1.09)

While both studies showed an increased risk of ‘ICU admission’ in ‘Atrial fibrillation’ populations, the results were not statistically significant. However, the population of the Helmy 2023 study is very small, indicating that a larger sample size is needed to investigate the true effect of this analysis.

###### 2.4.8.4. Supplemental eFigure 22. Atrial Fibrillation, Combined Outcome (No. of Subgroups: 5)

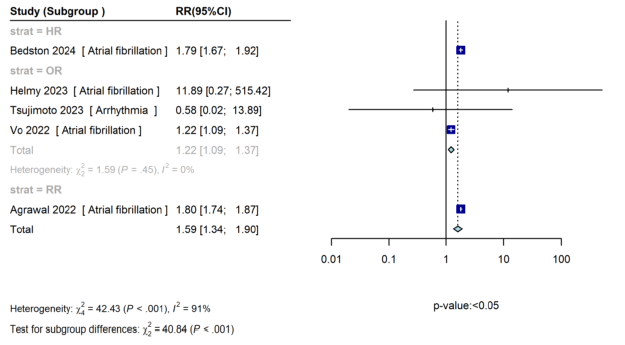

CI, confidence interval; HR, hazard ratio; OR, odds ratio; RR, risk ratio.

##### 2.4.9. Heart Disease

###### 2.4.9.1. Supplemental eFigure 23. Heart Disease, Death Outcome (No. of Subgroups: 29)

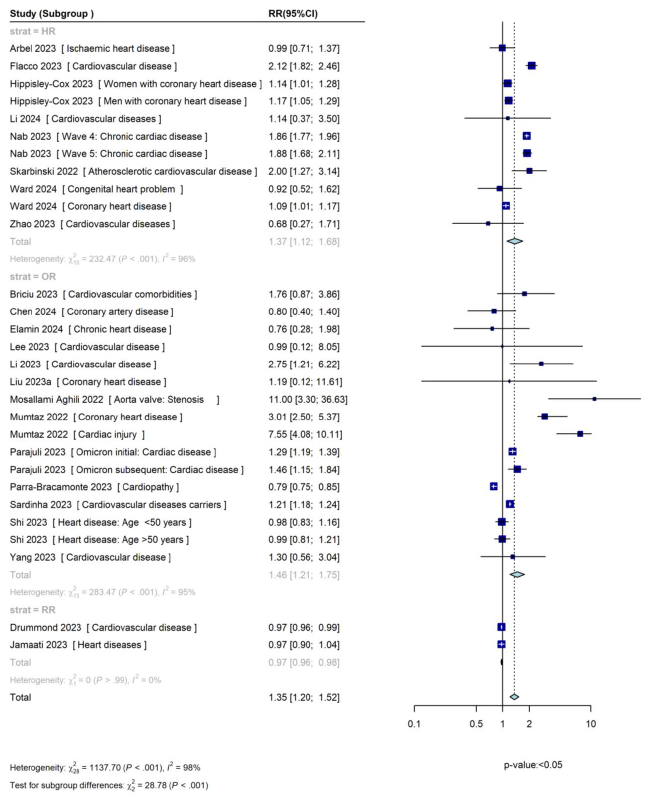

CI, confidence interval; HR, hazard ratio; OR, odds ratio; RR, risk ratio.

###### 2.4.9.2. Supplemental eFigure 24. Heart Disease, Hospitalization Outcome (No. of Subgroups: 8)

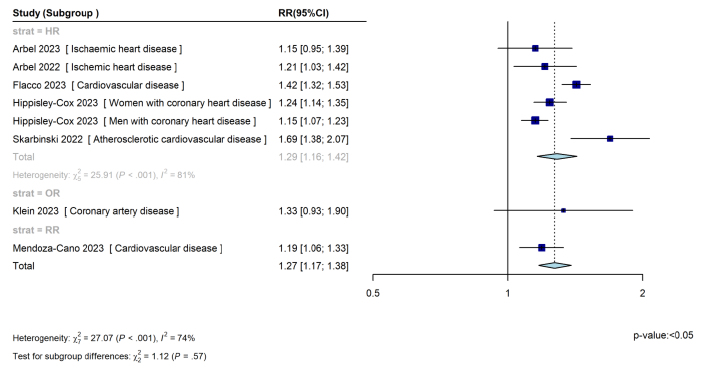

CI, confidence interval; HR, hazard ratio; OR, odds ratio; RR, risk ratio.

###### 2.4.9.3. Supplemental eFigure 25. Heart Disease, ICU Admission Outcome (No. of Subgroups: 11)

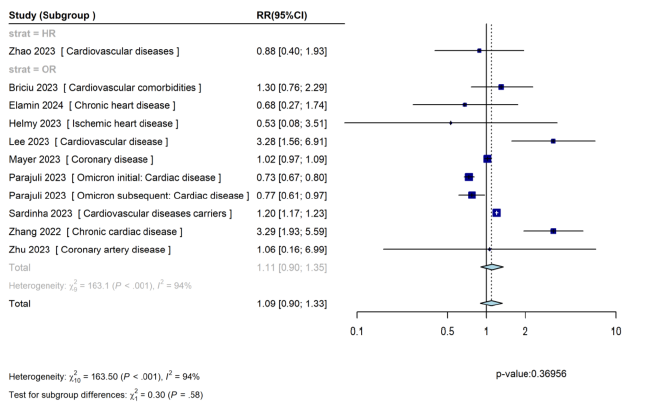

CI, confidence interval; HR, hazard ratio; OR, odds ratio; RR, risk ratio.

###### 2.4.9.4. Supplemental eFigure 26. Heart Disease, Combined Outcome (No. of Subgroups: 17)

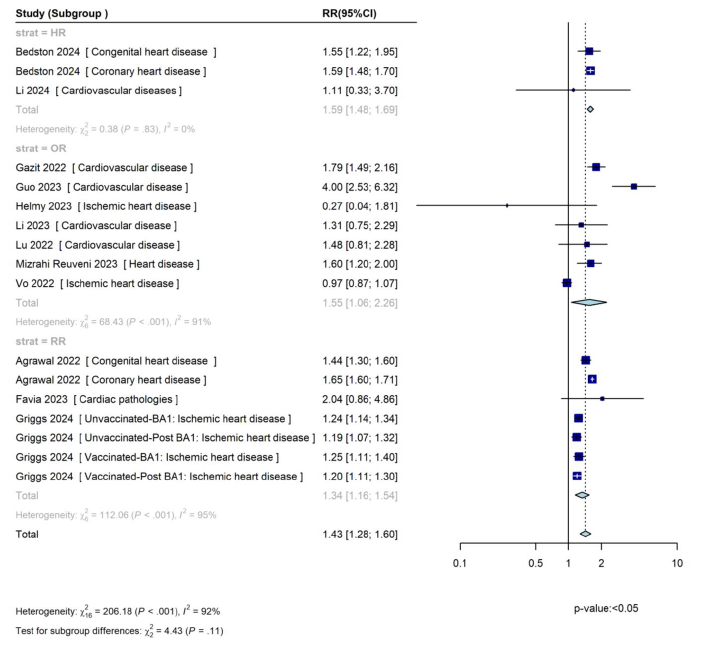

CI, confidence interval; HR, hazard ratio; OR, odds ratio; RR, risk ratio.

##### 2.4.10. Heart Failure

###### 2.4.10.1. Supplemental eFigure 27. Heart Failure, Death Outcome (No. of Subgroups: 11)

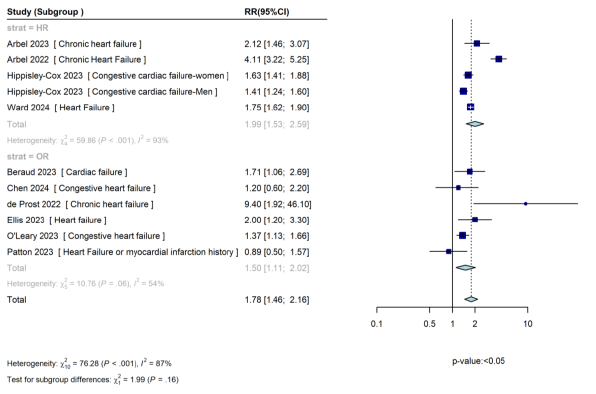

CI, confidence interval; HR, hazard ratio; OR, odds ratio; RR, risk ratio.

###### 2.4.10.2. Supplemental eFigure 28. Heart Failure, Hospitalization Outcome (No. of Subgroups: 7)

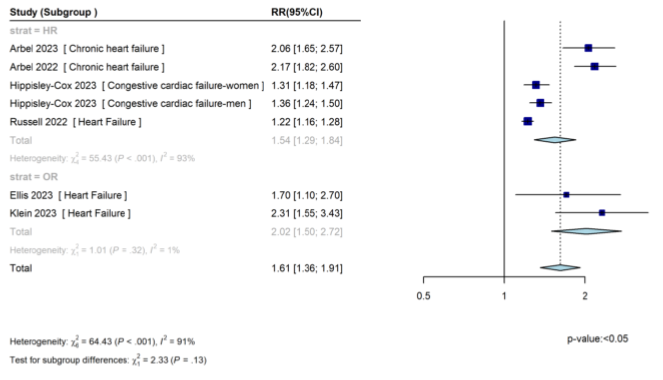

CI, confidence interval; HR, hazard ratio; OR, odds ratio; RR, risk ratio.

###### 2.4.10.3. Supplemental eFigure 29. Heart Failure, ICU Admission Outcome (No. of Studies: 3; No. of Subgroups Included in ‘Main’ Analysis: 3)

The studies reported the following results for the ‘ICU’ outcome in the ‘Heart failure’ populations:

- Beraud 2023^15^ (Cardiac failure): OR (95% CI): 1.39 (0.86, 2.19)
- Mayer 2023^49^ (Chronic heart failure): OR (95% CI): 1.05 (0.98, 1.14)
- Russell 2022^62^ (Heart failure): HR (95% CI): 1.24 (1.08, 1.44)

Based on the CIs, 1 out of the 3 studies (Russell 2022) yielded a significant result, although all studies exhibited a consistent trend – increase in the risk of the ‘ICU’ outcome.

###### 2.4.10.4. Supplemental eFigure 30. Heart Failure, Combined Outcome (No. of Subgroups: 7)

CI, confidence interval; HR, hazard ratio; OR, odds ratio; RR, risk ratio.

##### 2.4.11. Hypertension

###### 2.4.11.1. Supplemental eFigure 31. Hypertension, Death Outcome (No. of Subgroups: 23)

CI, confidence interval; HR, hazard ratio; OR, odds ratio; RR, risk ratio.

###### 2.4.11.2. Supplemental eFigure 32. Hypertension, Hospitalization Outcome (No. of Subgroups: 5)

CI, confidence interval; HR, hazard ratio; OR, odds ratio; RR, risk ratio.

###### 2.4.11.3. Supplemental eFigure 33. Hypertension, ICU Admission Outcome (No. of Subgroups: 7)

CI, confidence interval; OR, odds ratio; RR, risk ratio.

###### 2.4.11.4. Supplemental eFigure 34. Hypertension, Combined Outcome (No. of Subgroups: 17)

CI, confidence interval; HR, hazard ratio; OR, odds ratio; RR, risk ratio.

##### 2.4.12. Thrombosis

###### 2.4.12.1. Thrombosis, Death Outcome (No. of Studies: 2; No. of Subgroups Included in ‘Main’ Analysis: 3)

The studies reported the following results for the ‘Death’ outcome in the ‘Thrombosis’ populations:

- Hippisley-Cox 2023^36^ (Women with thromboembolism): HR (95% CI): 1.42 (1.23, 1.63)
- Hippisley-Cox 2023^36^ (Men with thromboembolism): HR (95% CI): 1.29 (1.11, 1.50)
- Ward 2024^72^ (Thrombosis or pulmonary embolus): HR (95% CI): 1.93 (0.62, 5.99)

Based on the CIs, the results of 2 out of the 3 population subgroups of interest (men and women with thromboembolism) were statistically significant, although all studies exhibited a consistent trend – increase in the risk of the ‘Death’ outcome.

###### 2.4.12.2. Thrombosis, Hospitalization Outcome (No. of Studies: 1; No. of Subgroups Included in ‘Main’ Analysis: 2)

The studies reported the following results for the ‘Hospitalization’ outcome in the ‘Thrombosis’ populations:

- Hippisley-Cox 2023^36^ (Women with thromboembolism-women): HR (95% CI): 1.50 (1.38, 1.63)
- Hippisley-Cox 2023^36^ (Men with thromboembolism-men): HR (95% CI): 1.44 (1.31, 1.57)

Based on the CIs, the results of both population subgroups showed statistically significant increased associations between ‘Thrombosis’ and ‘Hospitalization’.

###### 2.4.12.3. Thrombosis, ICU Admission Outcome (No. of Studies: 0)

‘ICU’ outcome analysis in the ‘Thrombosis’ population was not feasible as there were no studies included.

###### 2.4.12.4. Thrombosis, Combined Outcome (No. of Studies: 2; No. of Subgroups Included in ‘Main’ Analysis: 2)

The studies reported the following results for the ‘Combined’ outcome in the ‘Thrombosis’ populations:

- Agrawal 2022^8^ (Thrombosis or pulmonary embolus): RR (95% CI): 2.27 (2.15, 2.40)
- Bedston 2024^13^ (Thrombosis or pulmonary embolus): HR (95% CI): 1.93 (1.71, 2.18)

Based on the CIs, the results of both studies showed a statistically significant increase in the ‘Combined’ outcome of ‘Thrombosis’ populations.

### 3. References

**1.**	Wells G SB, O’Connell D, Peterson J, Welch V, Losos M. The Newcastle-Ottawa Scale (NOS) for assessing the quality of nonrandomised studies in meta-analyses. <https://www.ohri.ca/programs/clinical_epidemiology/oxford.asp>.

**2.**	JBI. Critical appraisal tools. <https://jbi.global/critical-appraisal-tools>.

**3.**	DerSimonian R, Laird N. Meta-analysis in clinical trials. *Control Clin Trials*. 1986;7(3):177-188.

**4.**	Higgins JPT TJ, Chandler J, Li T, Page MJ, Welch V, eds. *Cochrane Handbook for Systematic Reviews of Interventions [Internet]. Version 6.4 Cochrane, updated August 2023. [cited 2024 May 28]. Available from:* [*https://training.cochrane.org/handbook/current*](https://training.cochrane.org/handbook/current).

**5.**	Stuck AE, Rubenstein LZ, Wieland D. Bias in meta-analysis detected by a simple, graphical test. Asymmetry detected in funnel plot was probably due to true heterogeneity. *BMJ*. 1998;316(7129):469; author reply 470-461.

**6.**	Viechtbauer W. Conducting meta-analyses in R with the metafor package. *J Stat Softw*. 2010;36(3):1-48.

**7.**	Vo AD, La J, Wu JT, et al. Factors associated with severe COVID-19 among vaccinated adults treated in US veterans affairs hospitals. *JAMA Netw Open*. 2022;5(10):e2240037.

**8.**	Agrawal U, Bedston S, McCowan C, et al. Severe COVID-19 outcomes after full vaccination of primary schedule and initial boosters: pooled analysis of national prospective cohort studies of 30 million individuals in England, Northern Ireland, Scotland, and Wales. *Lancet*. 2022;400(10360):1305-1320.

**9.**	AlBahrani S, AlBarrak A, Al-Musawi T, et al. COVID-19 vaccine had a significant positive impact on patients with SARS-COV-2 during the third (Omicron) wave in Saudi Arabia. *J Infect Public Health*. 2022;15(11):1169-1174.

**10.**	Arbel R, Peretz A, Sergienko R, et al. Effectiveness of a bivalent mRNA vaccine booster dose to prevent severe COVID-19 outcomes: a retrospective cohort study. *Lancet Infect Dis*. 2023;23(8):914-921.

**11.**	Arbel R, Sergienko R, Friger M, et al. Effectiveness of a second BNT162b2 booster vaccine against hospitalization and death from COVID-19 in adults aged over 60 years. *Nat Med*. 2022;28(7):1486-1490.

**12.**	Bao S, Lu G, Kang Y, et al. A diagnostic model for serious COVID-19 infection among older adults in Shanghai during the Omicron wave. *Front Med (Lausanne)*. 2022;9:1018516.

**13.**	Bedston S, Almaghrabi F, Patterson L, et al. Risk of severe COVID-19 outcomes after autumn 2022 COVID-19 booster vaccinations: a pooled analysis of national prospective cohort studies involving 7.4 million adults in England, Northern Ireland, Scotland and Wales. *Lancet Reg Health Eur*. 2024;37:100816.

**14.**	Benites-Godínez V, Mendoza-Cano O, Trujillo X, et al. Survival analysis and contributing factors among PCR-confirmed adult inpatients during the endemic phase of COVID-19. *Diseases*. 2023;11(3):119.

**15.**	Beraud G, Bouetard L, Civljak R, et al. Impact of vaccination on the presence and severity of symptoms in hospitalized patients with an infection of the Omicron variant (B.1.1.529) of the SARS-CoV-2 (subvariant BA.1). *Clin Microbiol Infect*. 2023;29(5):642-650.

**16.**	Briciu V, Topan A, Calin M, et al. Comparison of COVID-19 severity in vaccinated and unvaccinated patients during the Delta and Omicron wave of the pandemic in a Romanian tertiary infectious diseases hospital. *Healthcare*. 2023;11(3):373.

**17.**	Brosh-Nissimov T, Hussein K, Wiener-Well Y, et al. Hospitalized patients with severe coronavirus disease 2019 during the Omicron wave in Israel: benefits of a fourth vaccine dose. *Clin Infect Dis*. 2022;76(3):e234-e239.

**18.**	Bulgaresi M, Rivasi G, Tarantini F, et al. Impact of SARS-CoV2 infection on mortality and hospitalization in nursing home residents during the “Omicron era”. *Aging Clin Exp Res*. 2023;35(6):1393-1399.

**19.**	Chan M, Owens L, Gray ML, et al. Asthma and susceptibility to COVID-19 in Australian children during Alpha, Delta and Omicron waves of the COVID-19 pandemic. *J Asthma Allergy*. 2023;16(null):1139-1155.

**20.**	Chen CL, Teng CK, Chen WC, et al. Clinical characteristics and treatment outcomes among the hospitalized elderly patients with COVID-19 during the late pandemic phase in central Taiwan. *J Microbiol Immunol Infect*. 2024;57(2):257-268.

**21.**	Choi S-H, Choi JH, Lee JK, et al. Clinical characteristics and outcomes of children with SARS-CoV-2 infection during the Delta and Omicron variant-dominant periods in Korea. *J Korean Med Sci*. 2023;38(9):e65.

**22.**	de Prost N, Audureau E, Heming N, et al. Clinical phenotypes and outcomes associated with SARS-CoV-2 variant Omicron in critically ill French patients with COVID-19. *Nat Commun*. 2022;13(1):6025.

**23.**	Drummond PD, de Salles DB, de Souza NSH, et al. Profile and outcomes of hospitalized COVID-19 patients during the prevalence of the Omicron variant according to the Brazilian regions: a retrospective cohort study from 2022. *Vaccines*. 2023;11(10):1568.

**24.**	Elamin MY, Maslamani YA, Alsheikh FA, et al. Impact of vaccination on morbidity and mortality in adults hospitalized with COVID-19 during the omicron wave in the Jazan Region, Saudi Arabia. *Saudi Med J*. 2024;45(2):179-187.

**25.**	Ellis RJ, Moffatt CR, Aaron LT, et al. Factors associated with hospitalisations and deaths of residential aged care residents with COVID-19 during the Omicron (BA.1) wave in Queensland. *Med J Aust*. 2023;218(4):174-179.

**26.**	Favia G, Barile G, Tempesta A, et al. Relationship between oral lesions and severe SARS-CoV-2 infection in intensive care unit patients. *Oral Dis*. 2024;30(3):1296-1303.

**27.**	Finkas LK, Ramesh N, Block LS, et al. Asthma and COVID-19 outcomes: a prospective study in a large health care delivery system. *J Asthma Allergy*. 2023;16:1041-1051.

**28.**	Flacco ME, Acuti Martellucci C, Soldato G, et al. Predictors of SARS-CoV-2 infection and severe and lethal COVID-19 after three years of follow-up: a population-wide study. *Viruses*. 2023;15(9):1794.

**29.**	Flisiak R, Zarębska-Michaluk D, Dobrowolska K, et al. Change in the clinical picture of hospitalized patients with COVID-19 between the early and late period of dominance of the Omicron SARS-CoV-2 variant. *J Clin Med*. 2023;12(17):5572.

**30.**	Gazit S, Saciuk Y, Perez G, et al. Short term, relative effectiveness of four doses versus three doses of BNT162b2 vaccine in people aged 60 years and older in Israel: retrospective, test negative, case-control study. *BMJ*. 2022;377:e071113.

**31.**	Geng Y, Nie Q, Liu F, et al. Understanding clinical characteristics influencing adverse outcomes of Omicron infection: a retrospective study with propensity score matching from a Fangcang hospital. *Front Cell Infect Microbiol*. 2023;13:1115089.

**32.**	Grannec F, Meddeb L, Tissot-Dupont H, Gentile S, Brouqui P. Pre-hospital management of patients with COVID-19 and the impact on hospitalization. *Medicina*. 2023;59(8):1440.

**33.**	Griggs EP, Mitchell PK, Lazariu V, et al. Clinical epidemiology and risk factors for critical outcomes among vaccinated and unvaccinated adults hospitalized with COVID-19-VISION Network, 10 states, June 2021-March 2023. *Clin Infect Dis*. 2024;78(2):338-348.

**34.**	Guo Y, Guo Y, Ying H, et al. In-hospital adverse outcomes and risk factors among chronic kidney disease patients infected with the omicron variant of SARS-CoV-2: a single-center retrospective study. *BMC Infect Dis*. 2023;23(1):698.

**35.**	Helmy MA, Milad LM, Hasanin AM, et al. Parasternal intercostal thickening at hospital admission: a promising indicator for mechanical ventilation risk in subjects with severe COVID-19. *J Clin Monit Comput*. 2023;37(5):1287-1293.

**36.**	Hippisley-Cox J, Khunti K, Sheikh A, Nguyen-Van-Tam JS, Coupland CAC. Risk prediction of covid-19 related death or hospital admission in adults testing positive for SARS-CoV-2 infection during the omicron wave in England (QCOVID4): cohort study. *BMJ*. 2023;381:e072976.

**37.**	Jamaati H, Karimi S, Ghorbani F, et al. Effectiveness of different vaccine platforms in reducing mortality and length of ICU stay in severe and critical cases of COVID-19 in the Omicron variant era: a national cohort study in Iran. *J Med Virol*. 2023;95(3):e28607.

**38.**	Karageorgou V, Papaioannou AI, Kallieri M, et al. Patients hospitalized for COVID-19 in the periods of Delta and Omicron variant dominance in Greece: determinants of severity and mortality. *J Clin Med*. 2023;12(18):5904.

**39.**	Kim SH, Kim T, Choi H, Shin TR, Sim YS. Clinical outcome and prognosis of a nosocomial outbreak of COVID-19. *J Clin Med*. 2023;12(6):2279.

**40.**	Klein EY, Fall A, Norton JM, et al. Severity outcomes associated with SARS-CoV-2 XBB variants, an observational analysis. *J Clin Virol*. 2023;165:105500.

**41.**	Lee CM, Kim M, Park SW, et al. Clinical outcomes and immunological features of COVID-19 patients receiving B-cell depletion therapy during the Omicron era. *Infect Dis (Lond)*. 2024;56(2):116-127.

**42.**	Lewnard JA, Hong VX, Patel MM, et al. Clinical outcomes associated with SARS-CoV-2 Omicron (B.1.1.529) variant and BA.1/BA.1.1 or BA.2 subvariant infection in Southern California. *Nat Med*. 2022;28(9):1933-1943.

**43.**	Li D-J, Zhou C-C, Huang F, Shen F-M, Li Y-C. Clinical features of omicron SARS-CoV-2 variants infection associated with co-infection and ICU-acquired infection in ICU patients. *Front Public Health*. 2024;11:1320340.

**44.**	Li H, Jia X, Wang Y, et al. Differences in the severity and mortality risk factors for patients hospitalized for COVID-19 pneumonia between the early wave and the very late stage of the pandemic. *Front Med (Lausanne)*. 2023;10:1238713.

**45.**	Liu Y, Chen D, Li J, et al. Metabolic syndrome is associated with poor Omicron infection prognosis while inactivated vaccine improves the outcome of Coronavirus Disease 2019 among Chinese inhabitants: a retrospective observational study from a Chinese municipality. *Vaccines*. 2023;11(10):1554.

**46.**	Liu Y, Qi Z, Bai M, et al. Combination of chest computed tomography value and clinical laboratory data for the prognostic risk evaluation of patients with COVID-19. *Int J Gen Med*. 2023;16:3829-3842.

**47.**	Lu G, Zhang Y, Zhang H, et al. Geriatric risk and protective factors for serious COVID-19 outcomes among older adults in Shanghai Omicron wave. *Emerg Microbes Infect*. 2022;11(1):2045-2054.

**48.**	Manchanda V, Mitra S, Rafique I, et al. Is Omicron really mild? – Comparative analysis of comorbidities and disease outcomes associated with SARS-CoV-2 Omicron (B.1.1.529) and Delta (B.1.617.2) variants. *Indian J Med Microbiol*. 2023;45:100391.

**49.**	Mayer C, Woo MS, Brehm TT, et al. History of cerebrovascular disease but not dementia increases the risk for secondary vascular events during SARS-CoV-2 infection with presumed Omicron variant: a retrospective observational study. *Eur J Neurol*. 2023;30(8):2297-2304.

**50.**	McNeil T, Zhang F, Moffatt S, Emeto TI, Tucker E. Nosocomial COVID-19 infection in the era of vaccination and antiviral therapy. *Intern Med J*. 2024;54(3):374-381.

**51.**	Mendoza-Cano O, Trujillo X, Ríos-Silva M, et al. Association between vaccination status for COVID-19 and the risk of severe symptoms during the endemic phase of the disease. *Vaccines*. 2023;11(10):1512.

**52.**	Mizrahi Reuveni M, Kertes J, Shapiro Ben David S, et al. Risk stratification model for severe COVID-19 disease: a retrospective cohort study. *Biomedicines*. 2023;11(3):767.

**53.**	Morris CP, Eldesouki RE, Sachithanandham J, et al. Omicron subvariants: clinical, laboratory, and cell culture characterization. *Clin Infect Dis*. 2023;76(7):1276-1284.

**54.**	Mosallami Aghili SM, Khoshfetrat M, Asgari A, et al. Association of echocardiographic findings with in-hospital mortality of COVID-19 patients and their changes in one-month follow-up; a cohort study. *Arch Acad Emerg Med*. 2022;10(1):e85.

**55.**	Mumtaz A, Rehman E, Rahaman MA, Rehman S. Inflammatory biomarkers and cardiac injury in COVID-19 patients. *Front Public Health*. 2022;10:1024535.

**56.**	Nab L, Parker EPK, Andrews CD, et al. Changes in COVID-19-related mortality across key demographic and clinical subgroups in England from 2020 to 2022: a retrospective cohort study using the OpenSAFELY platform. *Lancet Public Health*. 2023;8(5):e364-e377.

**57.**	O'Leary AL, Wattengel BA, Carter MT, Drye AF, Mergenhagen KA. Risk factors associated with mortality in hospitalized patients with laboratory confirmed SARS-CoV-2 infection during the period of omicron (B.1.1.529) variant predominance. *Am J Infect Control*. 2023;51(6):603-606.

**58.**	Parajuli P, Sabo R, Alsaadawi R, et al. Fibrosis-4 (FIB-4) index as a predictor for mechanical ventilation and 30-day mortality across COVID-19 variants. *J Clin Transl Sci*. 2023;7(1):e213.

**59.**	Parra-Bracamonte GM, Lopez-Villalobos N, Velazquez MA, et al. Comparative analysis of risk factors for COVID-19 mortality before, during and after the vaccination programme in Mexico. *Public Health*. 2023;215:94-99.

**60.**	Patton MJ, Orihuela CJ, Harrod KS, et al. COVID-19 bacteremic co-infection is a major risk factor for mortality, ICU admission, and mechanical ventilation. *Crit Care*. 2023;27(1):34.

**61.**	Radhakrishnan N, Liu M, Idowu B, et al. Comparison of the clinical characteristics of SARS-CoV-2 Delta (B.1.617.2) and Omicron (B.1.1.529) infected patients from a single hospitalist service. *BMC Infect Dis*. 2023;23(1):747.

**62.**	Russell SL, Klaver BRA, Harrigan SP, et al. Clinical severity of Omicron subvariants BA.1, BA.2, and BA.5 in a population-based cohort study in British Columbia, Canada. *J Med Virol*. 2023;95(1):e28423.

**63.**	Sardinha DM, Ferreira ALdS, Guimarães RJdPSe, Lima KVB, Lima LNGC. Clinical characteristics and outcomes among vaccinated and unvaccinated patients with cardiovascular disease who were hospitalized for COVID-19 in Brazil: retrospective cohort. *Vaccines*. 2023;11(4):861.

**64.**	Shakor ASaA, Samsudin EZ, Chen XW, Ghazali MH. Factors associated with COVID-19 brought-in deaths: a data-linkage comparative cross-sectional study. *J Infect Public Health*. 2023;16(12):2068-2078.

**65.**	Shi HJ, Yang J, Eom JS, et al. Clinical characteristics and risk factors for mortality in critical COVID-19 patients aged 50 years or younger during Omicron wave in Korea: comparison with patients older than 50 years of age. *J Korean Med Sci*. 2023;38(28):e217.

**66.**	Simmons AE, Amoako A, Grima AA, et al. Vaccine effectiveness against hospitalization among adolescent and pediatric SARS-CoV-2 cases between May 2021 and January 2022 in Ontario, Canada: a retrospective cohort study. *PLoS One*. 2023;18(3):e0283715.

**67.**	Skarbinski J, Wood MS, Chervo TC, et al. Risk of severe clinical outcomes among persons with SARS-CoV-2 infection with differing levels of vaccination during widespread Omicron (B.1.1.529) and Delta (B.1.617.2) variant circulation in Northern California: a retrospective cohort study. *Lancet Reg Health Am*. 2022;12:100297.

**68.**	Sonaglioni A, Lombardo M, Albini A, et al. Charlson comorbidity index, neutrophil-to-lymphocyte ratio and undertreatment with renin-angiotensin-aldosterone system inhibitors predict in-hospital mortality of hospitalized COVID-19 patients during the omicron dominant period. *Front Immunol*. 2022;13:958418.

**69.**	Stepanova M, Lam B, Younossi E, et al. The impact of variants and vaccination on the mortality and resource utilization of hospitalized patients with COVID-19. *BMC Infect Dis*. 2022;22(1):702.

**70.**	Tsujimoto Y, Kobayashi M, Oku T, et al. Outcomes in novel hospital-at-home model for patients with COVID-19: a multicentre retrospective cohort study. *Fam Pract*. 2023;40(5-6):662-670.

**71.**	Wang X, Zein J, Ji X, Lin DY. Impact of vaccination, prior infection, and therapy on Omicron infection and mortality. *J Infect Dis*. 2023;227(8):970-976.

**72.**	Ward IL, Robertson C, Agrawal U, et al. Risk of COVID-19 death in adults who received booster COVID-19 vaccinations in England. *Nat Commun*. 2024;15(1):398.

**73.**	Xin S, Chen W, Yu Q, Gao L, Lu G. Effect of the number of coronavirus disease 2019 (COVID-19) vaccination shots on the occurrence of pneumonia, severe pneumonia, and death in SARS-CoV-2-infected patients. *Front Public Health*. 2024;11:1330106.

**74.**	Yang H, Wang Z, Zhang Y, et al. Clinical characteristics and factors for serious outcomes among outpatients infected with the Omicron subvariant BF.7. *J Med Virol*. 2023;95(8):e28977.

**75.**	Ying-Hao P, Yuan-Yuan G, Hai-Dong Z, et al. Clinical characteristics and analysis of risk factors for disease progression of patients with SARS-CoV-2 Omicron variant infection: a retrospective study of 25207 cases in a Fangcang hospital. *Front Cell Infect Microbiol*. 2022;12:1009894.

**76.**	Zhang Y, Han J, Sun F, et al. A practical scoring model to predict the occurrence of critical illness in hospitalized patients with SARS-CoV-2 omicron infection. *Front Microbiol*. 2022;13:1031231.

**77.**	Zhao Q, Zheng B, Han B, et al. Is azvudine comparable to nirmatrelvir-ritonavir in real-world efficacy and safety for hospitalized patients with COVID-19? a retrospective cohort study. *Infect Dis Ther*. 2023;12(8):2087-2102.

**78.**	Zhu Z, Cai J, Tang Q, et al. Circulating eosinophils associated with responsiveness to COVID-19 vaccine and the disease severity in patients with SARS-CoV-2 omicron variant infection. *BMC Pulm Med*. 2023;23(1):177.
